# Inexpensive multi-patient respiratory monitoring system for helmet ventilation during COVID-19 pandemic

**DOI:** 10.1101/2020.06.29.20141283

**Authors:** The Princeton Open Ventilation Monitor Collaboration, Philippe Bourrianne, Stanley Chidzik, Daniel Cohen, Peter Elmer, Thomas Hallowell, Todd J. Kilbaugh, David Lange, Andrew M. Leifer, Daniel R. Marlow, Peter D. Meyers, Edna Normand, Janine Nunes, Myungchul Oh, Lyman Page, Talmo Periera, Jim Pivarski, Henry Schreiner, Howard A. Stone, David W. Tank, Stephan Thiberge, Christopher Tully

**Affiliations:** Department of Mechanical and Aerospace Engineering, Princeton University, Princeton, NJ, 08544, USA; Department of Physics, Princeton University, Princeton, NJ, 08544, USA; Department of Anesthesiology and Critical Care Medicine, Children’s Hospital of Philadelphia, Philadelphia, PA 19104, USA; Princeton Neuroscience Institute, Princeton University, Princeton, NJ, 08544, USA; Department of Molecular Biology, Princeton University, Princeton, NJ, 08544, USA; Rutgers Robert Wood Johnson, New Brunswick, NJ, 08901 USA; Princeton Institute for Computational Science and Engineering, Princeton University, Princeton, NJ, 08544, USA

**Keywords:** COVID-19, non-invasive ventilation, CPAP, helmet, hood, respiratory profile monitor, remote monitoring, critical care, low-cost ventilator, emergency ventilator

## Abstract

**Background:** Helmet continuous positive applied pressure is a form of non-invasive ventilation (NIV) that has been used to provide respiratory support to COVID-19 patients. Helmet NIV is low-cost, readily available, provides viral filters between the patient and clinician, and may reduce the need for invasive ventilation. Its widespread adoption has been limited, however, by the lack of a respiratory monitoring system needed to address known safety vulnerabilities and to monitor patients. To address these safety and clinical needs, we developed an inexpensive respiratory monitoring system based on readily available components suitable for local manufacture. Open-source design and manufacturing documents are provided. The monitoring system comprises flow, pressure and CO_2_ sensors on the expiratory path of the helmet circuit and a central remote station to monitor up to 20 patients.

**Methods:** The system is validated in bench tests, in human-subject tests on healthy volunteers, and in experiments that compare respiratory features obtained at the expiratory path to simultaneous ground-truth measurements from proximal sensors.

**Findings:** Measurements of flow and pressure at the expiratory path are shown to deviate at high flow rates, and the tidal volumes reported via the expiratory path are systematically underestimated.

**Interpretation:** Helmet monitoring systems exhibit high-flow rate, non-linear effects from flow and helmet dynamics. These deviations are found to be within a reasonable margin and should, in principle, allow for calibration, correction and deployment of clinically accurate derived quantities.

**Funding:** This project is supported by Princeton University, and by National Science Foundation grants OAC-1836650, PHY-2031509 and IOS-1845137. The funding sources provided no role in the design or execution of the the work or in the preparation of the manuscript.

**Research in context:** *Evidence before this study:* Respiratory monitoring is standard when treating intubated patients undergoing invasive mechanical ventilation. In contrast, respiratory monitoring systems have not been developed for helmet non-invasive ventilation (NIV). Previous measurements of CO_2_ concentration in the helmet versus flow rate have been published and serve as the primary guide for setting the minimum flow rate for patient treatment of helmet NIV. Similar studies have explored optimal PEEP settings for clinical treatment. However, in practice, respiratory profiles are not measured during helmet treatment and more evidence is needed to evaluate whether clinically useful quantities, such as tidal volume, can be accurately measured during helmet NIV, to provide the same level of clincially relevant monitoring that is standard with invasive ventliation.

*Added value of this study:* Due to the widespread need for inexpensive multi-patient respiratory monitoring systems to cope with the COVID-19 pandemic, a helmet NIV monitoring system was developed and validated with bench tests, human-subject tests on healthy volunteers, and in experiments that compare respiratory features obtained at the expiratory path to simultaneous ground-truth measurements from proximal sensors. At high flow rate, the non-linear effects from the flow and helmet dynamics are observed and have a measurable effect on the estimation of tidal volumes and derived quantities.

*Implications of all the available evidence:* Helmet monitoring systems for NIV are in wide-spread use for the treatment of the coronavirus disease 2019. The introduction of respiratory monitoring systems for helmet NIV addresses important safety concerns and opens up the possibility of providing clinically relevant derived quantities to track disease progression. A systematic study of deviations between expiratory path measurements and ground-truth proximal sensors was conducted in bench tests and human-subject tests of health volunteers. The non-linear flow and helmet dynamics effects the accuracy of derived quantities at high flow rates. These deviations are found to be within a reasonable margin and should, in principle, allow for calibration, correction and deployment of clinically accurate derived quantities. An inexpensive implementation of the respiratory monitoring system was achieved to cope with the immense scale of the COVID-19 pandemic. Further steps to improve the quality of care for COVID-19 helmet NIV treatment can be achieved through the additional of respiratory monitoring systems that adjust for high flow-rate deviations in the estimation of tidal volumes and derived quantities.

## 1 Introduction

Non-invasive ventilation systems, such as continuous positive application of pressure (CPAP) and high flow nasal canula, have emerged as important tools for treating coronavirus disease 2019 (COVID-19) patients who need respiratory support but not intubation [1, 2]. Such non-invasive approaches conserve traditional ventilators and improve patient outcomes by reducing or eliminating the need for invasive ventilation [3]. Additionally, COVID-19 patients show surprisingly poor outcomes on invasive ventilation [4], making non-invasive ventilation (where applicable) uniquely valuable during the pandemic.

Helmet non-invasive ventilation (NIV) is a form of CPAP that uses a clear, polyvinyl chloride (PVC), bubble-like helmet, attached to a soft collar that seals around a patient’s neck [5]. The helmet delivers an air-oxygen mixture to the patient at a higher than atmospheric pressure with a a Positive End Expiratory Pressure (PEEP) valve. The PEEP serves to keep the patient’s airways open and the patient oxygenated, increases the Functional Residual Capacity (FRC) of the lung, recruiting collapsed alveoli and decreasing the left ventricular transmural pressure, and assists through other mechanisms in augmenting patient respiratory function [6]. Helmet NIV is appealing for pandemic use because it can be run directly from a constant flow of air-oxygen, which is readily available in hospitals [7]. Helmets cost less than 300 USD, are straightforward to manufacture, and have been available during the pandemic, despite supply chain disruptions for other ventilators [8, 9]. Importantly, Helmet NIV is an enclosed system that uses viral filters to protect clinicians and other patients from droplets or aerosolized SARS-CoV-2 viral particles shed by the patient [10]. Helmet NIV is permitted by the FDA for COVID-19 treatment [11].

Despite these advantages, lingering safety concerns have prevented wider adoption of the helmet in the United States. Steady airflow is required to clear the patient’s expired CO_2_ from accumulating in the helmet [12]. An unexpected drop of airflow caused by a disruption in gas supply or blockage in the circuit could lead to rebreathing of CO_2_ and asphyxiation [13]. Therefore, one of the greatest concerns when using helmets is the lack of an effective monitoring and alarm system. Antisuffocation valves can help mitigate this risk, but ultimately a dedicated monitoring and alarm system is needed to employ helmet NIV safely [14]. Partly for this reason, helmets in the US have been primarily restricted to the ICU, where clinician-to-patient ratios are high enough to allow clinicians to observe patients around-the-clock.

We have addressed these safety risks and clinical needs by developing an inexpensive respiratory profile monitoring system, which we call the Princeton Open Ventilator Monitor (POVM), that is based on readily available commercial components and is suitable for rapid fabrication during a pandemic. The system is designed for use in conjunction with non-invasive helmet ventilator systems, such as the Sea-Long Medical Systems COVID-19 Helmet [8] and the Subsalve Oxygen Treatment Hood [9], but is modular and can be used in other systems. The system comprises one or more flow, pressure and CO_2_ sensors per helmet, and includes a central station that can be used to remotely monitor up to 20 patients simultaneously. The system reports flow, pressure and CO_2_ concentration in the helmet, the patient’s equivalent tidal volume (≈ TV), respiratory rate (RR), ratio of inspiratory to expiratory time (I:E ratio), peak inspiratory pressure (PIP), and peak end expiratory pressure (PEEP). The system allows helmets to be deployed more broadly on the wards, since a single clinician can now monitor many patients simultaneously. The respiratory profile provided by the system allows clinicians to track disease progression and informs treatment decisions, including decisions about when to intubate.

We have produced and tested 50 such devices in a matter of weeks at a marginal cost of under 300 USD. Commercial monitoring devices designed for mechanical ventilation, like the Philips Respirionics NM3 monitor, offer some of the functionality of our monitoring system, but those systems are currently unavailable due in part to disruptions in the medical supply chain. In addition, the commercial alternatives are single-patient devices that cost 20 to 40 times more than our device, thus making them impractical for rapid scale-up and deployment in a pandemic setting. To facilitate local manufacture, we provide detailed parts lists, computer-aided design files, software, and instructions for assembly and testing.

## 2 Materials and methods

### 2.1 Design and construction of the device and software

The Princeton Open Ventilation Monitor device consists of a flow-sensor assembly and an interface box located at the patient’s bedside, as well as a multi-patient remote monitoring station, as shown in Fig 1. When used with a helmet for monitoring patients undergoing non-invasive ventilation, the flow-sensor assembly is inserted into the expiratory path. A schematic of the system is shown in Fig 2. Detailed information can be found in Supplementary Design and Construction of the Device and Software.

**Fig 1.**
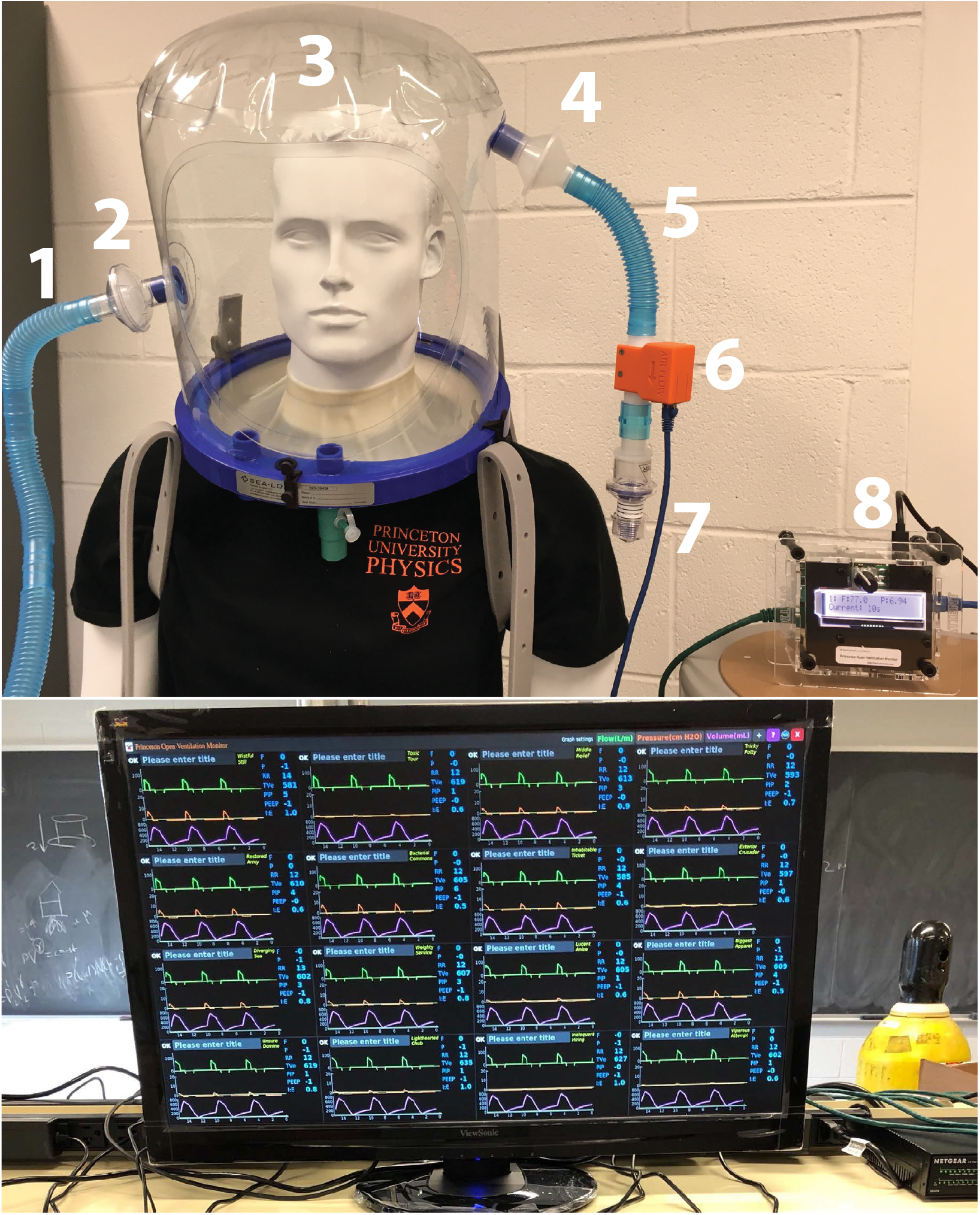
Device monitors respiratory features by measuring pressure and flow of air leaving a helmet. Top: Device is shown installed in a typical circuit. Optional CO_2_ sensor is not shown. (1) Inspiratory path. (2) Filter. (3) Patient in helmet. (4) Filter. (5) Expiratory path. (6) Flow-sensor assembly. (7) PEEP valve. (8) Interface box. The interface box reports basic respiratory information and announces audible and visual alarms if flow, pressure or respiratory rate cross clinician defined thresholds. Bottom: The remote monitoring station displays flow, pressure, and volume waveforms and clinically relevant quantities including equivalent tidal volume, respiratory rate, I:E ratio, PIP and PEEP from up to 20 devices. Here 16 devices in a test-circuit are being monitored simultaneously.

**Fig 2.**
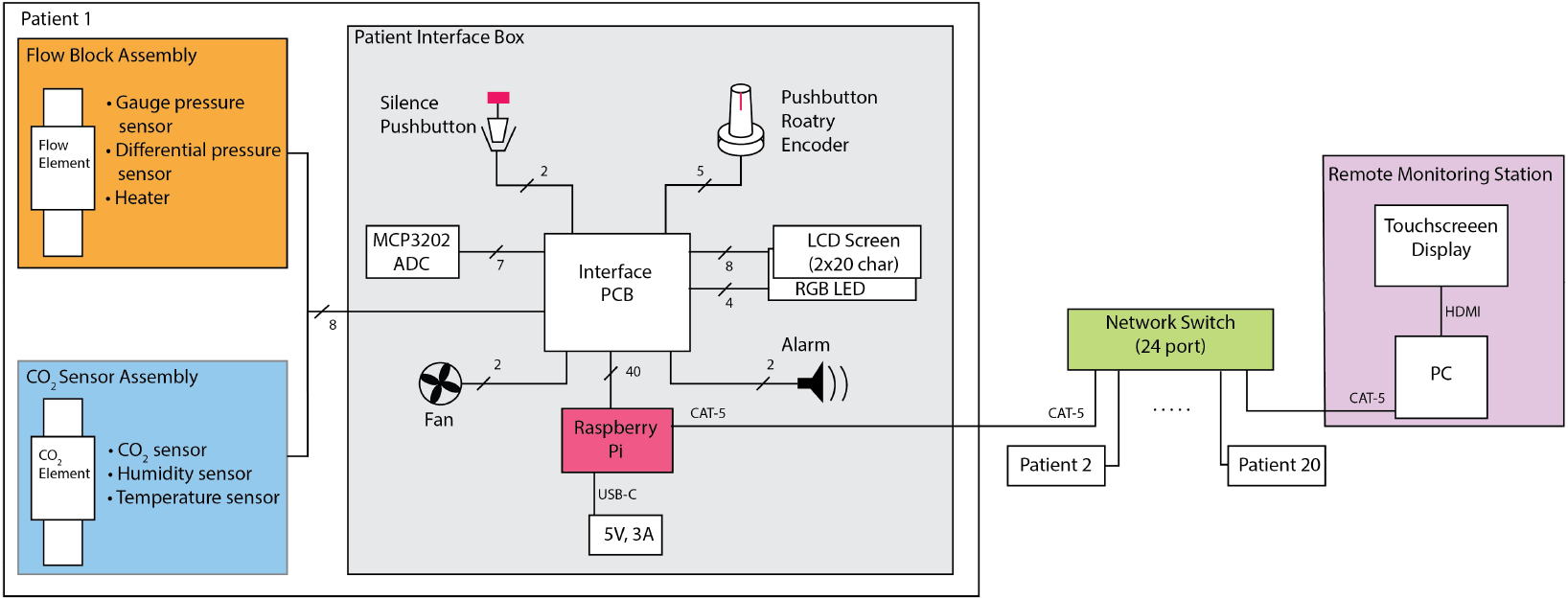
Schematic of system design.

### 2.2 Methods for testing and evaluation of the device and software

#### 2.2.1 Software testing

Software was designed with the following testing strategies in place: Static type analysis is performed to ensure basic validity of the code on every commit. Some portions of the code, such as the custom rolling window, have unit tests. A simulation and a data recording playback feature provide tools to perform integration tests for the hardware, analysis, and the clinician GUI. An alternative patient loop that runs using Qt instead of requiring the hardware rotary, screen, and silence button enables testing with the playback even without a physical device present.

#### 2.2.2 Flow-sensor assembly calibration tests

The flow-sensor assembly was calibrated by using a mass flow controller (Alicat Scientific MCR-100SLPM-D). As sketched in Fig. 3c, the inlet port of the mass flow controller (MFC) was connected to compressed air and the outlet port was connected to the flow element by a long segment (1.5 m) of medical tubing. To minimize the effect of tubing curvature in the flow calibration (see tests below), the tubing upstream of the flow element was maintained horizontal and straight, and downstream the flow element is directly connected to atmospheric pressure. Successive steps of constant flow *Q* ranging from 10 L/min to 100 L/min were imposed while the differential pressure Δ*P* at the flow sensor was measured using our device (see Fig. 3). The details of the sweep were chosen to probe both the steady-state relation between *Q* and Δ*P* and temporal dynamics during changes of applied flow. The 0 to 100 L/min flow range accessible to our MFC is clinically relevant to the helmet NIV application and corresponds to differential pressure Δ*P* readings from 0 to 150 Pa (Fig. 3b). The relation between Δ*P* (Fig. 3b, red) and *Q* (Fig. 3a, blue), averaged over many flow elements, gives the calibration in Fig. 4b. Flow elements with obvious manufacturing defects had *Q*-Δ*P* relations that deviated (see Fig. S3), and were removed from the global calibration.

**Fig 3.**
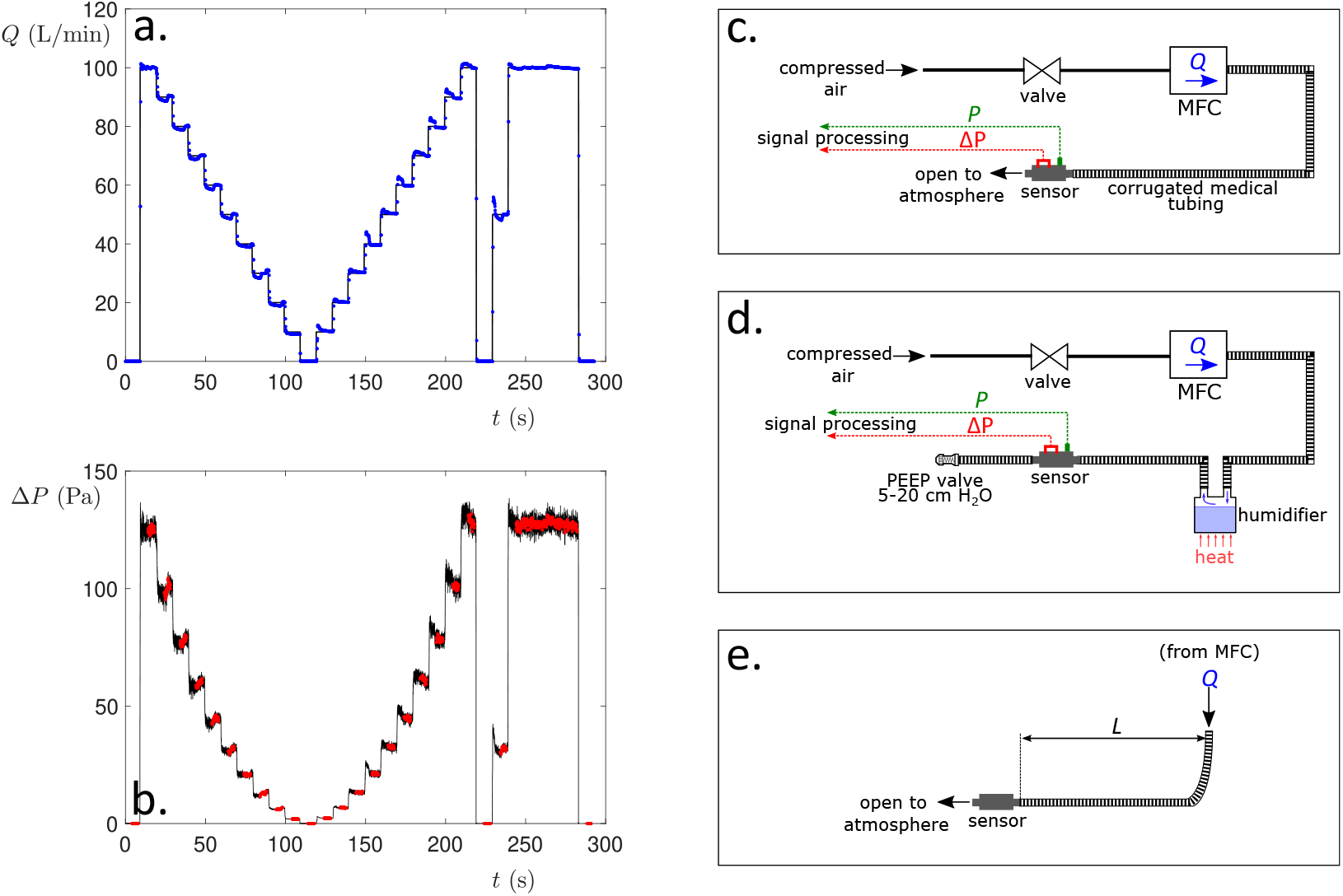
Calibration of the flow sensor. a) Standard flow sweep used to calibrate the flow sensor. Successive steps of flow *Q* (L/min) were imposed – the blue points are the flow measured in the mass flow controller (MFC). b) The differential pressure Δ*P* (Pa) as measured by our flow sensor. Black points are raw data measured by the sensor at 50 Hz. Red points are derived from the black points by eliminating the beginning and end of each step and averaging to match the 10 Hz of data from the MFC. c) Sketch of the standard configuration of our calibration setup. The MFC delivers a controlled flow *Q* to our flow element. d) More tests were performed by adding a humidifier and a PEEP valve. e) The influence of the tubing curvature was systematically tested by forcing a 90° bend at a distance *L* of the flow block.

**Fig 4.**
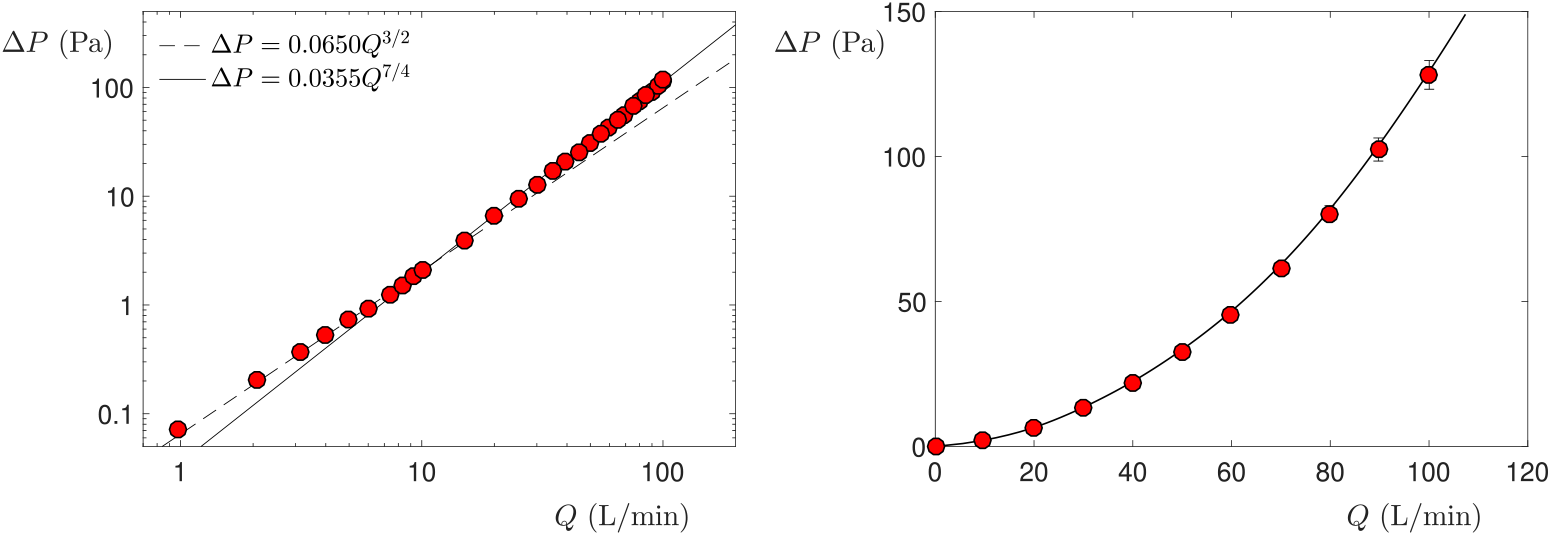
Calibration curve for measuring flow from differential pressure. Left: Measurements of Δ*P* as function of the imposed flow *Q* for a single flow element. For low flow (below 15 L/min), Δ*P* ∝ *Q*^3/2^ whereas Δ*P* ∝ *Q*^7/4^ for larger flows. Right: Averaged measurements of Δ*P* as a function of the flow *Q* on 42 flow blocks machined by commercial firm Xometry. The error bars represent standard deviations. The solid line shows the lookup table that is installed in all of our devices.

Similar calibrations were carried out in high relative humidity environment (*RH* = 100%) for several hours (Fig. 3d) to verify the resilience of the sensor to humid air and possible condensation. The effect of the presence of a PEEP valve was also tested. The effect of the curvature of the tubing was systematically estimated by running calibration tests while forcing a 90° bend at a distance *L* of the input of the flow element (Fig. 3e).

#### 2.2.3 Relationship between differential pressure and flow

Our device calculates the reported flow *Q* from a differential pressure measure Δ*P* across a portion of the flow element. A single global lookup table is used for all devices to convert from Δ*P* to *Q* (see Fig. 4). The relationship between *Q* and Δ*P* matches existing knowledge from fluid dynamics.

The geometry of the flow element and the Reynolds number determine this relation. The Darcy–Weisbach equation relates flow (here described as a flow velocity *V*) to differential pressure Δ*P*,

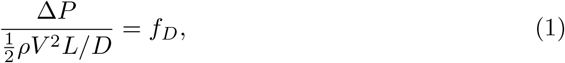

given the length *L* and diameter *D* of a pipe, and a Darcy friction factor *f*_*D*_ that depends on the Reynolds number. At 100 L/min our device operates at an intermediate Reynolds number of Re ≈ 1800, which is neither in the high (Re ≫ 2000) nor low (Re ≪ 2000) turbulence limits. (Re = *V D*/*ν*, where the kinematic viscosity of air *ν ≃* 2 *×* 10^5^ m^2^/s. At *Q* = 100 L/min, air through one of the 19 honeycomb channels of diameter *D ≃* 3 mm travels at approximately *V*_1_ *≃* 12 m/s.)

The relation between a Darcy friction factor *f*_*D*_ and Reynolds number can be looked up in standard Moody diagrams [15]. For turbulent flow in sufficiently long smooth tubes, *f* ∝ Re^*-*1*/*4^, which implies Δ*P* ∝ *Q*^7/4^. When our flow is lower, the Reynolds number drops. At a flow of 10 L/min (Re = *O*(200)), the flow in a shorter tube will have a growing boundary layer. In such laminar flow cases we expect Δ*P* ∝ *Q*^3/2^. We note that our calibration lookup table follows the Δ*P* ∝ *Q*^7/4^ for high flows and the Δ*P* ∝ *Q*^3/2^ for lower flows in agreement with expectations (Fig. 4).

#### 2.2.4 Gauge pressure test

The gauge pressure sensor was calibrated by imposing a constant pressure with a pressure controller (Fluigent MFCS-EZ). The sensor saturates at a value around 3000 Pa and exhibits a linear response, see Fig. 5.

**Fig 5.**
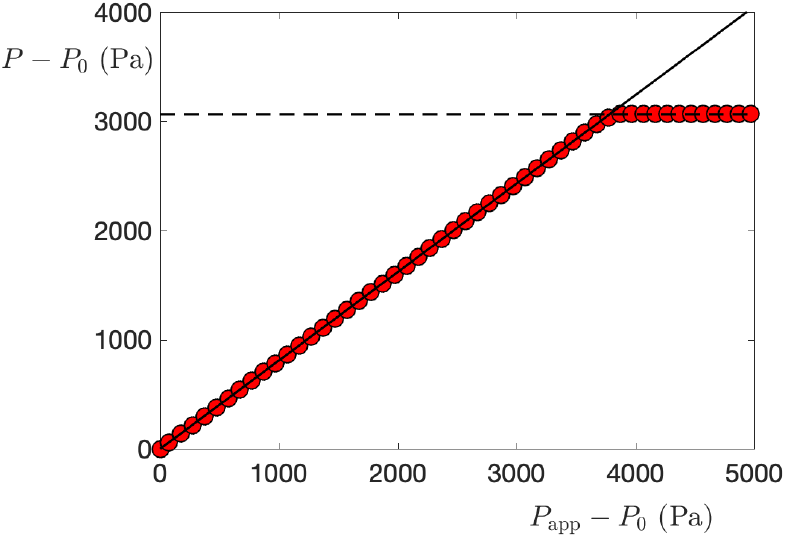
Pressure sensor calibration. The pressure *P* measured varies linearly (solid line of slope 0.81) with the pressure *P*_app_ applied with the Fluigent pressure controller and saturates above 3000 Pa.

#### 2.2.5 Methods for bench test comparison to commercial medical systems

To compare our device against commercial systems, the April 22^nd^ 2020 version of our device was tested on a commercial test lung (IngMar ASL5000) driven by a commercial ventilator (GE Avance CS2 Anaesthesia System) in series with a commercial respiratory monitoring system (Philips Respironics NM3). Our device and the commercial test monitoring system were both situated in-line on the inspiratory path of the ventilator circuit. Simultaneous recordings of pressure and flow were made from our device, the commercial monitor, and the test lung system. Ground-truth volume information was also recorded from the test lung and precisely time-aligned. Pressure information from our device was also compared to the commercial monitor (NM3). Respiratory rate, PIP and PEEP were also recorded from our device and occasionally compared to the display of the commercial ventilator. Our device was tested under a variety of ventilation modes including Pressure Control Ventilation, Volume Control Ventilation, and Synchronized Intermittent-Mandatory Ventilation with Pressure Control (SIMV PC). The test lung’s compliance was varied from 80 mL/cm-H_2_O, typical for a healthy adult, to 20 mL/cm-H_2_O which is more typical of a diseased lung. Respiratory rate was set on either the ventilator or the lung to be approximately 15 breaths per minute.

#### 2.2.6 Quality control and acceptance testing of final assembled devices

We developed a quality control protocol that could be used to test finished assembled devices prior to delivering to patients. The test demonstrates that each flow-sensor assembly’s combination of specific flow element and sensors gives measured results for flow and pressure within a set range, for example ± 10% of actual values. This was done by providing a known flow (using an Aalborg GFC47 mass flow controller) or known pressure (measured at a low flow using a water manometer sampling the flow just upstream of the flow element), recording sensor data with an interface box, analyzing the data using the fixed average calibrations discussed in Sec. 2.2.3 and 2.2.4, and comparing the recorded data to the known values.

### 2.3 Methods for human subject test

Human subject tests were conducted in accordance with IRB protocol # 12857 approved by Princeton University’s Institutional Review Board. Written consent was obtained from a healthy adult volunteer. The subject was placed in a helmet (Sea Long Model PN5404) fed by 80 L/min of medical air (AirGas) configured as shown in Fig 1 with the PEEP valve set to its lowest limit, nominally 5 cm-H_2_O. A one way valve (Teleflex Model 1644) was added to one of the helmet ports to serve as an anti-suffocation valve. The subject was in a standing position. The subject’s spontaneous breathing was recorded on our respiratory monitor device over two 20 min trials as the subject was instructed to breath normally. A water manometer in our air delivery system appeared to contribute to dynamic changes to the volume in the helmet circuit coincident with the subject’s breathing, so the manometer was inactivated during portions of the recording.

#### 2.3.1 Proximal sensor configuration

A special setup was configured to make direct comparisons of proximal sensor readings to those measured in the expiratory path of the helmet. In these experiments the human plugged their nose and breathed through a hose into the helmet, while the subject themselves remained outside the helmet. The auxiliary port of the helmet was used for external breathing, as shown in Fig. 6. The human subject breathed though a short 10-cm-long 22-mm-diameter tube with an inline flow and pressure sensor monitor, the proximal sensor. A spirometer mouthpiece and nose clip were used by the human subject.

**Fig 6.**
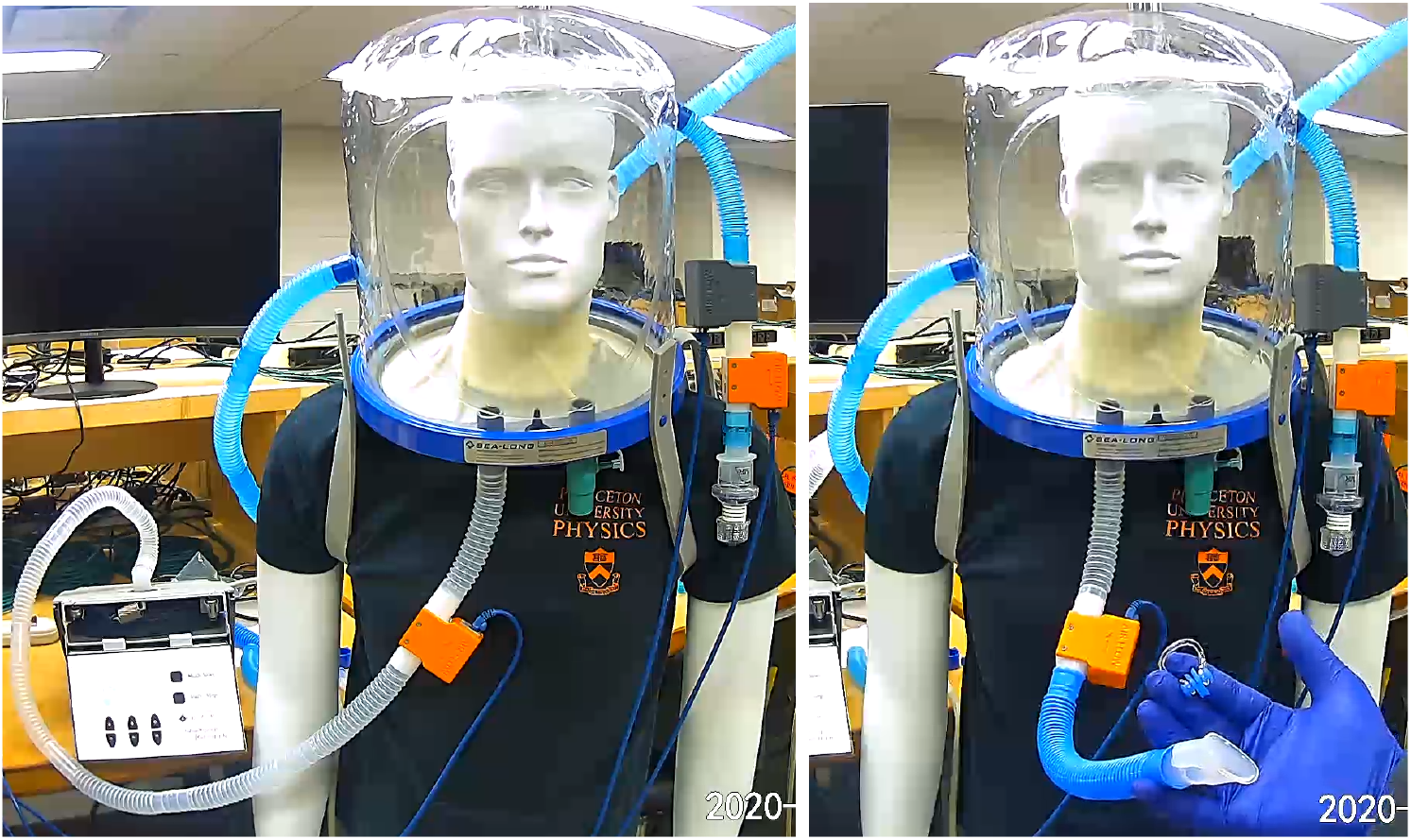
Proximal sensor configuration. The auxiliary port of the helmet is used for external breathing, shown on the left with a mechanical breather and on the right with a spirometer mouthpiece. The expiratory path is instrumented with a CO_2_ sensor in grey followed by a flow sensor in orange followed by a PEEP valve. The proximal flow sensor is placed directly inline between the spirometer mouthpiece and the auxiliary port.

Some bench tests were also performed in this configuration. In those instances, a mechanical breather instead of a human subject was connected through the inline flow sensor to the auxiliary port.

## 3 Results

### 3.1 Flow, pressure and tidal volume agree with commercial test systems during mechanical ventilation

To test our device’s ability to measures of flow and pressure generally, we placed a prototype of our device in a circuit with a commercial single-patient respiratory monitoring system (Philips NM3) on a test lung (ASL-5000) driven by a commercial mechanical ventilator. We compared recordings made by our device to those made by the test lung and the commercial monitoring system. Recordings from our device show strong agreement with the test lung for flow and volume, as shown in Figs. 7, 8 and 9.

**Fig 7.**
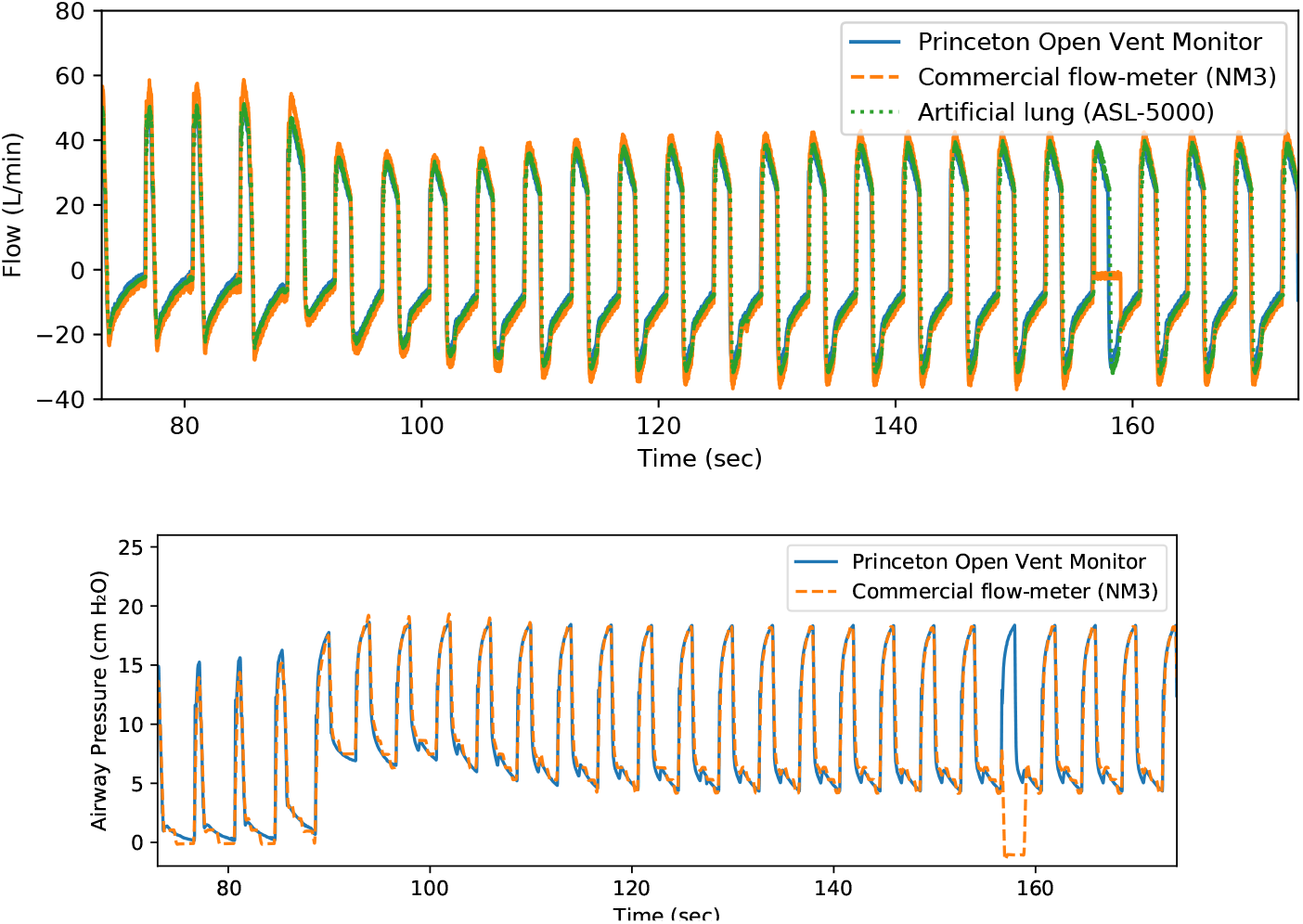
Comparison to commercial monitor. Overlay of flow and pressure measured simultaneously from the Princeton Open Vent Monitor, a commercial respiratory monitor (NM3), and an artificial lung (ASL-5000) driven by a commercial ventilator.

**Fig 8.**
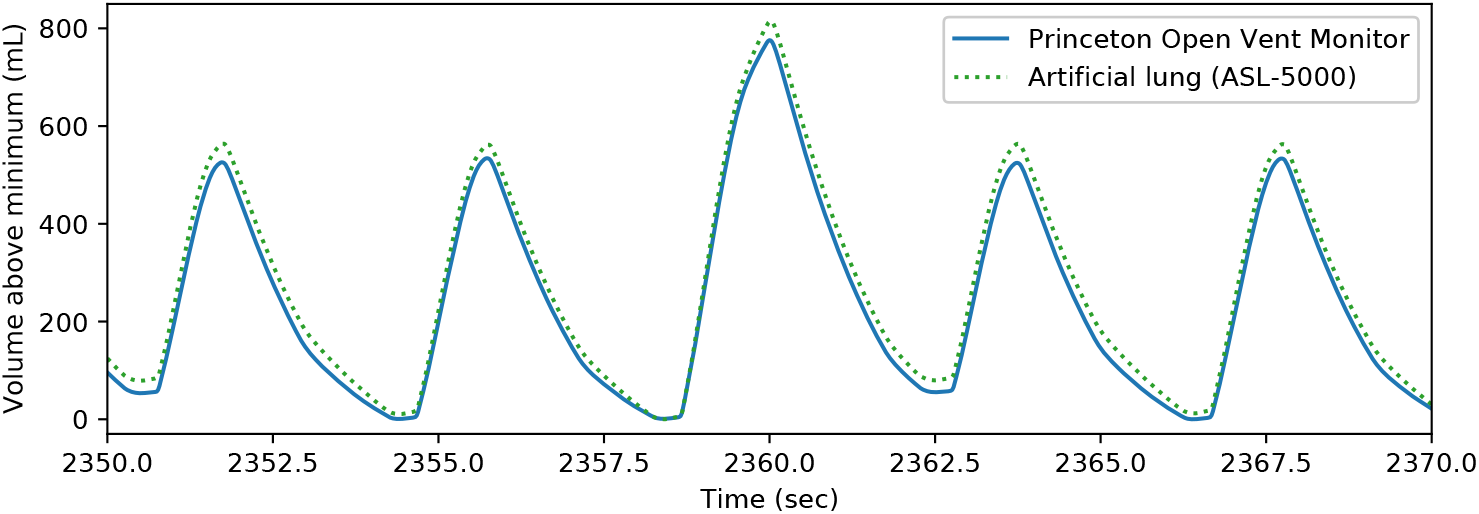
Volume comparison to test lung. Overlay of derived volume from the Princeton Open Vent Monitor (using the high-pass filter with the weaker 0.0004 Hz Butterworth critical frequency) and measured volume from an artificial lung (ASL-5000), driven by a commercial ventilator.

**Fig 9.**
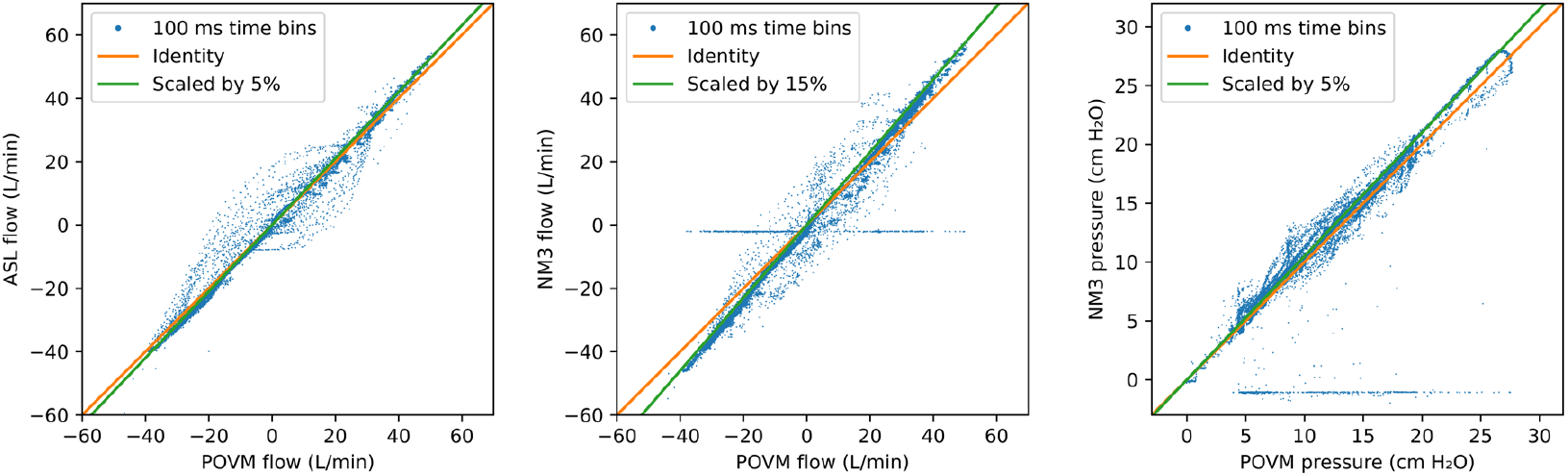
Fine comparison of Princeton Open Ventilation Monitor (POVM), the artificial lung simulator (ASL), and a commercial respiratory monitor (NM3) in matched time-bins (100 ms wide). The commercial monitor occasionally drops to zero and has discretization effects at low pressure. POVM and ASL agree in flow up to a 5% scale factor; POVM and NM3 agree in flow up to a 15% scale factor, and POVM and NM3 agree in pressure up to a 5% scale factor.

Our device’s flow measurement showed closer agreement to the test lung than the commercial monitoring system that was measuring flow simultaneously. There was a scaling factor of about 5% discrepancy between our device and the test lung’s measured flow, while the commercial monitor reported flows that differed from our device by a scaling factor of about 15% and an offset of 2 L/min. The commercial respiratory monitor also would occasionally show a flat line drift for flow or pressure during a breath, while our system captured dynamics of all breaths (see artifact at approximately 158 s in Fig. 7 and as a horizontal spread around zero in Fig. 9). This flat line (drift) from NM3 monitor is likely caused by the periodic flushing of the system to avoid condensation in the pressure lines.

In measures of pressure, our system had close agreement to the commercial respiratory monitor. We note that our system seemed to avoid quantization artifacts found in the commercial monitoring system at low pressures (see steps in pressure around 80 s in NM3 recording in Fig. 7).

We also compared derived quantities—the equivalent tidal volume, PIP, PEEP, and breathing rate—with those reported by the mechanical ventilator at the same time. Our system largely agrees with the ventilator’s own settings (see Table 1).

**Table 1.** Comparison of derived quantities. Princeton Open Vent Monitor (POVM) derived tidal volume equivalent (≈ TV), peak inspiratory (PIP) and end-expiratory (PEEP) pressure, and breathing rate (RR) compared with contemporaneous readings/settings from the ventilator and readings from the commercial monitor (NM3).

|  | POVM derivation | Ventilator reading | NM3 reading |
| --- | --- | --- | --- |
| TV | 440 mL | 500 mL (setting) | 460 mL |
| PIP | 28.0 cm H<sub>2</sub>O | 29 cm-H<sub>2</sub>O |  |
| PEEP | 4.5 cm-H<sub>2</sub>O | 5 cm-H<sub>2</sub>O |  |
| RR | 15.0 bpm | 15 bpm |  |
| I:E | 0.66 |  |  |

### 3.2 Human volunteer helmet NIV study

A healthy consenting adult volunteer was placed in a helmet and received medical air at 80 L/min while undergoing monitoring with our device. The device captured qualitatively reasonable waveforms for flow, pressure and equivalent tidal volume, as shown in Fig. 10. The measured respiratory rate matched that reported by the subject. Measured flow waveforms appear qualitatively similar to previous reports of human subjects in a helmet [12]. Recorded device data stream is available in Supplementary Data Files 1.

**Fig 10.**
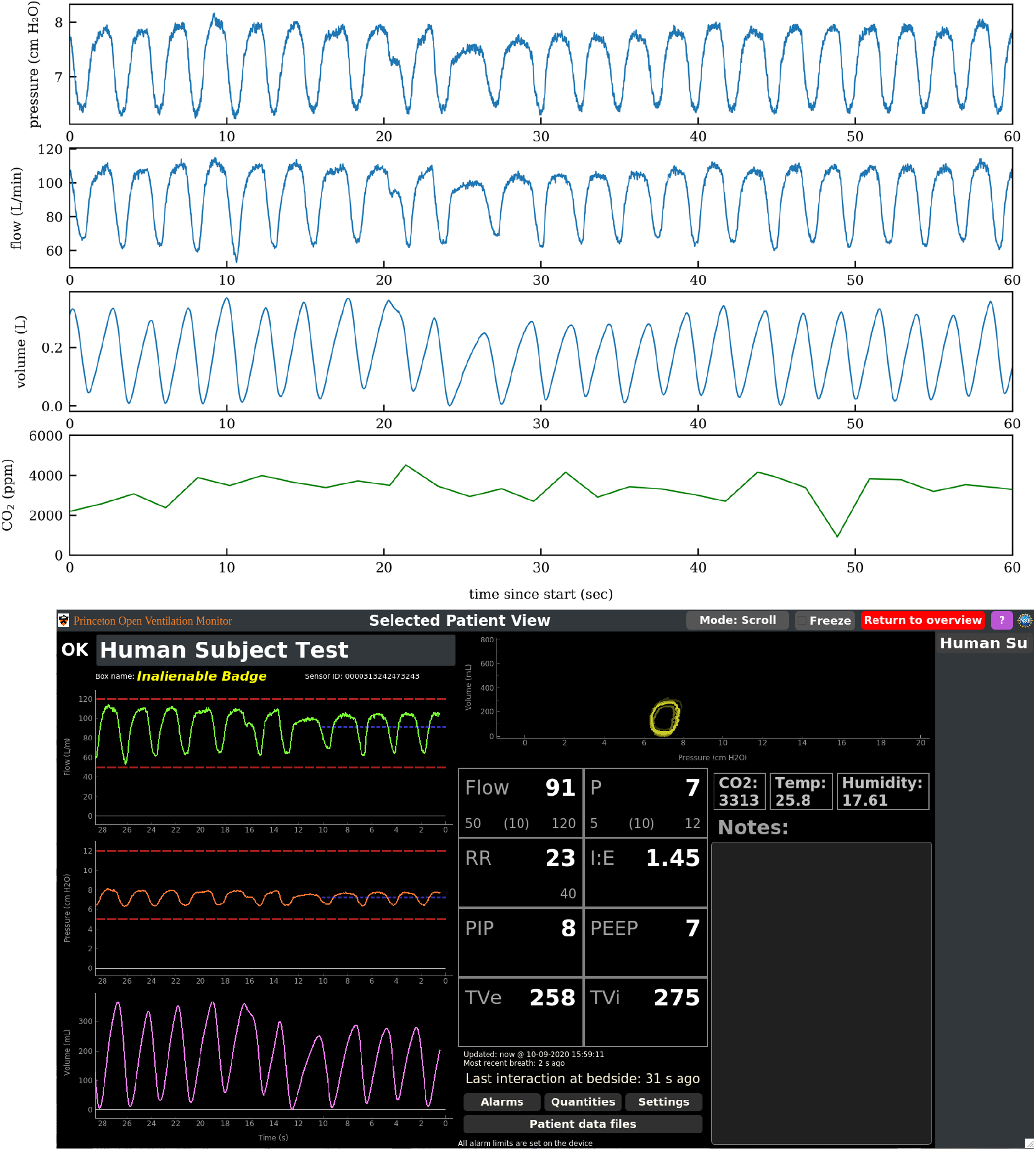
Respiratory profile of healthy human subject in helmet. Top: Pressure, flow, equivalent tidal volume and CO_2_ concentration on the expiratory path of a helmet worn by a healthy human subject are shown. Positive flow is defined to be gas leaving the helmet through the expiratory path. Bottom: Derived quantities, including tidal volume, respiratory rate, etc are visible on the remote monitoring station (shown here in “drilldown” screen).

#### 3.2.1 Comparison of tidal volume measured at helmet expiratory path to that measured proximally

A central question in the applicability of helmet monitoring for clinicians is whether the patient’s tidal volume and waveforms are modified by the pliable helmet. We had independently validated our device’s volume estimates during mechanical ventilation with the ASL-5000 artificial lung (Fig. 8 where there was no helmet in the circuit. To quantify the effect of a helmet on our device’s ability to estimate volume, we next conducted human-subject tests on volunteers wearing a helmet and directly compared simultaneous recordings at the expiratory path to measurements proximal to the patient. Here the proximal sensor served as “truth” in the sense that it provides an ideal respiratory monitoring data with direct monitoring of flows, pressures and computed tidal volumes from the human subject that should be unaffected by the helmet. The human stood outside of the helmet, plugged their nose and breathed into the helmet through tubing connected to the helmet’s auxiliary port. Readings were made simultaneously at the expiratory path. From the perspective of the expiratory path, the airflow behaves as if the patient were inside the helmet. The comparison of the proximal sensor (auxport) and expiratory pressures, flows and estimated tidal volumes are shown in Fig. 11. Leaks were monitored through an inspiratory-to-expiratory path average flow rate comparison. Leaks were observed to be as large as 1 to 4 LPM, depending on the configuration and quality of the neck seal. Leaks impact the tidal volume baseline calculation and the dynamical response of the volume modulation of the helmet described in more detail below.

**Fig 11.**
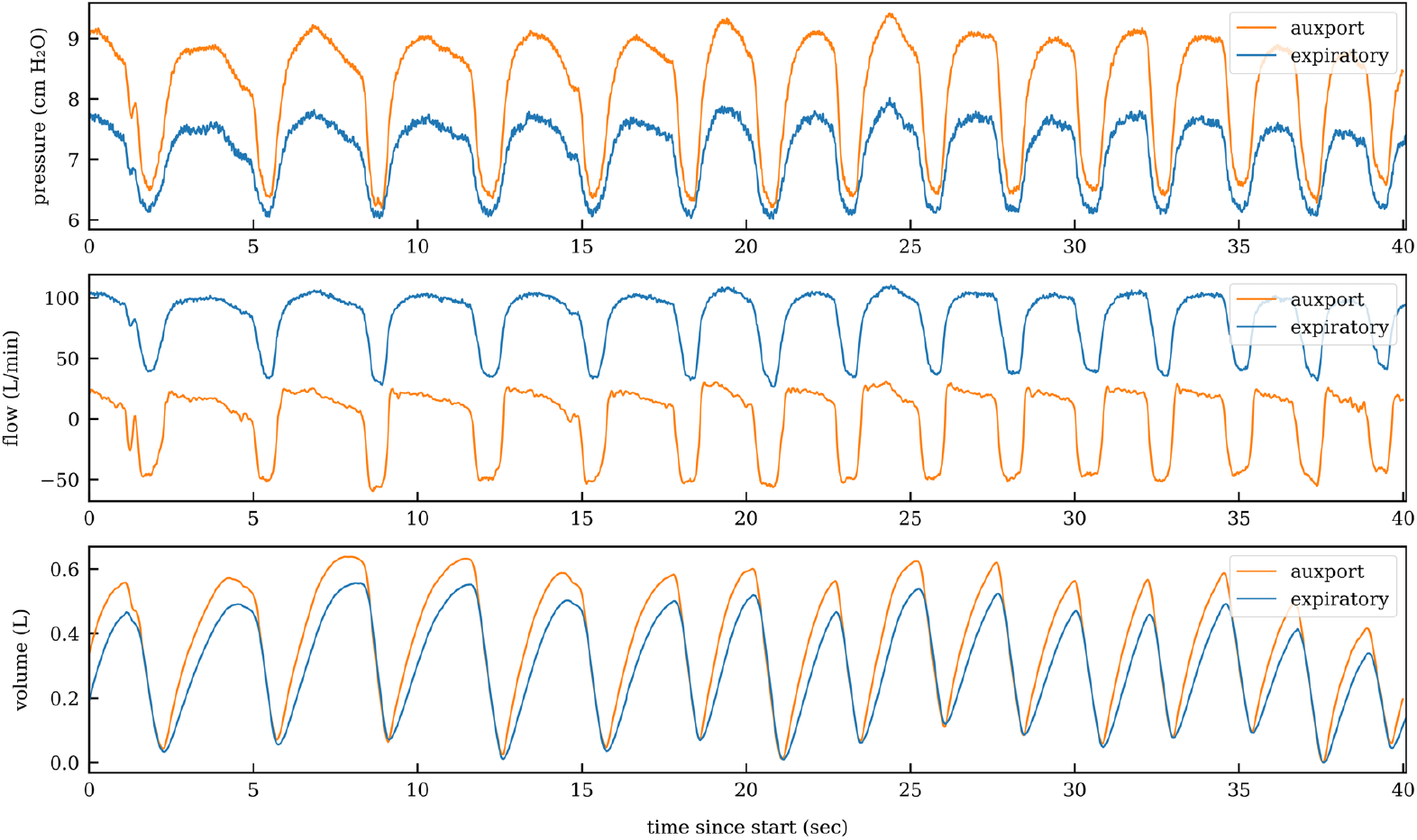
Proximal vs expiratory sensor comparison, with human subject. The respiratory waveforms recorded directly by a proximal sensor (auxport) are compared to measurements made at the expiratory path. A slight pressure increase is seen by the human subject due to breathing through the proximal sensor and tube through the auxiliary port. Expiratory path’s measure of flow appears slightly smoother compared to the proximal sensor, and the expiratory path measure of volume consistently underestimates that of the proximal sensor.

By comparing the proximal to expiratory path, several observations can be made that reveal effects of the helmet system on one’s ability to measure respiratory features. The first is a discrepancy in pressure. In the top plot of Fig. 11, there is an additional pressure on the human subject side of the flow block of the proximal sensor of approximately 1 cm-H_2_O. This pressure increase is attributed to the flow restriction inherent in the flow block and the short length of 22-mm tube to access the auxiliary port. The second is an expected change in baseline flow. From the middle plot of Fig. 11, the expiratory waveforms are offset from zero by the flushing rate of medical air through the helmet. In contrast, the average flow in and out of the proximal sensor is zero, as this is only connected to the human lung (nose clips are worn by the human subject).

Third is is an observation that the expiratory waveforms appear to be slightly smoother than the proximal ones and in certain cases when a sharp feature appears in the proximal sensor, the expiratory path response is slower. To further explore the slow down of response to the respiratory waveforms, a mechanical breather (QuickLung Breather from IngMar Medical) was connected through the auxiliary port in the same configuration as the proximal sensor human subject test. The mechanical breather was configured to generate sharp respiratory waveforms (compliance, *C*, of 50 mL/cm-H_2_O and a resistance, *R*_*p*_, of 5 cm-H_2_O/L/s). A comparison of the proximal sensor and expiratory sensor at an 80 L/min average flow rate through the Sea-Long helmet are shown in Fig. 12. The slow down in response as measured via the expiratory path is visible in the middle plot. Similar effects to those in Fig. 11 are observed in the pressure and estimated tidal volumes. Additional measurements were made at different settings of the mechanical breather, including resistance at 20 cm-H_2_O/L/s and 50 cm-H_2_O/L/s. At 50 cm-H_2_O/L/s (*R*_*p*_*C* = 2 · 5 s), many of the sharp features in the mechanical breather waveforms are removed and the expiratory and proximal sensor waveforms agree. The apparently longer resistance-compliance time is an artifact of the helmet monitoring.

**Fig 12.**
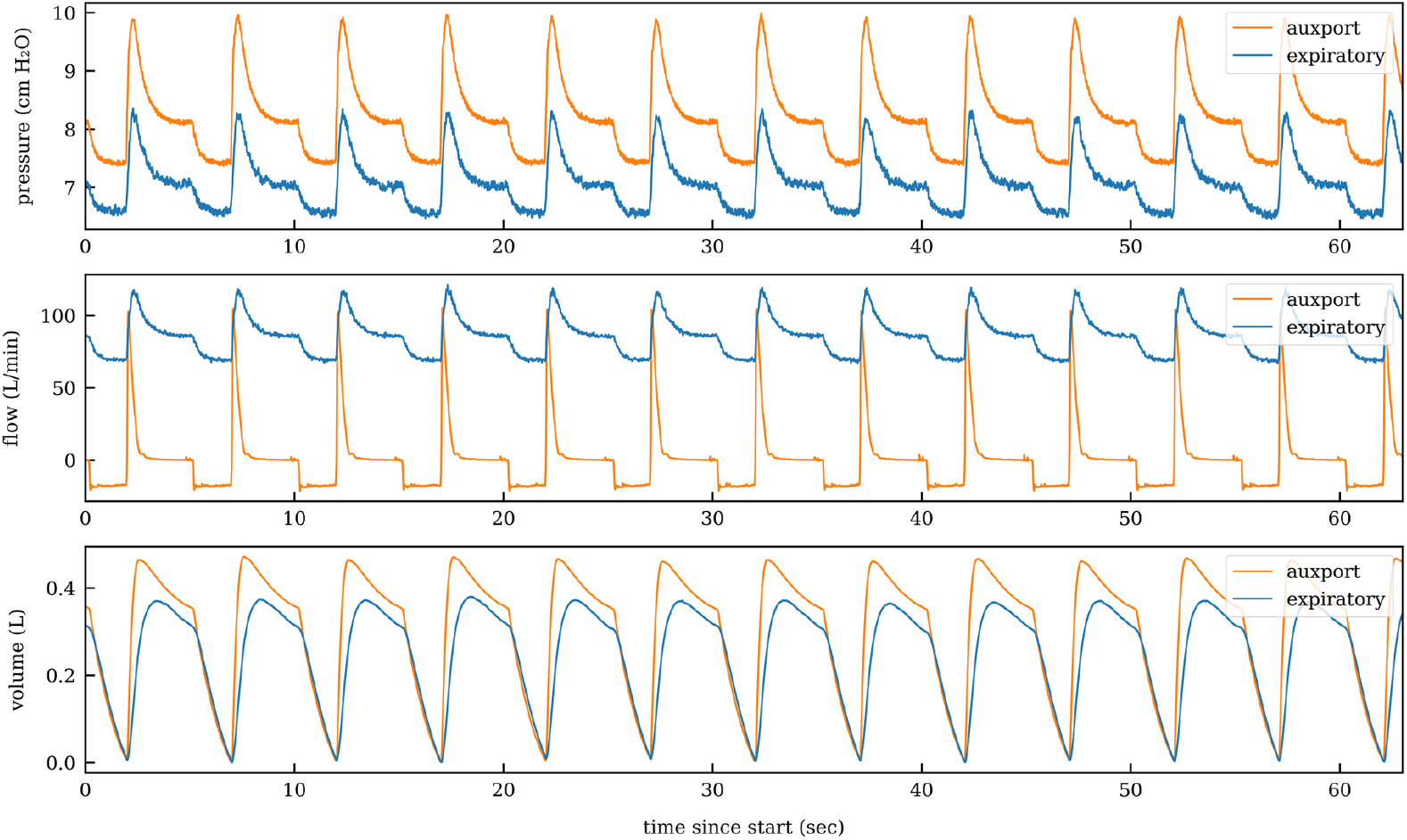
Proximal vs expiratory sensor comparison, with mechanical breather. The IngMar Medical QuickLung Breather is connected in the proximal sensor configuration to the auxiliary port of the Seal-Long helmet and monitored simultaneously through the expiratory path of the helmet. The mechanical breather is set to a compliance of 50 mL/cm-H_2_O and a resistance of 5 cm-H_2_O/L/s to generate sharp respiratory waveforms at 12 pbm. The slow down in response as measured via the expiratory path is visible.

Finally, on the bottom plot of Fig. 11, the equivalent tidal volumes underestimates the expiratory path tidal volumes by roughly 20%. The accuracy of the equivalent tidal volume estimates from the expiratory path in the proximal sensor configuration was further evaluated with a calibrated 1-L volume syringe (Aspire 1 Liter Volume Calibration Syringe) that was manually operated. The data from this comparison is shown in Fig. 13 for helmet flow rates of 50 L/min and 80 L/min. The underestimation of the equivalent tidal volumes is up to -20% at high flow rate.

**Fig 13.**
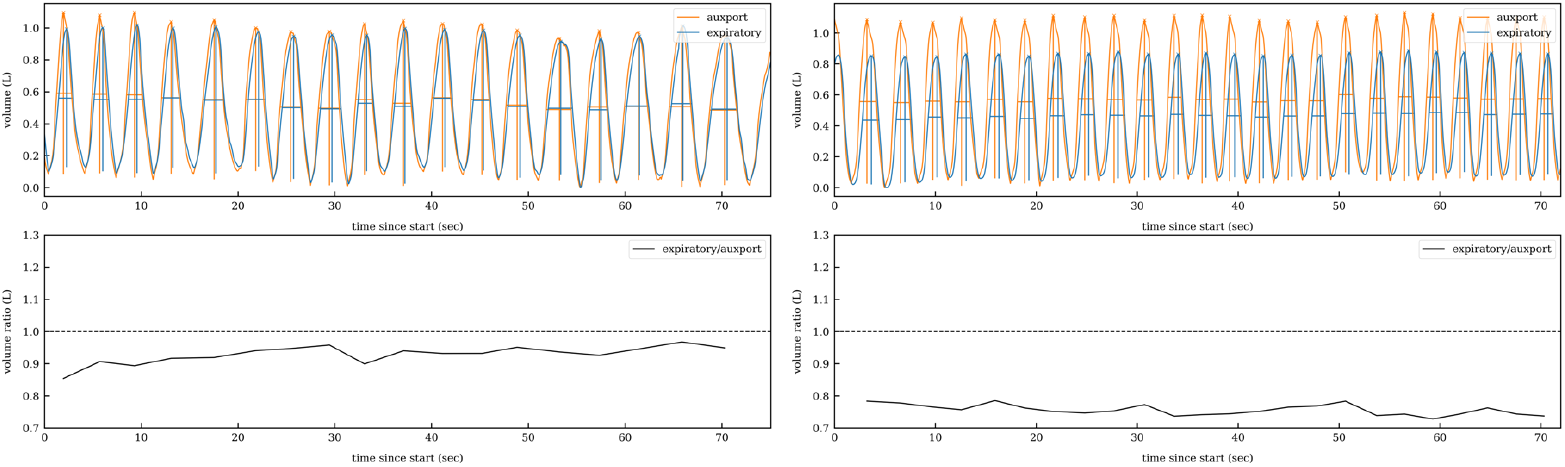
Proximal vs expiratoy sensor comparison, with 1L calibrated syringe. The Aspire 1 Liter Volume Calibration Syringe is operated manually in the proximal sensor configuration. The left plot is for a 50 L/min flow rate through the helmet and the right plot is for 80 L/min. The bottom plots are the corresponding ratios of the peak heights, where the peak heights are the equivalent tidal volume estimates. The underestimation of the equivalent tidal volumes by the expiratory sensor is up to 20% at high flow rate compared to the proximal sensor (auxport).

The origin of this deviation is inherent and universal to the helmet continuous positive applied pressure setup and therefore of upmost important to clinicians that make use of these systems to infer tidal volume information about the patient. The physics origin of this deviation in tidal volume estimation is of similar origin to the slow down in response presented in Fig. 12. The pressure differential required to increase the flow rate through the expiratory path grows non-linearly as Δ*P* ∝ *Q*^7/4^ for large flows, as shown in Fig. 4. Therefore, a fraction of the exhaled volume at high flow rate will preferentially inflate the pliable helmet volume. Similarly, the inhaled volume will tend to contract a fraction of the helmet volume. Slight changes in the helmet volume (inflating and contracting) at high flow rate through the helmet are observed and can be exacerbated by deep, rapid, breathing of large tidal volumes.

It is therefore vital that helmet monitoring systems provide a tidal volume estimation that accounts for the inflation and deflation of the helmet volume and leaks, as the expiratory flow does not directly measure the volume change within the helmet. The bias toward underestimation of the equivalent tidal volumes at high flow rate can be removed with a correction factor that follows the same functional form of Δ*P* ∝ *Q*^7/4^ for large flows. After this flow-rate dependent correction, the deviations on tidal volumes are within *±*5%. We have further added this correction as an option into the system so that a clinician can see the corrected value.

#### 3.2.2 CO_2_ in the helmet

The CO_2_ concentration in the helmet was monitored with the CO_2_ sensor block at the expiratory path, Fig. 14. When Medical Air gas was flowed through the helmet at approximately 80 L/min without any human, the CO_2_ concentration was measured to be 412*±*92 ppm, in agreement with expected CO_2_ levels for fresh air.

**Fig 14.**
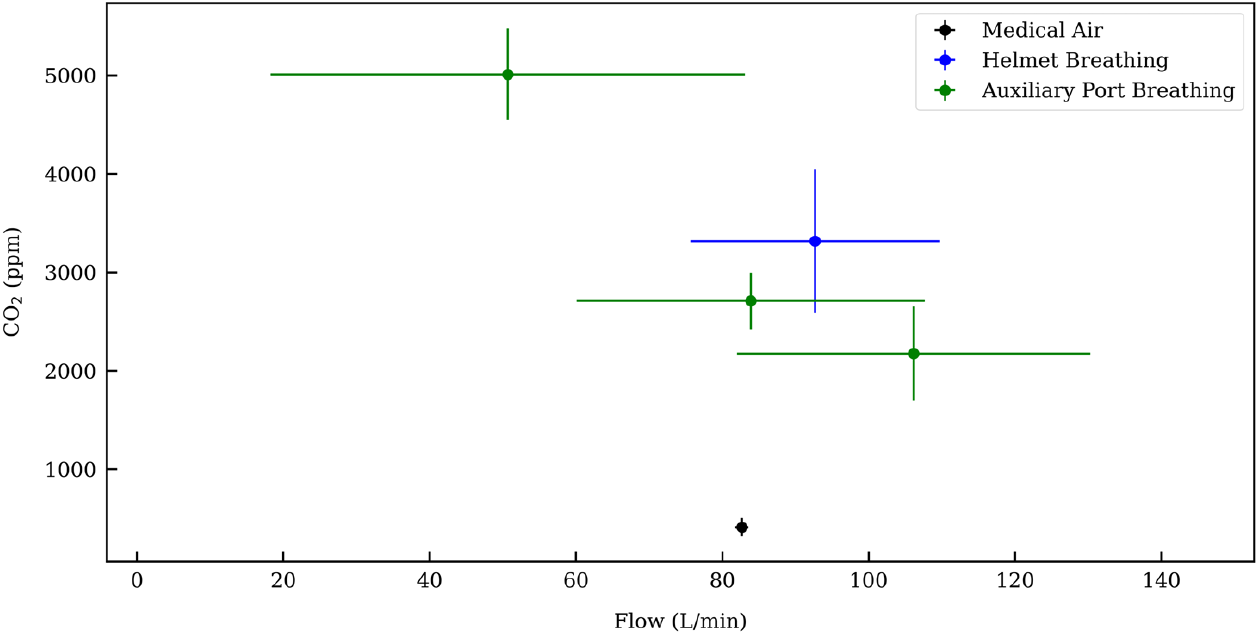
CO_2_ concentration in the helmet. The CO_2_ concentration in the helmet as a function of flow rate reproduces previous findings [12]. The horizontal uncertainties are the RMS values of the flow measurements coming from the breathing of the human subject. The vertical uncertainties are the RMS values of the CO_2_ concentration measurements in the expiratory path, indicating incomplete mixing of the exhaled breaths with the helmet flow. The steady-flow medical air measurement through the helmet shows that the intrinsic CO_2_ concentration measurements are not a large contribution to the uncertainties compared to breathing.

CO_2_ concentrations at the expiratory port were than measured when a human, standing external to the helmet, breathed into the helmet through the auxiliary port (Fig. 6). Measurements were made during three different flow rates, ranging from 50 L/min to over 100 L/min, Fig. 14 “Auxiliary Port Breathing”. The CO_2_ concentration measured in the helmet as a function of flow rate reproduces previous findings [12].

CO_2_ concentration was also measured at the expiratory port when the human test subject breathed from within the helmet (“Helmet Breathing” in Fig. 14, flow rate of approximately 93 L/min). At the nominal average flow rate of 80 L/min, the CO_2_ concentration was measured to be approximately 3000 ppm. At 50 L/min, the average CO_2_ concentration grows to 5000 ppm. The CO_2_ concentration, measured every 2 seconds, fluctuates significantly. In the human subject test at an average flow rate of 93 L/min, the mean value of the CO_2_ concentration was 3317 ppm and the root-mean-squared fluctuations of measurements were 732 ppm. Some of these fluctuations are due in part to the changes in flow rate that occur from subject breathing. But, even more so, we suspect the large fluctuations arise from incomplete mixing of the subject’s exhaled CO_2_ with the air in the helmet before it reaches the sensors in the expiratory path.

Fluctuations in measured CO_2_ makes it difficult to use CO_2_ to reliably monitor fast time-varying changes to CO_2_ dynamics. In contrast, flow and pressure can be monitored quickly, and they also correlate well with average CO_2_ concentration.

### 3.3 External sources of variability

We tested the influence of relative humidity on our sensors, Fig. 3(d). In bench tests, we challenged our device by measuring 20 hours of continuous flow of saturated humid air (*RH* = 100%). Both sensors performed even in the presence of significant condensation in the flow element, which suggests that our heater circuit, designed to counter condensation, functions effectively. In tests with variable humidity, we observed that the flow *Q* increased slightly by less than 3% while the humidity rose from 5 to 100%.

Finally, the presence of curvature in tubing has been reported to affect measures of downstream flow [16, 17]. This is relevant because our device may dangle off the helmet at an angle, as can be seen in Fig. 1 and Fig. 6. We introduced a 90° bend into the tubing a variable distance *L* upstream of our device and observed the change in our device’s reported flow *Q* under a constant imposed flow of 100 L/min, Fig. 3(e) and Fig. 15. We observed a systematic increase in the flow *Q* reported by our device for bends less than 30 cm upstream of our device. The largest deviations from the imposed flow occurred when the bend was closest to the device (5% for *L* = 5 cm).

**Fig 15.**
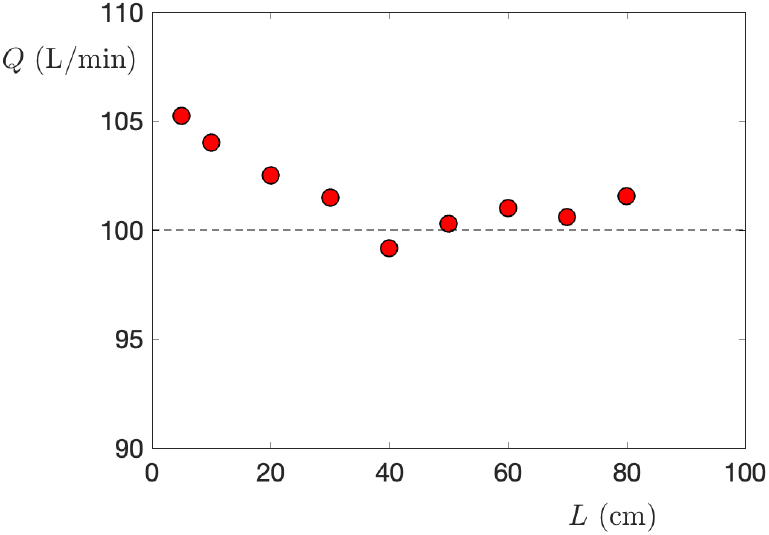
Effect of bent tubing. Flow *Q* measured by our flow sensor in the presence of a 90° bend positioned a distance *L* upstream of the flow block. A 100 L/min flow was imposed by a Mass Flow Controller.

### 3.4 Results of production run

We completed a production run of 50 devices. Producing devices at scale posed additional challenges regarding calibration and quality control. In particular manufacturing defects in the flow element altered the relationship between the measured pressure difference Δ*P* and the flow *Q* across the device, as shown in Fig. S3. Initial determination of Δ*P* vs. *Q* for each flow element with a fixed sensor array allowed us to identify and remove defective devices, and eliminate outliers from the determination of the average calibration. After each flow element is integrated into its final flow-sensor assembly, it will undergo the acceptance testing described in Sec. 2.2.6. An example test result is shown in Fig. 16. A production yield of 88% was achieved on the flow-sensor assemblies after requiring the calibrations to stay within a 10% band of the nominal calibration above 15 L/min and above 5 cm-H_2_O as a quality control measure.

**Fig 16.**
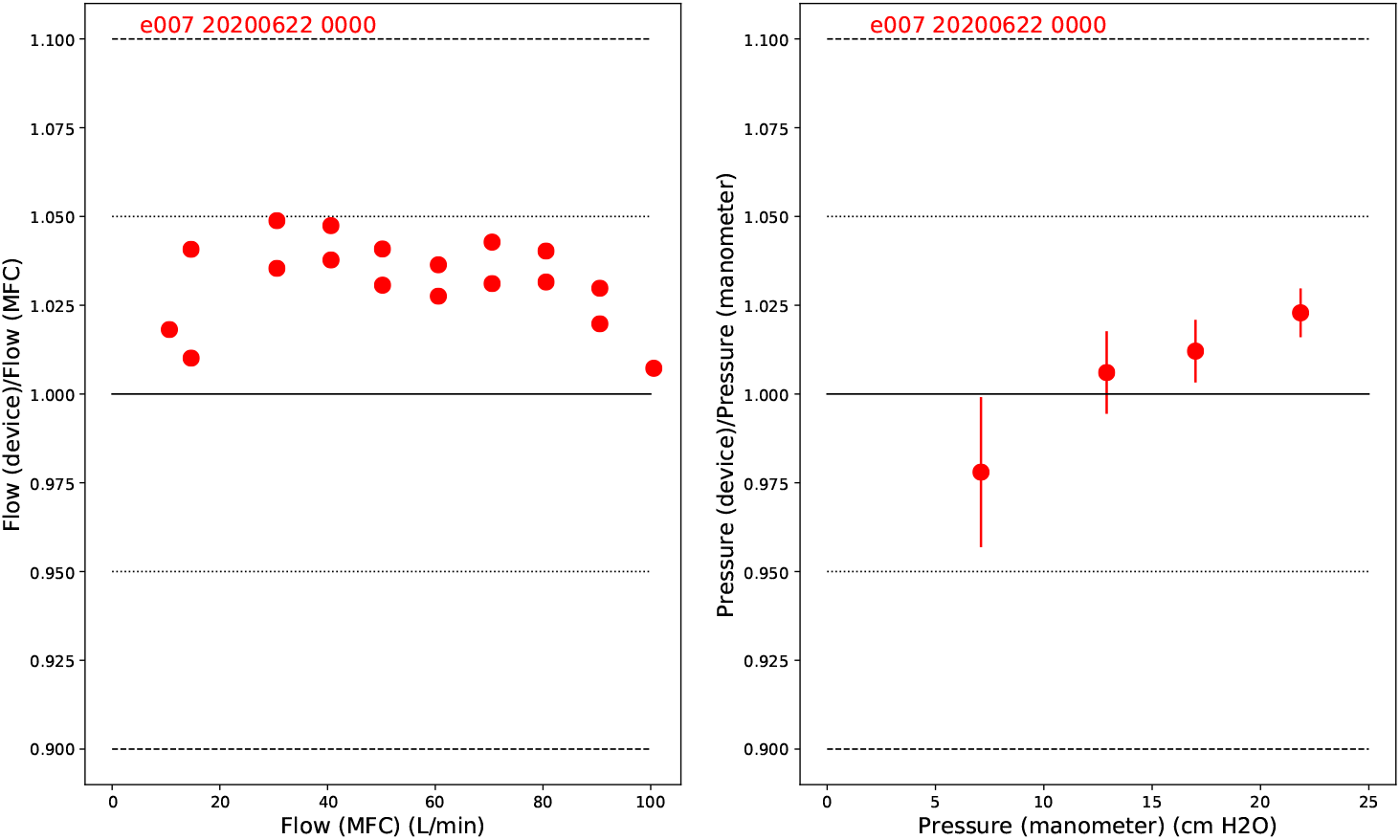
Results of acceptance testing of a final flow-sensor assembly. Left: comparison of calibrated flow reported by the device under a test to flow set by a Mass Flow Controller. Right: comparison of calibrated pressure reported by the device under test to pressure measured with a water manometer. Red text uniquely identifies the flow-sensor assembly.

## 4 Discussion and Conclusion

We designed and built an inexpensive respiratory monitoring device from conception to production in a six week period coincident with the initial peak of the COVID-19 pandemic in our region. In bench tests against a test lung undergoing mechanical ventilation, our device performed similarly to a single-patient commercial monitoring system that is currently unavailable. Measurements of a healthy volunteer undergoing helmet NIV taken by our device are reasonable and match prior reports. We quantified the effect of the helmet on estimates of tidal volume by directly comparing simultaneous measurements obtained at the expiratory path to ground-truth measurements proximal to the human-subject. Expiratory path flow and pressure monitoring tracked proximal sensor readings. At high flow rates, however, the tidal volumes reported by our device from measurements on the expiratory path underestimated tidal volumes derived from the proximal sensors by up to -20%. The deviations on tidal volumes after a flow-rate dependent calibration procedure brought this discrepancy to within only ± 5%.

Our device has the potential to improve patient outcomes and to mitigate well-documented safety concerns regarding CO_2_ re-breathing that have prevented the helmet’s widespread adoption. Our device also introduces the ability to monitor many patients simultaneously. Importantly, our device uses off-the-shelf components that have shown themselves to be robust to supply chain disruptions. The device is well suited for local manufacture during a pandemic.

## Supporting information

Supplementary Design File 1

Supplementary Design File 3

Supplementary Design File 2

Supplementary Git Repository

## Data Availability

Relevant human subject data is included as supplementary material. All design files are included as supplementary material. Links to software repositories are included in the manuscript.

## 5 Acknowledgements

This project is supported by Princeton University, including generous support by the Provost’s Office, and by National Science Foundation grants OAC-1836650, PHY-2031509 and IOS-1845137. Any opinions, findings, conclusions or recommendations expressed in this material are those of the authors and do not necessarily reflect the views of the National Science Foundation or Princeton University.

We acknowledge contributions from all members of the Princeton Open Ventilation Monitor Collaboration, including Lauren Callahan, Matthew Creamer, Sophie Dvali, Alexander E. Gaillard, Alex Glaser, Bert G. Harrop, Darryl Johnson, Julienne M. LaChance, Theodore H. Lewis III, Martina Macakova, Mala Murthy, Jonathan Prevost, P. Dylan Rich, William R. Sands and Lisa Scalice. We also acknowledge Glenn Atkinson, Matt Komor, Patrick Bradshaw from the Jadwin machine shop.

We would like to thank Dr. Ronah Harris for shooting and editing the supplementary video. We are grateful for productive conversations with Dr. Jacob Brenner, Dr. James Weimer, Dr. Maurizio Cereda, Dr. Barry Fuchs, Mike Frazer and Victoria Berehnolz all of Penn Health Tech. We thank Sebastian Seung and his volunteers for feedback. We thank Theodor Brasoveanu and his students at the Princeton Day School for supplies. We thank Meghan Testerman of Princeton University Libraries for assistance.

## 6 Author contributions

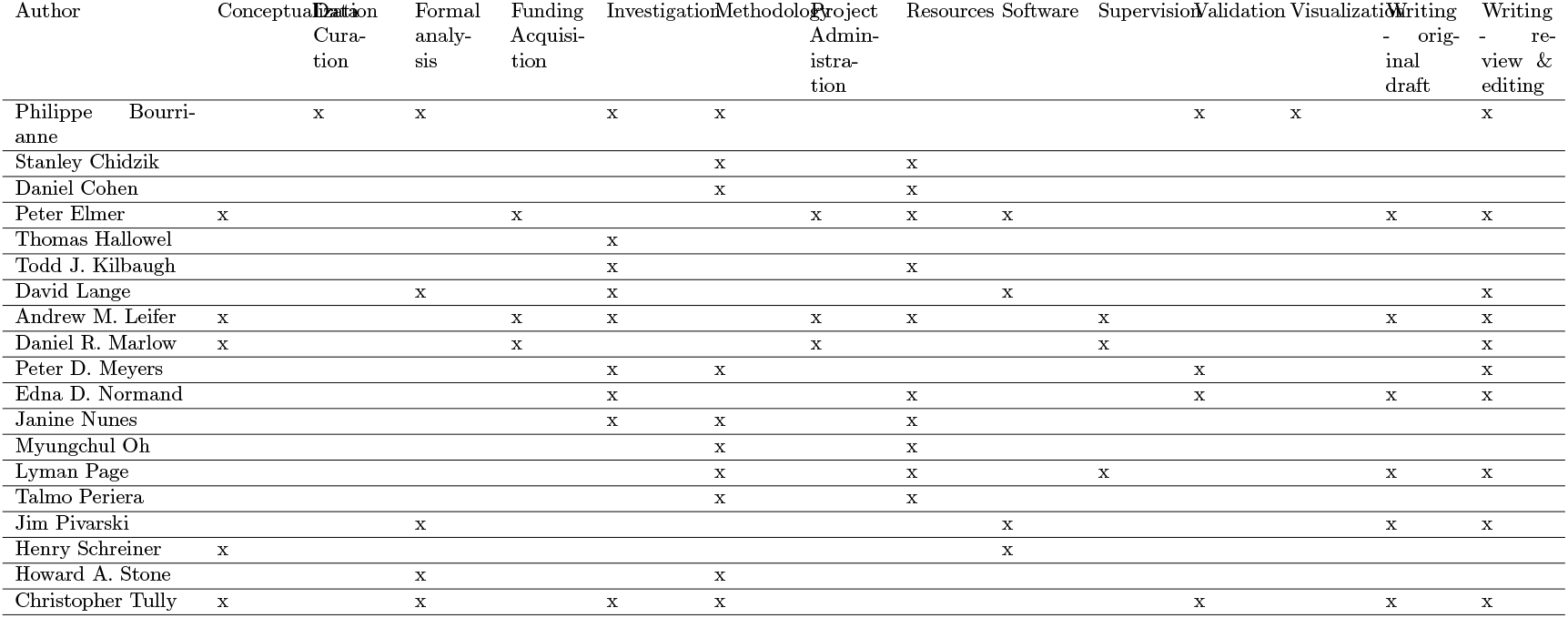

## 8 Supporting information

**Supplementary Design and Construction of the Device and Software**.

### 8.1 Differential pressure sensor

The flow meter consists of a differential pressure sensor that measures the pressure drop across a flow channel. This pressure drop is then used to calculate flow through the channel. The differential pressure sensor was chosen to be capable of measuring relatively small pressure drops (± 500 Pa) with good dynamic range. Stable operation in a humid environment was also a key consideration, as were cost and availability. The device selected was the Sensirion SDP31 differential pressure sensor. This part works by diverting a small portion of the flow and measuring the change in temperature as the gas passes over a small internal heating element. The temperature change in the gas as it flows through the sensor is related to the mass flow rate of the diverted flow stream, which in turn is related to the pressure difference across the sense ports. Readings from the SDP31 are routed to the controller over an I2C bus at a rate of 120 readings per second.

The SDP31 flow sensor, which comes in a surface mount package, is mounted to a small printed circuit board (PCB, see Supplementary Design File 1) that is fixed to the flow block as shown in Fig. S1. To prevent condensation from forming in the narrow flow-sensor channel, a heater has been added. Heat is generated in a TIP32 transistor housed in a TO-220 package. The TIP32 is mounted on the opposite side of the printed circuit board from the SDP31. Operating the transistor at a power of less than 1 W suffices to raise the temperature in the SDP31 sensor to 45 °C, which is above the dew point of the air in the system. The operating temperature is regulated using a pulse-width modulation signal from the readout controller.

### 8.2 Gauge pressure sensor

A second sensor, the MP3V5004G from Freescale Semicondutor, is included to measure the gauge pressure in the flow block. The MP3V5004G measures pressures in the range 0 < *p* < 3920 Pa (approximately 0 to 40 cm H_2_O) and provides an analog output in the range 0·6 < *V* < 3·0 V. The analog output is digitized using a Microchip MCP3202 12-bit analog to digital converter. PCB layout and schematics are included in Supplementary Design File 1.

### 8.3 Flow-sensor assembly enclosure

A 3D printed enclosure houses an RJ-45 jack and surrounds the PCBs and flow element. The enclosure was printed from PLA by a commercial fabrication house (3DGence), see Supplementary Design File 2.

### 8.4 Flow element design

The flow element, shown in Fig. S2, is a 19-mm-diameter tube, partially obstructed by a 10 mm thick wall perforated by nineteen 3 · 175-mm-diameter channels arranged in a hexagonal pattern. One of the channels is partially obstructed and is used to guide a small fraction of the air flow through the flow sensor. The obstruction is only partial, as a portion of the air flow can directly cross the central wall through a small aperture. We tuned the diameter of this aperture in order to match the dynamic range of the sensor to the typical flow rates to be measured. We found that a 1 59-mm-diameter aperture is well adapted to a 500 Pa dynamic range differential pressure sensor.

**Fig S1.**
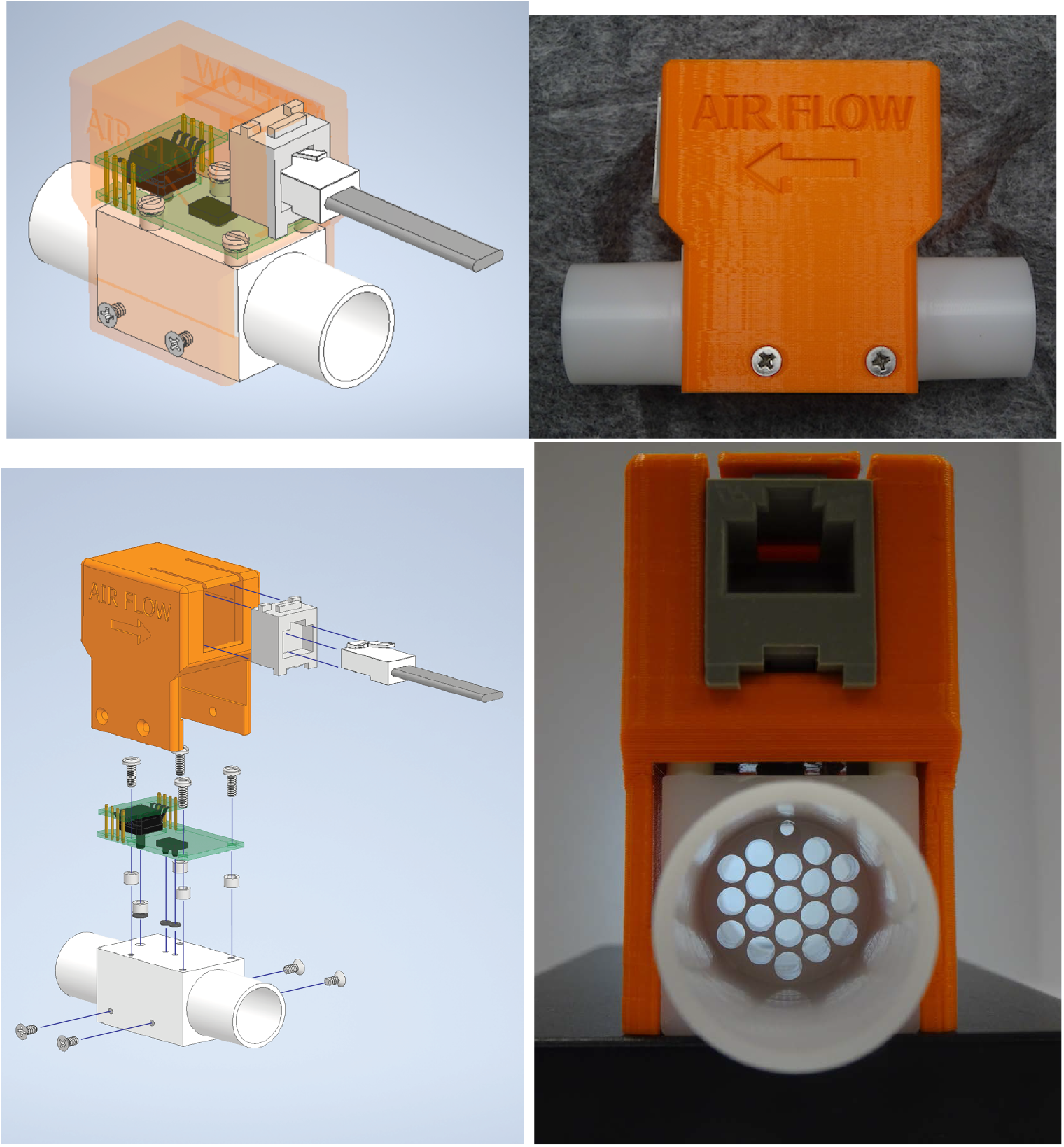
Flow-sensor assembly. A flow and pressure sensor are exposed to the flow element. The electronic sensor elements are mounted on circuit boards that connect to an external RaspberryPi controller via a standard RJ-45 jack.

**Fig S2.**
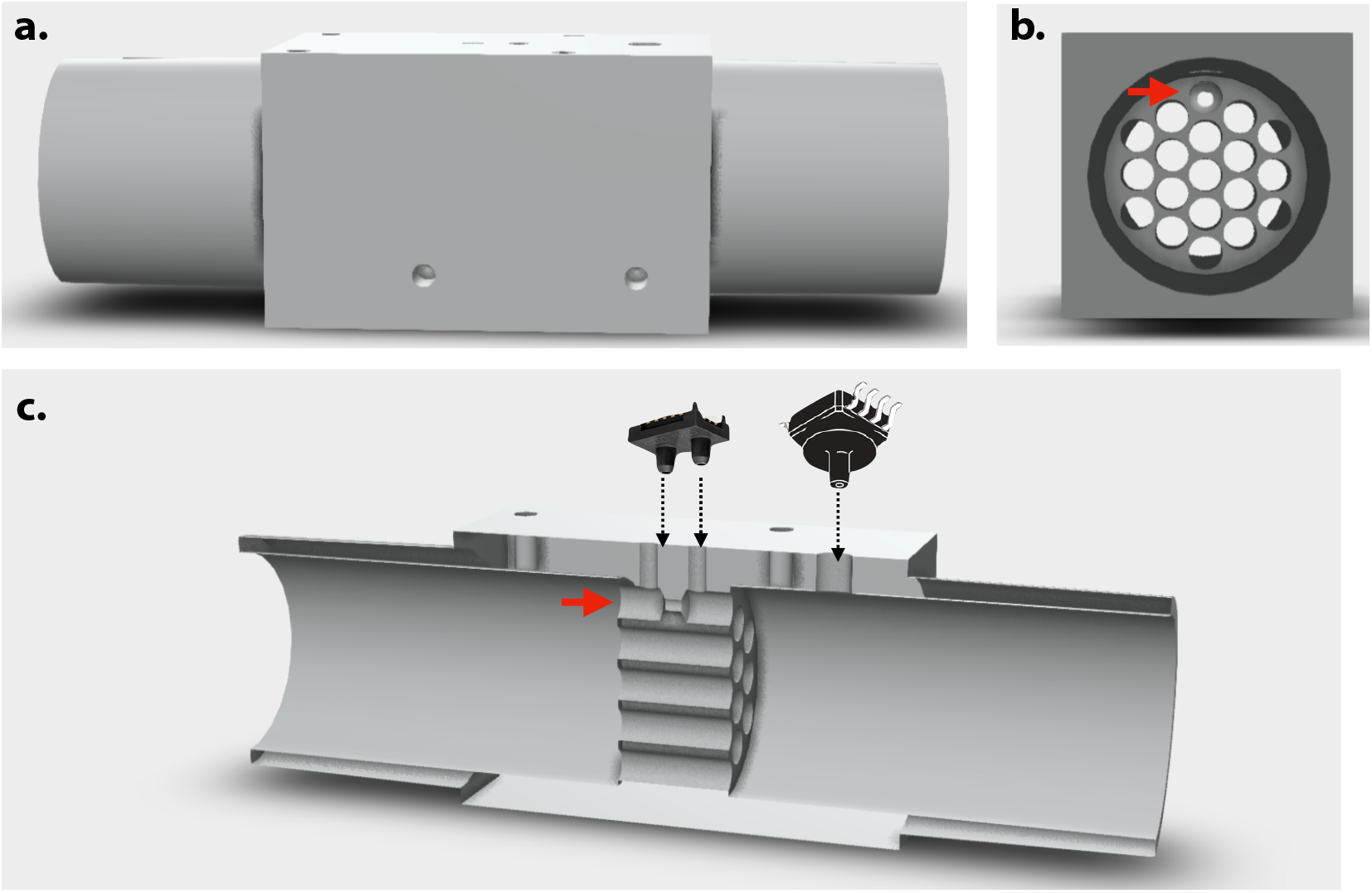
Flow element. The flow element is a 19 mm diameter tube of length 82 mm with an obstruction to create a pressure drop. **a**. Exterior view. **b**. View along the direction of airflow. A honeycomb pattern of holes decreases turbulence. **c**. Interior view. Flow is detected as a pressure drop across a small obstruction in one of the channels (red arrow), as measured by a differential pressure sensor (two pronged device, pictured). An additional sensor measures gauge pressure.

### 8.5 Flow element construction

Flow elements are manufactured on a CNC mill from 2·54 × 2·54 cm^2^ white Delrin Acetal resin rod (McMaster-Carr part number 8739K92). The material is commonly used in FDA approved medical devices, meets ASTM D6100 standards, and is readily available. The CAD machining file is available in Supplementary Design File 2. One of the goals of the machining was to make the blocks repeatable enough that the flow versus pressure drop of all blocks could be characterized with a just a few parameters, eliminating the need for individual calibration. It was particularly important to maintain the uniformity and spacing of the eighteen 3·175 mm-diameter holes (for example, see Fig. S3). Drill wander was minimized by using stiff bits and starting the hole by “pecking.” Deburring is also important, as there is a tendency for residual Delrin “hair” to form at the entry and exit of the hole, see S4. This was reduced by touching up the holes with a slightly larger bit. With proper jigging and after some practice, one block could be machined in under 20 minutes. The yield is about 80%.

**Fig S3.**
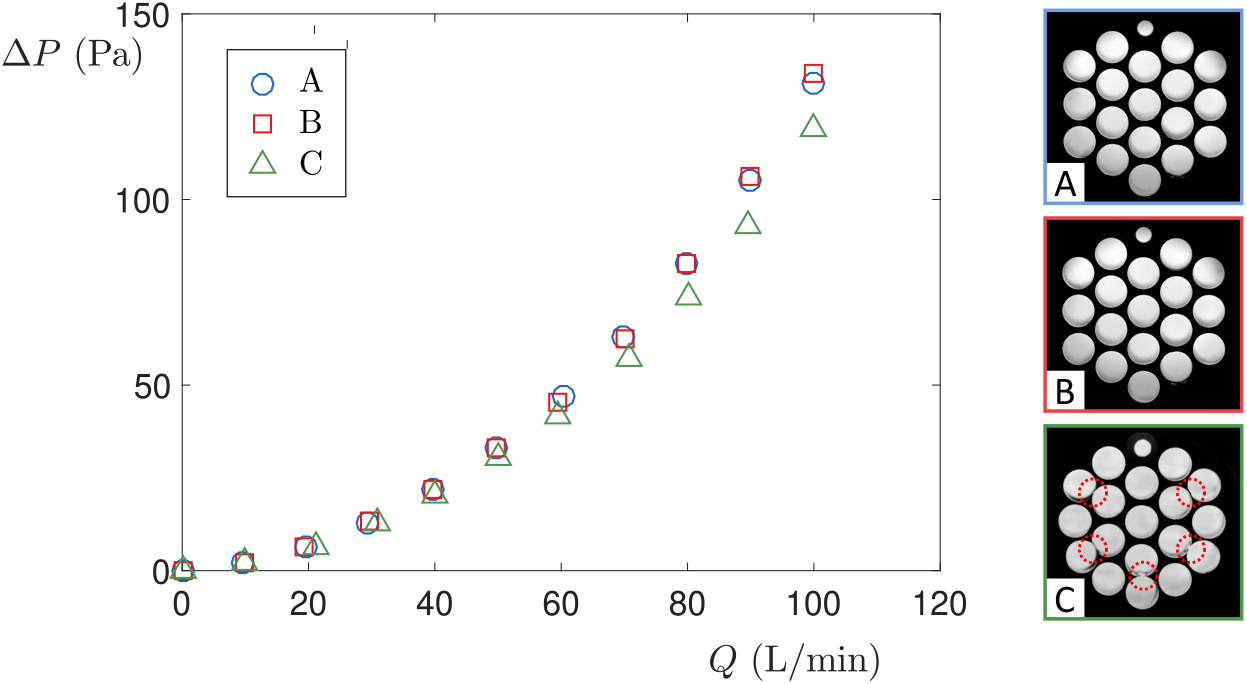
Influence of large machining defects on flow calibration. Measurements on flow element A and B overlap nicely, but measurements on C are affected by the obvious defects in the honeycomb pattern (red dashed circles in the bottom picture).

Cleaning the blocks to “medical grade” required some care. The primary goal was to remove all residual Delrin hair. This was most successfully done by a commercial company [18], using a frozen CO_2_ deburring technique, see Fig. S4. The hair could also be melted away by application of a heat gun. In addition, we had success by mounting a 3 175-mm-diameter “low scratch” tube brush dipped in 80 grit water-based slurry in a hand drill and then polishing each hole. These last two methods require some care with the spinning brush being preferable. After the mechanical processes, blocks were cleaned with soap and water and then sonicated in isopropanol for 10 minutes.

### 8.6 CO_*2*_ sensor

The CO_2_ sensor assembly is shown in Fig. S5. The Sensirion SCD30 environmental sensor is used to collect CO_2_ concentration, temperature and relative humidity data at 2 second intervals along the flow axis. The block assembly is similar though slightly larger than the flow assembly to accommodate the SCD30 board, which we position along the axis of the flow tube. The input port is a male 22 mm connection, but the output port is a female 22 mm connection, allowing the CO_2_ sensor block to be placed directly on the input of the flow block in series with the flow. The communication protocol is I2C. Due to the unique device address of the SCD30 we use common bus lines to readout the flow block and CO_2_ sensor. A “Y”-cable is used to split the patient-side RJ-45 connector into two input ports to allow for the flow block and CO_2_ sensors to be connected to the same patient box. The use of the CO_2_ monitoring is optional. The enclosure was printed from PLA, see Supplementary Design File 2.

**Fig S4.**
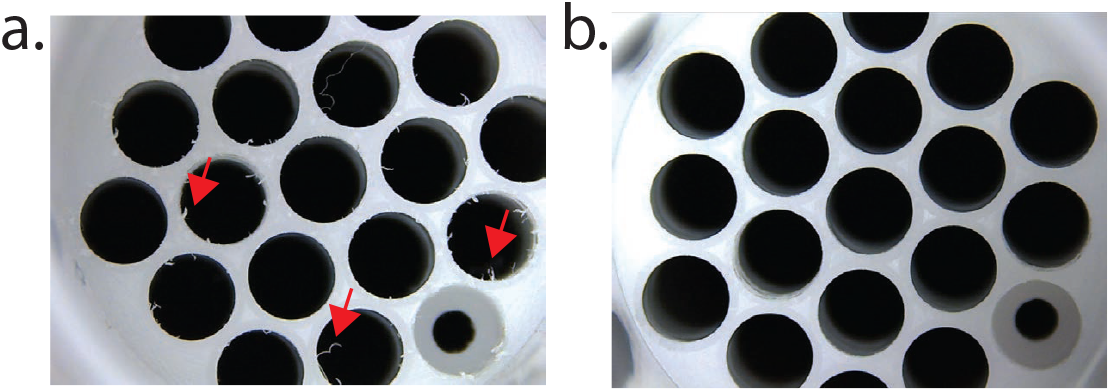
Delrin hair removal. Flow block is show a) before and b) after frozen CO_2_ deburring. Note the small delrin hair filaments (red arrows) are gone.

**Fig S5.**
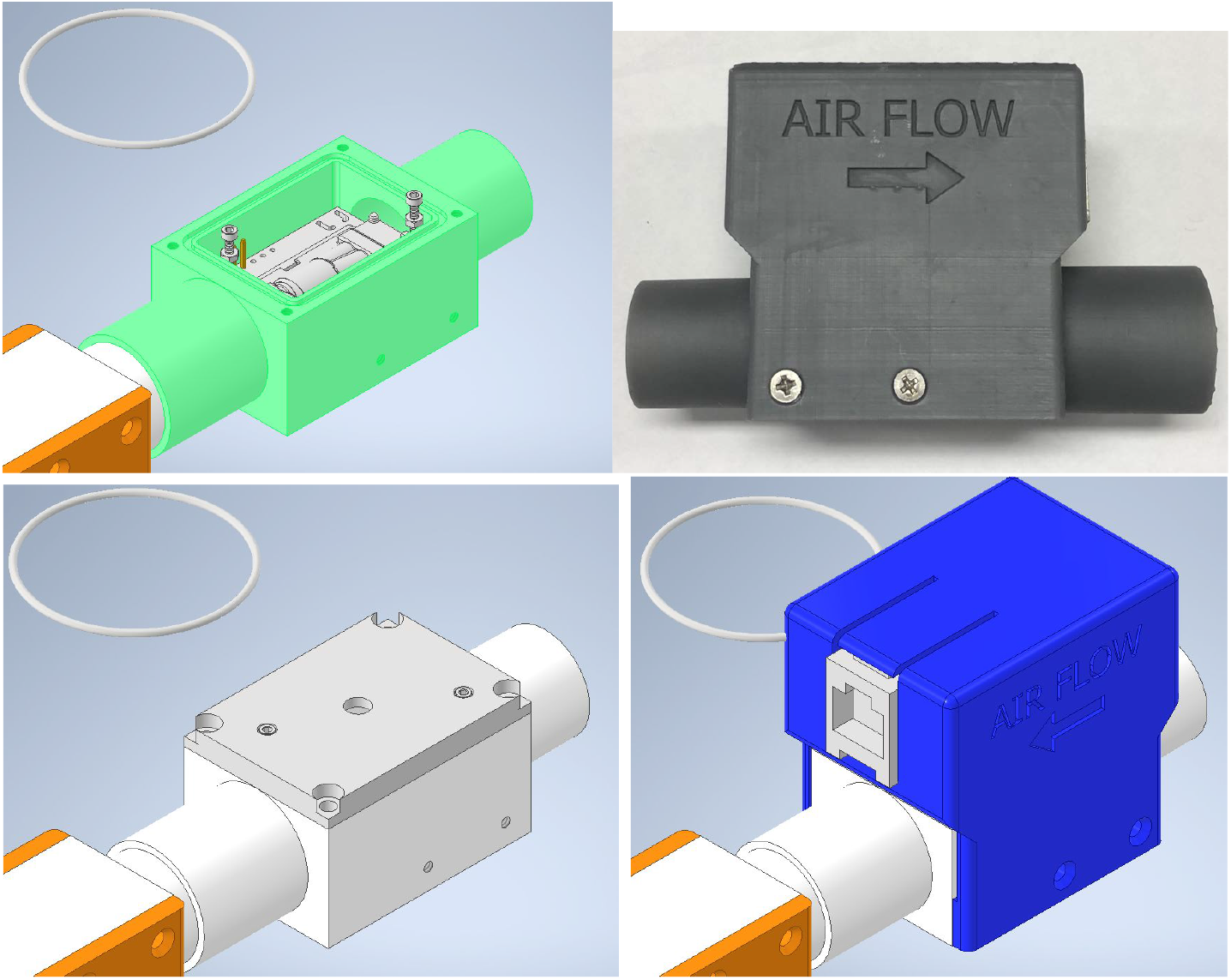
CO_2_ sensor assembly. The SCD30 sensor board is mounted on the center axis for inline flow monitoring. The diameter of the output side of the block fits directly over the flow element input, allowing the sensors to be directly connected in series. The sensor communicates over a common I2C bus protocol to an external RaspberryPi controller via a standard RJ-45 jack.

### 8.7 Interface box

The interface box reads out the flow, pressure, and CO_2_ sensors, provides an alarm system with a human interface, and transmits data to a central remote monitoring station for remote, multi-patient monitoring. The core element of the interface box is the microcontroller, which was chosen to be the Raspberry Pi (RPi) Model 4B, running the Raspbian operating system. The RPi is low in cost, widely available, and enjoys the support of a broad user community. It also provides flexibility and allows for additional sensors and interfaces. Wired and wireless network ethernet interfaces are available on the RPi. The human interface was chosen to be simple and minimal. The setting and management of alarm functions is accommodated by a two-line 20-character-per-line LCD screen with individual Red/Green/Blue LED backlights, a pushbutton rotary encoder, and a red-colored “Silence” pushbutton.

**Fig S6.**
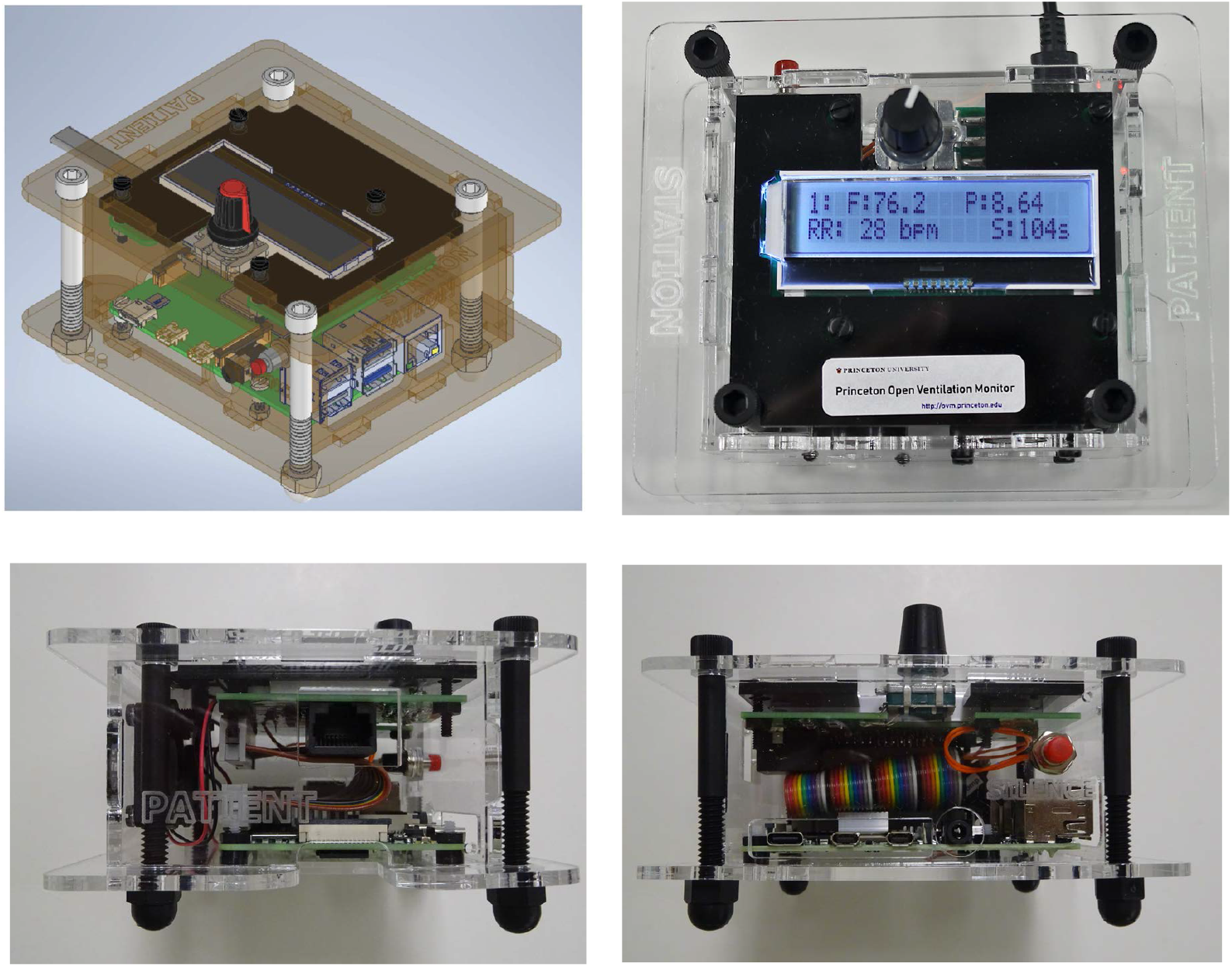
Interface box. Raspberry Pi based interface box sits at patient bedside. It provides an alarm system with a human interface and it relays sensor information to the central remote monitoring station.

On the sensor readout side, the digital bus interfaces were restricted to comply with 3·3V I2C and SPI and the control elements to 3·3V PWM and GPIO. The data transmission distance of the I2C bus imposes a proximity requirement on the interface box to the flow element of approximately 2 m. The time-base of the readout is derived from the RPi system clock using timer interrupts. The human interface is based on asynchronous signal interrupts.

The SDP31 differential pressure sensor is placed into continuous, average until read, mode. The RPi reads the SDP31 over I2C at a rate of 50 Hz, where each reading is the arithmetic average of 40 individual readings taken at 0·5 ms intervals. The MP3V5004G pressure sensor has an analog output that is digitized by an MCP3202 12-bit ADC from Microchip. The MCP3202 is read out at a rate of 200 Hz over SPI. A 4-sample arithmetic average is formed and recorded at a rate of 50 Hz on the same time-base as the SDP31. The temperature of the SDP31 sensor is read out at 1 Hz.

The preferred operating temperature of the flow block sensors is 45°C to avoid condensation from hot, humid vapor. A heating circuit located on the flow block is adjusted using a convergent “take-back-half” servo algorithm operating on a PWM control signal. The heating circuit has a maximum heating power of 1 W and reaches a maximum temperature of 64°C. A convergent algorithm enables the circuit to operate at a fixed heating value approximately 200 seconds after power-up, with infrequent adjustment. Small shifts of the analog signal from the pressure sensor are observed when the heating power is adjusted.

The addition of a single, customized two-layer PCB was found to be sufficient to make the connections between the interface box hardware and the 40-pin GPIO through a short ribbon cable, see Supplementary Design File 1. This construction made it simple to design an enclosure to host the RPi and interface box components, including: a 2×20 character LCD screen with RGB backlight, pushbutton rotary encoder, silence pushbutton, RJ45 connector, alarm buzzer and cooling fan. Laser cut design files for the enclosure and an assembly video are in Supplementary Design File 2 and Supplementary Video 1.

The initial system software for the RPi is installed on a 32GB microSD memory card with a separate data partition. The disk image is then replicated using multiple USB3 hubs running in parallel to provide identical systems for all interface boxes. The custom enclosure allows the microSD card to be removed and replaced, but to minimize handling of the microSD card, the preferred method of software update is through a local repository on the remote monitoring station.

The alarm is based on a 10 s running average of the flow rate and pressure, as well as the average respiratory rate, calculated using a decaying weight on the last several breaths. Maximum and minimum values are entered directly via the pushbutton rotary knob. Rotation of the knob cycles through alarm setting menus, and a “push and turn” operation is used to change the alarm threshold settings.

The sounding of the alarm generates a PWM volume-controlled buzzer and introduces a red back light to the LCD display. A red silence pushbutton on the interface box may be used to temporarily silence the audible alarm, turning the back light to orange. Silence can be invoked preemptively or post-alarm. The silence duration defaults to 120 s, but can also be manually adjusted upon invoking the silence button. The alarm status and all monitoring data is continuously streamed to the remote monitoring station through a wired ethernet interface. The unique digital identification of the flow element sensors, RPi ethernet MAC address, and a unique interface box naming system allows the remote monitoring station to unambiguously assign alarms to patients, as described in Section 8.15.

### 8.8 Remote Monitoring Station

The remote monitoring station is the primary means of providing clinicians with detailed patient information. The remote monitoring station comprises a networked computer and a 27 inch monitor. Both an Intel NUC miniPC and a Raspberry Pi were tested, although any modern PC running Python on any operating system will suffice.

The remote monitoring station provides a simultaneous overview of all patient interface box data and alarm states. For each patient it shows flow, pressure and volume waveforms and a set of derived quantities, including respiratory rate and equivalent tidal volume (see Fig. 1). In a “drilldown” view it provides full-screen plots of the waveforms for a single patient as well as a pressure-volume plot and derived quantities, while still displaying alarm status information on all patients, see Fig. 10.

The remote monitoring station receives data from interface boxes over a wired or wireless network. In a hospital setting we expect an air-gapped wired network to be used. Our system was tested on an ethernet network with a 24 port hub. The monitoring station acts as an always-on appliance and handles the automatic discovery, pairing and logging of patient interface boxes, described in Section 8.15. Because the interface boxes lack battery-backed real-time clocks, the remote monitoring station maintains a master time record for recorded data streams.

### 8.9 Bill of materials

A summary level bill of materials is shown in Table S1 and detailed bill of materials is in Supplementary Design File 3.

**Table S1.** Summary level bill of materials. Full details are available in Supplementary Design File 3. Circuit board costs include assembly and minor components. Each remote monitoring station accommodates up to 20 patients.

| Subsystem | Item | Cost |
| --- | --- | --- |
| Flow-Sensor Assembly | Flow element | \$ 34 |
| | SDP31 Sensor | \$ 25 |
| | MPV35 Pressure Sensor | \$ 9 |
| | Circuit Boards | \$ 27 |
| | Enclosure | \$ 13 |
| | **Flow-Sensor Assembly Total** | **\$ 108** |
| Interface Box | Microcontroller, LVPS, and disk | \$ 76 |
| | Display and Encoder Knob | \$ 11 |
| | Circuit Board | \$ 30 |
| | Enclosure | \$ 9 |
| | Ethernet cable | \$ 8 |
| | **Interface Box Total** | **\$ 134** |
| Remote Monitoring Station | Intel NUC Computer | \$ 359 |
| | Touch Screen Monitor | \$ 561 |
| | Ethernet Cables | \$ 148 |
| | Network Switch | \$ 80 |
| | **Remote Station Total** | **\$ 1148** |

### 8.10 Software overview

Software is released as open source under the MIT License and is available at https://github.com/Princeton-Penn-Vents/princeton-penn-flowmeter, see also Supplementary GitHub Code Repository. The software integrates together the data collection and handling in the interface boxes and remote monitoring stations, provides an analysis suite for monitoring and alarm determination, drives the rotary-dial interface of the interface box and implements the graphical user interface of the remote monitoring station. It has been designed to work with minimal computational resources, to be easily portable, to have few external dependencies, and to use standard, well supported, tools when external dependencies are used.

The software is written entirely in Python and uses only open-source components. On top of a base Python 3·7 installation, additional requirements are currently NumPy, SciPy, PyQt5, pyqtgraph, pyzmq, confuse/pyyaml, zeroconf and the Python Raspberry Pi tools. Qt was selected for both its ability to display real-time graphics with minimal computation resources, and for its portability across computing platforms including for embedded devices. The sections below discuss the software design and implementation of the interface boxes, the remote monitoring station and the communication between them.

### 8.11 Interface box software

The interface box simultaneously runs two Python applications called device loop and patient loop.

The device loop collects raw data from the sensors and converts it to JSON, which it both transmits through a ZeroMQ broadcast to port 5556 and logs to a simple file. A basic JSON “packet” is produced at 50 Hz containing the monotonic clock, pressure, and flow data. The flow data is averaged over four readings taken at 200 Hz. Every second, an extended packet is produced, which includes the device temperature measurement, heater duty cycle, sensor identification number, and the path of the current file used for logging. This application does not read the main configuration file, and is completely stand-alone from the rest of the system. It is started when the device starts up and runs until the device is powered down.

The second application is the patient loop This application is responsible for reading the ZeroMQ stream from the device loop, powering the on-device LCD display, taking input from the rotary and silence button, applying calibrations, computing the alarm and respiratory analysis, and producing a new 50 Hz JSON ZeroMQ broadcast stream for the remote monitoring station to ingest. The new 50 Hz stream is similar to the original stream, except calibrations have been applied. A different type of JSON structure, with rotary information (including the clinician-set alarm thresholds for respiratory rate, flow and pressure), unique per-device MAC address, current on-device timestamp, silence timer, and last interaction time is sent at 1 Hz or whenever the rotary is changed. A configuration file in YAML format allows most settings to be changed or overridden without changing the Python code.

The patient loop runs several threads. There is a collector thread, which reads the input stream and broadcasts to the output stream (the actual reads and writes to the stream are buffered inside the internal ZeroMQ thread). There is also an analysis thread, which runs a lightweight analysis several times per second and a full breath analysis every few seconds (both rates are configurable); this thread can update the display if a displayed value has changed. The rotary and silence button are interrupt driven, though the silencer does launch timers which run in a thread. Locks are used in several places to protect communication of rotary settings and analysis products.

All communication from the device loop to the patient loop is one-way; and all communication from the patient loop to the remote monitoring station is also one-way. This provides a simpler architecture that was easy to make robust, and ensures that one box can be monitored by multiple stations if needed. One side effect of this is that the real-time clocks on the patient stations are not synchronized, so synchronization is made possible by the timestamps on the remote monitoring station.

Configuration is handled through several YAML files. There is a `processor/config_defaults.yaml` file that stores the defaults for most settings. There is a top-level file `povm.yml` that stores copies of settings that might be commonly configured in a deployment. And then, when running, the patient software stores a `povm-live.yml` file in the data directory that keeps track of runtime changes to the alarm limits, so that the alarm settings get restored when the system is restarted. This file is not intended to be edited directly, and is only searched for the rotary settings. If this file were to be corrupted, this is not a failure - the settings are just not restored. A dedicated thread stores the rotary settings every 10 seconds if they have changed.

### 8.12 Calculating derived quantities

Patient breaths and respiratory features are analyzed both locally on the interface box and on the remote monitoring station.

The interface box sensors produce two direct observables as a function of time: absolute pressure (cm-H_2_O) and air flow (L/min). These two quantities are presented with a digital readout on the bedside interface box. The remote monitoring station displays pressure and air flow as continuous time-series waveforms, along with air volume (mL), integrated from the flow. Since the helmet is designed to have a large constant flow through the circuit, the raw integral of flow steadily increases with time. To obtain a times series that shows volume fluctuations associated with breathing, we apply a weak high-pass filter (Butterworth critical frequency of 0·004 Hz) that removes the slow drift while preserving the shape on timescales relevant to breathing. See Figure S7-left.

We call our estimate of tidal volume an “equivalent Tidal Volume” because it represents our best estimate of the tidal volume as measured on top of the constant flow through the helmet. To estimate equivalent tidal volume (≈ TV), the volume per-breath is needed. Here we use a slightly different scheme to estimate the volume. We subtract off the mean of the flow before integrating and then use a weaker high-pass filter (Butterworth critical frequency of 0·0004 Hz). This improves accuracy in our per-breath estimate of tidal volume when used in an invasive ventilator context, and it offers a good compromise between controlling drift and distorting the shape of each waveform when computing derived quantities.

**Fig S7.**
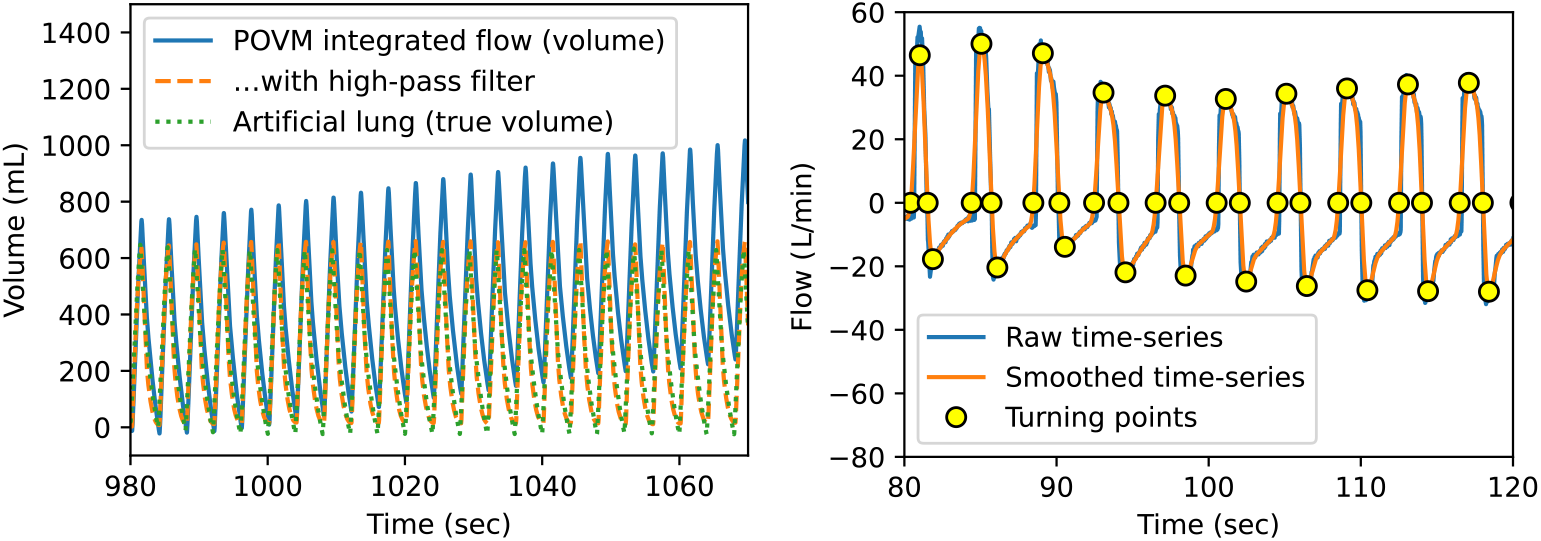
Signal processing to calculate volume and identify breaths. Left: effect of a high-pass filter in removing drift in volume (integral of flow). Recording here is with a test lung. In a helmet circuit the integral drifts because the ventilation system is designed to continuously vent expired air. Right: identifying breath cycles in the time-series data via turning points: roots of smoothed flow and its derivative.

Many derived quantities of interest, such as equivalent tidal volume (≈ TVi and ≈ TVe), respiratory rate (RR), inspiratory-to-expiratory time ratio (I:E), peak inspiratory pressure (PIP), and peak expiratory endpoint pressure (PEEP), are functions of a breath cycle. Breaths can vary in duration, so we need to identify landmarks on the time series that define points along the breath cycle. We do this by first smoothing the absolute pressure and air flow time-series using locally estimated scatterplot smoothing (LOESS) with a Gaussian kernel that has a 0·2 second standard deviation, then finding the roots of the smoothed curve and its derivative as the four turning points of the breath cycle. These four turning points correspond to the time points during a breath cycle when the lungs are least inflated, mid-inhale, most inflated, and mid-exhale, as shown in Figure S7. Once these turning points have been identified, each quantity of interest is derived by analyzing an extremum or a difference of pressure, volume, or time between these points. Breath-based derived quantities are listed in Table S2.

**Table S2.** Quantities derived for each breath from the original timeseries (flow, pressure, and volume versus time).

|  |
| --- |
| When flow is minimal (inhaling) or maximal (exhaling): |
| Time (sec) |
| Flow, as measured (L/min) |
| Flow, smoothed by 200 ms (L/min) |
| Change in pressure per unit time, smoothed by 200 ms (cm-H<sub>2</sub>O) |
| Compliance: change in smoothed flow per unit pressure (L/min/cm-H<sub>2</sub>O) |
| When volume is minimal (empty lungs) or maximal (full lungs): |
| Time (sec) |
| Exhale time: difference between empty and previous full |
| Pressure, as measured (cm-H<sub>2</sub>O) |
| PIP: maximum pressure achieved |
| PEEP: pressure when lungs are empty |
| Volume, derived with 0.004 Hz Butterworth frequency (mL) |
| Inspiratory tidal volume (TV<sub>i</sub>): full volume minus previous empty |
| Expiratory tidal volume (TV<sub>e</sub>): empty volume minus previous full |
| Time between breaths, measured at successive empty lungs (sec) |
| Respiratory rate: inverse of time between breaths (1/min) |

Breath records are logged and accumulate as a growing list. Cumulative measurements are derived from these breath-based measurements as exponentially weighted moving averages (EWMA) of breath records with *α* = 0·3, meaning that it takes approximately 1/0·3 = 3·3 consecutive breaths for a new trend to emerge. In particular, the respiratory rate, inspiratory and expiratory-endpoint pressures (PIP and PEEP), inspiratory and expiratory tidal volume equivalents (≈TVi and ≈TVe), and compliance are all reported as moving averages.

### 8.13 Software detection of Alarms

Alarms are raised when a monitored quantity exceeds a predefined threshold (either too high or too low) and are immediately removed once the quantity returns to a suitable value. Thresholds are set via the rotary dial on each interface box and are passed to the analysis software together with other data. Some alarms are raised on time-window averaged time series, specifically, flow and pressure, while others are raised on EWMA averages of derived quantities, currently only respiratory rate. In addition, “stale data” alarms are raised if any derived quantity is not received after a time window, which could indicate a bad connection.

The analysis functionality is implemented using the rolling buffer functionality described above together with numerical functions from the NumPy and SciPy Python packages. It produces Python dictionaries for breath measurements, cumulative measurements, and alarm states. The analysis is run on both the interface box and the remote monitoring station.

### 8.14 Software on Remote Monitoring Station

The back-end software is shared between the interface box and the remote monitoring station. A Generator is the core class, which handles the analysis thread and most of the data structure. The subclass that does the on-device collection and rebroadcast thread as described in Section 8.11 is the Collector. There is also a RemoteGenerator subclass of Generator, and this instead only reads from an interface box broadcast in a thread. This is used by the main Graphical User Interface (GUI) to monitor each connected interface box.

The remote monitoring station GUI is implemented in PyQt [19]. The GUI main window displays a grid of patient stations, each of which shows waveforms for flow, pressure, and volume and derived quantities. The grid can be rearranged via drag and drop. Clinicians can adjust the waveform axes. Clicking the status button shows advanced options, such as removing a disconnected sensor. Alarm status is displayed using both colors and symbols, as described in Table S3.

**Table S3.** Alarm status GUI scheme for remote monitoring station.

| Alarm Status | Color | Color Applied To | Status Window Text |
| --- | --- | --- | --- |
| No alarm | Black | Status window<br>Title background<br>Figure background | OK |
| No alarm; Silence mode | Black | Status window<br>Title background<br>Figure background | S |
| Alarm | Red | Status window<br>Title background<br>Figure background | ! |
| Alarm; Silence Mode | Yellow | Title background | S! |
| Device Disconnected | Blue | Status window<br>Title background<br>Figure background | D |

Clicking any of the grid tiles on the main screen will bring up a “drilldown” screen of information for that interface box (see Fig. 10). The drilldown screen shows a larger set of waveforms and additional derived quantities. It also includes a Volume vs. Pressure plot and additional information about alarm limits and averaging times, the duration elapsed since the last seen breath, the time elapsed since the last interaction at the patient station, and the countdown until the end of any silence mode. An overview status bar shows status and alarm information from all of the other interface boxes.

The device logs both software state and sensor data. Software-related logging is performed by the main thread and at the level of each Generator. Python errors, changes to the rotary knob, alarms and silencing events are all logged. Sensor data is stored in a CSV file, and breath data is stored in JSON files.

### 8.15 Device discovery and pairing

When patient interface boxes are connected to the network, they are automatically discovered by the remote monitoring station using the python-zeroconf framework. Clinicians then need only to associate the interface box with a given patient by entering the patients name, room number or other identifier.

To reduce the chance of error when associating interface boxes with patients, each interface box has an effectively-unique human-friendly name. The interface box and remote monitoring station share a deterministic naming algorithm that assigns adjective-noun combinations to the patient interface box’s ethernet address. Two word names are chosen from 4096 curated adjectives and nouns each (for a total of 16· 7 million possible names), using a pseudorandom algorithm seeded by the bits of the ethernet address. Unlike assigned names, which could be lost due to memory or disk errors, and which require careful records to avoid collisions, the device names are fully determined from the hardware ethernet address. Names are displayed on each interface box, and also on the remote monitoring station.

The names of the first 20 devices we produced are given in Table S4.

**Table S4.** Hardware ethernet addresses and associated human-readable names for the first 20 patient interface boxes produced.

| Ethernet address | Device name |
| --- | --- |
| dc:a6:32:8f:67:58 | cloudy pat |
| dc:a6:32:8f:67:46 | time tooth |
| dc:a6:32:82:f6:a2 | valuable canyon |
| dc:a6:32:83:73:79 | experimental chunk |
| dc:a6:32:8f:55:64 | regal tactile |
| dc:a6:32:83:73:4f | eastern wardrobe |
| dc:a6:32:8f:67:8b | untenable text |
| dc:a6:32:87:49:e7 | unfamiliar outset |
| dc:a6:32:8f:67:0e | abundant creek |
| dc:a6:32:83:73:b5 | ceaseless doubter |
| dc:a6:32:83:71:84 | dressy clavicle |
| dc:a6:32:80:73:99 | glum seeder |
| dc:a6:32:83:6f:fb | warlike bull |
| dc:a6:32:87:4a:11 | timorous puzzle |
| dc:a6:32:83:73:3d | such feather |
| dc:a6:32:87:4c:a2 | liable ability |
| dc:a6:32:84:e4:2e | conformable lily |
| dc:a6:32:83:71:45 | cohesive macro |
| dc:a6:32:71:76:71 | international clock |
| dc:a6:32:82:99:ba | unhurried camp |

**Supplementary Video 1. Instructional video showing interface box assembly**. Video available online at https://vimeo.com/433023739.

**Supplementary Design File 1. Electrical design files**. A zip file, `SuppDesignFilesElectrical.zip` (568 kB), containing electronic schematics, board layout, PCB design file, part placement and loading figures and GERBER fabrication files. Contents are described in Table S5.

**Supplementary Design File 2. Mechanical design files**. A zip file, `SuppDesignFilesMechanical.zip` (2·3 MB), containing computer aided design files for laser cutting, 3D printing and CNC machining the interface box enclosure, flow-sensor assembly cap, flow element and CO_2_ sensor block. Contents are described in Table S6.

**Supplementary Design File 3. Detailed Bill of Materials**. Spreadsheet in Excel format, `BillOfMaterials.xlsx` (22 kB), containing detailed pricing and part numbers for all components.

**Supplementary Data Files 1. Human subject recording**. `HumanSubjectTestDataAndAnalysis.zip` (18·9 MB) Recording of a healthy human subject volunteer breathing normally in a helmet. Analysis code is provided to separately access the data from the auxiliary and expiratory paths in the case of the proximal sensor configuration, and to access the expiratory path for the human subject breathing from within the helmet. A portion of this recording is shown in Fig. 10. The compressed zip file contains two sets of files within it: the first, `device_raw_waveforms_expiratory.json` and `device_raw_waveforms_auxport.json`, are the data streams recorded from the Princeton Open Ventilator Monitor in JSON format that includes raw sensor readings. The second, `calibrated_waveforms_expiratory.json.csv` and `calibrated_waveforms_auxport.json.csv`, are the corresponding CSV files that contain recorded pressure, flow and volume over time in calibrated units. The CSV file contains two volumes corresponding to the more- or less-aggressively high-passed filtered volumes discussed in the text. Note the baseline of the volume data have undefined offsets due to the persistent flow through the helmet. Example Jupyter notebook analysis files are included with the data. The analysis codes provide the time markers for the start and stop of multiple periods of data-taking, including two stable breathing periods with the human subject within the helmet, three external breathing configurations with three different flow rates, and a null data setup with data taken for an empty helmet and a constant flow. Examples of how the respiratory rates are computed and the extraction of per breath quantities are also included in the code. The analysis codes read the flow calibration file `flowcalib_ave200619.yml`.

**Table S5.**
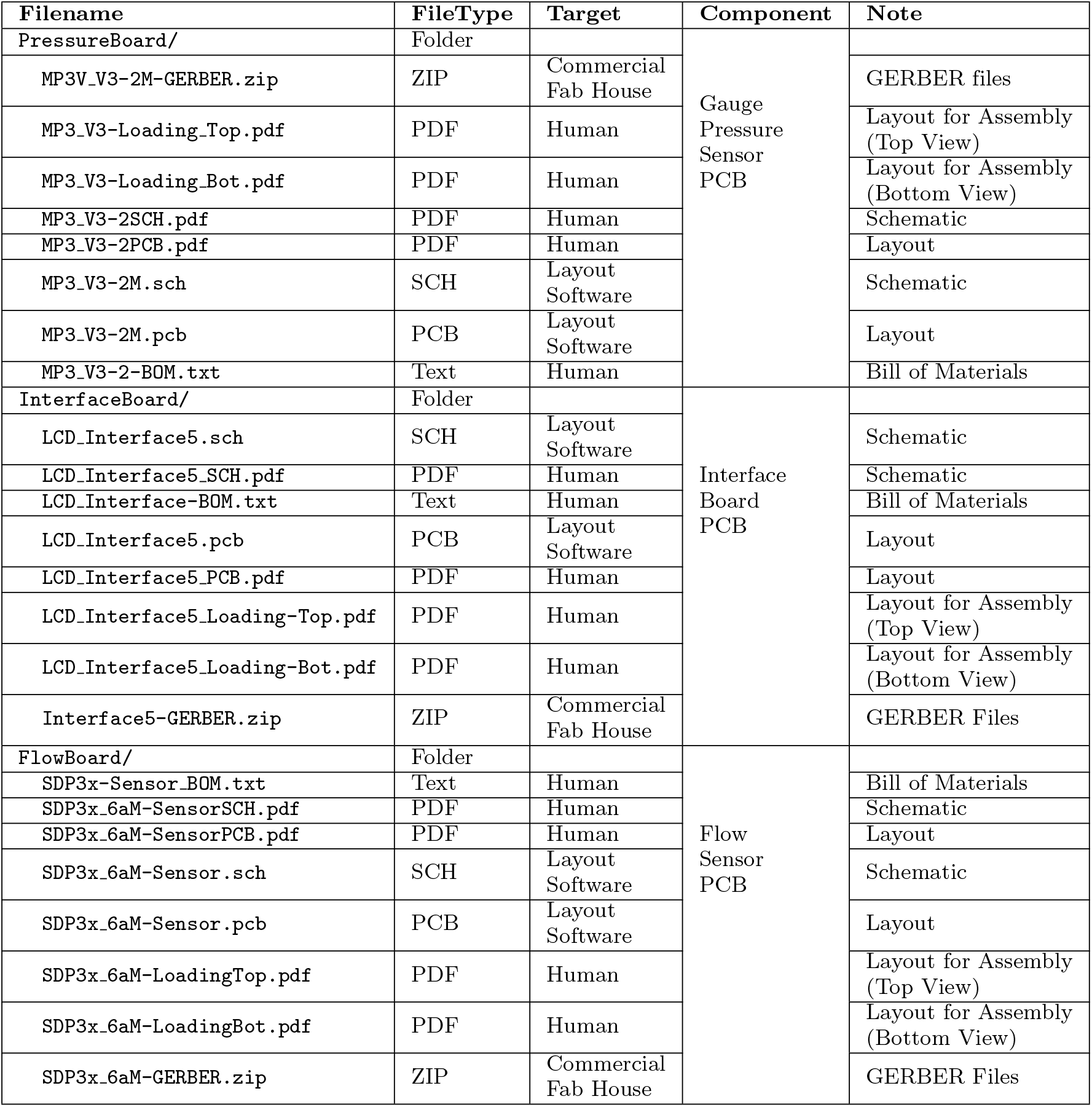
Contents of Supplementary Design File 1: `SuppDesignFilesElectrical.zip`.

**Table S6.**
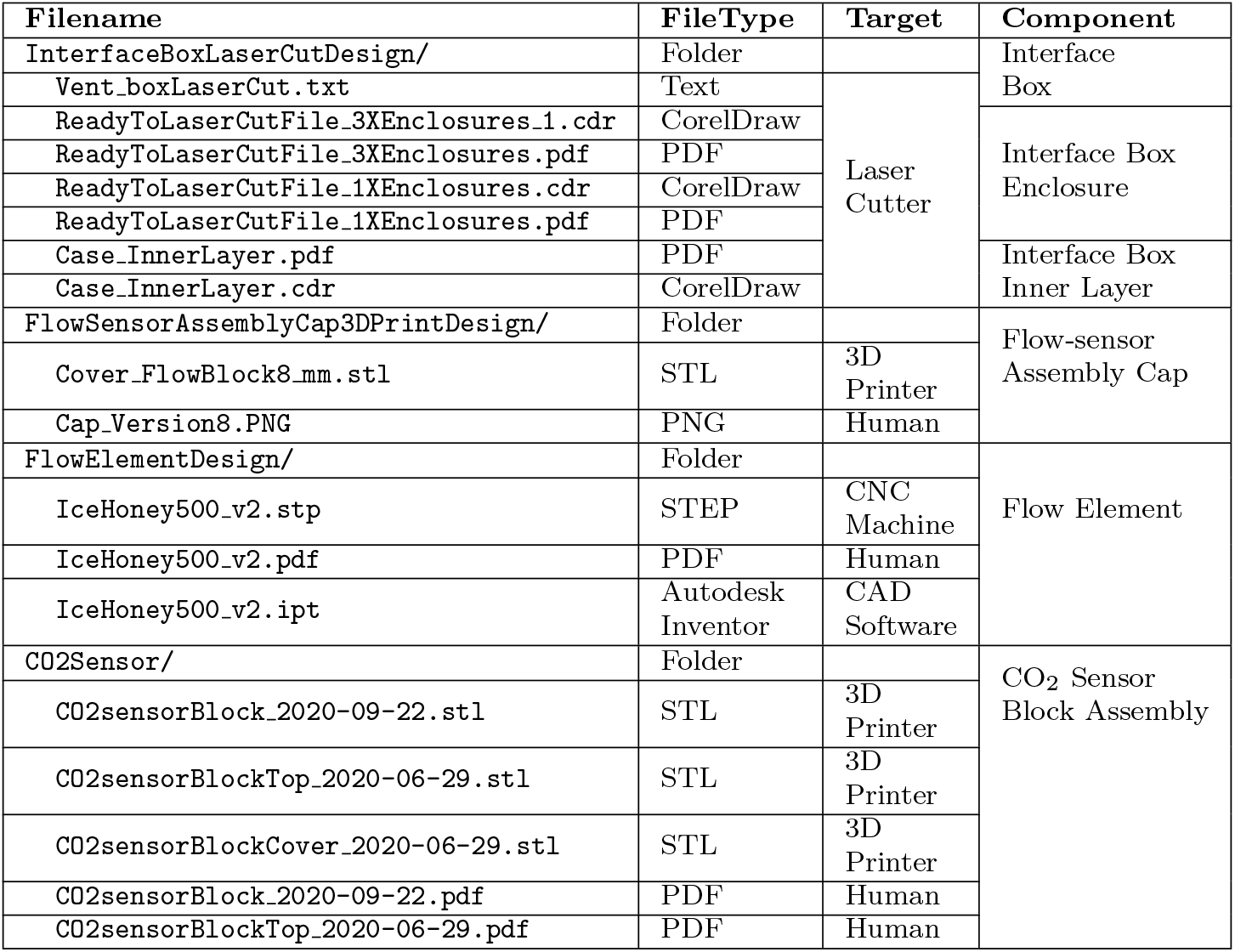
Contents of Supplementary Design File 2: `SuppDesignFilesMechanical.zip`.

**Supplementary GitHub Code Repository. Code repository**. The software for operating the interface box, the patient loop and the multi-patient remote monitoring station is available online at: https://github.com/Princeton-Penn-Vents/princeton-penn-flowmeter. A zip file, `princeton-penn-flowmeter-0.8.1.zip` (249 kB), contains a tagged version (v0*·*8*·*1) of the software repository.

