## Supplementary Design File 1 for "Inexpensive multi-patient respiratory monitoring system for helmet ventilation during COVID-19 pandemic": LCD_Interface5_SCH.pdf

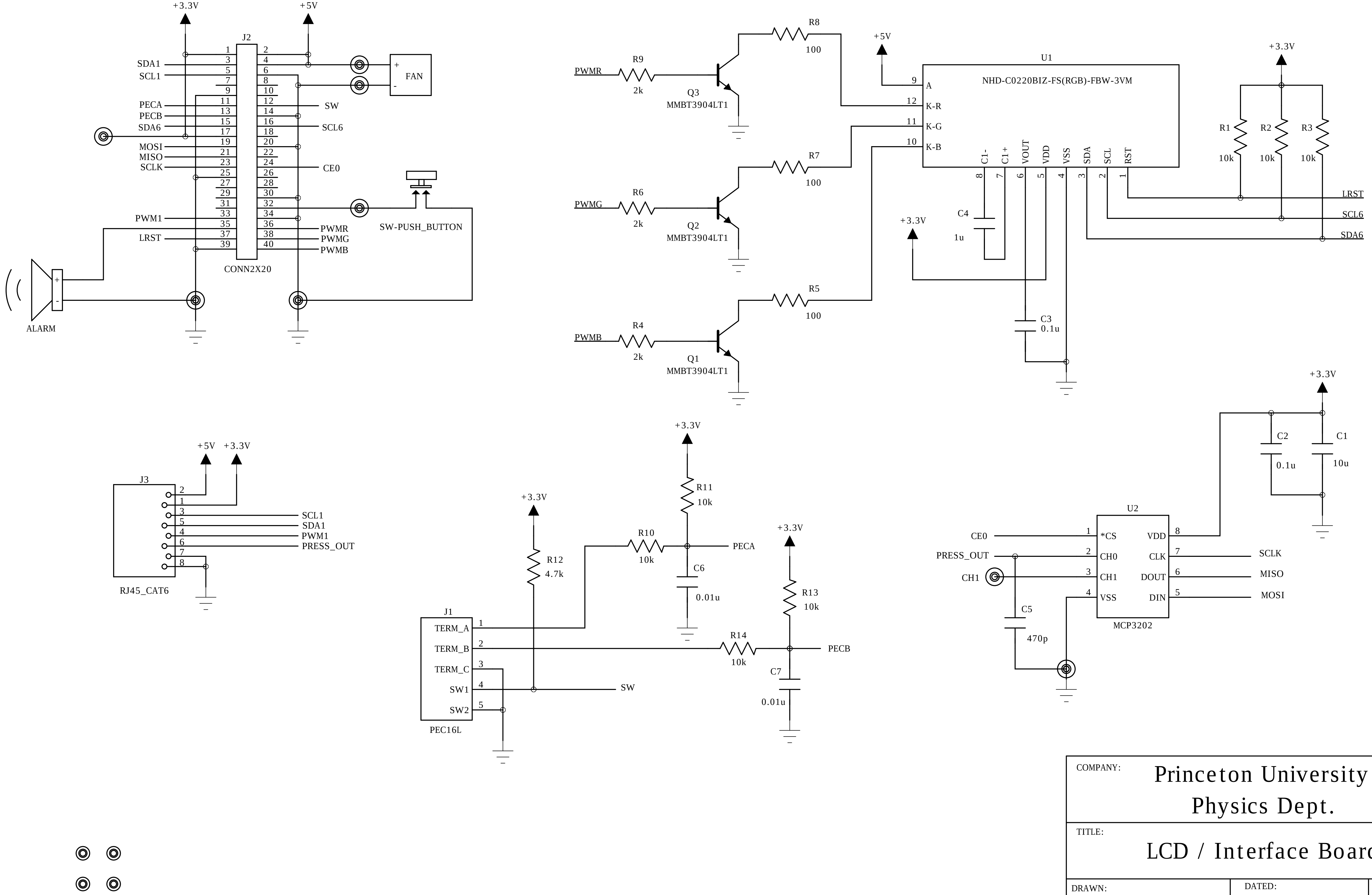


COMPANY: Princeton University Physics Dept.

TITLE: LCD / Interface Board

DRAWN: stanc

DATED: 26October2020

SHEET: 1 OF 1
