## Supplementary Design File 1 for "Inexpensive multi-patient respiratory monitoring system for helmet ventilation during COVID-19 pandemic": MP3_V3-2SCH.pdf

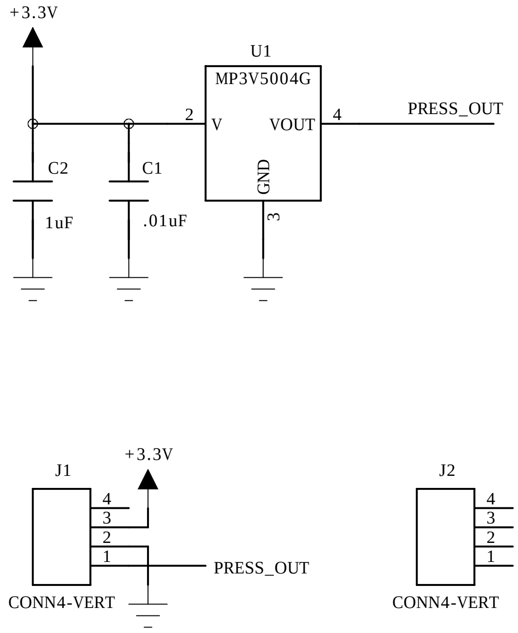


| | | | |
|---|---|---|---|
| COMPANY: Princeton University Physics Dept. | | | |
| TITLE: MP3V Sensor Board | | | |
| DRAWN: stanc | DATED: 18April2020 | REV: 2 | SHEET: 1 OF 1 |
