## Supplementary figures and images for "Inexpensive multi-patient respiratory monitoring system for helmet ventilation during COVID-19 pandemic"

### Cap_Version8.PNG

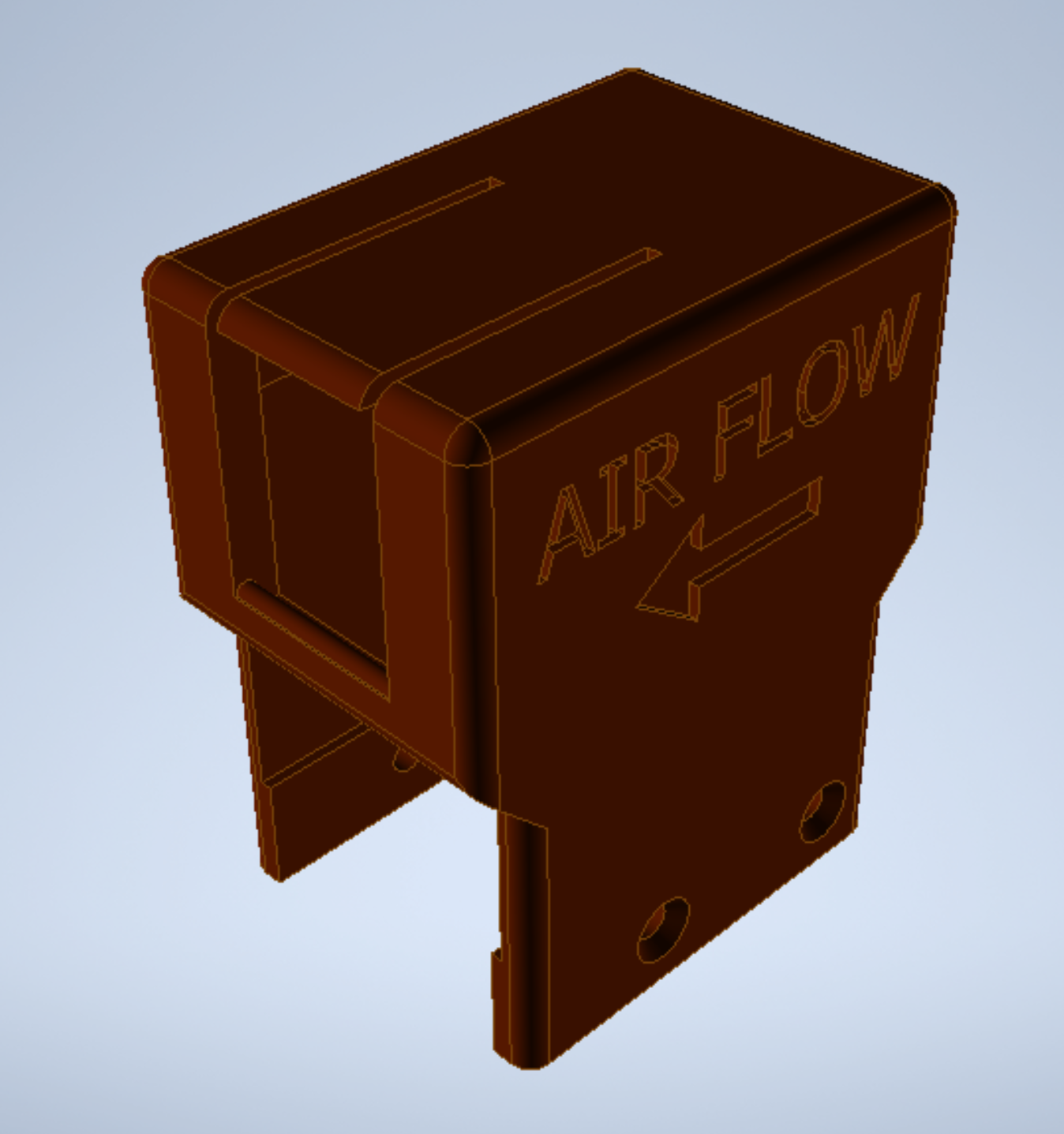

### Case_InnerLayer.pdf

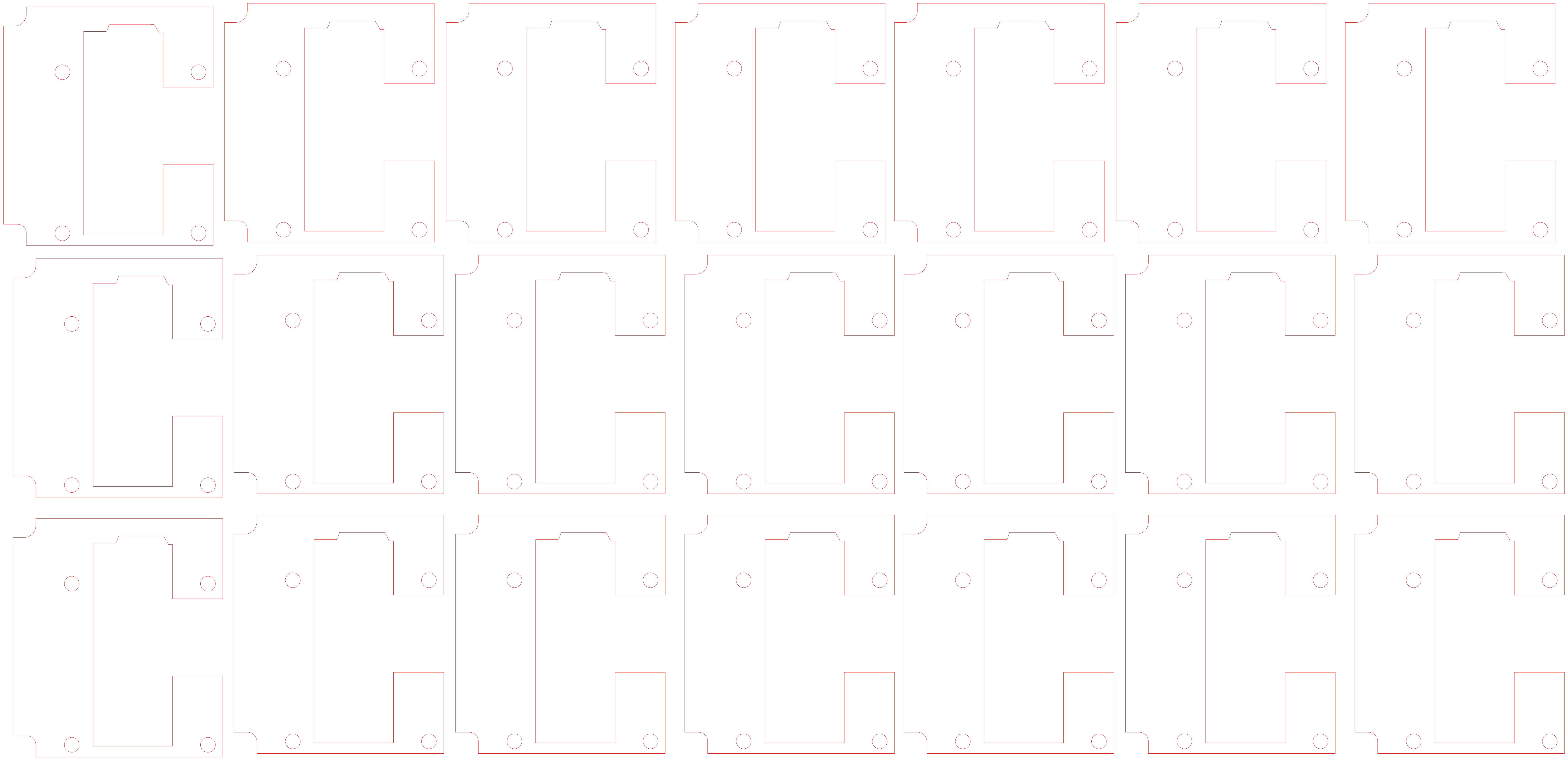

### LCD_Interface5_Loading-Bot.pdf

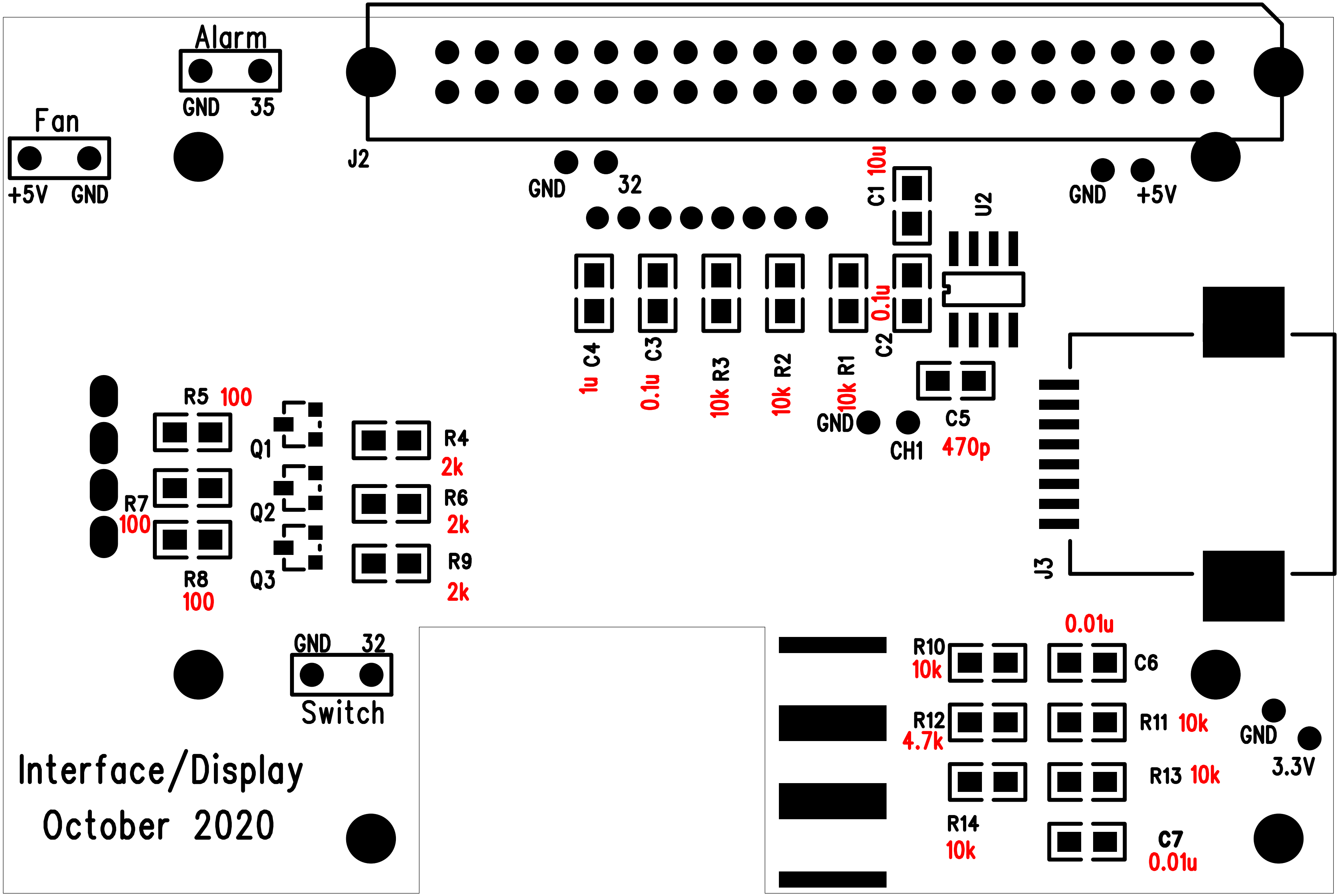

### LCD_Interface5_Loading-Top.pdf

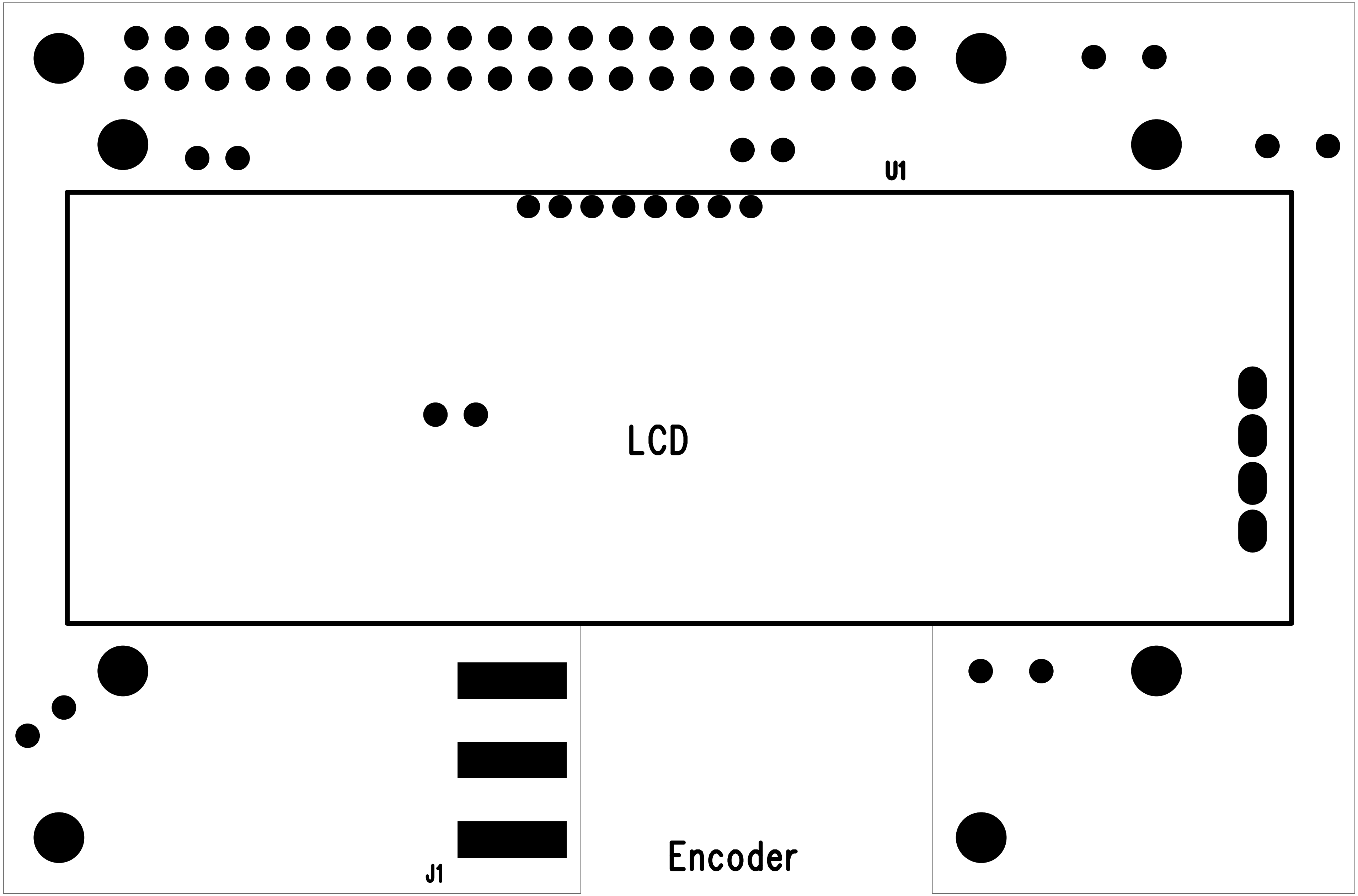

### LCD_Interface5_PCB.pdf

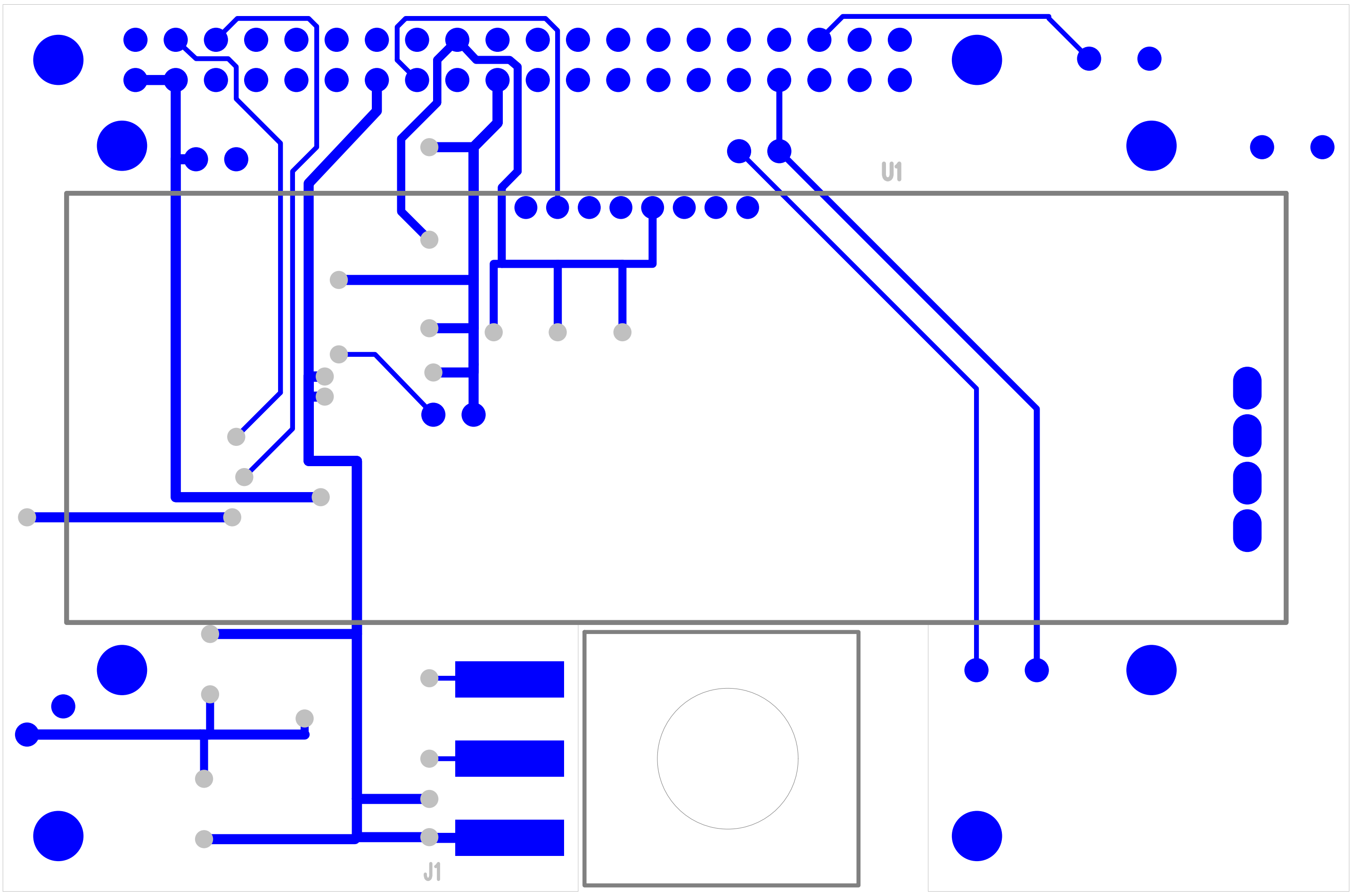

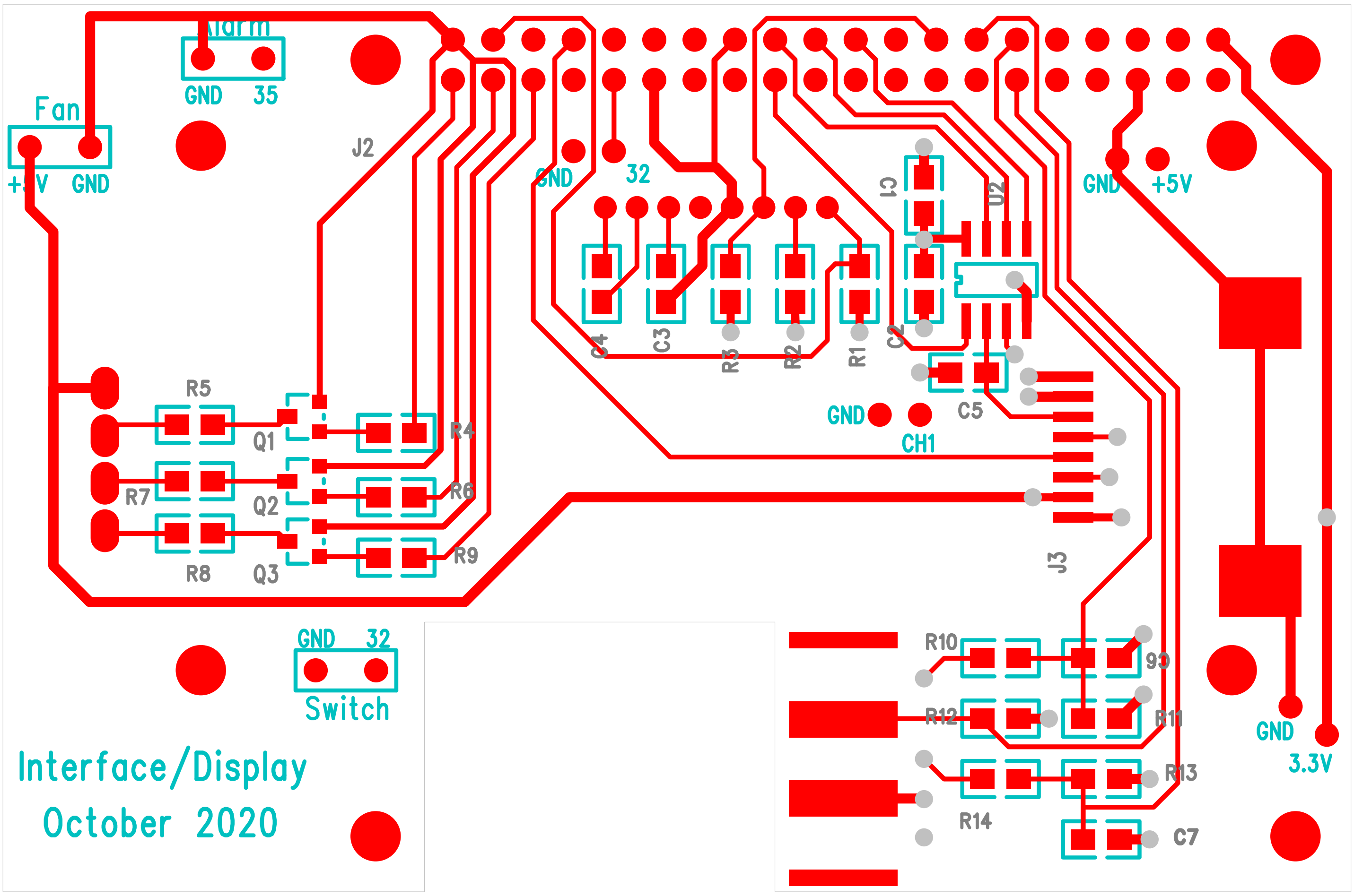

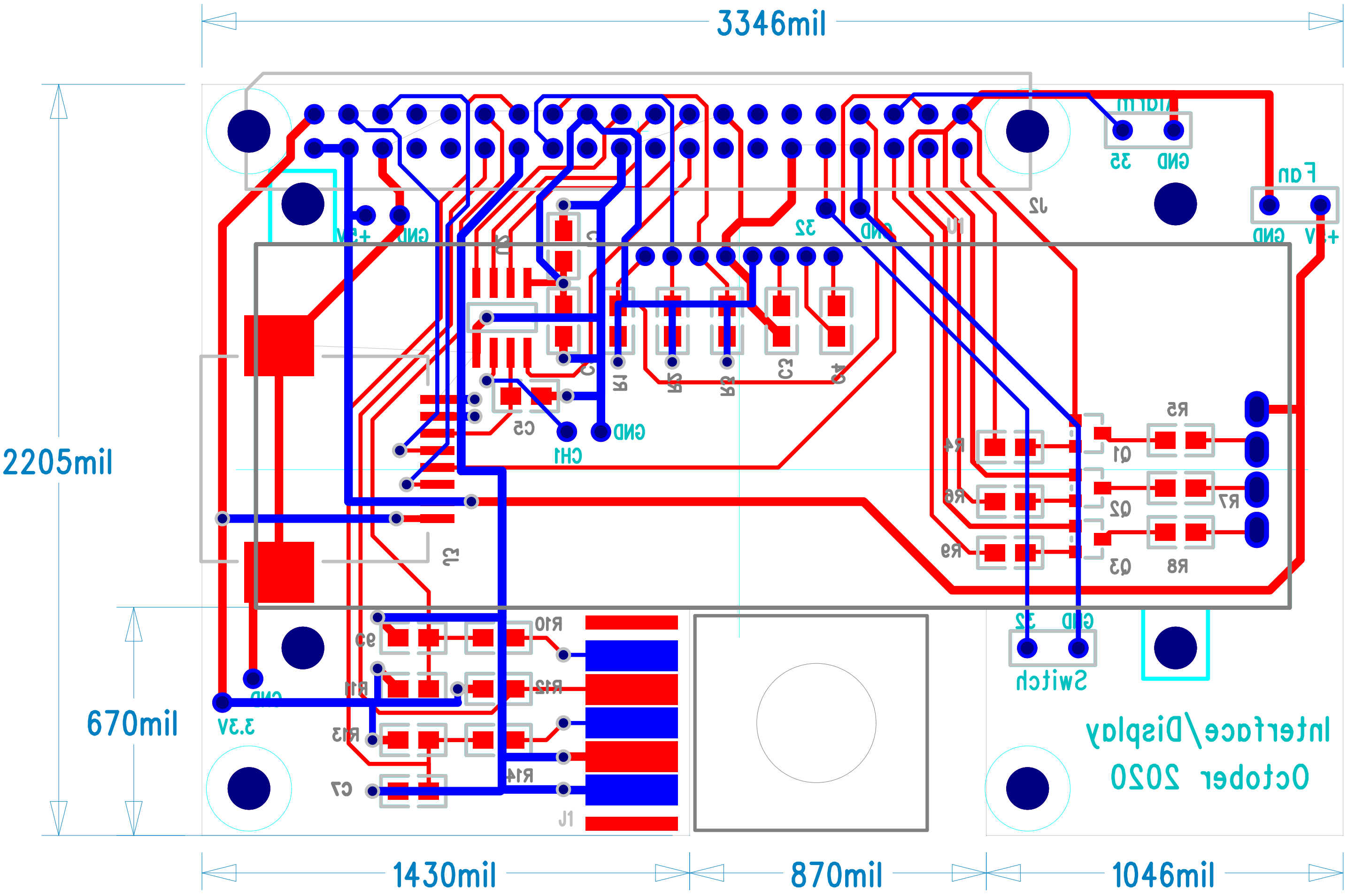

### MP3_V3-2PCB.pdf

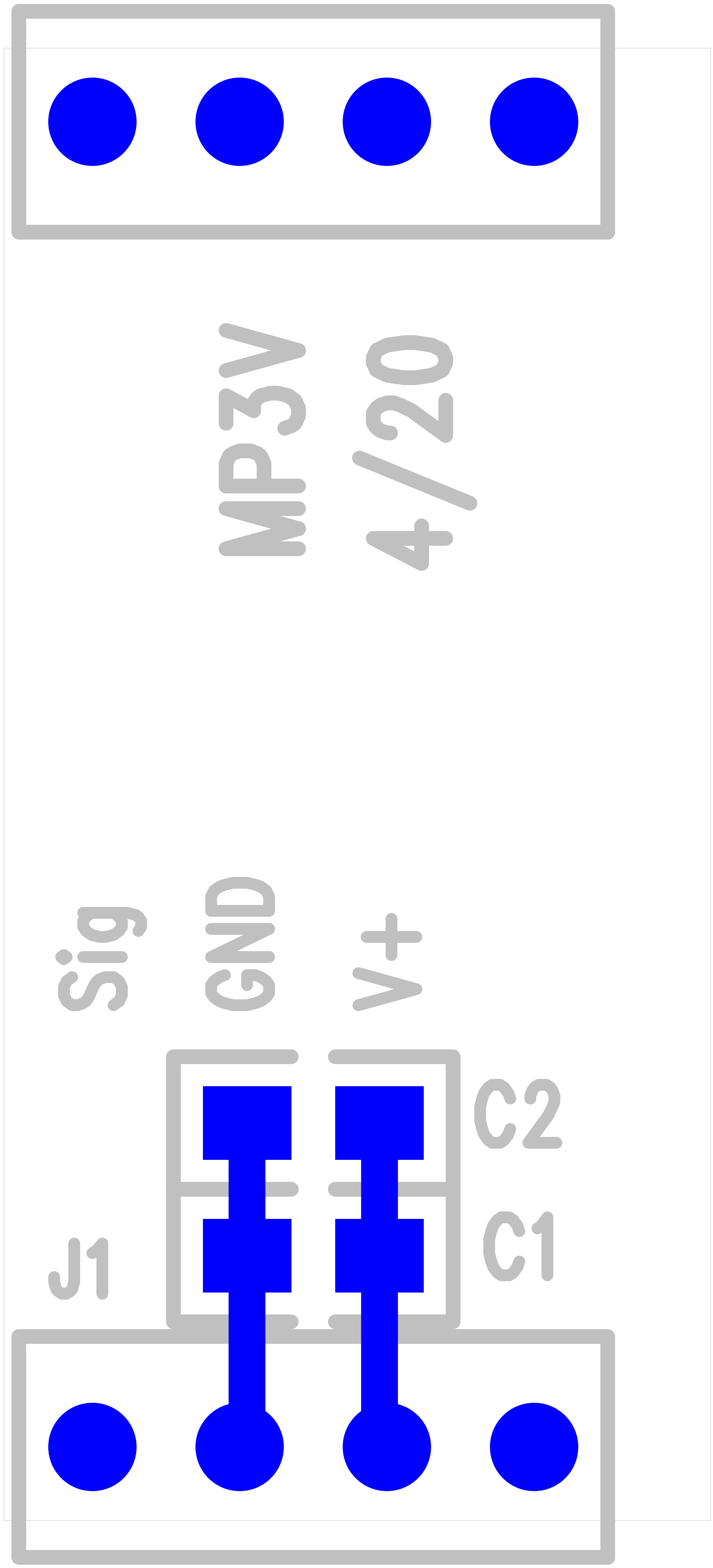

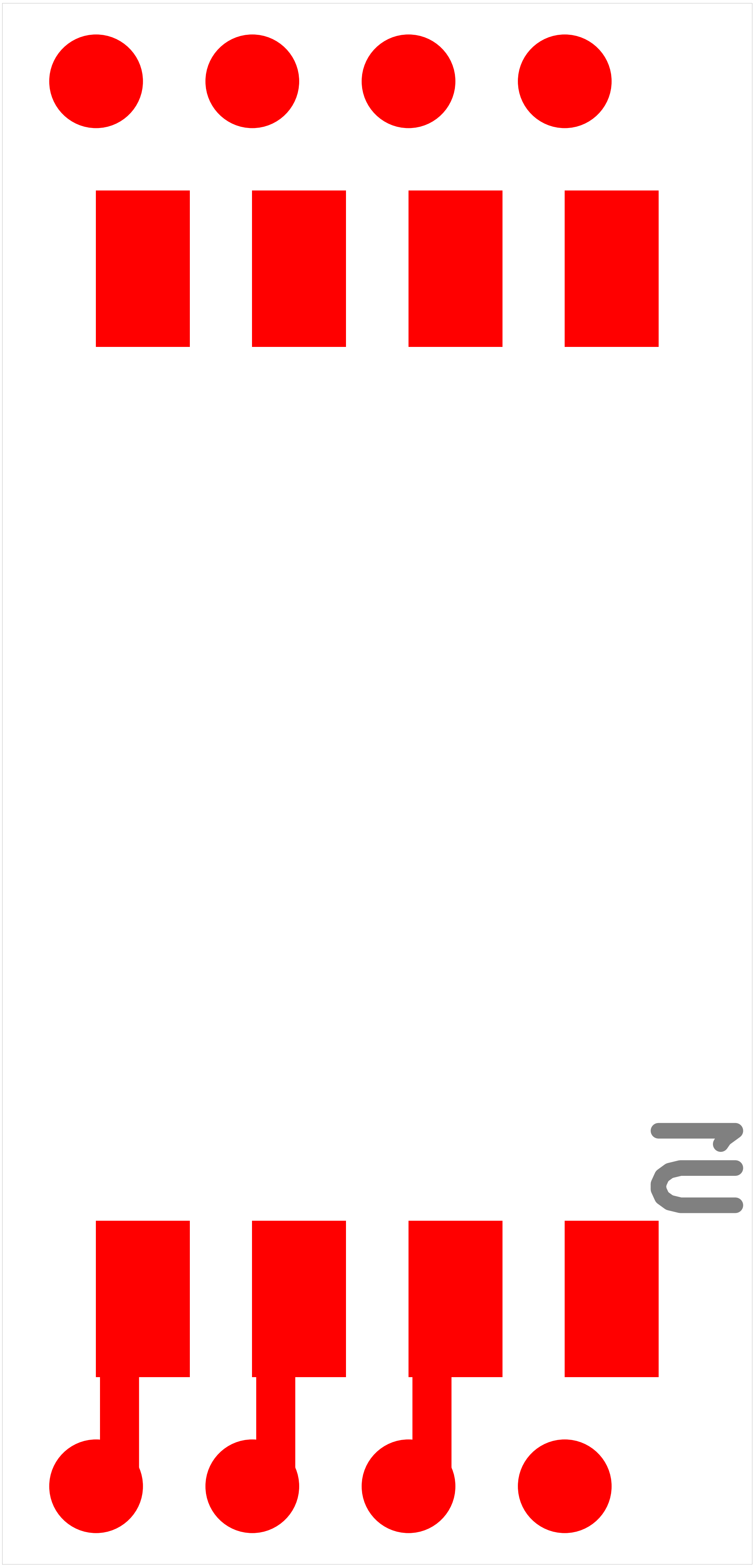

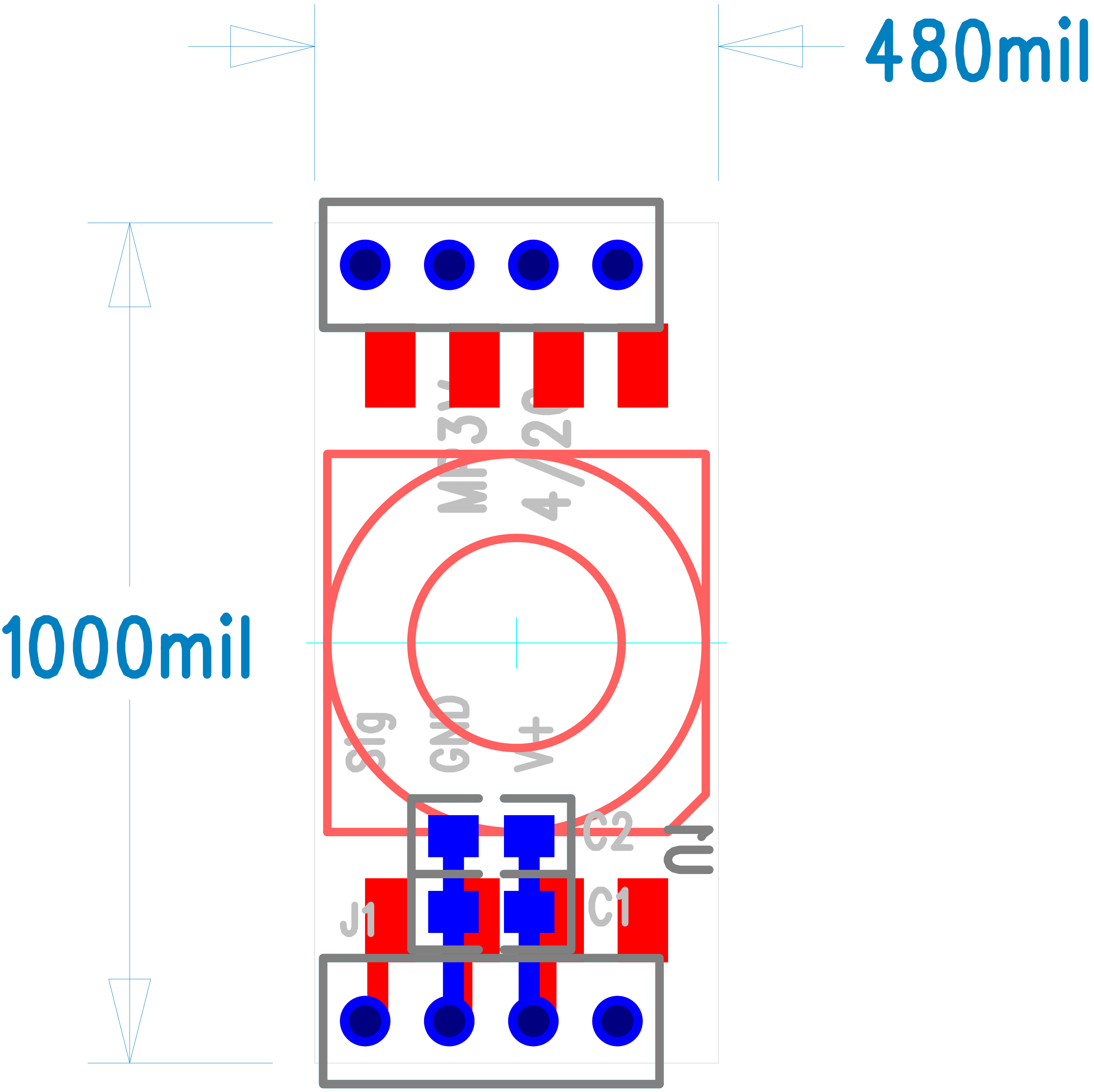

### MP3_V3-Loading_Bot.pdf

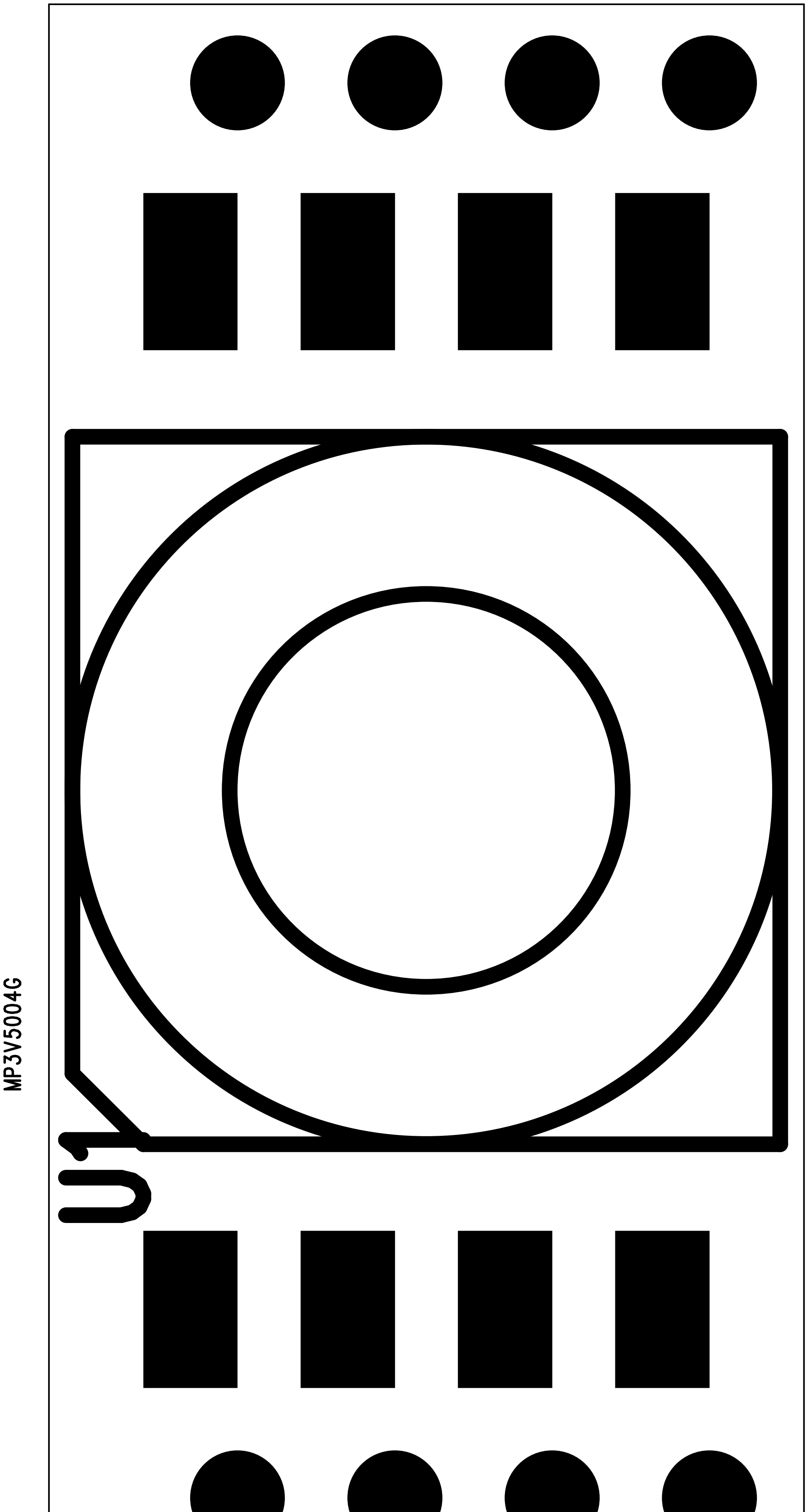


MP3_V3-2.pcb - Tue Apr 21 22:09:35 2020

### MP3_V3-Loading_Top.pdf

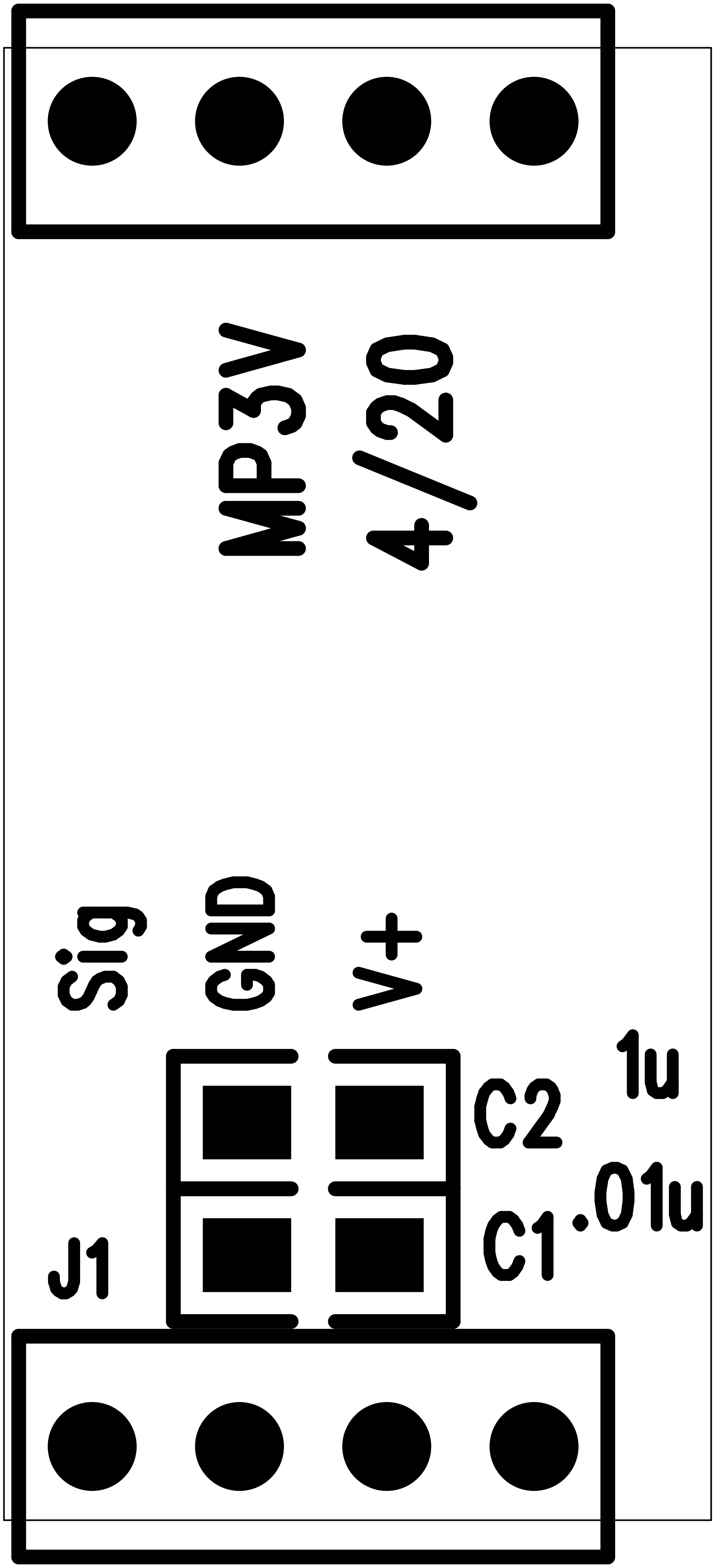

### nsf-logo-100.png

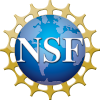

### PUsig2-158C-shield.png

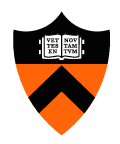

### ReadyToLaserCutFile_1XEnclosures.pdf

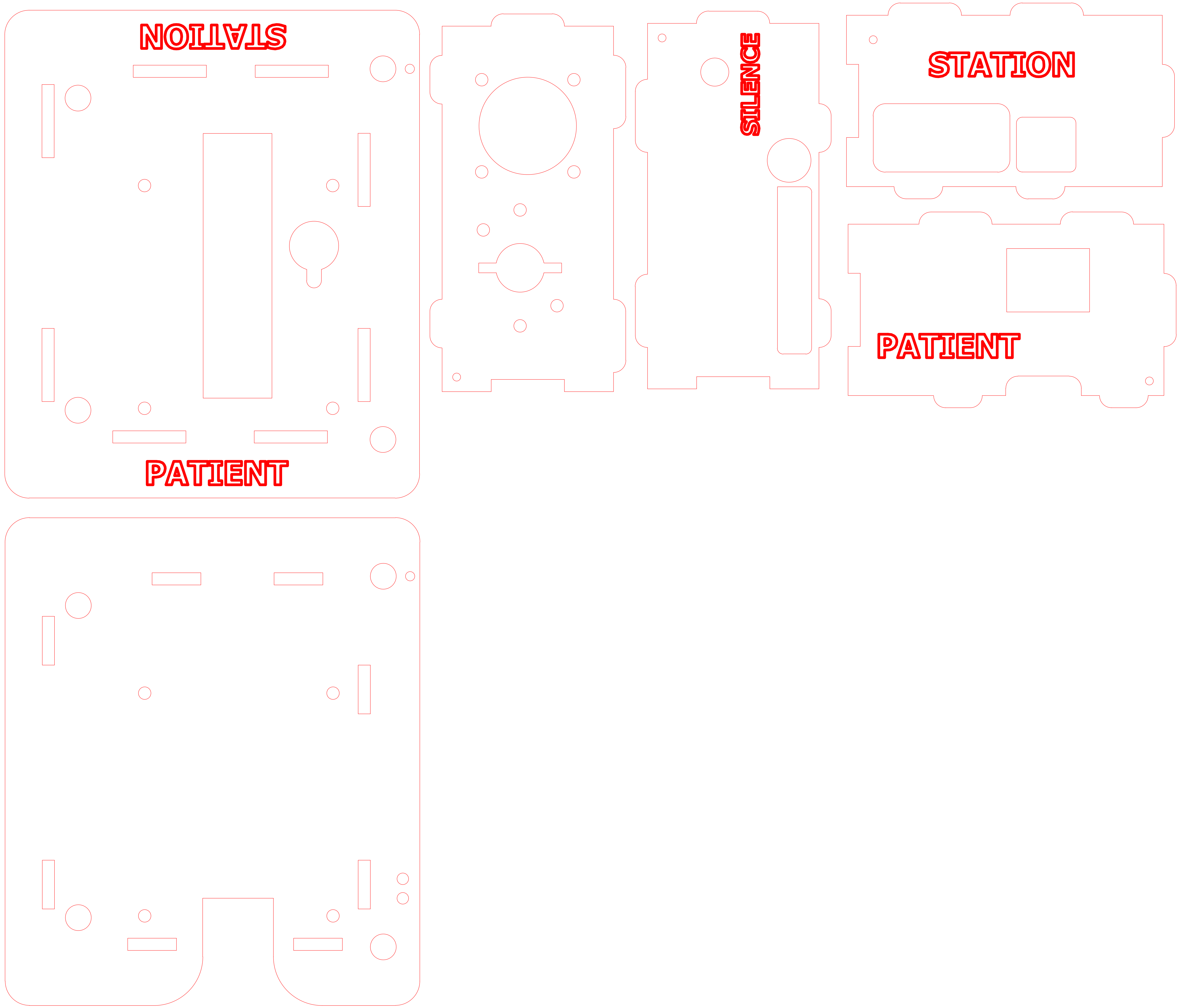

### ReadyToLaserCutFile_3XEnclosures.pdf

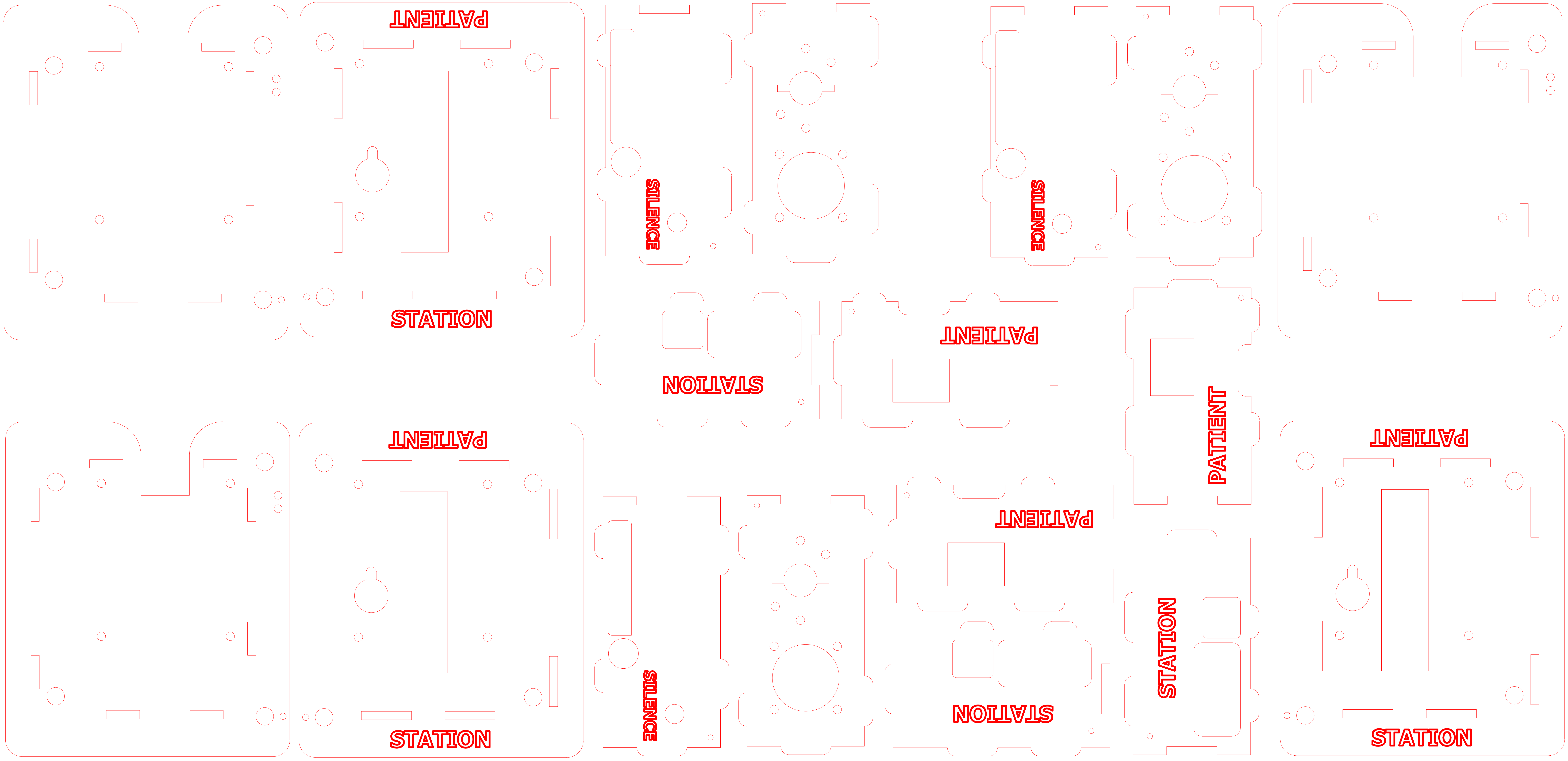

### SDP3x_6aM-LoadingBot.pdf

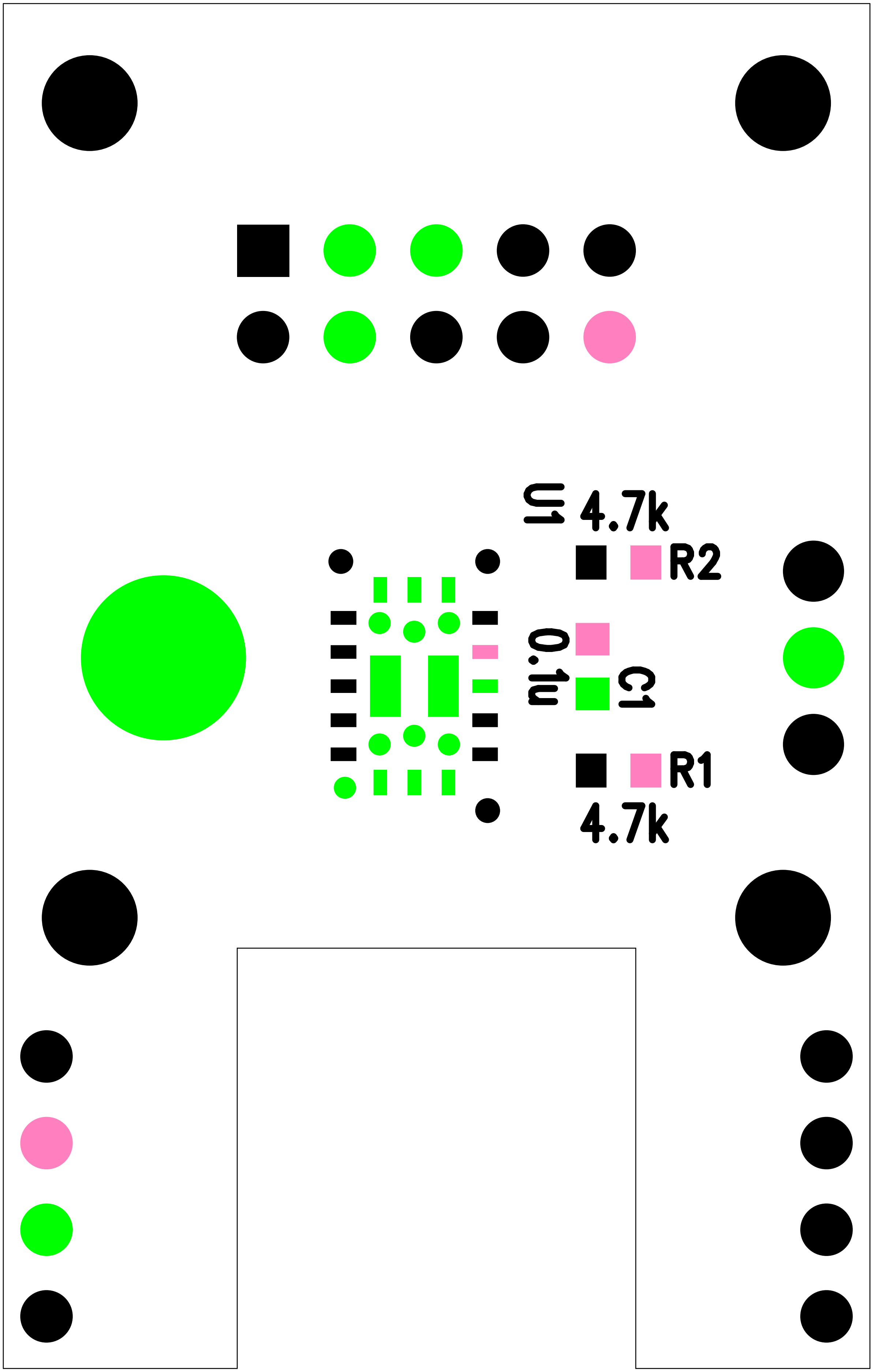


SDP3_6aM-Sensor.pcb - Fri Jun 26 16:45:42 2020

### SDP3x_6aM-LoadingTop.pdf

SDP3_6aM-Sensor.pcb - Fri Jun 26 16:45:04 2020

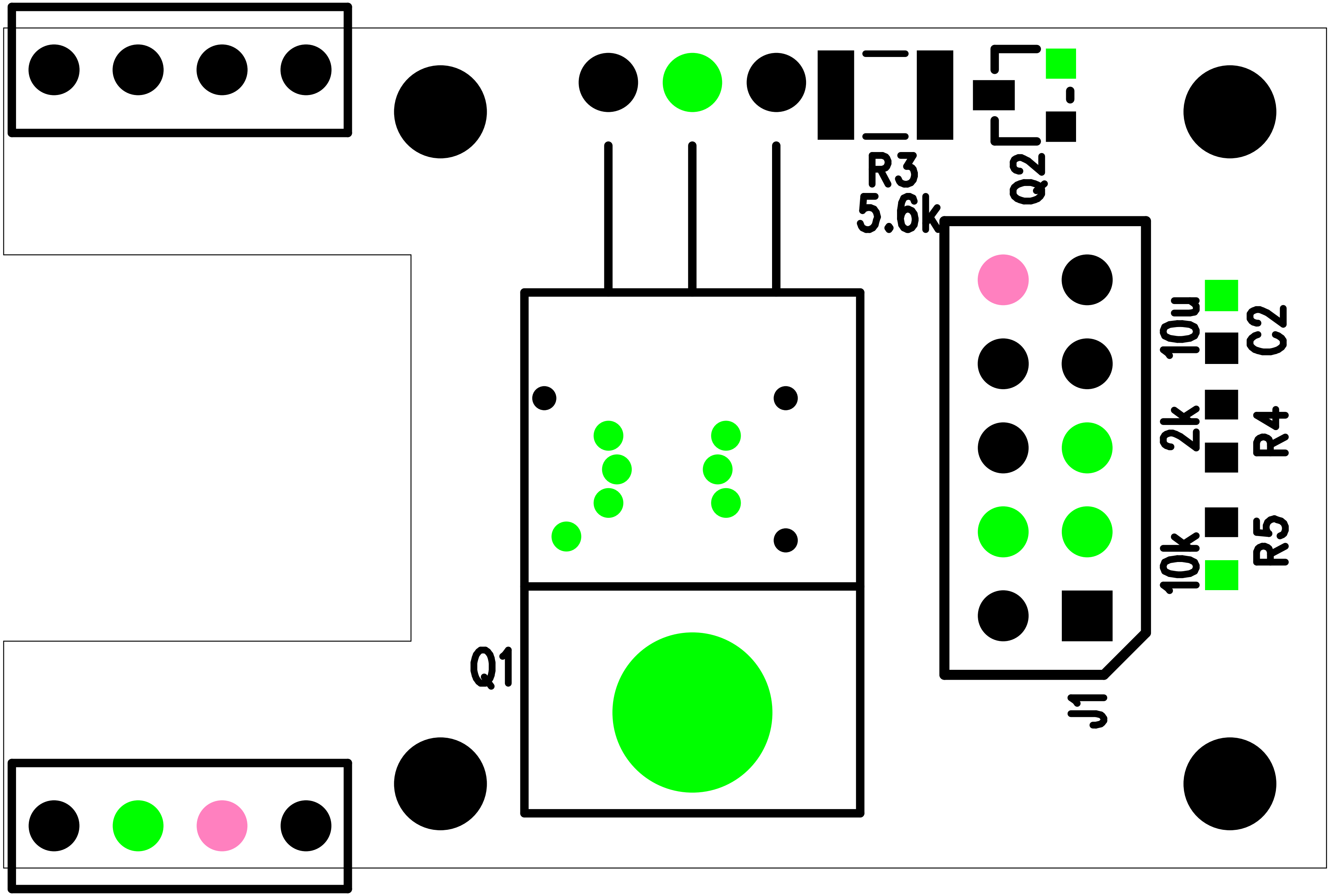

### SDP3x_6aM-SensorPCB.pdf

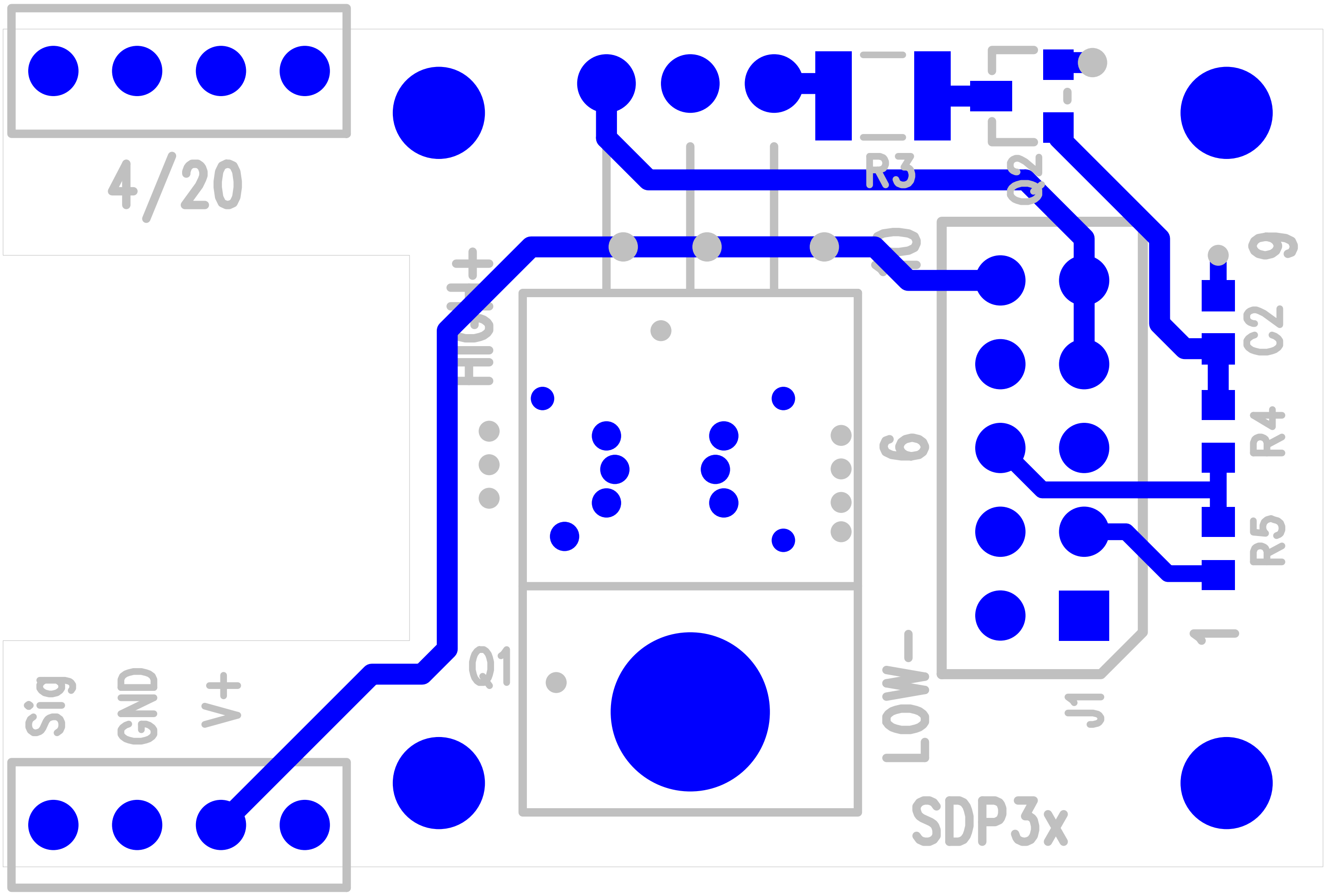

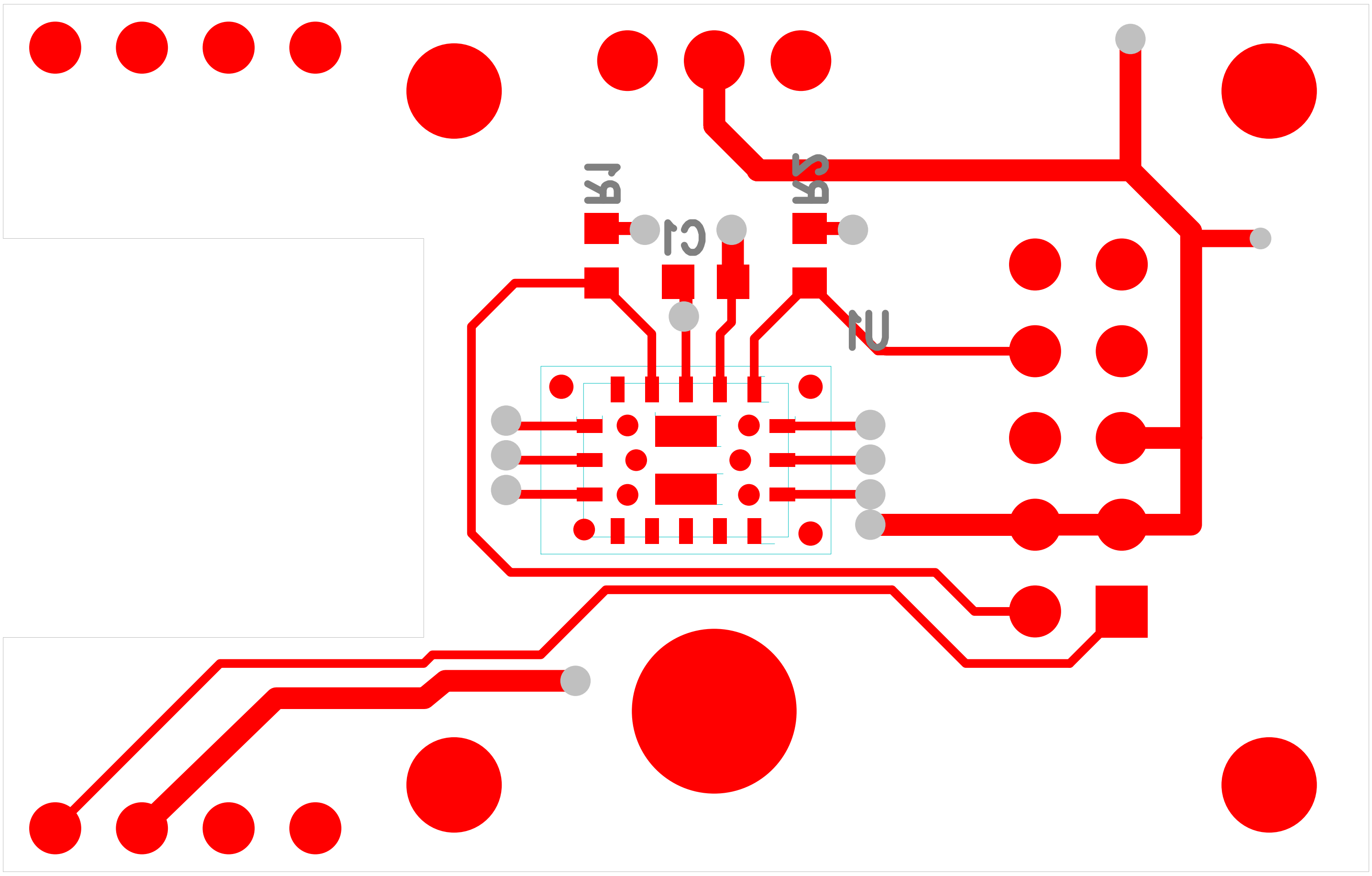

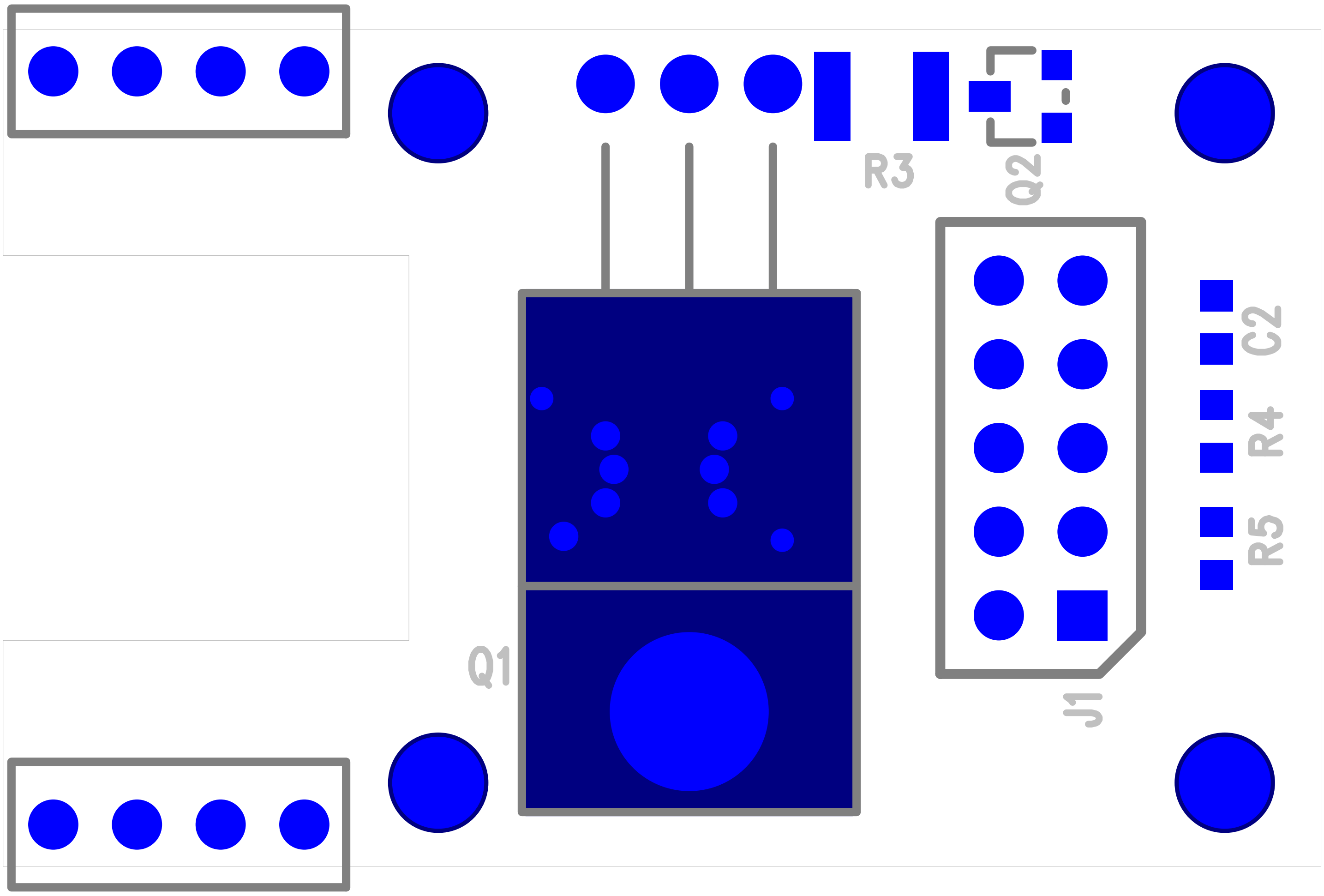

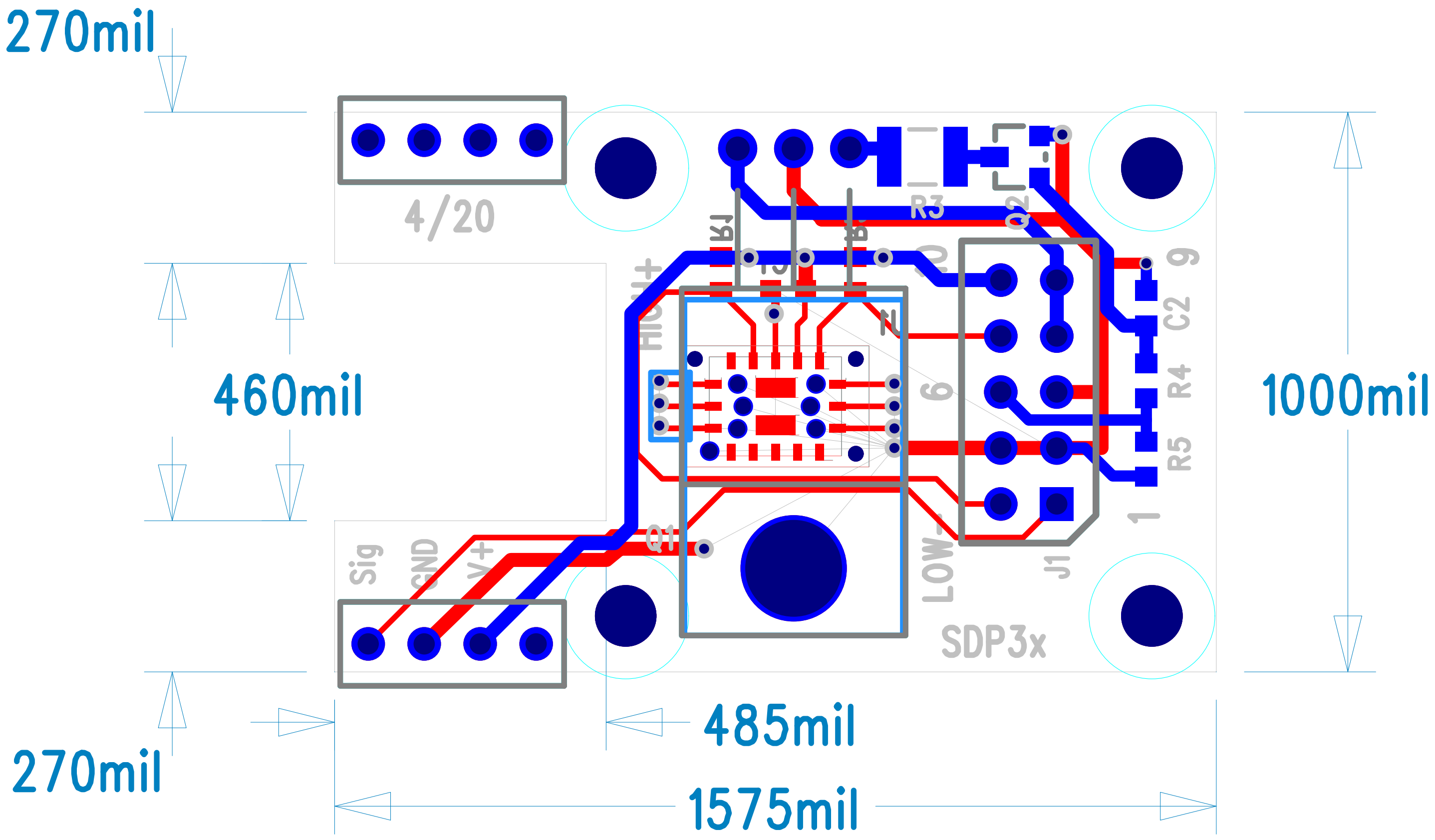

### SDP3x_6aM-SensorSCH.pdf

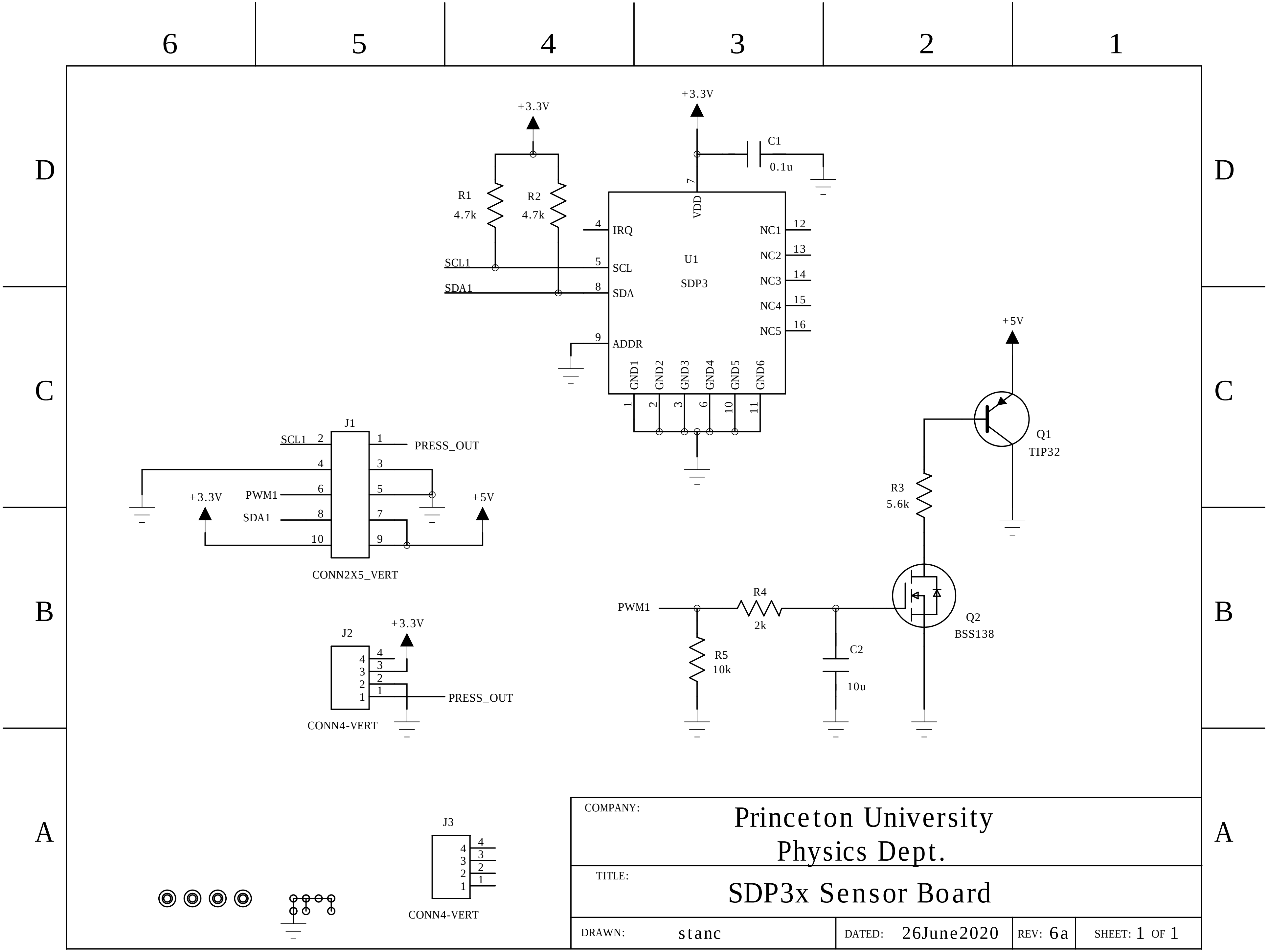
