## Supplementary Design File 2 for "Inexpensive multi-patient respiratory monitoring system for helmet ventilation during COVID-19 pandemic": CO2sensorBlock_2020-09-22.pdf

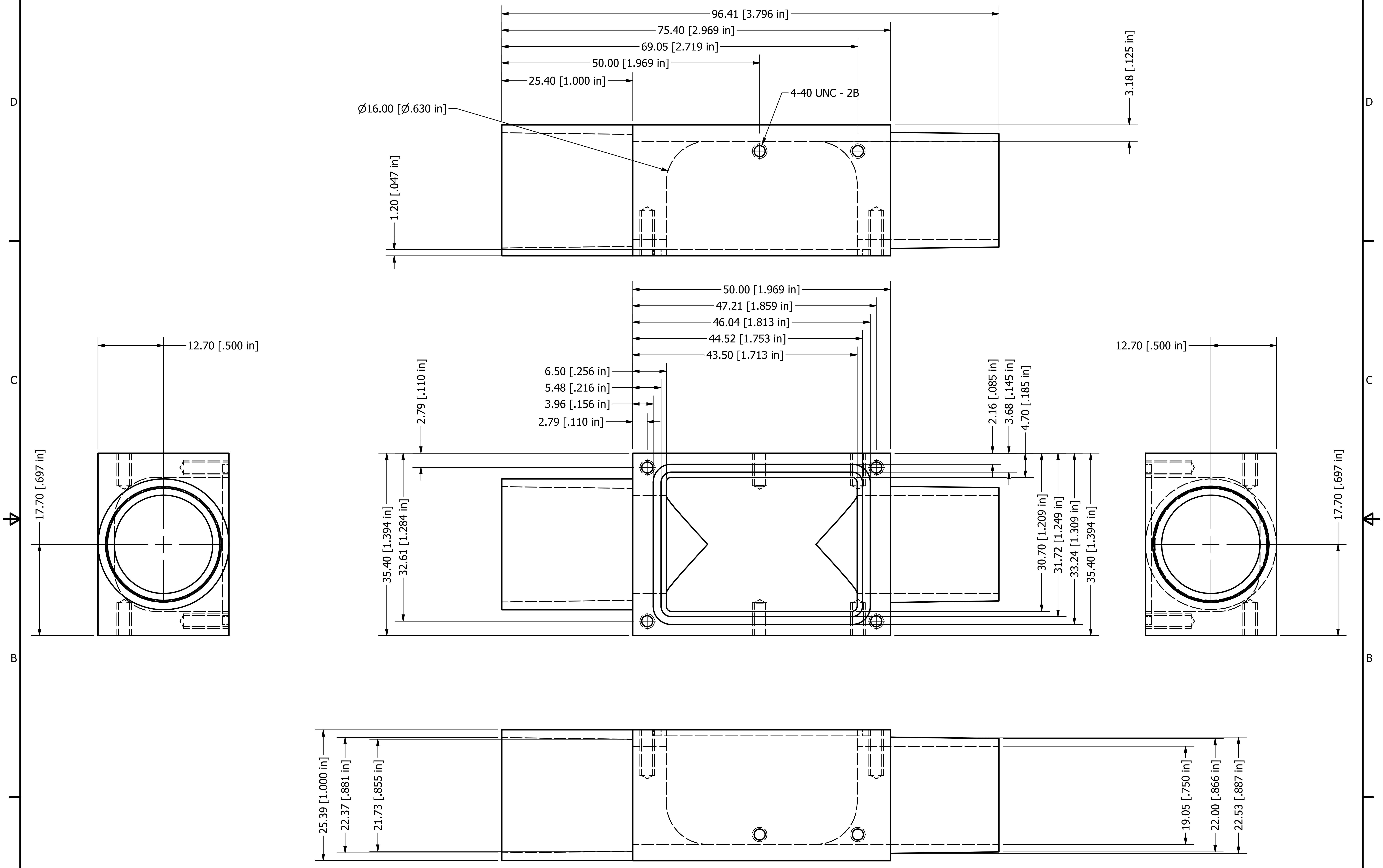


| | | | | | |
|---|---|---|---|---|---|
| DRAWN thiberge-admin | 10/21/2020 | | | | |
| CHECKED | | TITLE | | | |
| QA | | | | | |
| MFG | | | | | |
| APPROVED | | SIZE C | | DWG NO CO2sensorBlock_2020-09-22 | REV |
| | | SCALE | 2 : 1 | SHEET 1 OF 1 | |
