## Supplementary Design File 2 for "Inexpensive multi-patient respiratory monitoring system for helmet ventilation during COVID-19 pandemic": CO2sensorBlockTop_2020-06-29.pdf

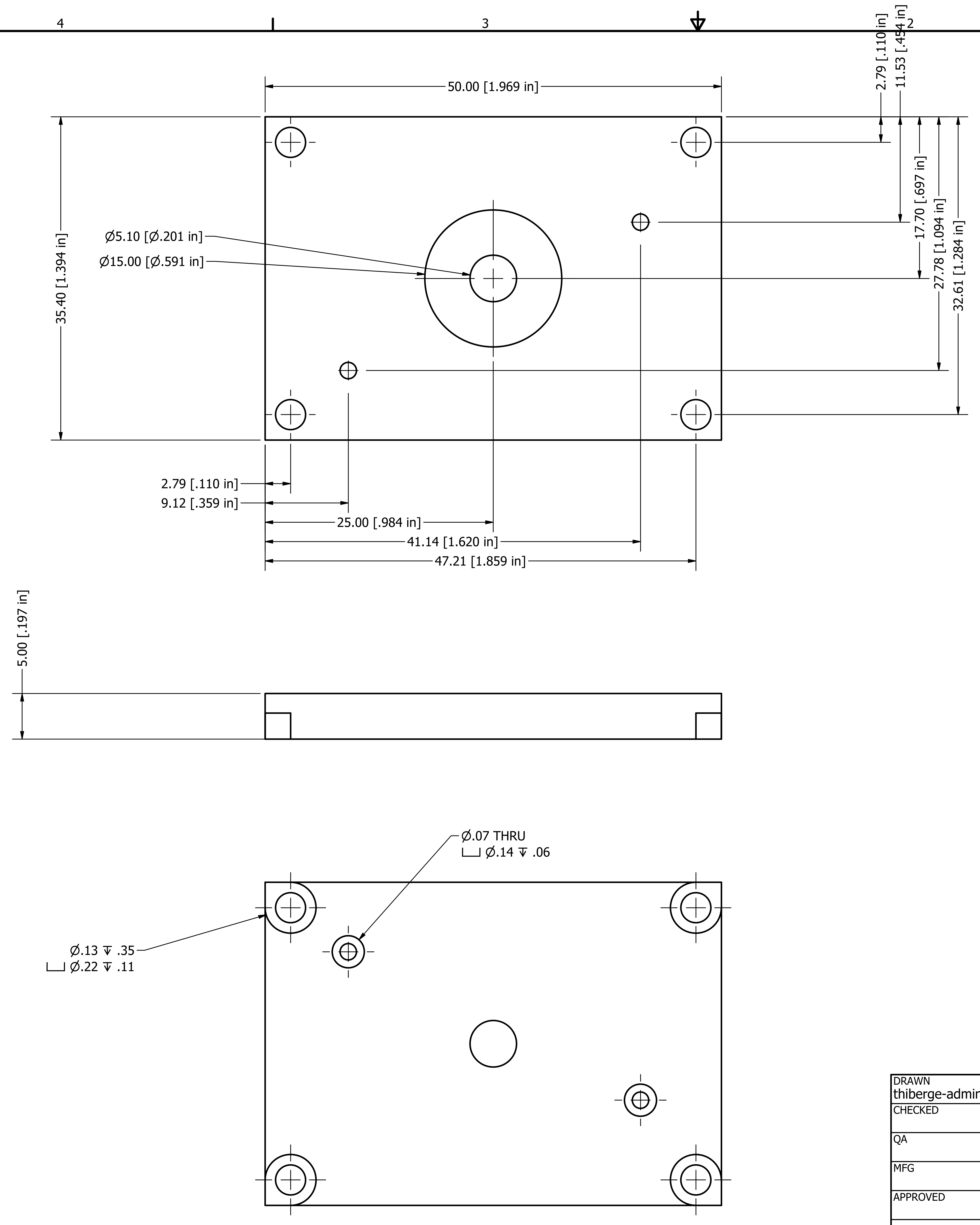


| | | |
|---|---|---|
| DRAWN thiberge-admin | 10/21/2020 | |
| CHECKED | | TITLE |
| QA | | |
| MFG | | |
| APPROVED | | |
| | | SIZE C · DWG NO CO2sensorBlockTop_2020-06-29 · REV |
| | | SCALE 3 : 1 · SHEET 1 OF 1 |
