## Supplementary Design File 2 for "Inexpensive multi-patient respiratory monitoring system for helmet ventilation during COVID-19 pandemic": IceHoney500_v2.pdf

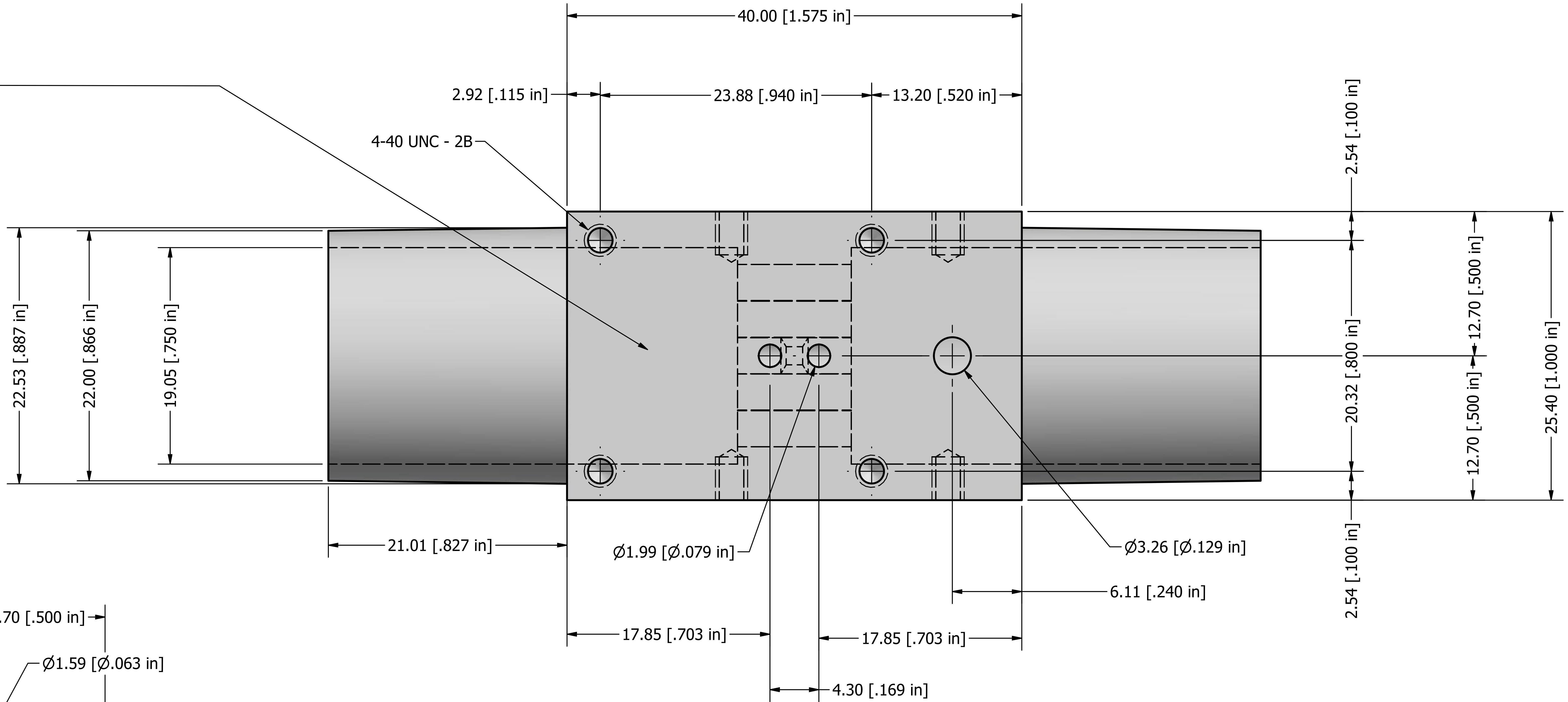


**Tolerance for the Nozzles**
Length .827" [-0.000 + 0 .010 ]
End diameter .866" +- .004"
Root diameter .887" +-.004 "

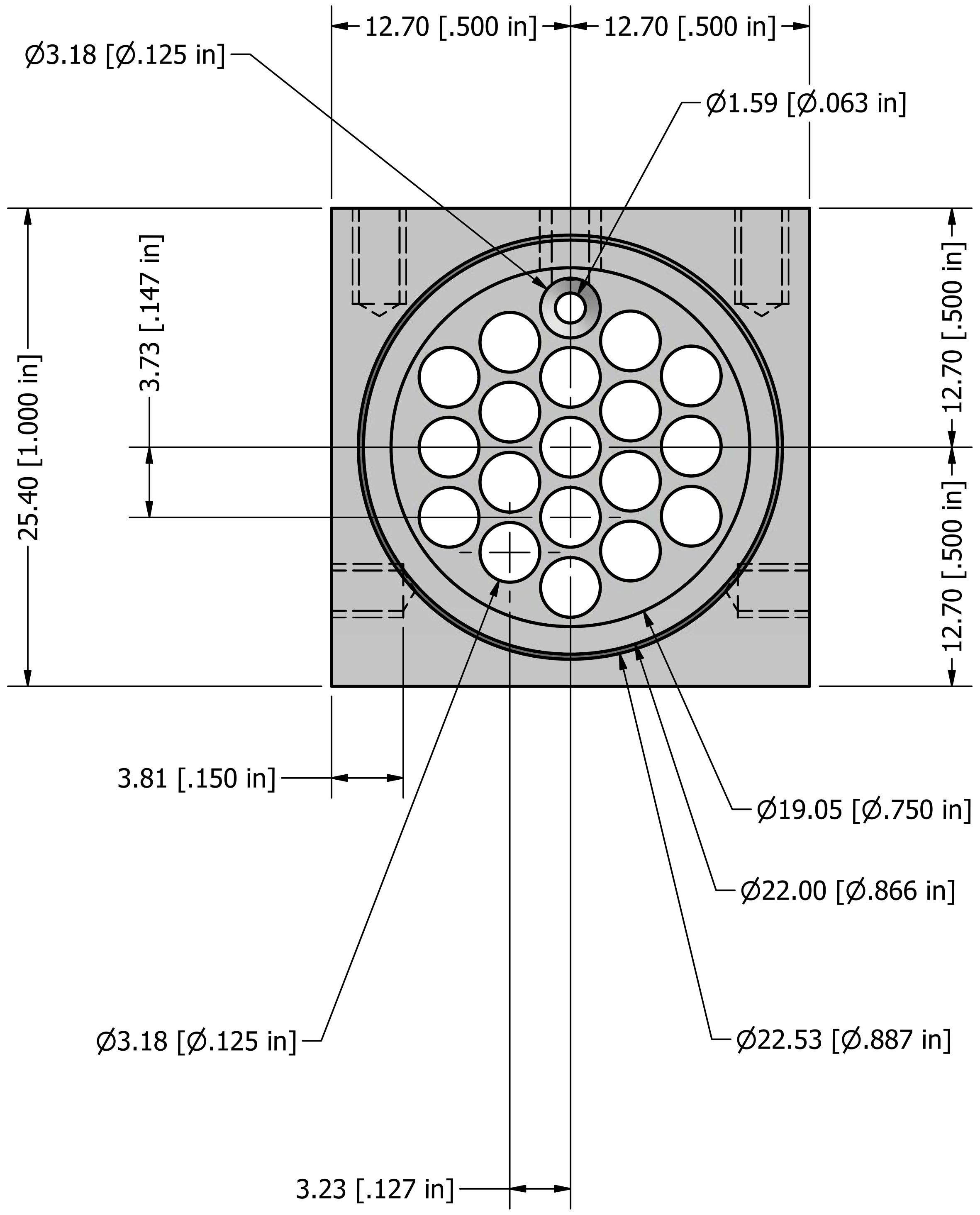


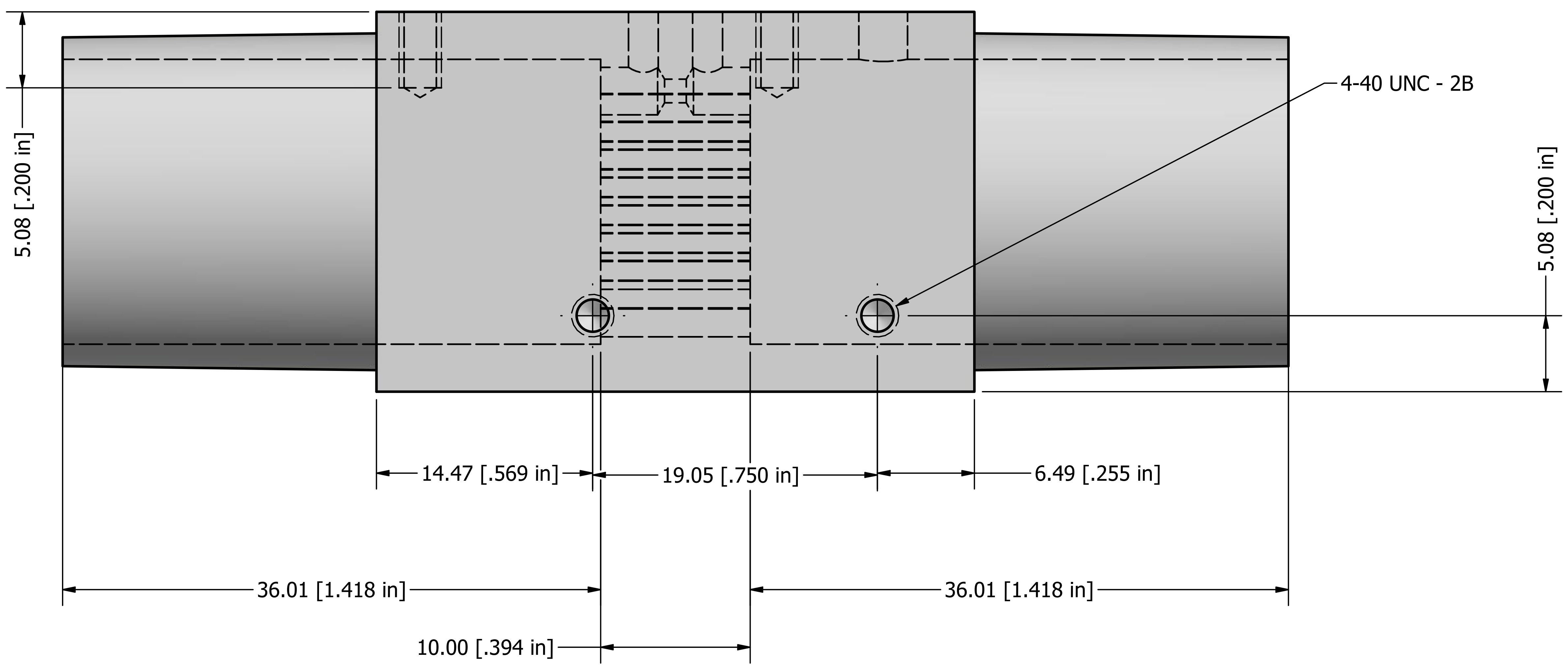


WHITE DELRIN

| | | |
|---|---|---|
| DRAWN Princeton Open Vent Monitor | 4/11/2020 | Princeton University |
| CHECKED | | TITLE |
| QA | | IceHoney500 |
| MFG | | |
| APPROVED | | SIZE C — DWG NO IceHoney500_v2 — REV |
| | | SCALE 3 : 1 — SHEET 1 OF 2 |

1.99mm [0.079"] is the diameter of Drill bit #47.

3.26mm is diameter of drill bit #30

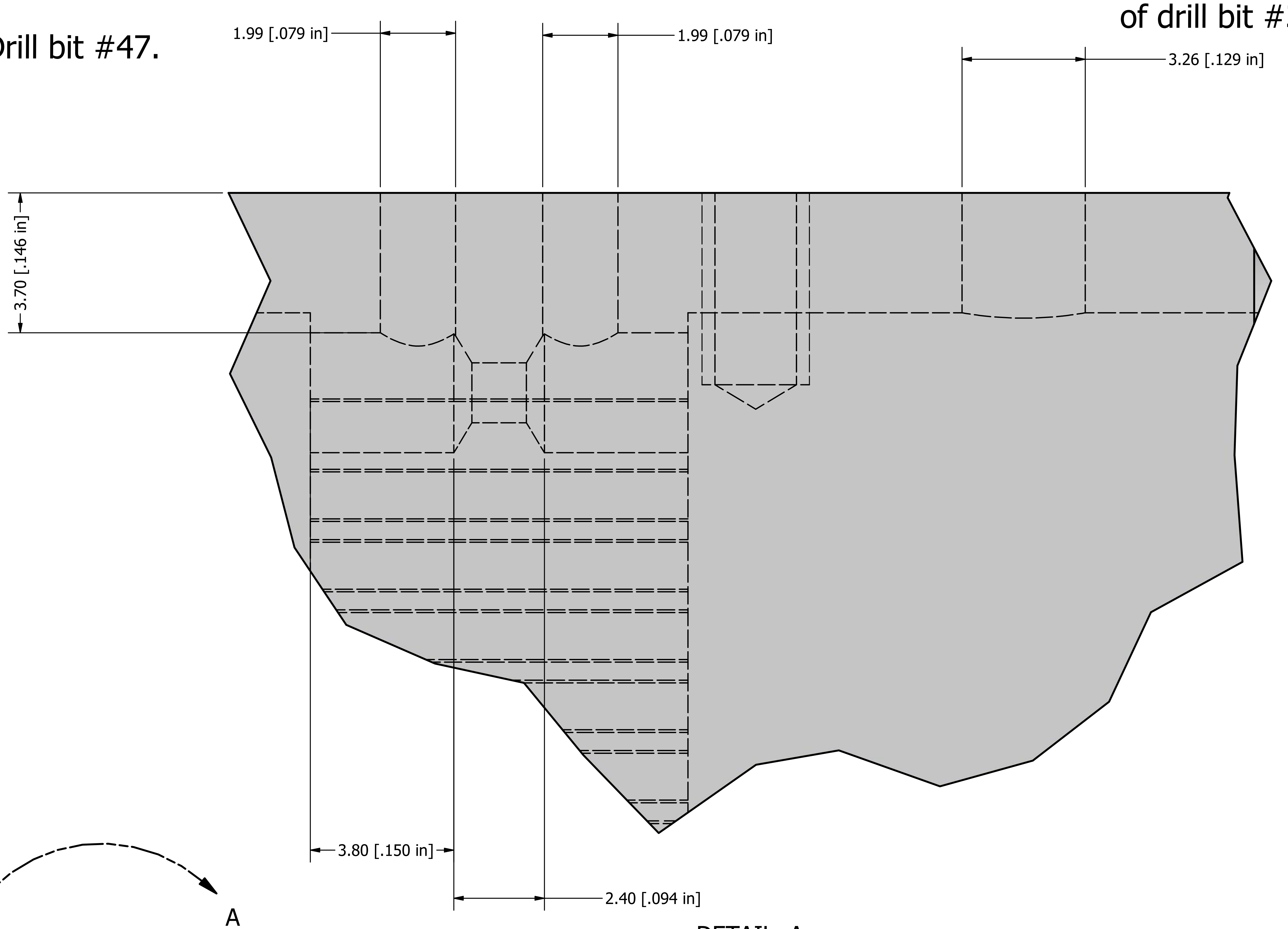


DETAIL A
SCALE 10:1

Page 2 of 2

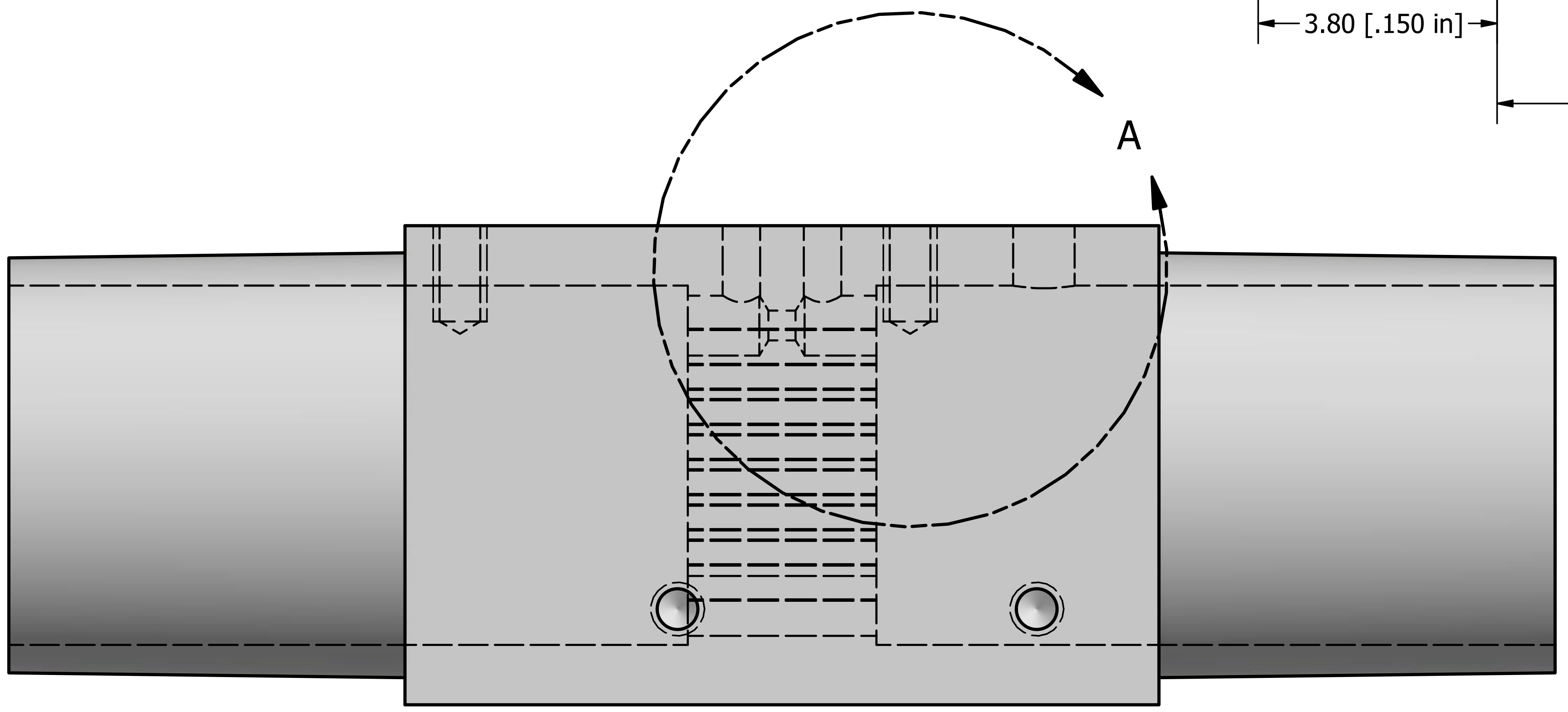


| | | |
|---|---|---|
| DRAWN Princeton Open Vent Me | 4/11/2020 | Princeton University |
| CHECKED | | |
| QA | | TITLE |
| MFG | | IceHoney500 |
| APPROVED | | |
| | | SIZE C / DWG NO IceHoney500_v2 / REV |
| | | SCALE 3 : 1 / SHEET 2 OF 2 |
