## Supplementary Git Repository for "Inexpensive multi-patient respiratory monitoring system for helmet ventilation during COVID-19 pandemic": config.txt

# For more options and information see
# http://rpf.io/configtxt
# Some settings may impact device functionality. See link above for details
#core_freq=250

# uncomment if you get no picture on HDMI for a default "safe" mode
#hdmi_safe=1

# uncomment this if your display has a black border of unused pixels visible
# and your display can output without overscan
disable_overscan=1

# uncomment the following to adjust overscan. Use positive numbers if console
# goes off screen, and negative if there is too much border
#overscan_left=16
#overscan_right=16
#overscan_top=16
#overscan_bottom=16

# uncomment to force a console size. By default it will be display's size minus
# overscan.
#framebuffer_width=1280
#framebuffer_height=720

# uncomment if hdmi display is not detected and composite is being output
#hdmi_force_hotplug=1

# uncomment to force a specific HDMI mode (this will force VGA)
#hdmi_group=1
#hdmi_mode=1

# uncomment to force a HDMI mode rather than DVI. This can make audio work in
# DMT (computer monitor) modes
#hdmi_drive=2

# uncomment to increase signal to HDMI, if you have interference, blanking, or
# no display
#config_hdmi_boost=4

# uncomment for composite PAL
#sdtv_mode=2

#uncomment to overclock the arm. 700 MHz is the default.
#arm_freq=800

# Uncomment some or all of these to enable the optional hardware interfaces
dtparam=i2c_arm=on,baudrate=200000
#dtparam=i2s=on
dtparam=spi=on

dtoverlay=i2c1,pins_2_3
dtoverlay=i2c6,pins_22_23
# Uncomment this to enable infrared communication.
#dtoverlay=gpio-ir,gpio_pin=17
#dtoverlay=gpio-ir-tx,gpio_pin=18

# Additional overlays and parameters are documented /boot/overlays/README

# Enable audio (loads snd_bcm2835)
dtparam=audio=on

[pi4]
# Enable DRM VC4 V3D driver on top of the dispmanx display stack
dtoverlay=vc4-fkms-v3d
max_framebuffers=2

[all]
#dtoverlay=vc4-fkms-v3d
# NOOBS Auto-generated Settings:
