## Supplementary Git Repository for "Inexpensive multi-patient respiratory monitoring system for helmet ventilation during COVID-19 pandemic": isc-dhcp-server

# Defaults for isc-dhcp-server (sourced by /etc/init.d/isc-dhcp-server)

# Path to dhcpd's config file (default: /etc/dhcp/dhcpd.conf).
#DHCPDv4_CONF=/etc/dhcp/dhcpd.conf
#DHCPDv6_CONF=/etc/dhcp/dhcpd6.conf

# Path to dhcpd's PID file (default: /var/run/dhcpd.pid).
#DHCPDv4_PID=/var/run/dhcpd.pid
#DHCPDv6_PID=/var/run/dhcpd6.pid

# Additional options to start dhcpd with.
#	Don't use options -cf or -pf here; use DHCPD_CONF/ DHCPD_PID instead
#OPTIONS=""

# On what interfaces should the DHCP server (dhcpd) serve DHCP requests?
#	Separate multiple interfaces with spaces, e.g. "eth0 eth1".
INTERFACESv4="eth0"
INTERFACESv6=""
