## Supplementary Git Repository for "Inexpensive multi-patient respiratory monitoring system for helmet ventilation during COVID-19 pandemic": device_json_to_csv.py

```
#!/usr/bin/env python3

from processor.argparse import ArgumentParser
from processor.config import config

parser = ArgumentParser(log_dir=None, log_stem=None)
parser.add_argument("input", help="Input single-line json file")
parser.add_argument("output", help="Output csv file")
args = parser.parse_args()

import json
import csv

flow_scale = config["device"]["flow"]["scale"].get(float)
flow_offset = config["device"]["flow"]["offset"].get(float)
pressure_scale = config["device"]["pressure"]["scale"].get(float)
pressure_offset = config["device"]["pressure"]["offset"].get(float)

with open(args.input) as fin, open(args.output, "w") as fout:
    writer = csv.writer(fout, delimiter=",")

    # Heading
    writer.writerow(["time", "Flow", "Pressure", "Current_temp", "DC_temp"])

    for line in fin:
        d = json.loads(line)
        writer.writerow(
            [
                d["t"],
                d["F"] * flow_scale - flow_offset,
                d["P"] * pressure_scale - pressure_offset,
                d.get("C"),
                d.get("D"),
            ]
        )
```
