## Supplementary Git Repository for "Inexpensive multi-patient respiratory monitoring system for helmet ventilation during COVID-19 pandemic": device_json_to_plot.py

```
#!/usr/bin/env python3

import argparse

parser = argparse.ArgumentParser()
parser.add_argument("input", help="Input single-line json file")
parser.add_argument("--output", default=None, help="Output png/pdf file")
parser.add_argument(
    "--interactive",
    action="store_true",
    help="Pop up a window to show the plot interactively",
)
parser.add_argument(
    "--no-curtemp",
    action="store_true",
    help="Don't print out current temperature values found in the data stream",
)
parser.add_argument(
    "--no-dctemp",
    action="store_true",
    help="Don't print out dc temperature values found in the data stream",
)
args = parser.parse_args()


import json
import warnings
import datetime

import matplotlib
import matplotlib.pyplot as plt

tP = []
tF = []
P = []
F = []
tmin, tmax, tprev = None, None, None

with open(args.input) as fin:
    for line in fin:
        try:
            d = json.loads(line)
        except json.JSONDecodeError:
            warnings.warn("line is not valid JSON: {}".format(repr(line)))
            break

        if d.get("v", None) != 1:
            warnings.warn(
                "key 'v' (version) is missing or not equal to 1: {}".format(
                    d.get("v", None)
                )
            )

        if "t" not in d or not isinstance(d["t"], (int, float)):
            warnings.warn(
                "key 't' (time) is missing or not a number: {}".format(d.get("t", None))
            )

        else:
            t = datetime.datetime.fromtimestamp(d["t"] / 1000.0)
            if tmin is None or t < tmin:
                tmin = t
            if tmax is None or t > tmax:
                tmax = t

            if "P" not in d or not isinstance(d["P"], (int, float)):
                warnings.warn(
                    "key 'P' (ADC avg pressure) is missing or not a number: {}".format(
                        d.get("P", None)
                    )
                )
            else:
                tP.append(t)
                P.append(d["P"])

            if "F" not in d or not isinstance(d["F"], (int, float)):
                warnings.warn(
                    "key 'F' (flow; a.k.a. delta pressure) is missing or not a number: {}".format(
                        d.get("F", None)
                    )
                )
            else:
                tF.append(t)
                F.append(d["F"])

            if tprev is not None and tprev >= t:
                warnings.warn("times are not strictly monotonic")
            tprev = t

        if "C" in d:
            print(
                "time: {}, cur temperature: {}".format(
                    t.isoformat().replace("T", " "), d["C"]
                )
            )
        if "D" in d:
            print(
                "time: {}, dc temperature:  {}".format(
                    t.isoformat().replace("T", " "), d["D"]
                )
            )

if tmin is None or tmax is None:
    warnings.warn("no time data in file")
else:
    print("starting time: {}".format(tmin.isoformat().replace("T", " ")))
    print("stopping time: {}".format(tmax.isoformat().replace("T", " ")))

    fig, (ax1, ax2) = plt.subplots(2, 1, figsize=(15, 10))
    ax1.plot(tP, P)
    ax1.set_xlabel("time")
    ax1.set_ylabel("ADC avg pressure")
    ax2.plot(tF, F)
    ax2.set_xlabel("time")
    ax2.set_ylabel("flow; a.k.a. delta pressure")
    if args.interactive:
        print("plotting interactively")
        with matplotlib.rc_context(rc={"interactive": False}):
            plt.show()
    else:
        print("to plot interactively, use the --interactive flag")

    if args.output is not None:
        print("saving as '{}'".format(args.output))
        fig.savefig(args.output)
    else:
        print("to save a file, pass '--output file.png' (or PDF) on the commandline")
```
