## Supplementary Git Repository for "Inexpensive multi-patient respiratory monitoring system for helmet ventilation during COVID-19 pandemic": device_json_to_socket.py

```
#!/usr/bin/env python3

import argparse
import zmq
import contextlib
import time
import json

parser = argparse.ArgumentParser()
parser.add_argument("input", help="Input single-line json file")
parser.add_argument(
    "--repeat", default=1, type=int, help="Number of times to repeat, 0 for forever"
)
parser.add_argument("--ff", type=int, help="Fast forward until this timestamp is found")
args = parser.parse_args()


rate = 50  # Hz


@contextlib.contextmanager
def controlled_time(t):
    tic = time.monotonic()
    yield
    toc = time.monotonic()
    remaining = t - (tic - toc)
    time.sleep(max(remaining, 0))


with zmq.Context() as ctx, ctx.socket(zmq.PUB) as pub_socket:
    pub_socket.bind("tcp://*:5556")

    with open(args.input) as f:
        for line in f:
            if args.ff and args.ff > json.loads(line)["t"]:
                continue
            with controlled_time(1 / rate):
                pub_socket.send_string(line)

    before = json.loads(line)
    timestamp = before["t"]

    i = 1
    while args.repeat == 0 or i < args.repeat:
        print(f"Re-serving: {i}")

        with open(args.input) as f:
            for line in f:
                with controlled_time(1 / rate):
                    current = json.loads(line)
                    timestamp += 20
                    current["t"] = timestamp
                    pub_socket.send_json(current)

        i += 1
```
