## Supplementary Git Repository for "Inexpensive multi-patient respiratory monitoring system for helmet ventilation during COVID-19 pandemic": device_json_to_timeseries.py

```
#!/usr/bin/env python3

import argparse

parser = argparse.ArgumentParser()
parser.add_argument("input", help="Input single-line json file")
parser.add_argument(
    "--drop-header",
    action="store_true",
    help="If enabled, the header line will be dropped",
)
parser.add_argument(
    "--deglitch-pressure",
    action="store_true",
    help="If enabled, pass pressure through pressure_deglitch_smooth",
)
parser.add_argument(
    "--start-time-at-zero",
    action="store_true",
    help="If enabled, the timeseries starts at zero",
)
args = parser.parse_args()

import json

import numpy

from processor.config import config
from processor import analysis
from processor.flow_calibrator import flow_calibrator

caliber = flow_calibrator()

flow_scale = config["device"]["flow"]["scale"].as_number()
flow_offset = config["device"]["flow"]["offset"].as_number()
pressure_scale = config["device"]["pressure"]["scale"].as_number()
pressure_offset = config["device"]["pressure"]["offset"].as_number()

time = []
pressure = []
flow = []

starttime = None
with open(args.input) as fin:
    for line in fin:
        try:
            j = json.loads(line)
        except json.JSONDecodeError:
            continue
        if j.get("v", None) != 1:
            continue
        if "t" not in j or not isinstance(j["t"], (int, float)):
            continue
        if "P" not in j or not isinstance(j["P"], (int, float)):
            continue
        if "F" not in j or not isinstance(j["F"], (int, float)):
            continue

        # FIXME: this will be a lookup function!
        # I could not extract this correction from the main codebase; we'll
        # have to fix it both here and there.
        t = j["t"] / 1000.0
        p = j["P"] * pressure_scale - pressure_offset
        # this was the old flow calibration
        # fO = math.copysign(abs(j["F"]) ** (4 / 7), j["F"]) * flow_scale - flow_offset
        f = caliber.Q(j["F"])

        if starttime is None:
            starttime = t
        if args.start_time_at_zero:
            t -= starttime

        time.append(t)
        pressure.append(p)
        flow.append(f)

if args.deglitch_pressure:
    pressure = analysis.pressure_deglitch_smooth(numpy.array(pressure))

volume = analysis.flow_to_volume(
    numpy.array(time),
    None,
    numpy.array(flow),
    None,
    critical_frequency=0.004,
)
volume -= numpy.min(volume)

minbias_volume = analysis.flow_to_volume(
    numpy.array(time),
    None,
    numpy.array(flow),
    None,
    critical_frequency=0.0004,
)
minbias_volume -= numpy.min(minbias_volume)

if not args.drop_header:
    print(
        "     time (sec), pressure (cm H2O), flow (L/min), volume (mL), minbias volume (mL)"
    )
for t, p, f, v, mv in zip(time, pressure, flow, volume, minbias_volume):
    print(f"{t:15.3f}, {p:17.4f}, {f:12.4f}, {v:11.4f}, {mv:19.4f}")
```
