## Supplementary Git Repository for "Inexpensive multi-patient respiratory monitoring system for helmet ventilation during COVID-19 pandemic": device_loop.py

```
#!/usr/bin/env python3


import argparse

parser = argparse.ArgumentParser(formatter_class=argparse.ArgumentDefaultsHelpFormatter)
parser.add_argument("--name", default="data.out", help="Base filename to record to")
parser.add_argument("--co2", action="store_true", help="Also read CO2 sensor")
parser.add_argument(
    "--dir", help="Directory to record to (device_log will be appended)"
)
arg = parser.parse_args()


# hardware interfaces:
# SDP3 diff pressure sensor - I2C handled by pigpio.pi()
# MCP3008 ADC readings - SPI handled by spidev.SpiDev()

import time
import signal
import pigpio
import spidev
import struct
import json
import zmq
import threading
import shutil
from datetime import datetime
from pathlib import Path
from typing import Optional, TextIO, Iterator, TYPE_CHECKING, Dict, Any, List
from contextlib import contextmanager, ExitStack

if TYPE_CHECKING:
    from typing_extensions import Final

DIR = Path(__file__).parent.resolve()

GB = 1_000_000_000
HOUR = 60 * 60

if arg.dir:
    directory = Path(arg.dir).expanduser().resolve() / "device_log"
else:
    directory = DIR / "device_log"

if arg.name:
    print("Logging to", directory)
    directory.mkdir(parents=True, exist_ok=True)

ReadoutHz: "Final" = 50.0
oversampleADC: "Final" = 4

NReadoutTemp: "Final" = 50 * oversampleADC
NReadout: "Final" = 0
nbytesPN: "Final" = 18
nbytesSF: "Final" = 9
nbytesTEMP: "Final" = 6
nbytesDP: "Final" = 3
hystTEMP: "Final" = 1.0  # change temperature is difference is higher than hystTemp
minTEMP: "Final" = 42.0
operTEMP: "Final" = 45.0
nbytesFW: "Final" = 3
nbytesRDY: "Final" = 3
nbytesCO2: "Final" = 18
maxTEMP: "Final" = 48.0
dcSTEP: "Final" = 1
dcMAX: "Final" = 9000
dcRANGE: "Final" = 10000
dcSTROBE: "Final" = 1 * NReadoutTemp
pinPWM: "Final" = 13


def sgn(a: float) -> float:
    return (a > 0) - (a < 0)


chanMP3V5004: "Final" = 0

DEVICE_SDP3: "Final" = 0x21  # grounded ADDR pin
DEVICE_SCD3: "Final" = 0x61  # grounded ADDR pin


def getADC(spi: spidev.SpiDev, channel: int) -> int:
    # Check channel valid
    # if ((channel > 7) or (channel < 0)):
    #  return -1
    # Perform SPI (spiMCP3008.xfer2 keeps CS asserted)
    r = spi.xfer2([1, (8 + channel) << 4, 0])
    # Reformat
    adcOut = ((r[1] & 0xF) << 8) + r[2]
    return adcOut


@contextmanager
def pi_cleanup() -> Iterator[pigpio.pi]:
    """
    This ensures that the π is closed, with the heater turned off, even if an error happens.
    """
    pi = pigpio.pi()
    try:
        yield pi
    finally:
        pi.set_PWM_dutycycle(pinPWM, 0)
        pi.stop()


def frequency(length: float, event: threading.Event) -> Iterator[None]:
    """
    This pauses as long as needed after running
    """
    while not event.is_set():
        start_time = time.monotonic()
        yield
        diff = time.monotonic() - start_time
        left = length - diff
        if left > 0:
            event.wait(left)


def open_next(mypath: Path) -> TextIO:
    """
    Open the next available file
    """
    while True:
        dt = datetime.now().strftime("%Y%m%d_%H%M%S")
        name = "{n}_{dt}{s}".format(n=mypath.stem, dt=dt, s=mypath.suffix)
        new_file_path = mypath.with_name(name)
        try:
            return open(str(new_file_path), "x")
        except FileExistsError:
            time.sleep(1)


def delete_oldest(mypath: Path, size: int) -> None:
    """
    Delete the oldest file(s) in a directory. Give the filename you want to create. The
    size parameter tells you how much to delete.
    """

    path_list = mypath.parent.glob("*." + mypath.suffix)
    # Sort by creation time
    paths = sorted(path_list, key=lambda x: x.stat().st_ctime)

    total_deleted = 0

    for path in paths:
        total_deleted += path.stat().st_size
        path.unlink()
        if total_deleted > size:
            break
    else:
        print("Warning: all log files deleted and still not enough space reclaimed!")


def read_loop(
    spi: spidev.SpiDev,
    pi: pigpio.pi,
    hSDP3: "pigpio.Handle",
    hSCD3: "Optional[pigpio.Handle]",
    running: threading.Event,
    myfile: Optional[TextIO],
) -> None:
    if hSCD3 is not None:
        # SCD3 handle
        print("SCD3 handle {}".format(hSCD3))

        # first issue stop command
        pi.i2c_write_device(hSCD3, [0x01, 0x04])

        time.sleep(0.02)

        # read firmware version
        pi.i2c_write_device(hSCD3, [0xD1, 0x00])
        dataSCD3 = pi.i2c_read_device(hSCD3, nbytesFW)
        bdataSCD3 = dataSCD3[1]
        if dataSCD3[0] != nbytesFW:
            return
        fw = (bdataSCD3[0] << 8) | bdataSCD3[1]
        print("fw", fw)

        # start continuous measurement without without pressure compensation
        pi.i2c_write_device(hSCD3, [0x00, 0x10, 0x00, 0x00, 0x81])

        time.sleep(0.02)

        # read first measurement
        pi.i2c_write_device(hSCD3, [0x03, 0x00])
        dataSCD3 = pi.i2c_read_device(hSCD3, nbytesCO2)
        bdataSCD3 = dataSCD3[1]
        if dataSCD3[0] != nbytesCO2:
            return
        (co2,) = struct.unpack(
            ">f", bytes([bdataSCD3[0], bdataSCD3[1], bdataSCD3[3], bdataSCD3[4]])
        )
        (tp,) = struct.unpack(
            ">f", bytes([bdataSCD3[6], bdataSCD3[7], bdataSCD3[9], bdataSCD3[10]])
        )
        (hd,) = struct.unpack(
            ">f", bytes([bdataSCD3[12], bdataSCD3[13], bdataSCD3[14], bdataSCD3[15]])
        )
        print(
            "CO2 concentration={:.2f} Temperature={:.2f} Humidity={:.2f}".format(
                co2, tp, hd
            )
        )

    # SDP3 handle
    print("SDP3 handle {}".format(hSDP3))

    # first issue stop command
    pi.i2c_write_device(hSDP3, [0x3F, 0xF9])

    time.sleep(0.5)

    pi.i2c_write_device(hSDP3, [0x36, 0x7C])
    pi.i2c_write_device(hSDP3, [0xE1, 0x02])
    dataSDP3 = pi.i2c_read_device(hSDP3, nbytesPN)
    bdataSDP3 = dataSDP3[1]
    if dataSDP3[0] != nbytesPN:
        return

    pnmsw = int.from_bytes(bdataSDP3[0:2], byteorder="big", signed=False)
    pnlsw = int.from_bytes(bdataSDP3[3:5], byteorder="big", signed=False)
    pn = (pnmsw << 16) | pnlsw
    snmmsw = int.from_bytes(bdataSDP3[6:8], byteorder="big", signed=False)
    snmsw = int.from_bytes(bdataSDP3[9:11], byteorder="big", signed=False)
    snlsw = int.from_bytes(bdataSDP3[12:14], byteorder="big", signed=False)
    snllsw = int.from_bytes(bdataSDP3[15:17], byteorder="big", signed=False)
    sn = (snmmsw << 48) | (snmsw << 32) | (snlsw << 16) | snllsw
    print("pn", hex(pn), "sn", hex(sn))

    # start continuous readout with averaging with differential pressure temperature compensation
    pi.i2c_write_device(hSDP3, [0x36, 0x15])

    time.sleep(0.020)

    # get initial values of differential pressure, temperature and the differential pressure scale factor
    dataSDP3 = pi.i2c_read_device(hSDP3, nbytesSF)
    if dataSDP3[0] != nbytesSF:
        return
    bdataSDP3 = dataSDP3[1]
    dp = int.from_bytes(bdataSDP3[0:2], byteorder="big", signed=True)
    temp = (bdataSDP3[3] << 8) | bdataSDP3[4]
    dpsf = float((bdataSDP3[6] << 8) | bdataSDP3[7])
    print("Time", time.time(), "dp", dp, "temp", temp, "dpsf", dpsf)

    # setup PWM heating
    dcTEMP = 3300
    curTEMP = temp / 200.0
    pi.set_PWM_range(pinPWM, dcRANGE)
    pi.set_PWM_dutycycle(pinPWM, dcTEMP)
    dcFREQ = pi.get_PWM_frequency(pinPWM)
    print(
        "time: {} dp / dpsf: {:.4f} temp: {:.4f}".format(
            time.time() * 1000, dp / dpsf, temp / 200.0
        )
    )
    print(
        "PWM settings: Range = {} Freq = {} Step = {} Strobe = {}".format(
            dcRANGE, dcFREQ, dcSTEP, dcSTROBE
        )
    )

    # initialize take-back-half servo
    last_errorTEMP = 1.0  # + sign helps with cold start
    first_crossTEMP = True
    dcTEMP_at_cross: int = 0

    # initialize ADC sampling
    NReadout = 0
    ADCsamples: List[float] = []

    for _ in frequency(1.0 / (ReadoutHz * oversampleADC), running):  # Readout in Hz

        NReadout += 1
        tmpADC = getADC(spi, chanMP3V5004)
        ADCsamples.append(tmpADC)

        # Continue if we are currently in oversampling
        if (NReadout % oversampleADC) != 0:
            continue

        ADCavg = 0.0
        if len(ADCsamples):
            ADCavg = sum(ADCsamples) / len(ADCsamples)
        ADCsamples = []

        ts = int(1000 * time.monotonic())

        if (NReadout % NReadoutTemp) == 0:
            nbytes = nbytesTEMP
        else:
            nbytes = nbytesDP

        tmpdataSDP3 = pi.i2c_read_device(hSDP3, nbytes)
        if tmpdataSDP3[0] != nbytes:
            return
        btmpdataSDP3 = tmpdataSDP3[1]
        tmpdp = int.from_bytes(btmpdataSDP3[0:2], byteorder="big", signed=True)

        d = {"v": 1, "t": ts, "P": ADCavg, "F": tmpdp}  # type: Dict[str, Any]

        if len(btmpdataSDP3) == nbytesTEMP:
            tmptemp = ((btmpdataSDP3[3] << 8) | btmpdataSDP3[4]) / 200.0
            print(ts, tmptemp, dcTEMP)
            if (NReadout % dcSTROBE) == 0:
                # Take-back-half algorithm
                errorTEMP = operTEMP - curTEMP
                if sgn(errorTEMP) != sgn(last_errorTEMP):
                    if first_crossTEMP:
                        first_crossTEMP = False
                    else:
                        dcTEMP = (dcTEMP + dcTEMP_at_cross) // 2
                    dcTEMP_at_cross = dcTEMP
                last_errorTEMP = errorTEMP

                # Standard servo (loop gain response)
                deltatemp = tmptemp - curTEMP
                if (deltatemp < hystTEMP / 10.0) and (curTEMP < minTEMP):
                    dcTEMP += dcSTEP
                    dcTEMP = min(dcMAX, dcTEMP)
                if ((curTEMP > operTEMP) and (deltatemp > hystTEMP / 20.0)) or (
                    curTEMP > maxTEMP
                ):
                    dcTEMP -= dcSTEP
                    dcTEMP = max(0, dcTEMP)
                pi.set_PWM_dutycycle(pinPWM, dcTEMP)

            curTEMP = tmptemp

            d.update({"C": curTEMP, "D": dcTEMP, "sn": sn})

            if myfile is not None:
                d["file"] = myfile.name

            if hSCD3 is not None:
                # read CO2 data ready status
                pi.i2c_write_device(hSCD3, [0x02, 0x02])
                dataSCD3 = pi.i2c_read_device(hSCD3, nbytesRDY)
                bdataSCD3 = dataSCD3[1]
                if dataSCD3[0] != nbytesRDY:
                    return
                rdy = (bdataSCD3[0] << 8) | bdataSCD3[1]
                # print("rdy", rdy) # if rdy, then read
                if rdy:
                    pi.i2c_write_device(hSCD3, [0x03, 0x00])
                    dataSCD3 = pi.i2c_read_device(hSCD3, nbytesCO2)
                    bdataSCD3 = dataSCD3[1]
                    if dataSCD3[0] != nbytesCO2:
                        return
                    (co2,) = struct.unpack(
                        ">f",
                        bytes([bdataSCD3[0], bdataSCD3[1], bdataSCD3[3], bdataSCD3[4]]),
                    )
                    (tp,) = struct.unpack(
                        ">f",
                        bytes(
                            [bdataSCD3[6], bdataSCD3[7], bdataSCD3[9], bdataSCD3[10]]
                        ),
                    )
                    (hd,) = struct.unpack(
                        ">f",
                        bytes(
                            [bdataSCD3[12], bdataSCD3[13], bdataSCD3[14], bdataSCD3[15]]
                        ),
                    )
                    print(
                        "Time={} CO2 concentration={:.2f} Temperature={:.2f} Humidity={:.2f}".format(
                            time.time() * 1000, co2, tp, hd
                        )
                    )
                    d.update({"CO2": co2, "Tp": tp, "H": hd})

        ds = json.dumps(d)
        pub_socket.send_string(ds)

        if myfile is not None:
            print(ds, file=myfile)


with ExitStack() as stack, ExitStack() as filestack:

    pi = stack.enter_context(pi_cleanup())
    spiMCP3008 = stack.enter_context(spidev.SpiDev())
    ctx = stack.enter_context(zmq.Context())
    pub_socket = stack.enter_context(ctx.socket(zmq.PUB))

    pub_socket.bind("tcp://*:5556")

    myfile = None  # type: Optional[TextIO]

    if arg.name:
        file_path = directory / arg.name
        myfile = filestack.enter_context(open_next(file_path))
        time_since_file_start = time.monotonic()
        print("Logging:", myfile.name)

    running = threading.Event()

    def close(_number, _frame):
        print("Closing down server...")
        signal.signal(signal.SIGINT, signal.SIG_DFL)
        signal.signal(signal.SIGTERM, signal.SIG_DFL)
        running.set()

    signal.signal(signal.SIGINT, close)
    signal.signal(signal.SIGTERM, close)

    spiMCP3008.open(0, 0)
    spiMCP3008.max_speed_hz = 1350000  # 500000

    while not running.is_set():
        # Get I2C bus handle
        hSDP3 = pi.i2c_open(1, DEVICE_SDP3)
        hSCD3 = pi.i2c_open(1, DEVICE_SCD3) if arg.co2 else None

        if myfile:
            # If the disk is full, we should delete a few files.
            total, used, free = shutil.disk_usage(str(myfile.name))
            if free < 2 * GB:
                delete_this_much = int(2.5 * GB - free)
                delete_oldest(file_path, delete_this_much)

            # Also we should make a new file if it's been more than an hour
            # (Currently only triggered by a read error, like removing the
            # sensor and replacing)
            if time.monotonic() > time_since_file_start + 1 * HOUR:
                filestack.pop_all().close()
                myfile = filestack.enter_context(open_next(file_path))
                time_since_file_start = time.monotonic()

        try:
            read_loop(spiMCP3008, pi, hSDP3, hSCD3, running, myfile)
        except pigpio.error:
            # 1 second of "fake" readings
            for i in range(int(ReadoutHz)):
                d = {"v": 1, "t": int(1000 * time.monotonic())}  # type: Dict[str, Any]
                if i == 0 and myfile is not None:
                    d["file"] = myfile.name

                pub_socket.send_json(d)

                running.wait(1 / ReadoutHz)
                if running.is_set():
                    break
        finally:
            pi.i2c_close(hSDP3)
            if hSCD3 is not None:
                pi.i2c_close(hSCD3)
```
