## Supplementary Git Repository for "Inexpensive multi-patient respiratory monitoring system for helmet ventilation during COVID-19 pandemic": device_read.py

```
#!/usr/bin/env python3

import numpy as np
import zmq

from processor.rolling import Rolling

max_pressure_roll = Rolling(window_size=1000 // 50)
pressure_roll = Rolling(window_size=1000)
flow_roll = Rolling(window_size=1000)
time_roll = Rolling(window_size=1000, dtype=np.int64)


with zmq.Context() as ctx, ctx.socket(zmq.SUB) as sub_socket:
    print("Collecting data")

    sub_socket.connect("tcp://localhost:5556")
    sub_socket.subscribe(b"")

    for _ in range(100):

        j = sub_socket.recv_json()
        print(f"Received: {j}")

        if "max_pressure" in j:
            max_pressure_roll.inject(j["max_pressure"])

        pressure_roll.inject_value(j["P"])
        flow_roll.inject_value(j["F"])
        time_roll.inject_value(j["t"])

    print(max_pressure_roll)
    print(pressure_roll)
    print(time_roll)
    print(flow_roll)
```
