## Supplementary Git Repository for "Inexpensive multi-patient respiratory monitoring system for helmet ventilation during COVID-19 pandemic": device_sim.py

```
#!/usr/bin/env python3

# This simulates a sensor.

from datetime import datetime
import time
import zmq

from sim.start_sims import start_sims

start_time = int(1000 * datetime.now().timestamp())  # milliseconds
sims = start_sims(1, start_time, 12000000)  # milliseconds
sim = sims[0]

start_loop = datetime.now().timestamp()

write_alarms_every = 50

# Examples (not final) - will also have temp
alarms = {"max_pressure": 50.0, "max_flow": 100.0}
nC = 0

with zmq.Context() as ctx, ctx.socket(zmq.PUB) as pub_socket:
    pub_socket.bind("tcp://*:5556")

    while True:
        t = int(1000 * datetime.now().timestamp())  # milliseconds
        d = sim.get_next()
        d["t"] = t

        if (nC % write_alarms_every) == 0:
            d = {**d, **alarms}

        pub_socket.send_json(d)

        time.sleep(1 / 50)
        nC += 1
```
