## Supplementary Git Repository for "Inexpensive multi-patient respiratory monitoring system for helmet ventilation during COVID-19 pandemic": analysis-products.md

# Analysis products

The analysis stage of the workflow takes two time-series as input:

   * **flow (L/min):** the primary observable of the flow-meter, after conversion/calibration from delta pressure in Pa through a 4/7 power. Positive **flow** is into the lung; negative **flow** is out.
   * **pressure (cm-H₂O):** air pressure in the lungs relative to atmospheric pressure (i.e. this quantity is zero at atmospheric pressure). Positive **pressure** is above atmospheric pressure; negative **pressure** is below it.

Both are sampled in identical time-steps, and the time-steps themselves are given as a third array:

   * **realtime (sec):** floating point number of seconds since epoch (January 1, 1970). This includes millisecond resolution.

## Time-series products

Currently only one:

   * **volume (mL):** the time-integral of **flow**, after converting L to mL and sec to min. The zero-point for **volume** starts at the beginning of the **flow** integration, so positive **volume** is an inflated lung and negative **volume** is below the initial value.

**Note:** Since **volume** is determined from its differential, it is only known up to an additive constant. Negative volume is not physical, so we adjust the additive constant such that the lowest **volume** ever reached is identified with zero. At the beginning of a time-series, quantities based on absolute **volume** may be revised as new global minima are encountered, but this becomes increasingly rare as a time-series accumulates. The global minimum is the lowest value ever reached, which might predate the data in the rolling buffer: all **volume** values in memory might be greater than zero.

## Breath record products

The time-series data are summarized into records that each represent one breath. Breaths can be long or short, and represent a discretization of the original waveform. Four time points are identified on each wave:

   * **inhale:** when the **flow** is at a maximum.
   * **full:** when the **volume** is at a maximum.
   * **exhale:** when the **flow** is at a minimum (extremely negative).
   * **empty:** when the **volume** is at a minimum.

A breath record can have data on any phase of the breath that can be determined from the time-series, but the dividing line between two breaths is after **empty**. Although the beginning and end aren't guaranteed, each breath record is guaranteed to proceed in the same order: **inhale** → **full** → **exhale** → **empty** in time-order, without any skipped or duplicated steps. These four turning points are determined from a smoothed version of the data, to minimize spurious "breaths" due to noise; the Gaussian sigma for this smoothing procedure is 0.2 seconds.

A breath record may have any of the following fields. (Field names include spaces, but not parentheses or units.)

   * **inhale timestamp (sec):** the **realtime** (floating point seconds since epoch) when the **flow** is at a maximum.
   * **inhale flow (L/min):** the original, unsmoothed **flow** at the time of **inhale**.
   * **inhale dV/dt (mL/sec):** the smoothed **flow** at the time of **inhale**, converted into mL and seconds.
   * **inhale dP/dt (cm-H₂O/sec):** the rate of change of smoothed **pressure** at the time of **inhale**.
   * **inhale compliance (ml/cm-H₂O):** the ratio of **dV/dt** over **dP/dt** at the time of **inhale**.
   * **min pressure (cm-H₂O):** the minimum value of unsmoothed **pressure** between the **inhale** time and the previous **exhale**, if it exists.
   * **full timestamp (sec):** the **realtime** when the **volume** is at a maximum.
   * **full pressure (cm-H₂O):** the original, unsmoothed **pressure** at the time of **full**, which is often but not necessarily equal to **max pressure**.
   * **full volume (mL):** the original, unsmoothed **volume** at the time of **full**, which is always the maximum for this breath.
   * **expiratory tidal volume (mL):** the difference between the **full volume** and the previous **empty volume**, if it exists. This quantity is always positive and is a relative volume, computed within a single time-series buffer, so it is not sensitive to changes in the absolute **volume**.
   * **inhale time (sec):** the **realtime** difference between **empty** and **full**, through the inhale.
   * **exhale timestamp (sec):** the **realtime** when the **flow** is at a minimum (extremely negative).
   * **exhale flow (L/min):** the original, unsmoothed **flow** at the time of **exhale**.
   * **exhale dV/dt (mL/sec):** the smoothed **flow** at the time of **exhale**, converted into mL and seconds.
   * **exhale dP/dT (cm-H₂O/sec):** the rate of change of smoothed **pressure** at the time of **exhale**.
   * **exhale compliance (ml/cm-H₂O):** the ratio of **dV/dt** over **dP/dt** at the time of **exhale**.
   * **max pressure (cm-H₂O):** the maximum value of unsmoothed **pressure** between the **exhale** time and the previous **inhale**, if it exists.
   * **empty timestamp (sec):** the **realtime** when the **volume** is at a minimum.
   * **empty pressure (cm-H₂O):** the original, unsmoothed **pressure** at the time of **empty**, which is often but not necessarily equal to **min pressure**.
   * **empty volume (mL):** the original, unsmoothed **volume** at the time of **empty**, which is always the minimum for this breath.
   * **exhale time (sec):** the **realtime** difference between **full** and **empty**, through the exhale.
   * **average flow (L/min):** the average of **flow** from the last **empty** to the **empty** of this breath.
   * **average pressure (cm-H₂O):** the average of **pressure** from the last **empty** to the **empty** of this breath.
   * **time since last (sec):** the time difference between this breath (at **empty**) and the previous breath (at its **empty**).

## Recomputing breath records

Since the time-series has finite duration, it necessarily clips some part of the first breath and some part of the last breath in its range. As data are accumulated, old time-series data are dropped off one end of the buffer and new time-series data are added to the other. By repeating the analysis on overlapping segments of time, we can recover clipped breaths. Since breath records require much less memory than the original time-series, a longer history of breaths may be saved.

To recover all clipped breaths, the analysis must be performed at least once per `length of time-series` minus `length of one breath` and at most twice per `length of time-series`. Since breath lengths are variable and the analysis is computationally inexpensive, twice per `length of time-series` would be best. This also implies that the length of the time-series must be considerably longer than two breaths, which it is (currently 15 seconds).

**FIXME:** The analysis is currently being run once every second, which is considerably more often than twice per time-series. It could be a lot less frequent than that. Perhaps the **volume** (derived time-series) is desired at 1 second intervals, but the breath records do not need to be constructed this often.

When breath records are recomputed, they have to be matched with old breath records. Any of the four timestamps (**inhale**, **full**, **exhale**, **empty**) that match within 3 × the Gaussian smoothing sigma (0.6 seconds) qualifies as a match. When old and new breath records are identified, any fields that are present in old or new but not the other are kept, and new fields are kept if both old and new are present.

**FIXME:** The breath data accumulates indefinitely. At some point it should be batched to disk or dropped entirely.

## Cumulative measurements

The next data tier is that of cumulative measurements. Each of the following fields has a single value that gets updated with each run of the analysis, unlike the breath records, which accumulate as a growing list. Most are exponentially weighted moving averages (EWMA) of breath records with `alpha=0.3`, meaning that it takes about 3 consecutive breaths for a new trend to emerge.

   * **last breath timestamp (sec):** the **realtime** of the latest breath recorded.
   * **breath interval (sec):** the EWMA of **time since last** from breath records.
   * **RR (1/min):** the instantaneous reciprocal of **breath interval**, converted from seconds to minutes.
   * **PIP (cm-H₂O):** the EWMA of **max pressure** from breath records.
   * **PEEP (cm-H₂O):** the EWMA of **empty pressure** from breath records.
   * **TVe (mL):** the EWMA of **expiratory tidal volume** from breath records.
   * **TVi (mL):** the EWMA of **inspiratory tidal volume** from breath records.
   * **TV (mL):** the instantaneous average of **TVe** and **TVi**.
   * **inhale compliance (ml/cm-H₂O):** the EWMA of **inhale compliance** from breath records.
   * **exhale compliance (ml/cm-H₂O):** the EWMA of **exhale compliance** from breath records.
   * **breath volume rate (L/min):** the instantaneous product of **TV** and **RR**.
   * **inhale time (sec):** the EWMA of **inhale time**.
   * **exhale time (sec):** the EWMA of **exhale time**.
   * **I:E time ratio (unitless):** the instantaneous ratio of **inhale time** over **exhale time**.
   * **breath average flow (L/min):** the EWMA of **averge flow** from breath records.
   * **breath average pressure (cm-H₂O):** the EWMA of **averge pressure** from breath records.
   * **window average flow (L/min):** a dict of window widths (in seconds) to sliding window averages of the raw **flow** time-series (not calculated from breaths).
   * **window average pressure (cm-H₂O):** a dict of window widths (in seconds) to sliding window averages of the raw **pressure** time-series (not calculated from breaths).

## Cumulative timestamps

As a data tier that is parallel to the cumulative measurements, "cumulative timestamps" is a Python dict of the last time each field was updated. This time is not from the measurement stream (i.e. **realtime** of the time-series), it is the output of Python's `time.time()` function. If the time-series is generated on a computer with a different clock offset from the analysis procedure, these times can diverge.

The time associated with the empty string, `""`, is always updated every time `analyze` is called.

The other strings are all the cumulative measurements fields that have ever been set and the time when they were last set. A difference between one of these values and the value associated with `""` indicates a stale value.

## Alarms

Alarms are like cumulative measurements in that they are a fixed number of fields that change in place, rather than a growing sequence like the breath records. Alarms are raised when a cumulative value exceeds a predefined threshold, and are immediately removed once the quantity returns to a suitable value.

An alarm is either undefined (no values out of bounds) or a record with the following fields:

   * **first timestamp (sec):** the **realtime** when the quantity first went out of bounds.
   * **last timestamp (sec):** the **realtime** when the quantity was last observed out of bounds. (In other words, "now", because alarms disappear as soon as the quantity is in bounds.)
   * **extreme:** the most extreme value observed. For alarms of upper bounds, the **extreme** is the maximum value, and for alarms of lower bounds, the **extreme** is the minimum value.

Thresholds for all alarms are derived from the rotary dial.

The following alarms are defined:

   * **PIP Max:** upper bound on the cumulative **PIP** value.
   * **PIP Min:** lower bound on the cumulative **PIP** value.
   * **PEEP Max:** upper bound on the cumulative **PEEP** value.
   * **PEEP Min:** lower bound on the cumulative **PEEP** value.
   * **TVe Max:** upper bound on the cumulative **TVe** value.
   * **TVe Min:** lower bound on the cumulative **TVe** value.
   * **TVi Max:** upper bound on the cumulative **TVi** value.
   * **TVi Min:** lower bound on the cumulative **TVi** value.

The **Stale Data** alarm is configured with the rotary dial, but is otherwise special. It does not have the standard record structure: it is a dict from cumulative measurement field name to the time since its last measurement. See cumulative timestamps (above).

## Status of analysis products in the workflow

Currently, all of the above-mentioned analysis products are computed in the full workflow (every time the `Generator.analyze` method is called) and the results are attached to the `Generator` object. No further action is performed, such as passing them downstream to the nurse GUI.

**FIXME:** These analysis products must be passed downstream to the nurse GUI!

## Future analysis products

If the **volume** and **flow** are presented on a 2-dimensional plot, they form a phase diagram that is independent of breath starting time, unlike the time-series. This could be useful for characterizing the breath as a shape. These shapes could start and end on the **inhale**, **full**, **exhale**, **empty** turning points to avoid overlaps that clutter the display. However, only a few breaths can be displayed at a time, and the data can't be saved indefinitely because they require the full time-series.
