## Supplementary Git Repository for "Inexpensive multi-patient respiratory monitoring system for helmet ventilation during COVID-19 pandemic": environment.yml

```
name: flowmeter
channels:
  - conda-forge
dependencies:
  - black
  - matplotlib
  - mypy
  - numpy
  - pyqt
  - pyqtgraph
  - python==3.7
  - pyyaml
  - pyzmq
  - scipy
  - confuse
  - zeroconf
  - markdown
  - flake8
  - flake8-bugbear
```
