## Supplementary Git Repository for "Inexpensive multi-patient respiratory monitoring system for helmet ventilation during COVID-19 pandemic": common.py

```
from __future__ import annotations

import numpy as np

from nurse.qt import QtGui


# Replace with proper importlib.resources if made a package
from processor.config import get_internal_file

style_path = get_internal_file("nurse/style.css")

guicolors = {
    "ALERT": QtGui.QColor(0, 0, 100),
    "DISCON": QtGui.QColor(0, 0, 200),
}

INFO_STRINGS = {
    "Avg Flow": ".0f",
    "Avg Pressure": ".0f",
    "RR": ".0f",  # (breaths/min)
    "TVe": ".0f",  # (mL)
    "PIP": ".0f",  # (cm H2O)
    "PEEP": ".0f",  # (cm H2O)
    "I:E time ratio": ".1f",
}

HOVER_STRINGS = {
    "Avg Flow": "Flow in L/m averaged over the last 10 seconds",
    "Avg Pressure": "Pressure in cm H20 averaged over the last 10 seconds",
    "RR": "Respiratory rate, in breaths per minute",
    "TVe": "Expiratory tidal volume, in ML",
    "TVi": "Expiratory tidal volume, in ML",
    "PIP": "Peak inspiratory pressure, in cm H2O",
    "PEEP": "Positive end-expiratory pressure, in cm H2O",
    "I:E time ratio": "The inspiratory over expiratory time ratio",
}


class GraphInfo:
    def __init__(self):
        # the y limits ought to be configurable.
        self.graph_labels = ["flow", "pressure", "volume"]
        self.all_graph_labels = self.graph_labels + ["co2"]
        self.graph_names = {key: key.capitalize() for key in self.graph_labels}
        self.graph_names["co2"] = "CO2"

        self.graph_pens = {
            "flow": (120, 255, 50),
            "pressure": (255, 120, 50),
            "volume": (255, 128, 255),
            "co2": (255, 50, 50),
        }

        self.yMax = {"flow": 300, "pressure": 100, "volume": 5000, "co2": 5000}
        self.yMin = {"flow": -300, "pressure": -100, "volume": 0, "co2": 0}
        self.yStep = {"flow": 10, "pressure": 2, "volume": 100, "co2": 10}

        self.yLimKeywords = {
            "flow": {"minYRange": 5, "yMin": -200, "yMax": 200},
            "pressure": {"minYRange": 2, "yMin": -15, "yMax": 40},
            "volume": {"minYRange": 0.5},
            "co2": {"minYRange": 100, "yMin": 0, "yMax": 40_000},
        }

        self.graph_pen_qcol = {k: QtGui.QColor(*v) for k, v in self.graph_pens.items()}

        self.yLims = {
            "flow": (-20, 140),
            "pressure": (-1, 20),
            "volume": (0, 800),
            "co2": (-5, 6000),
        }
        self.units = {"flow": "L/m", "pressure": "cm H2O", "volume": "mL", "co2": "ppm"}


def rolling_mean(data: np.ndarray, size: int) -> np.ndarray:
    assert size % 2 == 1, "Only odd sizes supported"
    edge = (size - 1) // 2
    cumsum = np.empty(len(data) + 1, dtype=float)
    cumsum[0] = 0
    np.cumsum(data, dtype=float, out=cumsum[1:])
    counts = np.arange(len(data))
    right = np.clip(counts + edge + 1, 0, len(data))
    left = np.clip(counts - edge, 0, len(data))
    return (cumsum[right] - cumsum[left]) / (right - left)
```
