## Supplementary Git Repository for "Inexpensive multi-patient respiratory monitoring system for helmet ventilation during COVID-19 pandemic": config.yaml

```
rotary:

  PIP Max:
    type: Incr
    name: PIP Max
    default: 30
    min: 0
    max: 50
    incr: 1
    unit: cm-H2O

  PIP Min:
    type: Incr
    default: 5
    min: 1
    max: 25
    incr: 1
    unit: cm-H2O
    name: PIP Min

  PEEP Max:
    type: Incr
    default: 8
    min: 1
    max: 25
    incr: 1
    unit: cm-H2O
    name: PEEP Max

  PEEP Min:
    type: Incr
    default: 0
    min: 0
    max: 25
    incr: 1
    unit: cm-H2O
    name: PEEP Min

  TVe Max:
    type: Incr
    default: 700
    min: 50
    max: 2000
```
