## Supplementary Git Repository for "Inexpensive multi-patient respiratory monitoring system for helmet ventilation during COVID-19 pandemic": connection_dialog.py

```
from __future__ import annotations

from urllib.parse import urlparse
from processor.listener import FindBroadcasts, Detector
from typing import List, TYPE_CHECKING

if TYPE_CHECKING:
    from nurse.main_window import MainStack

from nurse.qt import (
    QtWidgets,
    QtGui,
    Qt,
    Slot,
    PopdownTitle,
)


class ManualTab(QtWidgets.QWidget):
    def __init__(self, parent: TabbedConnection, address: str):
        super().__init__(parent)

        layout = QtWidgets.QFormLayout(self)

        self.ip_address = QtWidgets.QLineEdit()
        layout.addRow("IP Address:", self.ip_address)

        self.port = QtWidgets.QLineEdit()
        validator = QtGui.QIntValidator()
        self.port.setValidator(validator)
        layout.addRow("Port:", self.port)

        parsed = urlparse(address)
        self.ip_address.setText(parsed.hostname)
        self.port.setText(str(parsed.port))

    def current_url(self) -> str:
        port = int(self.port.text())
        ip_address = self.ip_address.text()
        return f"tcp://{ip_address}:{port}"


class DetectedTab(QtWidgets.QWidget):
    def __init__(self, parent: TabbedConnection, listener: FindBroadcasts):
        super().__init__(parent)

        layout = QtWidgets.QFormLayout(self)

        self.detected = QtWidgets.QComboBox()
        layout.addRow("Detected:", self.detected)

        self.detected.setMinimumWidth(290)

        self.items = list(listener.detected)
        items = [str(d) for d in self.items]
        self.detected.addItems(items)

    def current_url(self) -> str:
        return self.items[self.detected.currentIndex()].url


class LocalTab(QtWidgets.QWidget):
    def __init__(self, parent: TabbedConnection):
        super().__init__(parent)

        self.items: List[int] = []

        layout = QtWidgets.QFormLayout(self)

        self.generators = QtWidgets.QComboBox()
        layout.addRow("Local generators:", self.generators)

        self.generators.setMinimumWidth(290)

        stack: MainStack = self.parent().parent().parent()
        for i in range(24):
            mac = f"dc:a6:32:00:00:{i+1:02x}"
            mac_seen = any(
                graph.gen.record.mac == mac for graph in stack.graphs.values()
            )
            if not mac_seen:
                self.generators.addItem(mac)
                self.items.append(i)

    def current_selection(self) -> int:
        return self.items[self.generators.currentIndex()]


class TabbedConnection(QtWidgets.QTabWidget):
    def __init__(
        self,
        parent: ConnectionDialog,
        address: str,
        listener: FindBroadcasts,
        sim: bool,
    ):
        super().__init__(parent)

        self.detected_tab = DetectedTab(self, listener)
        self.addTab(self.detected_tab, "Detected")

        self.manual_tab = ManualTab(self, address)
        self.addTab(self.manual_tab, "Manual IP")

        if sim:
            self.local_tab = LocalTab(self)
            self.addTab(self.local_tab, "Simulation")


class ConnectionDialog(QtWidgets.QDialog):
    def __init__(
        self,
        parent: QtWidgets.QWidget,
        listener: FindBroadcasts,
        i: int,
        address: str,
        sim: bool,
    ):
        super().__init__(parent)

        self.setWindowTitle(f"Patient box {i} connection")

        self.i = i
        self.listener = listener
        self.items: List[Detector] = []

        self.setWindowModality(Qt.ApplicationModal)

        layout = QtWidgets.QVBoxLayout(self)

        layout.addWidget(PopdownTitle("Add a device"))

        self.tabbed = TabbedConnection(self, address, listener, sim)
        layout.addWidget(self.tabbed)

        buttons = QtWidgets.QDialogButtonBox(
            QtWidgets.QDialogButtonBox.Ok | QtWidgets.QDialogButtonBox.Cancel
        )
        buttons.accepted.connect(self.accept)
        buttons.rejected.connect(self.reject)

        self.ok, cancel = buttons.buttons()

        layout.addWidget(buttons)

        if not self.tabbed.detected_tab.items:
            self.tabbed.setCurrentIndex(1)
            self.tabbed.setTabEnabled(0, False)

    @Slot()
    def accept(self):
        if self.tabbed.currentIndex() < 2:
            self.parent().add_new_by_address(self.connection_address())
        else:
            self.parent().add_new_generator(self.tabbed.local_tab.current_selection())

        super().accept()

    def connection_address(self) -> str:
        if self.tabbed.currentIndex() == 0:
            return self.tabbed.detected_tab.current_url()
        else:
            return self.tabbed.manual_tab.current_url()
```
