## Supplementary Git Repository for "Inexpensive multi-patient respiratory monitoring system for helmet ventilation during COVID-19 pandemic": dragdrop.py

```
from __future__ import annotations

from nurse.qt import (
    QtWidgets,
    QtGui,
    QtCore,
    Qt,
    swap_grid,
)

from nurse.gen_record_gui import GenRecordGUI

from typing import Optional


class DraggableSensor(QtWidgets.QFrame):
    MIME_NAME = "DraggableSensor"

    def mime_type(self: DraggableSensor) -> str:
        return f"application/{self.MIME_NAME}"

    def __init__(self: DraggableSensor, *args, **kwargs):
        super().__init__(*args, **kwargs)
        self.setAcceptDrops(True)
        self._start_pos: Optional[QtGui.QMouseEvent] = None

    def mousePressEvent(self: DraggableSensor, event: QtGui.QMouseEvent) -> None:
        if event.button() == Qt.LeftButton:
            self._start_pos = event.pos()

    def mouseMoveEvent(self: DraggableSensor, evt: QtGui.QMouseEvent) -> None:
        if self._start_pos is None or not (evt.buttons() & Qt.LeftButton):
            return
        if (
            evt.pos() - self._start_pos
        ).manhattanLength() < QtWidgets.QApplication.startDragDistance():
            return

        hot_spot = evt.pos()

        mime_data = QtCore.QMimeData()
        mime_data.setData(
            self.mime_type(),
            QtCore.QByteArray.number(hot_spot.x())
            + b" "
            + QtCore.QByteArray.number(hot_spot.y()),
        )

        dpr = QtWidgets.QApplication.instance().devicePixelRatio()
        pixmap = QtGui.QPixmap(self.size() * dpr)
        pixmap.setDevicePixelRatio(dpr)
        self.render(pixmap)

        drag = QtGui.QDrag(self)
        drag.setMimeData(mime_data)
        drag.setPixmap(pixmap)
        drag.setHotSpot(hot_spot)

        self.setVisible(False)
        drag.exec()

        if drag.target() and drag.source() != drag.target():
            parent = self.parent()
            swap_grid(parent.grid_layout, drag.source(), drag.target())
            for item in (drag.source(), drag.target()):
                if hasattr(item, "gen"):
                    record: GenRecordGUI = item.gen.record
                    ind = parent.grid_layout.indexOf(item)
                    x, y, _width, _height = parent.grid_layout.getItemPosition(ind)
                    record.position = (x, y)
            parent.drop_final_row_or_column_if_needed()

        self.setVisible(True)

    def dragEnterEvent(self: DraggableSensor, evt: QtGui.QDragEnterEvent) -> None:
        if evt.mimeData().hasFormat(self.mime_type()):
            evt.accept()
            eff = QtWidgets.QGraphicsOpacityEffect(self)
            eff.setOpacity(0.5)
            self.setGraphicsEffect(eff)
        else:
            evt.ignore()

    def dragLeaveEvent(self: DraggableSensor, evt: QtGui.QDragEnterEvent) -> None:
        self.setGraphicsEffect(None)

    def dropEvent(self: DraggableSensor, evt):
        if evt.mimeData().hasFormat(self.mime_type()):
            evt.accept()
            self.setGraphicsEffect(None)
        else:
            evt.ignore()
```
