## Supplementary Git Repository for "Inexpensive multi-patient respiratory monitoring system for helmet ventilation during COVID-19 pandemic": drilldown.py

```
from __future__ import annotations

import pyqtgraph as pg

from typing import Optional, Any, Dict, TYPE_CHECKING
import logging
import subprocess
import os
import sys

import numpy as np

from nurse.qt import (
    QtCore,
    QtWidgets,
    Qt,
    QtGui,
    Slot,
    HBoxLayout,
    VBoxLayout,
    GridLayout,
    BoxName,
    DraggableMixin,
)

from nurse.common import GraphInfo, HOVER_STRINGS, rolling_mean
from nurse.header import DrilldownHeaderWidget
from nurse.gen_record_gui import GenRecordGUI, GeneratorGUI
from processor.generator import Status
from nurse.generator_dialog import GeneratorDialog
from processor.config import config
from processor.remote_generator import RemoteGenerator

if TYPE_CHECKING:
    from nurse.main_window import MainStack

logger = logging.getLogger("povm")


class BoxHeader(QtWidgets.QLabel):
    pass


class DrilldownCumulative(QtWidgets.QLabel):
    pass


class DrilldownLimit(QtWidgets.QLabel):
    pass


class DrilldownLabel(QtWidgets.QLabel):
    pass


class DrilldownSince(QtWidgets.QLabel):
    pass


class DisplayBoxBase(QtWidgets.QFrame):
    @property
    def status(self) -> Status:
        return Status[self.property("alert_status")]

    @status.setter
    def status(self, value: Status):
        if value.name != self.property("alert_status"):
            self.setProperty("alert_status", value.name)

            self.style().unpolish(self)
            self.style().polish(self)

            self.cumulative.style().unpolish(self.cumulative)
            self.cumulative.style().polish(self.cumulative)


class DisplayBox(DisplayBoxBase):
    def __init__(self, *, key: str, label: str, fmt: str = ""):
        super().__init__()
        self.key = key
        self.fmt = fmt
        self.window = config["global"]["avg-window"].get(int)
        layout = QtWidgets.QVBoxLayout(self)

        upper_layout = QtWidgets.QHBoxLayout()
        layout.addLayout(upper_layout)

        title = DrilldownLabel(label)
        title.setToolTip(HOVER_STRINGS[key])
        upper_layout.addWidget(title)
        upper_layout.addStretch()

        self.cumulative = DrilldownCumulative()
        self.cumulative.setToolTip(HOVER_STRINGS[key])
        upper_layout.addWidget(self.cumulative)

        self.since = DrilldownSince()
        layout.addWidget(self.since, 0, Qt.AlignHCenter)

        lower_layout = QtWidgets.QHBoxLayout()
        layout.addLayout(lower_layout)

        self.lower_limit = DrilldownLimit()
        self.lower_limit.setToolTip("Lower limit for alarm")
        lower_layout.addWidget(self.lower_limit)

        lower_layout.addStretch()

        if self.key.startswith("Avg "):
            self.avg_time = DrilldownLimit()
            self.avg_time.setToolTip("Time averaged over in seconds")
            lower_layout.addWidget(self.avg_time)
            lower_layout.addStretch()

        self.upper_limit = DrilldownLimit()
        self.upper_limit.setToolTip("Upper limit for alarm")
        lower_layout.addWidget(self.upper_limit)

        self.update_cumulative()
        self.update_limits()

    def update_cumulative(self):
        gen: Optional[GeneratorGUI] = (
            self.parent().parent().gen
            if (self.parent() and self.parent().parent())
            else None
        )

        if gen is None:
            value = None
        elif self.key == "Avg Flow":
            value = gen.average_flow[self.window]
        elif self.key == "Avg Pressure":
            value = gen.average_pressure[self.window]
        else:
            value = gen.cumulative.get(self.key)

        if value is not None:
            assert gen is not None, "Should not be possible if value is not None"

            self.cumulative.setText(format(value, self.fmt))
            if f"{self.key} Max" in gen.alarms:
                self.status = (
                    Status.ALERT_SILENT
                    if isinstance(gen, RemoteGenerator)
                    and gen.time_left is not None
                    and gen.time_left > 0
                    else Status.ALERT
                )
                item = gen.alarms[f"{self.key} Max"]
                if "first timestamp" in item:
                    over = (gen.realtime[-1] - item["first timestamp"]) + gen.tardy
                    self.since.setText(f"Over for {over:.0f} s")
            elif f"{self.key} Min" in gen.alarms:
                self.status = (
                    Status.ALERT_SILENT
                    if isinstance(gen, RemoteGenerator)
                    and gen.time_left is not None
                    and gen.time_left > 0
                    else Status.ALERT
                )
                item = gen.alarms[f"{self.key} Min"]
                if "first timestamp" in item:
                    under = (gen.realtime[-1] - item["first timestamp"]) + gen.tardy
                    self.since.setText(f"Under for {under:.0f} s")
            else:
                self.status = (
                    Status.SILENT
                    if isinstance(gen, RemoteGenerator)
                    and gen.time_left is not None
                    and gen.time_left > 0
                    else Status.OK
                )
                self.since.setText("")
        else:
            self.cumulative.setText("---")
            self.status = Status.DISCON

    def update_limits(self):
        min_key = f"{self.key} Min"
        max_key = f"{self.key} Max"

        gen = (
            self.parent().parent().gen
            if (self.parent() and self.parent().parent())
            else None
        )

        if gen is not None:
            if min_key in gen.rotary:
                self.lower_limit.setText(format(gen.rotary[min_key].value, self.fmt))

            if max_key in gen.rotary:
                self.upper_limit.setText(format(gen.rotary[max_key].value, self.fmt))

            if hasattr(self, "avg_time"):
                avg_window = config["global"]["avg-window"].get(int)
                self.avg_time.setText(f"({avg_window})")


class DisplayBoxCO2(DisplayBoxBase):
    def __init__(self, label: str):
        super().__init__()
        layout = QtWidgets.QVBoxLayout(self)

        title = DrilldownLabel(label)
        layout.addWidget(title)

        self.cumulative = DrilldownCumulative(label)
        layout.addWidget(self.cumulative)


class AllDisplays(QtWidgets.QWidget):
    def __init__(self):
        super().__init__()

        self.boxes = [
            DisplayBox(key="Avg Flow", label="Flow", fmt=".0f"),
            DisplayBox(key="Avg Pressure", label="P", fmt=".0f"),
            DisplayBox(key="RR", label="RR", fmt=".0f"),
            DisplayBox(key="I:E time ratio", label="I:E", fmt=".2f"),
            DisplayBox(key="PIP", label="PIP", fmt=".0f"),
            DisplayBox(key="PEEP", label="PEEP", fmt=".0f"),
            DisplayBox(key="TVe", label="TVe", fmt=".0f"),
            DisplayBox(key="TVi", label="TVi", fmt=".0f"),
        ]

        layout = GridLayout(self)
        for n, box in enumerate(self.boxes):
            layout.addWidget(box, *divmod(n, 2))

    def update_cumulative(self):
        for box in self.boxes:
            box.update_cumulative()

    def update_limits(self):
        for box in self.boxes:
            box.update_limits()


class CO2Displays(QtWidgets.QWidget):
    def __init__(self) -> None:
        super().__init__()
        layout = QtWidgets.QHBoxLayout(self)

        self.co2 = DisplayBoxCO2("CO2:")
        layout.addWidget(self.co2)

        self.co2_temp = DisplayBoxCO2("Temp:")
        layout.addWidget(self.co2_temp)

        self.humidity = DisplayBoxCO2("Humidity:")
        layout.addWidget(self.humidity)

        self.setVisible(False)

    def update_co2(self) -> None:
        gen: Optional[GeneratorGUI] = self.parent().gen if self.parent() else None

        if gen is not None:
            if len(gen.co2):
                co2 = np.mean(gen.co2[-5:])
                co2_temp = np.mean(gen._co2_temp[-5:])
                humidity = np.mean(gen._humidity[-5:])

                self.co2.cumulative.setText(f"{co2:.0f}")
                self.co2_temp.cumulative.setText(f"{co2_temp:.1f}")
                self.humidity.cumulative.setText(f"{humidity:.2f}")

                if not self.isVisible():
                    self.setVisible(True)

                alarming = "Avg CO2 Max" in gen.alarms or "Avg CO2 Min" in gen.alarms
                self.co2.status = Status.ALERT if alarming else Status.OK


class DisplayText(QtWidgets.QTextEdit):
    def __init__(self):
        super().__init__()

        self.setReadOnly(True)
        self._text = ""

    def update_if_needed(self, text, html=False):
        if self._text != text:
            self._text = text
            val = self.verticalScrollBar().value()
            if html:
                self.setHtml(text)
            else:
                self.setPlainText(text)
            self.verticalScrollBar().setValue(val)


class PatientTitle(QtWidgets.QWidget):
    def __init__(self):
        super().__init__()
        layout = HBoxLayout(self)

        self.name_lbl = QtWidgets.QPushButton("i")
        self.name_lbl.setToolTip("Click for details")
        self.name_lbl.clicked.connect(self.click_number)
        layout.addWidget(self.name_lbl)

        self.name_edit = QtWidgets.QLineEdit()
        self.name_edit.setPlaceholderText("Please enter title")
        layout.addWidget(self.name_edit)

        self.name_edit.editingFinished.connect(self.update_title)

    @property
    def record(self) -> GenRecordGUI:
        return self.parent().gen.record

    def repolish(self) -> None:
        self.name_lbl.style().unpolish(self.name_lbl)
        self.name_edit.style().polish(self.name_edit)

    def activate(self) -> None:
        self.name_edit.setText(self.record.title)
        self.record.master_signal.title_changed.connect(self.external_update_title)

    def deactivate(self) -> None:
        if self.parent().gen is not None:
            self.record.master_signal.title_changed.disconnect(
                self.external_update_title
            )

    @Slot()
    def external_update_title(self) -> None:
        self.name_edit.setText(self.record.title)

    @Slot()
    def update_title(self) -> None:
        self.record.title = self.name_edit.text()

    @Slot()
    def click_number(self) -> None:
        dialog = GeneratorDialog(self, self.parent().gen)
        dialog.open()


class DrilldownWidget(QtWidgets.QWidget):
    def __init__(self, parent: QtWidgets.QWidget):
        super().__init__(parent)

        layout = VBoxLayout(self)

        self.header = DrilldownHeaderWidget()
        self.return_btn = self.header.return_btn
        layout.addWidget(self.header)

        columns_layout = HBoxLayout()
        layout.addLayout(columns_layout)

        self.patient = PatientDrilldownWidget()
        columns_layout.addWidget(self.patient)

        side_layout = VBoxLayout()
        columns_layout.addLayout(side_layout)

        self.alarms_layout = VBoxLayout()
        side_layout.addLayout(self.alarms_layout)
        side_layout.addStretch()

        self.alarm_boxes: Dict[int, AlarmBox] = {}

    def add_alarm_box(self, gen: GeneratorGUI, i: int):
        alarm_box = AlarmBox(gen, i)
        self.alarm_boxes[i] = alarm_box
        self.alarms_layout.addWidget(alarm_box)
        alarm_box.clicked.connect(self.click_alarm)

    def drop_alarm_box(self, ind: int):
        alarm_box = self.alarm_boxes.pop(ind)
        alarm_box.clicked.disconnect(self.click_alarm)
        self.alarms_layout.removeWidget(alarm_box)
        alarm_box.setParent(None)

    def sort_alarms(self):
        stacked: MainStack = self.parent().parent().main_stack
        positions = {
            tuple(reversed(graph.gen.record.position)): n
            for n, graph in stacked.graphs.items()
        }
        ordered = [positions[k] for k in sorted(positions)]
        for i in range(len(ordered)):
            if self.alarms_layout.itemAt(i).widget().i != ordered[i]:
                for j in range(i, len(ordered)):
                    if self.alarms_layout.itemAt(j).widget().i == ordered[i]:
                        item = self.alarms_layout.takeAt(j)
                        self.alarms_layout.insertWidget(i, item.widget())
                        break

    @Slot()
    def click_alarm(self):
        alarm_box: AlarmBox = self.sender()
        self.parent().parent().drilldown_activate(alarm_box.i)

    def activate(self, i: int):
        """
        Call this to activate or switch drilldown screens!
        """

        if self.patient.gen is not None:
            self.deactivate()

        self.sort_alarms()

        for n, box in self.alarm_boxes.items():
            box.active = i == n
            box.update_gen()

        main_stack = self.parent().parent().main_stack

        record: GenRecordGUI
        if self.patient.gen is not None:
            record = self.patient.gen.record
            record.master_signal.mac_changed.disconnect(
                self.patient.external_update_boxname
            )
            record.master_signal.sid_changed.disconnect(
                self.patient.external_update_sid
            )

        self.patient.gen = main_stack.graphs[i].gen
        assert (
            self.patient.gen is not None
        ), "Should never be possible to activate with None"

        self.patient.title.activate()

        record = self.patient.gen.record
        record.master_signal.mac_changed.connect(self.patient.external_update_boxname)
        record.master_signal.sid_changed.connect(self.patient.external_update_sid)
        record.master_signal.notes_changed.connect(self.patient.external_update_notes)

        self.patient.external_update_boxname()
        self.patient.external_update_sid()
        self.patient.external_update_notes()

        self.patient.update_plot(True)

    def deactivate(self) -> None:
        assert (
            self.patient.gen is not None
        ), "Should never be possible to activate with None"

        record: GenRecordGUI
        record = self.patient.gen.record

        record.master_signal.mac_changed.disconnect(
            self.patient.external_update_boxname
        )
        record.master_signal.sid_changed.disconnect(self.patient.external_update_sid)

        record.master_signal.notes_changed.disconnect(
            self.patient.external_update_notes
        )

        self.patient.title.deactivate()
        self.patient.gen = None
        self.patient.qTimer.stop()


class AlarmBox(QtWidgets.QPushButton):
    def __init__(self, gen: GeneratorGUI, i: int):
        super().__init__()
        self.gen: GeneratorGUI = gen
        record: GenRecordGUI = self.gen.record
        record.master_signal.title_changed.connect(self.update_gen)
        self.active = False
        self.i = i

        self.stack = QtWidgets.QStackedLayout()
        self.setLayout(self.stack)

        boxname_container = QtWidgets.QWidget()
        boxname_layout = VBoxLayout(boxname_container)
        self.stack.addWidget(boxname_container)
        self.upper = BoxName("Box name")
        self.lower = BoxName("Unknown")
        boxname_layout.addWidget(self.upper, 0, Qt.AlignHCenter)
        boxname_layout.addWidget(self.lower, 0, Qt.AlignHCenter)

        title_container = QtWidgets.QWidget()
        title_layout = VBoxLayout(title_container)
        self.title = QtWidgets.QLabel("")
        title_layout.addWidget(self.title, 0, Qt.AlignHCenter)
        self.stack.addWidget(title_container)

        self.update_gen()

    @Slot()
    def update_gen(self):
        if self.gen.record.title:
            self.title.setText(self.gen.record.title)
            self.title.setToolTip(
                f"{self.gen.record.title}\n{self.gen.record.box_name}"
            )
            self.stack.setCurrentIndex(1)
        else:
            upper, lower = self.gen.record.stacked_name.split("\n")
            self.upper.setText(upper)
            self.lower.setText(lower)
            self.stack.setCurrentIndex(0)

    @property
    def status(self) -> Status:
        return Status[self.property("alert_status")]

    @status.setter
    def status(self, value: Status):
        value_name = value.name + ("_ACTIVE" if self.active else "")
        if value_name != self.property("alert_status"):
            self.setProperty("alert_status", value_name)
            self.style().unpolish(self)
            self.style().polish(self)


class LogTextEdit(QtWidgets.QPlainTextEdit):
    def __init__(self) -> None:
        super().__init__()

        self.text_changed = False
        self.textChanged.connect(self.update_notes)

    @Slot()
    def update_notes(self) -> None:
        self.text_changed = True

    @property
    def record(self) -> GenRecordGUI:
        return self.parent().gen.record

    def focusOutEvent(self, e: QtGui.QFocusEvent) -> None:
        if self.text_changed:
            self.record.notes = self.toPlainText()
        self.text_changed = False


class DraggableMsg(QtWidgets.QMessageBox, DraggableMixin):
    def __init__(self, parent: QtWidgets.QWidget):
        super().__init__(parent)
        self.move(parent.geometry().center() - self.geometry().center())
        self.setTextFormat(Qt.RichText)
        self.setWindowFlags(Qt.Popup)


class PatientDrilldownWidget(QtWidgets.QFrame):
    def __init__(self) -> None:
        super().__init__()
        self.curves: Dict[str, pg.PlotCurveItem] = {}
        self.curves2: Dict[str, pg.PlotCurveItem] = {}
        self.upper: Dict[str, pg.PlotDataItem] = {}
        self.lower: Dict[str, pg.PlotDataItem] = {}
        self.current: Dict[str, pg.PlotDataItem] = {}
        self.co2_plot: Optional[pg.PlotItem] = None
        self.graphs: Dict[str, pg.PlotItem] = {}
        self.gen: Optional[GeneratorGUI] = None

        layout = HBoxLayout(self)

        left_layout = VBoxLayout()
        layout.addLayout(left_layout, 2)

        right_layout = VBoxLayout()
        layout.addLayout(right_layout, 1)

        self.title = PatientTitle()
        left_layout.addWidget(self.title)

        warning_layout = QtWidgets.QHBoxLayout()
        left_layout.addLayout(warning_layout)

        warning_layout.addStretch()

        warning_layout.addWidget(QtWidgets.QLabel("Box name: "))
        self.box_name = BoxName("???")
        warning_layout.addWidget(self.box_name)

        warning_layout.addStretch()

        self.sensor_id = QtWidgets.QLabel("Sensor ID: ???")
        warning_layout.addWidget(self.sensor_id)

        warning_layout.addStretch()

        self.graphview = pg.GraphicsView(parent=self)
        self.graph_layout = pg.GraphicsLayout()
        self.graphview.setCentralWidget(self.graph_layout)
        left_layout.addWidget(self.graphview, 4)

        phaseview = pg.GraphicsView(parent=self)
        self.phase_layout = pg.GraphicsLayout()
        phaseview.setCentralWidget(self.phase_layout)
        right_layout.addWidget(phaseview, 2)

        self.set_plots()

        side_by_side_layout = HBoxLayout()
        right_layout.addLayout(side_by_side_layout, 3)

        displays_layout = VBoxLayout()
        side_by_side_layout.addLayout(displays_layout)

        nurse_layout = VBoxLayout()
        side_by_side_layout.addLayout(nurse_layout)

        self.displays = AllDisplays()
        displays_layout.addWidget(self.displays)

        self.last_ts = QtWidgets.QLabel("Last updated: ---")
        self.last_ts.setObjectName("LastTS")
        displays_layout.addWidget(self.last_ts)

        self.last_interation = QtWidgets.QLabel("Last interaction at bedside: ---")
        self.last_interation.setObjectName("LastInteraction")
        displays_layout.addWidget(self.last_interation)

        self.time_left = QtWidgets.QLabel("")
        self.time_left.setObjectName("TimeLeft")
        displays_layout.addWidget(self.time_left)
        self.time_left.setVisible(False)

        button_box = QtWidgets.QWidget()
        button_box.setObjectName("DrilldownExtras")
        displays_layout.addWidget(button_box)

        stacked_button_box = QtWidgets.QVBoxLayout(button_box)

        buttons_layout = QtWidgets.QHBoxLayout()
        stacked_button_box.addLayout(buttons_layout)

        all_alarms = QtWidgets.QPushButton("Alarms")
        all_alarms.setToolTip("Frozen details of the current active alarms")
        all_alarms.clicked.connect(self.display_alarms)
        buttons_layout.addWidget(all_alarms, 1)

        all_cumulative = QtWidgets.QPushButton("Quantities")
        all_cumulative.setToolTip("Frozen cummulative quanities")
        all_cumulative.clicked.connect(self.display_cumulative)
        buttons_layout.addWidget(all_cumulative, 1)

        all_rotary = QtWidgets.QPushButton("Settings")
        all_rotary.setToolTip(
            "Frozen details of the current settings on the patient box"
        )
        all_rotary.clicked.connect(self.display_rotary)
        buttons_layout.addWidget(all_rotary, 1)

        buttons_2_layout = QtWidgets.QHBoxLayout()
        stacked_button_box.addLayout(buttons_2_layout)

        all_logs = QtWidgets.QPushButton("Patient data files")
        all_logs.setToolTip("Open the current logging directory")
        all_logs.clicked.connect(self.display_logs)
        buttons_2_layout.addWidget(all_logs, 1)

        # If a past history data viewer was added, it could be added here too

        lim_help = QtWidgets.QLabel("All alarm limits are set on the device")
        displays_layout.addWidget(lim_help)

        displays_layout.addStretch()

        self.co2_widget = CO2Displays()
        nurse_layout.addWidget(self.co2_widget)

        nurse_layout.addWidget(BoxHeader("Notes:"))
        self.log_edit = LogTextEdit()
        nurse_layout.addWidget(self.log_edit, 1)

        self.qTimer = QtCore.QTimer()
        self.qTimer.setSingleShot(True)
        self.qTimer.timeout.connect(self.update_plot)

    @property
    def status(self) -> Status:
        return Status[self.property("alert_status") or "NONE"]

    @status.setter
    def status(self, value: Status) -> None:
        if value.name != self.property("alert_status"):
            self.setProperty("alert_status", value.name)
            self.title.name_lbl.setText(value.value)
            self.title.repolish()

    @Slot()
    def display_cumulative(self) -> None:
        if self.gen is not None:
            cumulative = "\n".join(
                rf"<p>{k}: {v:.2f}</p>" for k, v in self.gen.cumulative.items()
            )
        else:
            cumulative = "Disconnected"
        if not cumulative:
            cumulative = "No computed values yet"

        box = DraggableMsg(self)
        box.setWindowTitle("Static computed values")
        box.setText(cumulative)
        box.show()

    @Slot()
    def display_alarms(self) -> None:
        assert self.gen is not None

        def expand(s: Dict[str, Any]) -> str:
            return "".join(f"<br>  {k}: {v}" for k, v in s.items())

        active_alarms = "\n".join(
            rf"<p>{k}: {expand(v)}</p>" for k, v in self.gen.alarms.items()
        )

        if not active_alarms:
            active_alarms = "No alarms currently."

        box = DraggableMsg(self)
        box.setWindowTitle("Active alarms (static)")
        box.setText(active_alarms)
        box.show()

    @Slot()
    def display_rotary(self) -> None:
        assert self.gen is not None
        rotary_text = "\n".join(
            rf"<p>{v.name}: {v.value} {v.unit}</p>" for v in self.gen.rotary.values()
        )

        box = DraggableMsg(self)
        box.setWindowTitle("Static alarm limits on the patient box")
        box.setText(rotary_text)
        box.show()

    @Slot()
    def display_logs(self) -> None:
        assert self.gen is not None
        if self.gen.saver_cml is not None:
            filepath = self.gen.saver_cml.filepath.parent

            startfile = getattr(os, "startfile", None)
            if startfile is not None:
                startfile(filepath, "open")
            elif sys.platform.startswith("darwin"):
                subprocess.check_call(["open", filepath])
            else:
                subprocess.check_call(["xdg-open", filepath])

        else:
            box = DraggableMsg(self)
            box.setWindowTitle("Debug mode")
            box.setText(
                "This was opened with the --debug flag. Logs are not being kept."
            )
            box.show()

    @Slot()
    def external_update_boxname(self) -> None:
        assert self.gen is not None
        text = self.gen.record.box_name
        if self.box_name.text() != text:
            self.box_name.setText(text)

    @Slot()
    def external_update_sid(self) -> None:
        assert self.gen is not None
        text = f"Sensor ID: {self.gen.record.sid:016X}"
        if self.sensor_id.text() != text:
            self.sensor_id.setText(text)

    @Slot()
    def external_update_notes(self) -> None:
        assert self.gen is not None
        text = self.gen.record.notes
        self.log_edit.setPlainText(text)

    def set_plot(self, key: str) -> pg.PlotItem:
        gis = GraphInfo()

        limit_pen = pg.mkPen(color=(170, 30, 30), width=3, style=QtCore.Qt.DashLine)
        current_pen = pg.mkPen(color=(50, 50, 180), width=3, style=QtCore.Qt.DotLine)

        if self.graph_layout.items:
            self.graph_layout.nextRow()

        graph = self.graph_layout.addPlot(
            x=None,
            y=None,
            name="CO2" if key == "co2" else key.capitalize(),
            autoDownsample=True,
            clipToView=True,
        )
        graph.invertX()
        graph.setRange(xRange=(28, 0))  # Actually shows a bit more that the range
        graph.setLabel(
            "left", gis.graph_names[key], "L" if key == "volume" else gis.units[key]
        )

        # Force autoscaling to avoid micro sized y ranges
        # Also keep x from going out of bounds
        graph.getViewBox().setLimits(
            xMin=0, xMax=30, minXRange=2, **gis.yLimKeywords[key]
        )

        # Axis line at 0
        graph.addItem(pg.PlotDataItem([0, 30], [0, 0]))

        self.upper[key] = pg.PlotDataItem(x=None, y=None, pen=limit_pen)
        graph.addItem(self.upper[key])
        self.lower[key] = pg.PlotDataItem(x=None, y=None, pen=limit_pen)
        graph.addItem(self.lower[key])
        self.current[key] = pg.PlotDataItem(x=None, y=None, pen=current_pen)
        graph.addItem(self.current[key])

        pen = pg.mkPen(color=gis.graph_pens[key], width=2)
        self.curves[key] = graph.plot(x=None, y=None, pen=pen)
        self.curves2[key] = graph.plot(x=None, y=None, pen=pen)

        return graph

    def set_plots(self) -> None:
        gis = GraphInfo()

        graphs = {key: self.set_plot(key) for key in gis.graph_labels}
        self.graphs = graphs

        graphs["volume"].setLabel("bottom", "Time", "s")

        graphs[gis.graph_labels[0]].setXLink(graphs[gis.graph_labels[1]])
        graphs[gis.graph_labels[1]].setXLink(graphs[gis.graph_labels[2]])

        # Phase plot
        self.phase_graph = self.phase_layout.addPlot(x=None, y=None, name="Phase")
        self.phase_graph.setRange(
            xRange=gis.yLims["pressure"],
            yRange=tuple(v / 1000 for v in gis.yLims["volume"]),
        )
        self.phases = list(
            reversed(
                [
                    self.phase_graph.plot(
                        x=None,
                        y=None,
                        pen=pg.mkPen(color=(i * 50, i * 50, i * 10), width=3),
                    )
                    for i in range(1, 6)
                ]
            )
        )
        self.phase_graph.setLabel("left", "Volume", units="L")
        self.phase_graph.setLabel("bottom", "Pressure", units="cm H2O")

    @Slot()
    def update_plot(self, first: bool = False) -> None:
        if self.isVisible() and self.gen is not None:
            gis = GraphInfo()
            scroll = self.parent().header.mode_scroll

            with self.gen.lock:
                if first or not self.parent().header.freeze_btn.checkState():
                    avg_window = config["global"]["avg-window"].get(int)

                    select = (
                        slice(np.searchsorted(-self.gen.time, -32), None)
                        if len(self.gen.time)
                        else slice(None)
                    )
                    co2_select = (
                        slice(
                            np.searchsorted(-self.gen.co2_time, -35),
                            None,
                        )
                        if len(self.gen.co2_time)
                        else slice(None)
                    )

                    # If we have CO2 sensor data
                    if len(self.gen.co2):
                        # Add the plot if not added already
                        if self.co2_plot is None:
                            self.co2_plot = self.set_plot("co2")
                            self.graphs["volume"].setLabel("bottom", "", "")
                            self.co2_plot.setLabel("bottom", "Time", "s")
                            self.graphs[gis.graph_labels[2]].setXLink(self.co2_plot)

                    labels = gis.all_graph_labels if self.co2_plot else gis.graph_labels
                    for key in labels:
                        if key == "co2":
                            xvalues = self.gen.co2_time[co2_select]
                            orig_yvalues = self.gen.co2[co2_select]
                            yvalues = rolling_mean(orig_yvalues, 5)
                            key_cap = "CO2"
                        else:
                            xvalues = self.gen.time[select]
                            yvalues = getattr(self.gen, key)[select]
                            key_cap = key.capitalize()

                        if key == "volume":
                            yvalues = yvalues / 1000

                        if scroll:
                            self.curves[key].setData(xvalues, yvalues)
                            x, _y = self.curves2[key].getData()
                            if x is not None and len(x) > 0:
                                self.curves2[key].setData(
                                    x=np.array([], dtype=float),
                                    y=np.array([], dtype=float),
                                )

                        else:
                            realtime = (
                                self.gen.co2_realtime[co2_select]
                                if key == "co2"
                                else self.gen.realtime[select]
                            )

                            # This should never be empty, but quit here if it is
                            if not len(realtime):
                                continue

                            last = realtime[-1]
                            breakpt = np.searchsorted(realtime, last - last % 30)
                            gap = 25

                            self.curves[key].setData(
                                30 - (realtime[breakpt:] % 30),
                                yvalues[breakpt:],
                            )
                            self.curves2[key].setData(
                                30 - (realtime[gap:breakpt] % 30),
                                yvalues[gap:breakpt],
                            )

                        min_key = f"Avg {key_cap} Min"
                        max_key = f"Avg {key_cap} Max"
                        if min_key in self.gen.rotary:
                            self.lower[key].setData(
                                [0, 30], [self.gen.rotary[min_key].value] * 2
                            )
                        if max_key in self.gen.rotary:
                            self.upper[key].setData(
                                [0, 30], [self.gen.rotary[max_key].value] * 2
                            )

                        if key == "flow":
                            self.current[key].setData(
                                [0, avg_window], [self.gen.average_flow[avg_window]] * 2
                            )
                        elif key == "pressure":
                            self.current[key].setData(
                                [0, avg_window],
                                [self.gen.average_pressure[avg_window]] * 2,
                            )
                        elif key == "co2":
                            avg_co2 = float(np.mean(orig_yvalues[xvalues < avg_window]))
                            self.current[key].setData(
                                [0, avg_window],
                                [avg_co2] * 2,
                            )

                    for i, phase in enumerate(self.phases):
                        range = slice(-(i + 1) * 50 * 3 - 1, -i * 50 * 3)
                        phase.setData(
                            self.gen.pressure[range], self.gen.volume[range] / 1000
                        )

                    if self.status != self.gen.status:
                        self.status = self.gen.status
                    self.displays.update_cumulative()
                    self.displays.update_limits()
                    self.co2_widget.update_co2()

                    time_str = (
                        "now" if self.gen.tardy < 1 else f"{self.gen.tardy:.0f}s ago"
                    )
                    date_str = (
                        "---"
                        if self.gen.last_update is None
                        else format(self.gen.last_update, "%m-%d-%Y %H:%M:%S")
                    )

                    for breath in reversed(self.gen.breaths):
                        if "full timestamp" in breath:
                            time_since = (
                                self.gen.realtime[-1] - breath["full timestamp"]
                            ) + self.gen.tardy
                            breath_str = f"Most recent breath: {time_since:.0f} s ago"
                            break
                    else:
                        breath_str = "No detected breaths yet"

                    self.last_ts.setText(
                        f"Updated: {time_str} @ {date_str}\n{breath_str}"
                    )

                    if (
                        isinstance(self.gen, RemoteGenerator)
                        and self.gen.last_interact is not None
                        and self.gen.current_monotonic is not None
                    ):
                        last_interaction = (
                            self.gen.tardy
                            + self.gen.current_monotonic
                            - self.gen.last_interact
                        )
                        self.last_interation.setText(
                            f"Last interaction at bedside: {last_interaction:.0f} s ago"
                        )

                    if (
                        isinstance(self.gen, RemoteGenerator)
                        and self.gen.time_left is not None
                        and self.gen.time_left > 0
                    ):
                        self.time_left.setText(
                            f"Silenced, time remaining: {self.gen.time_left:.0f} s"
                        )
                        if not self.time_left.isVisible():
                            self.time_left.setVisible(True)

                    elif self.time_left.isVisible():
                        self.time_left.setVisible(False)

            patient = self.parent()
            main_stack = patient.parent().parent().main_stack

            for ind in main_stack.graphs.keys():
                patient.alarm_boxes[ind].status = main_stack.graphs[ind].gen.status

        self.qTimer.start(50)
```
