## Supplementary Git Repository for "Inexpensive multi-patient respiratory monitoring system for helmet ventilation during COVID-19 pandemic": gen_record_gui.py

```
from __future__ import annotations

from dataclasses import dataclass, field
from typing import Tuple, Optional, Union
from pathlib import Path
from contextlib import suppress

import yaml

from processor.gen_record import GenRecord
from nurse.qt import Signal, QtCore
from processor.config import get_data_dir
from processor.remote_generator import RemoteGenerator
from processor.local_generator import LocalGenerator


class RemoteGeneratorGUI(RemoteGenerator):
    record: GenRecordGUI


class LocalGeneratorGUI(LocalGenerator):
    record: GenRecordGUI


GeneratorGUI = Union[RemoteGeneratorGUI, LocalGeneratorGUI]


class RecordSignals(QtCore.QObject):
    title_changed = Signal()
    mac_changed = Signal()
    sid_changed = Signal()
    notes_changed = Signal()


class NoMacError(Exception):
    pass


@dataclass
class GenRecordGUI(GenRecord):
    master_signal: RecordSignals = field(default_factory=RecordSignals)

    _path: Optional[Path] = None
    ip_address: Optional[str] = None
    _notes: str = ""
    _active: bool = True
    _position: Tuple[int, int] = (-1, -1)

    @property
    def position(self) -> Tuple[int, int]:
        return self._position

    @position.setter
    def position(self, value: Tuple[int, int]):
        self._position = value
        self.save()

    @property
    def notes(self) -> str:
        return self._notes

    @notes.setter
    def notes(self, value: str) -> None:
        self._notes = value
        self.master_signal.notes_changed.emit()
        self.save()

    @property
    def path(self) -> Path:
        if self._path is not None:
            return self._path
        if self._mac is not None:
            dirpath = get_data_dir() / "nurse_layout"
            dirpath.mkdir(exist_ok=True)
            self._path = dirpath / f"{self.safemac}.yml"
            return self._path
        else:
            raise NoMacError

    def title_changed(self) -> None:
        self.master_signal.title_changed.emit()
        self.save()

    def mac_changed(self) -> None:
        self.master_signal.mac_changed.emit()

        with suppress(NoMacError, FileNotFoundError), self.path.open() as f:
            info = yaml.safe_load(f)
            self.title = info["title"]
            self.notes = info["notes"]
            self._active = True  # Loading a device activates it!

        self.save()

    @property
    def active(self) -> bool:
        return self._active

    @active.setter
    def active(self, value: bool):
        self._active = value
        self.save()

    def save(self) -> None:
        with suppress(NoMacError), self.path.open("w") as f:
            d = {"title": self.title, "sid": self.sid, "notes": self.notes}
            if self.ip_address:
                d["ip_address"] = self.ip_address
            if self._position[0] >= 0 and self._position[1] >= 0:
                d["position"] = self._position

            d["active"] = self._active

            yaml.safe_dump(d, f)

    def sid_changed(self) -> None:
        self.master_signal.sid_changed.emit()
        self.save()
```
