## Supplementary Git Repository for "Inexpensive multi-patient respiratory monitoring system for helmet ventilation during COVID-19 pandemic": generator_dialog.py

```
from __future__ import annotations

from nurse.qt import QtWidgets, Slot
from processor.generator import Status
from nurse.gen_record_gui import (
    LocalGeneratorGUI,
    GeneratorGUI,
)

from typing import TYPE_CHECKING

if TYPE_CHECKING:
    from nurse.main_window import MainStack
    from nurse.grid import PatientSensor


class BasicTab(QtWidgets.QWidget):
    def __init__(self, gen: GeneratorGUI):
        super().__init__()

        layout = QtWidgets.QVBoxLayout(self)

        layout.addWidget(
            QtWidgets.QLabel(
                "Please make sure a title is set to make box identification easier.\n"
                "The first few characters will be shown in the drilldown alarm screen."
            )
        )

        form_layout = QtWidgets.QFormLayout()
        layout.addLayout(form_layout)

        self.title = QtWidgets.QLineEdit()
        self.title.setPlaceholderText("Please enter title")
        self.title.setText(gen.record.title)

        form_layout.addRow("Title:", self.title)
        form_layout.addRow("Box Name:", QtWidgets.QLabel(gen.record.box_name))


class DetailsTab(QtWidgets.QWidget):
    def __init__(self, gen: GeneratorGUI):
        super().__init__()

        layout = QtWidgets.QFormLayout(self)

        layout.addRow("MAC Addr:", QtWidgets.QLabel(gen.record.mac))
        layout.addRow("Sensor:", QtWidgets.QLabel(format(gen.record.sid, "016X")))
        layout.addRow(
            "IP Address:", QtWidgets.QLabel(getattr(gen, "address", "Simulation"))
        )


class GeneratorDialog(QtWidgets.QDialog):
    def __init__(
        self, parent: QtWidgets.QWidget, gen: GeneratorGUI, *, grid: bool = False
    ):
        super().__init__(parent)
        self.setWindowTitle(f"Info for {gen.record.box_name}")

        self.gen = gen
        layout = QtWidgets.QVBoxLayout(self)

        self.basic = BasicTab(gen)
        self.details = DetailsTab(gen)
        self.selection = QtWidgets.QTabWidget()
        self.selection.addTab(self.basic, "Basic")
        self.selection.addTab(self.details, "Details")
        layout.addWidget(self.selection)

        self.buttons = QtWidgets.QDialogButtonBox()
        self.buttons.addButton(QtWidgets.QDialogButtonBox.Ok)
        self.buttons.addButton(QtWidgets.QDialogButtonBox.Cancel)

        if grid:
            self.discon = self.buttons.addButton(
                "Remove", QtWidgets.QDialogButtonBox.DestructiveRole
            )
            self.discon.setEnabled(
                self.gen.status == Status.DISCON
                or isinstance(self.gen, LocalGeneratorGUI)
            )
            self.discon.setToolTip("You can only disconnect an unplugged sensor")
            self.discon.clicked.connect(self.disconnect_sensor)
            self.discon.setObjectName("Remove")

        layout.addWidget(self.buttons)

        self.buttons.accepted.connect(self.accept)
        self.buttons.rejected.connect(self.reject)

    @Slot()
    def disconnect_sensor(self) -> None:
        print("Diconnecting")
        parent: PatientSensor = self.parent()
        main: MainStack = parent.parent()
        main.drop_item(parent.i)
        self.reject()

    @Slot()
    def accept(self) -> None:
        self.gen.record.title = self.basic.title.text()
        super().accept()
```
