## Supplementary Git Repository for "Inexpensive multi-patient respiratory monitoring system for helmet ventilation during COVID-19 pandemic": grid.py

```
from __future__ import annotations

import pyqtgraph as pg
import numpy as np

from datetime import datetime

from typing import Dict, Any, Optional

from nurse.qt import (
    QtWidgets,
    QtGui,
    Qt,
    Slot,
    HBoxLayout,
    VBoxLayout,
    FormLayout,
    BoxName,
)

from nurse.common import GraphInfo, INFO_STRINGS, HOVER_STRINGS
from nurse.dragdrop import DraggableSensor
from nurse.generator_dialog import GeneratorDialog
from nurse.gen_record_gui import GeneratorGUI
from processor.generator import Status
from processor.config import config


class NumberLabel(QtWidgets.QLabel):
    pass


class NumbersWidget(QtWidgets.QWidget):
    def __init__(self):
        super().__init__()

        layout = FormLayout(self)

        self.val_widgets = {}

        for info in INFO_STRINGS:
            val_widget = NumberLabel("---")
            val_widget.setToolTip(HOVER_STRINGS[info])
            val_widget.setMinimumWidth(56)
            self.val_widgets[info] = val_widget
            title = QtWidgets.QLabel(
                info.split()[-1][0] if info.startswith("Avg") else info.split()[0]
            )
            title.setToolTip(HOVER_STRINGS[info])
            layout.addRow(
                title,
                self.val_widgets[info],
            )

            self.set_value(info, None)

    def set_value(self, info_str: str, value: float = None, ok: bool = True) -> None:
        val_widget = self.val_widgets[info_str]
        info_widget = self.layout().labelForField(val_widget)

        fmt = INFO_STRINGS[info_str]

        val_widget.setText("---" if value is None else f"{value:{fmt}}")

        prev = val_widget.property("measure")
        curr = "NONE" if value is None else ("OK" if ok else "ERR")

        if prev is None or prev != curr:
            val_widget.setProperty("measure", curr)
            info_widget.setProperty("measure", curr)
            val_widget.style().unpolish(val_widget)
            val_widget.style().polish(val_widget)
            info_widget.style().unpolish(info_widget)
            info_widget.style().polish(info_widget)

    def __iter__(self):
        return iter(self.val_widgets)


class PatientTitleWidget(QtWidgets.QWidget):
    def __init__(self, gen: GeneratorGUI):
        super().__init__()
        self.setToolTip("Click for details")
        self.gen = gen
        record = self.gen.record

        layout = HBoxLayout(self)

        self.name_btn = QtWidgets.QPushButton("i")
        layout.addWidget(self.name_btn)

        self.name_edit = QtWidgets.QLineEdit()
        self.name_edit.setText(self.gen.record.title)
        self.name_edit.setPlaceholderText("Please enter title")
        self.name_edit.editingFinished.connect(self.update_title)
        record.master_signal.title_changed.connect(self.external_update_title)
        layout.addWidget(self.name_edit, 1)

        self.box_name = BoxName(gen.record.stacked_name)
        layout.addWidget(self.box_name)
        record.master_signal.mac_changed.connect(self.external_update_mac)

    @Slot()
    def external_update_title(self):
        self.name_edit.setText(self.gen.record.title)

    @Slot()
    def external_update_mac(self):
        self.box_name.setText(self.gen.record.stacked_name)

    @Slot()
    def update_title(self):
        self.gen.record.title = self.name_edit.text()

    def repolish(self):
        self.name_btn.style().unpolish(self.name_btn)
        self.name_btn.style().polish(self.name_btn)

        self.name_edit.style().unpolish(self.name_edit)
        self.name_edit.style().polish(self.name_edit)


class EmptySensor(DraggableSensor):
    def __init__(self):
        super().__init__()

        layout = VBoxLayout(self)
        layout.addWidget(QtWidgets.QLabel("Empty"), 0, Qt.AlignHCenter)


class PatientSensor(DraggableSensor):
    @property
    def status(self):
        return Status[self.property("alert_status")]

    @status.setter
    def status(self, value: Status):
        if value.name != self.property("alert_status"):
            self.setProperty("alert_status", value.name)
            self.title_widget.name_btn.setText(value.value)
            self.title_widget.repolish()

            self.style().unpolish(self)
            self.style().polish(self)

    def __init__(self, *, i: int, gen: GeneratorGUI):
        super().__init__()
        self.last_status_change = int(1000 * datetime.now().timestamp())
        self.gen: GeneratorGUI = gen
        self.current_alarms: Dict[str, Any] = {}
        self.i = i

        layout = HBoxLayout(self)

        layout_left = VBoxLayout()
        layout.addLayout(layout_left)

        self.title_widget = PatientTitleWidget(gen)
        layout_left.addWidget(self.title_widget)

        self.graphview = pg.GraphicsView()
        self.graphview.setAttribute(Qt.WA_TransparentForMouseEvents)
        graphlayout = pg.GraphicsLayout()
        graphlayout.setContentsMargins(0, 5, 0, 0)
        self.graphview.setCentralWidget(graphlayout)
        layout_left.addWidget(self.graphview)

        gis = GraphInfo()
        self.graph = {}
        for j, key in enumerate(gis.graph_labels):
            self.graph[key] = graphlayout.addPlot(x=None, y=None, name=key.capitalize())
            self.graph[key].setMouseEnabled(False, False)
            self.graph[key].invertX()
            if j != len(gis.graph_labels):
                graphlayout.nextRow()

        self.status = self.gen.status
        self.title_widget.name_btn.clicked.connect(self.click_number)

        self.values = NumbersWidget()
        layout.addWidget(self.values)

        self.curves: Dict[str, Any] = {}
        self.dialog: Optional[GeneratorDialog] = None

    @Slot()
    def click_number(self) -> None:
        if self.dialog is not None:
            if self.dialog.isVisible():
                return

        self.dialog = GeneratorDialog(self, self.gen, grid=True)
        self.dialog.setWindowFlags(self.dialog.windowFlags() | Qt.WindowStaysOnTopHint)
        self.dialog.show()
        self.dialog.move(self.geometry().center() - self.dialog.rect().center())

    def set_plot(self):
        assert len(self.curves) == 0

        gis = GraphInfo()

        for i, (key, graph) in enumerate(self.graph.items()):
            pen = pg.mkPen(color=gis.graph_pens[key], width=2)

            self.curves[key] = graph.plot(x=None, y=None, pen=pen, autoDownsample=True)

            graph.setRange(xRange=(15, 0), yRange=gis.yLims[key])

            graph.getAxis("left").setStyle(
                tickTextOffset=2,
                textFillLimits=[
                    (2, 1),
                ],  # Never have less than two ticks
            )

            if i != len(gis.graph_labels) - 1:
                graph.hideAxis("bottom")

            graph.addLine(y=0)

        self.graph[gis.graph_labels[0]].setXLink(self.graph[gis.graph_labels[1]])
        self.graph[gis.graph_labels[1]].setXLink(self.graph[gis.graph_labels[2]])

    def update_plot(self):
        gis = GraphInfo()
        avg_window = config["global"]["avg-window"].get(int)

        with self.gen.lock:
            # Fill in the data
            for key in gis.graph_labels:
                if self.isVisible():
                    select = (
                        slice(np.searchsorted(-self.gen.time, -15), None)
                        if len(self.gen.time)
                        else slice(None)
                    )
                    xvalues = self.gen.time[select]
                    yvalues = getattr(self.gen, key)[select]

                    self.curves[key].setData(xvalues, yvalues)
                else:
                    self.curves[key].setData(x=None, y=None)

            # Change of status requires a background color change
            self.status = self.gen.status

            alarming_quanities = {key.rsplit(maxsplit=1)[0] for key in self.gen.alarms}

            for key in self.values:
                value: Optional[float]

                if key == "Avg Flow":
                    value = self.gen.average_flow[avg_window]
                elif key == "Avg Pressure":
                    value = self.gen.average_pressure[avg_window]
                else:
                    value = self.gen.cumulative.get(key)

                self.values.set_value(
                    key,
                    value=value,
                    ok=key not in alarming_quanities,
                )

    def mouseReleaseEvent(self, ev: QtGui.QMouseEvent):
        if (
            self._start_pos is not None
            and ev.button() == Qt.LeftButton
            and (ev.pos() - self._start_pos).manhattanLength()
            < QtWidgets.QApplication.startDragDistance()
        ):
            self._start_pos = None
            self.parent().parent().parent().drilldown_activate(self.i)
        else:
            self._start_pos = None
            ev.ignore()
```
