## Supplementary Git Repository for "Inexpensive multi-patient respiratory monitoring system for helmet ventilation during COVID-19 pandemic": header.py

```
from __future__ import annotations

from nurse.qt import QtWidgets, QtGui, Qt, Slot, HBoxLayout
from nurse.common import GraphInfo
from datetime import datetime

from nurse.limits_dialog import LimitDialog, NoLimitDialog
from nurse.help_dialog import HelpDialog


class WindowTitle(QtWidgets.QLabel):
    pass


class HeaderButton(QtWidgets.QPushButton):
    pass


class PrincetonLogoWidget(QtWidgets.QWidget):
    def __init__(self, *args, **kwargs):
        super().__init__(*args, **kwargs)
        layout = HBoxLayout(self)

        logo = QtGui.QPixmap("images/PUsig2-158C-shield.png").scaledToWidth(18)
        logolabel = QtWidgets.QLabel()
        logolabel.setPixmap(logo)

        text = QtWidgets.QLabel("Princeton Open Ventilation Monitor")
        text.setMinimumWidth(90)
        text.setAlignment(Qt.AlignLeft)
        layout.addWidget(logolabel, 0, Qt.AlignVCenter)
        layout.addWidget(text, 0, Qt.AlignVCenter)
        layout.setSpacing(0)


class NSFLogoWidget(QtWidgets.QWidget):
    def __init__(self, *args, **kwargs):
        super().__init__(*args, **kwargs)
        layout = HBoxLayout(self)

        logo = QtGui.QPixmap("images/nsf-logo-100.png").scaledToWidth(25)
        logolabel = QtWidgets.QLabel()
        logolabel.setPixmap(logo)
        layout.addWidget(logolabel)
        layout.setAlignment(Qt.AlignRight)


class DateTimeWidget(QtWidgets.QWidget):
    def __init__(self, *args, **kwargs):
        super().__init__(*args, **kwargs)
        layout = HBoxLayout(self)

        now = datetime.now()
        nowstring = now.strftime("%d %b %Y %H:%M:%S")
        text = QtWidgets.QLabel(nowstring)
        text.setAlignment(Qt.AlignLeft)
        layout.addWidget(text, 0, Qt.AlignVCenter)
        layout.addStretch()


class LimitButton(QtWidgets.QPushButton):
    def __init__(self, key):
        gis = GraphInfo()
        super().__init__(f"{key.capitalize()}({gis.units[key]})")
        self.setToolTip(f"Adjust the vertical display range for {key.capitalize()}")
        self.key = key

        self.setProperty("graph", key)
        self.clicked.connect(self.click_graph_info)

    @Slot()
    def click_graph_info(self):
        graphs = self.parent().parent().parent().graphs
        if not graphs:
            NoLimitDialog(self).exec()
        else:
            limit = LimitDialog(self.key, parent=self)
            limit.open()


class GraphLabelWidget(QtWidgets.QWidget):
    def __init__(self, *args, **kwargs):
        super().__init__(*args, **kwargs)

        layout = HBoxLayout(self)

        gis = GraphInfo()

        alpha = 140
        values = {}
        for key in gis.graph_pen_qcol:
            pen = gis.graph_pen_qcol[key]
            values[key] = "{r}, {g}, {b}, {a}".format(
                r=pen.red(), g=pen.green(), b=pen.blue(), a=alpha
            )

        text = QtWidgets.QLabel("Graph settings")
        text.setAlignment(Qt.AlignLeft)
        layout.addWidget(text, 0, Qt.AlignVCenter)

        for key in gis.graph_labels:
            name_btn = LimitButton(key)
            layout.addWidget(name_btn, 0, Qt.AlignVCenter)


class HeaderWidget(QtWidgets.QWidget):
    pass


class MainHeaderWidget(HeaderWidget):
    def __init__(self, *args, **kwargs):
        super().__init__(*args, **kwargs)
        layout = HBoxLayout(self)

        layout.addWidget(PrincetonLogoWidget())
        layout.addStretch()
        layout.addWidget(WindowTitle("Overview Screen"))
        layout.addStretch()

        graph_info = GraphLabelWidget()
        layout.addWidget(graph_info)

        self.add_btn = HeaderButton("+")
        self.add_btn.setToolTip("Add a device manually")
        layout.addWidget(self.add_btn)

        self.help_btn = HeaderButton("?")
        self.help_btn.setObjectName("help_btn")
        self.help_btn.setToolTip("Get help with the current screen")
        self.help_btn.clicked.connect(self.call_for_help)
        layout.addWidget(self.help_btn)

        nsf_logo = NSFLogoWidget()
        layout.addWidget(nsf_logo)

        self.fs_exit = HeaderButton("X")
        self.fs_exit.setObjectName("exit_btn")
        self.fs_exit.setToolTip("Exit (may auto-restart on some systems)")
        self.fs_exit.setVisible(False)
        layout.addWidget(self.fs_exit)

        # dt_info = DateTimeWidget()
        # layout.addWidget(dt_info, 6) # Would need to be updated periodically

    @Slot()
    def call_for_help(self):
        help = HelpDialog(self, 0)
        help.setWindowFlags(Qt.Popup)
        help.show()


class DrilldownHeaderWidget(HeaderWidget):
    def __init__(self):
        super().__init__()
        layout = HBoxLayout(self)

        layout.addWidget(PrincetonLogoWidget())
        layout.addStretch()
        layout.addWidget(WindowTitle("Selected Patient View"))
        layout.addStretch()

        self.mode_btn = QtWidgets.QPushButton("Mode: Scroll")
        self.mode_btn.setObjectName("mode_btn")
        self.mode_btn.setToolTip("Toggle visual graph update system between two styles")
        self.mode_btn.clicked.connect(self.mode_btn_callback)
        layout.addWidget(self.mode_btn)

        self.freeze_btn = QtWidgets.QCheckBox("Freeze")
        self.freeze_btn.setToolTip("Stop refreshing display (toggle)")
        layout.addWidget(self.freeze_btn)

        self.return_btn = QtWidgets.QPushButton("Return to overview")
        self.return_btn.setObjectName("return_btn")
        self.return_btn.setToolTip("Go back to main overview screen")
        layout.addWidget(self.return_btn, 0, Qt.AlignVCenter)

        self.help_btn = HeaderButton("?")
        self.help_btn.setObjectName("help_btn")
        self.help_btn.setToolTip("Get help with the current screen")
        self.help_btn.clicked.connect(self.call_for_help)
        layout.addWidget(self.help_btn)

        nsf_logo = NSFLogoWidget()
        layout.addWidget(nsf_logo)

    @property
    def mode_scroll(self):
        return self.mode_btn.text() == "Mode: Scroll"

    @Slot()
    def mode_btn_callback(self):
        if self.mode_scroll:
            self.mode_btn.setText("Mode: Overwrite")
        else:
            self.mode_btn.setText("Mode: Scroll")

    @Slot()
    def call_for_help(self):
        help = HelpDialog(self, 1)
        help.setWindowFlags(Qt.Popup)
        help.show()
```
