## Supplementary Git Repository for "Inexpensive multi-patient respiratory monitoring system for helmet ventilation during COVID-19 pandemic": help_dialog.py

```
from __future__ import annotations

import sys
from nurse.qt import QtWidgets, PopdownTitle, DraggableMixin
from processor.version import get_version
from string import Template
from nurse.common import style_path, GraphInfo


class HelpDialog(QtWidgets.QDialog, DraggableMixin):
    def __init__(self, parent: QtWidgets.QWidget = None, default_tab: int = 0):
        super().__init__(parent)
        self.setWindowTitle("Help")

        layout = QtWidgets.QVBoxLayout(self)

        layout.addWidget(PopdownTitle("Help for the Princeton Open Ventilator Monitor"))

        tabs = QtWidgets.QTabWidget()
        layout.addWidget(tabs)

        main_help = QtWidgets.QLabel(
            r"""
        <p>At the top of the screen, you have the main menu bar. The three color coded graph labels after "Graph Settings" are clickable;
        clicking on them opens a limit selection dialog that can adjust the graph limits for all sensors. The next item on the menu is a "+" symbol,
        and clicking on that will open the add sensor dialog (advanced, usually not needed). Finally, you will see the "?" button that brought you here.</p>

        <p> The main screen has a grid of sensors. You can click the status button to view information. You can click on the title to set it.
        After the title, you can see the box name. Cumulatve values are displayed on the right side of the graph. You can click/tap on a sensor to enter the "drilldown"
        for that sensor. You can drag one sensor on another to swap their positions.</p>

        <p>
        Status codes:
        <\p>

        <ul>
          <li>OK: No alarms present</li>
          <li>!: Alarm parameters exceeded (red)</li>
          <li>S: Silent mode, no alarms present (dark yellow)</li>
          <li>S!: Silent mode, one or more alarms present (bright yellow)</li>
          <li>D: Disconnected (blue)</li>
        </ul>
        """
        )
        main_help.setWordWrap(True)
        tabs.addTab(main_help, "Overview screen")

        drilldown_help = QtWidgets.QLabel(
            r"""
        <p> In the drilldown screen, there are a few new items in the top bar: </p>
        <ul>
          <li>Mode: can be set to Scroll or Overwrite</li>
          <li>Freeze: Will stop the screen from updating when selected</li>
          <li>Return to main window: Leave the drilldown</li>
        </ul>

        <p>
        The main screen on the left is similar to the one shown on the main page, but
        it allows direct manipulation (hover over an axes and turn the wheel, or drag).
        If you click the small "A" button, you can reset to view all.
        </p>

        <p>
        On the right side, you have the Volume vs. Pressure curve, a collection of cumulative
        values, and some timing information, such as the last time the patient box was adjusted.
        There is also a Notes field that can be used to enter information.
        </p>

        <p>
        The final column on the far right side of the screen provides an overview of all
        the boxes attached to the system. You can click on one to quickly switch, and the colors
        match the alarm status for that box.
        </p>
        """
        )
        drilldown_help.setWordWrap(True)
        tabs.addTab(drilldown_help, "Selected patient view")

        drilldown_rotary = QtWidgets.QLabel(
            r"""
        <p>
        The patient box has a screen and a dial on the front, and a red silence button on the top.
        The unique box name is displayed when the device is started (and can also be retrieved from
        the final menu item). The main screen is a live monitor screen, and shows the 10 second averaged
        Flow, Pressure, and Respiratory Rate. Alarms are shown on the far right of the screen, and the display
        turns red and a buzzer sounds when an alarm is active, unless the silence countdown timer is active, in which
        case the screen turns yellow and "S" for silence is replaced by "Q" for quiet. A number is shown here if more
        than one alarm is active.
        <\p>
        <p>
        To control the patient box:
        </p>
        <ul>
        <li>Turn to select items from the menu (item number is shown in the top left)</li>
        <li>Press and turn to set a value</li>
        <li>Press the silence button to put the device into silent mode for 120 seconds.</li>
        <li>Turn the knob while holding the silence button to adjust the timer from 0 to 995 seconds.</li>
        <li>Reset menu item only: Press and turn to activate the reset; release while the meter is full to reset the alarm settings.</li>
        <li>Box name menu item only: Press and turn to see sub-menu items.</li>
        </ul>
        """
        )
        drilldown_rotary.setWordWrap(True)
        tabs.addTab(drilldown_rotary, "Patient box")

        version = get_version() or "<unknown>"
        layout.addWidget(QtWidgets.QLabel(f"Version: {version}"))

        self.button = QtWidgets.QDialogButtonBox(QtWidgets.QDialogButtonBox.Ok)
        layout.addWidget(self.button)

        self.button.accepted.connect(self.close)

        tabs.setCurrentIndex(default_tab)


if __name__ == "__main__":
    app = QtWidgets.QApplication(sys.argv)

    if "Fusion" in QtWidgets.QStyleFactory.keys():
        QtWidgets.QApplication.setStyle(QtWidgets.QStyleFactory.create("Fusion"))
    else:
        print("Fusion style is not available, display may be platform dependent")

    dialog = HelpDialog()

    gis = GraphInfo()

    with open(style_path) as f:
        s = Template(f.read())
        t = s.substitute(gis.graph_pens)
        dialog.setStyleSheet(t)

    dialog.show()
    sys.exit(app.exec_())
```
