## Supplementary Git Repository for "Inexpensive multi-patient respiratory monitoring system for helmet ventilation during COVID-19 pandemic": limits_dialog.py

```
from nurse.qt import QtWidgets, Qt, Slot, PopdownTitle
from nurse.common import GraphInfo


class LimitSpinBox(QtWidgets.QDoubleSpinBox):
    def __init__(self, key, is_upper):
        super().__init__()
        self.key = key
        self.is_upper = is_upper
        gis = GraphInfo()

        self.setMaximum(gis.yMax[self.key])
        self.setMinimum(gis.yMin[self.key])
        self.setSingleStep(gis.yStep[self.key])
        self.setSuffix(" " + gis.units[self.key])


class GraphUnits(PopdownTitle):
    def __init__(self, key, *args, **kwargs):
        super().__init__(*args, **kwargs)
        self.setProperty("graph", key)


class LimitDialog(QtWidgets.QDialog):
    def __init__(self, name, parent):
        super().__init__(parent)
        gis = GraphInfo()
        self.key = name
        self.setWindowTitle(f"Change {name} visual display range")
        self.orig_lower: float = gis.yLims[self.key][0]
        self.orig_upper: float = gis.yLims[self.key][1]

        layout = QtWidgets.QVBoxLayout(self)

        layout.addWidget(
            GraphUnits(
                self.key, f"{gis.graph_names[self.key]} ({gis.units[self.key]})"
            ),
            0,
            Qt.AlignHCenter,
        )

        form_layout = QtWidgets.QFormLayout()
        layout.addLayout(form_layout)

        self.upper = LimitSpinBox(self.key, True)
        form_layout.addRow("Upper display:", self.upper)
        self.upper.valueChanged.connect(self.change_value)

        self.lower = LimitSpinBox(self.key, False)
        form_layout.addRow("Lower display:", self.lower)
        self.lower.valueChanged.connect(self.change_value)

        buttons = QtWidgets.QDialogButtonBox(
            QtWidgets.QDialogButtonBox.Ok | QtWidgets.QDialogButtonBox.Cancel
        )
        buttons.accepted.connect(self.accept)
        buttons.rejected.connect(self.reject)
        layout.addWidget(buttons)

        graphs = parent.parent().parent().parent().graphs
        graph = graphs[0]

        self.orig_lower, self.orig_upper = graph.graph[self.key].viewRange()[1]
        self.lower.setValue(self.orig_lower)
        self.upper.setValue(self.orig_upper)

    @Slot()
    def change_value(self):
        for graph in self.parent().parent().parent().parent().graphs.values():
            graph.graph[self.key].setRange(
                yRange=[self.lower.value(), self.upper.value()]
            )

    @Slot()
    def reject(self):
        self.lower.setValue(self.orig_lower)
        self.upper.setValue(self.orig_upper)
        self.change_value()
        super().reject()


class NoLimitDialog(QtWidgets.QMessageBox):
    def __init__(self, parent: QtWidgets.QWidget):
        super().__init__(
            QtWidgets.QMessageBox.Warning,
            "No sensors connected",
            "No sensors are connected, use the + button on the top right or plug in a sensor.",
            parent=parent,
        )
```
