## Supplementary Git Repository for "Inexpensive multi-patient respiratory monitoring system for helmet ventilation during COVID-19 pandemic": main_window.py

```
from __future__ import annotations

import pyqtgraph as pg

import logging
from string import Template
from typing import Optional, List, Tuple, Dict
import threading
import itertools

from nurse.qt import (
    QtCore,
    QtWidgets,
    Qt,
    Slot,
    Signal,
    VBoxLayout,
    GridLayout,
)

from nurse.common import style_path, GraphInfo
from nurse.header import MainHeaderWidget
from nurse.grid import PatientSensor, EmptySensor
from nurse.drilldown import DrilldownWidget
from nurse.connection_dialog import ConnectionDialog
from nurse.recover import collect_restore, restore_limits

from nurse.gen_record_gui import (
    GenRecordGUI,
    LocalGeneratorGUI,
    RemoteGeneratorGUI,
    GeneratorGUI,
)

from processor.listener import FindBroadcasts
from processor.logging import make_nested_logger


logger = logging.getLogger("povm")


class WaitingWidget(QtWidgets.QFrame):
    def __init__(self):
        super().__init__()
        layout = QtWidgets.QVBoxLayout(self)
        layout.addStretch()
        text = QtWidgets.QLabel("Waiting for a patient sensor to be discovered...")
        text.setAlignment(Qt.AlignCenter)
        layout.addWidget(text)
        layout.addStretch()


class InjectDiscovery(QtCore.QObject):
    inject = Signal()


class MainStack(QtWidgets.QWidget):
    def __init__(
        self,
        parent,
        *,
        listener: FindBroadcasts,
        displays: Optional[int],
        sim: bool,
        addresses: List[str],
        fresh: bool,
    ):
        super().__init__(parent)

        self.listener = listener
        self.sim = sim

        layout = VBoxLayout(self)

        self.header = MainHeaderWidget(self)
        layout.addWidget(self.header)

        grid_layout = GridLayout()
        self.grid_layout: QtWidgets.QGridLayout = grid_layout
        layout.addLayout(grid_layout)

        self.graphs: Dict[int, PatientSensor] = {}
        self.infos: List[WaitingWidget] = []
        self.next_graph: int = 0  # Value of next graph to add

        if displays or addresses:
            disp_addr = itertools.zip_longest(
                range(displays or len(addresses)), addresses or []
            )
            for i, addr in disp_addr:
                local_logger = make_nested_logger(i)
                ip_addr = addr or "tcp://127.0.0.1:8100"
                gen: GeneratorGUI = (
                    RemoteGeneratorGUI(
                        address=ip_addr,
                        logger=local_logger,
                        gen_record=GenRecordGUI(local_logger, ip_address=ip_addr),
                    )
                    if not sim
                    else LocalGeneratorGUI(
                        i=i + 1,
                        logger=local_logger,
                        gen_record=GenRecordGUI(local_logger),
                    )
                )
                gen.run()  # Close must be called
                self.add_item(gen)
        elif fresh:
            self.grid_layout.addWidget(WaitingWidget())
        else:
            restore_dict = collect_restore()
            if not restore_dict:
                self.grid_layout.addWidget(WaitingWidget())
            else:
                a, b = restore_limits(restore_dict)
                self.empty_grid(a, b)
                for mac, restore in restore_dict.items():
                    logger.info(f"Restoring {mac} @ {restore.ip_address}")
                    local_logger = make_nested_logger(self.next_graph)
                    gen = RemoteGeneratorGUI(
                        address=restore.ip_address,
                        logger=local_logger,
                        gen_record=GenRecordGUI(
                            local_logger, ip_address=restore.ip_address
                        ),
                    )
                    gen.run()
                    self.add_item(gen, restore.position)

        self.qTimer = QtCore.QTimer()
        self.qTimer.timeout.connect(self.update_plots)
        self.qTimer.setSingleShot(True)
        self.qTimer.start()

        self.header.add_btn.clicked.connect(self.add_item_dialog)

        self.injector = InjectDiscovery()
        self.queue_lock = threading.Lock()

        if not displays and not addresses:
            self.injector.inject.connect(self.add_from_queue)
            self.listener.inject = lambda: self.injector.inject.emit()
            self.injector.inject.emit()

    @Slot()
    def add_from_queue(self):
        # The header doesn't display properly if this runs at the same time via
        # different triggers
        with self.queue_lock:
            while not self.listener.queue.empty():
                address = self.listener.queue.get()
                for graph in self.graphs.values():
                    logger.debug(f"{graph.gen.record.ip_address} == {address.url}")
                    if graph.gen.record.ip_address == address.url:
                        break
                else:
                    self.add_new_by_address(address.url)

    @Slot()
    def add_item_dialog(self):
        dialog = ConnectionDialog(
            self,
            self.listener,
            self.grid_layout.count() + 1,
            "tcp://127.0.0.1:8100",
            self.sim,
        )
        dialog.open()

    def add_new_by_address(self, addr: str):
        local_logger = make_nested_logger(self.next_graph)
        gen = RemoteGeneratorGUI(
            address=addr,
            logger=local_logger,
            gen_record=GenRecordGUI(local_logger, ip_address=addr),
        )
        gen.run()
        self.add_item(gen)

    def add_new_generator(self, i: int):
        local_logger = make_nested_logger(self.next_graph)
        gen = LocalGeneratorGUI(
            i=i + 1,
            logger=local_logger,
            gen_record=GenRecordGUI(local_logger),
        )
        gen.run()
        self.add_item(gen)

    def _get_next_empty(self) -> Tuple[int, int]:
        # First, see if there's a open empty cell. If there is, return that index.
        old_ind = self.grid_layout.count()
        for i in range(old_ind):
            widget = self.grid_layout.itemAt(i).widget()
            if isinstance(widget, (EmptySensor, WaitingWidget)):
                i, j, _width, _height = self.grid_layout.getItemPosition(i)
                self.grid_layout.removeWidget(widget)
                widget.setParent(None)
                return i, j

        height, width = self.row_column_count()

        if old_ind == 0:
            self.grid_layout.setRowStretch(0, 1)
            self.grid_layout.setColumnStretch(0, 1)
            return 0, 0

        if height <= width:
            # Adding row
            self.grid_layout.setRowStretch(height, 1)
            for i in range(1, width):
                self.grid_layout.addWidget(EmptySensor(), height, i)
            return height, 0

        else:
            # Adding column
            self.grid_layout.setColumnStretch(width, 1)
            for i in range(1, height):
                self.grid_layout.addWidget(EmptySensor(), i, width)
            return 0, width

    def empty_grid(self, height: int, width: int) -> None:
        logger.debug(f"Setting up {height}x{width} empty grid")
        for i in range(height + 1):
            self.grid_layout.setRowStretch(i, 1)
        for j in range(width + 1):
            self.grid_layout.setColumnStretch(j, 1)
        for _ in range(height + 1):
            for _ in range(width + 1):
                self.grid_layout.addWidget(EmptySensor())

    def add_item(self, gen: GeneratorGUI, pos: Optional[Tuple[int, int]] = None):
        """
        If you use pos, you should make sure the grid is large enough (basically, you
        should have an empty grid). Currently only used by the restoring functionality.
        """
        ind = self.next_graph
        if (
            len(self.graphs) == 0
            and self.grid_layout.count() == 1
            and isinstance(
                self.grid_layout.itemAtPosition(0, 0).widget(), WaitingWidget
            )
        ):
            waiting = self.grid_layout.takeAt(0)
            waiting.widget().setParent(None)

        if pos is None or not isinstance(
            self.grid_layout.itemAtPosition(*pos).widget(), EmptySensor
        ):
            i, j = self._get_next_empty()
        else:
            i, j = pos
            item = self.grid_layout.itemAtPosition(i, j)
            ind = self.grid_layout.indexOf(item.widget())
            item = self.grid_layout.takeAt(ind)
            item.widget().setParent(None)

        gen.record.position = (i, j)

        drilldown: DrilldownWidget = self.parent().parent().drilldown
        drilldown.add_alarm_box(gen, i=ind)

        graph = PatientSensor(i=ind, gen=gen)
        self.graphs[ind] = graph

        self.grid_layout.addWidget(graph, i, j)
        graph.set_plot()

        self.next_graph += 1

    def drop_item(self, i: int) -> None:
        graph: PatientSensor = self.graphs.pop(i)
        record: GenRecordGUI = graph.gen.record
        record.active = False
        graph.gen.close()
        ind = self.grid_layout.indexOf(graph)
        x, y, *_ = self.grid_layout.getItemPosition(ind)
        self.grid_layout.removeWidget(graph)
        self.grid_layout.addWidget(EmptySensor(), x, y)
        graph.setParent(None)
        drilldown: DrilldownWidget = self.parent().parent().drilldown
        drilldown.drop_alarm_box(i)

        self.drop_final_row_or_column_if_needed()

    def row_column_count(self):
        height = self.grid_layout.rowCount()
        width = self.grid_layout.columnCount()

        rows = [False] * height
        columns = [False] * width

        for row in range(height):
            for column in range(width):
                if self.grid_layout.itemAtPosition(row, column) is not None:
                    rows[row] = True
                    columns[column] = True
        return sum(rows), sum(columns)

    def drop_final_row_or_column_if_needed(self) -> None:
        height, width = self.row_column_count()

        # Don't drop the final tile!
        if height <= 1 and width <= 1:
            return

        widgets = [
            self.grid_layout.itemAtPosition(row, width - 1).widget()
            for row in range(height)
        ]

        if all(isinstance(widget, EmptySensor) for widget in widgets):
            for widget in widgets:
                self.grid_layout.removeWidget(widget)
                widget.setParent(None)
            self.grid_layout.setColumnStretch(width - 1, 0)

            return self.drop_final_row_or_column_if_needed()

        widgets = [
            self.grid_layout.itemAtPosition(height - 1, column).widget()
            for column in range(width)
        ]

        if all(isinstance(widget, EmptySensor) for widget in widgets):
            for widget in widgets:
                self.grid_layout.removeWidget(widget)
                widget.setParent(None)
            self.grid_layout.setRowStretch(height - 1, 0)

            return self.drop_final_row_or_column_if_needed()

    @Slot()
    def update_plots(self) -> None:
        if self.isVisible():
            for graph in self.graphs.values():
                graph.update_plot()
            self.qTimer.start(50)
        else:
            self.qTimer.start(500)


class MatchingDialog(QtWidgets.QDialog):
    def __init__(self):
        super().__init__()
        self.msg = QtWidgets.QLabel()
        layout = QtWidgets.QVBoxLayout(self)
        layout.addWidget(self.msg)

    def setText(self, *args, **kwargs):
        self.msg.setText(*args, **kwargs)


class MainWindow(QtWidgets.QMainWindow):
    def __init__(self, *, displays: Optional[int], listener: FindBroadcasts, **kwargs):
        super().__init__()
        self.setObjectName("MainWindow")

        pg.setConfigOption("background", (0, 0, 0, 0))

        gis = GraphInfo()

        with open(style_path) as f:
            s = Template(f.read())
            t = s.substitute(gis.graph_pens)
            self.setStyleSheet(t)

        stacked_widget = QtWidgets.QStackedWidget(self)
        self.setCentralWidget(stacked_widget)

        self.drilldown = DrilldownWidget(stacked_widget)
        self.main_stack = MainStack(
            stacked_widget, listener=listener, displays=displays, **kwargs
        )

        stacked_widget.addWidget(self.main_stack)
        stacked_widget.addWidget(self.drilldown)

        self.main_stack.header.fs_exit.clicked.connect(self.close)
        self.drilldown.return_btn.clicked.connect(self.drilldown_deactivate)

        self.was_maximized: bool = False

    @Slot()
    def toggle_fs(self) -> None:
        if self.isFullScreen():
            if self.was_maximized:
                self.showMaximized()
            else:
                self.showNormal()
        else:
            self.was_maximized = self.isMaximized()
            self.showFullScreen()

    @Slot()
    def drilldown_deactivate(self) -> None:
        self.drilldown.deactivate()
        stacked_widget = self.centralWidget()
        stacked_widget.setCurrentIndex(0)

    def drilldown_activate(self, i: int) -> None:
        self.drilldown.activate(i)
        stacked_widget = self.centralWidget()
        stacked_widget.setCurrentIndex(1)

    def keyPressEvent(self, event):
        if event.key() == Qt.Key_Escape:
            stacked_widget = self.centralWidget()
            if stacked_widget.currentIndex() != 0:
                self.drilldown_deactivate()

    def changeEvent(self, evt):
        super().changeEvent(evt)
        if evt.type() == QtCore.QEvent.WindowStateChange:
            self.main_stack.header.fs_exit.setVisible(
                self.windowState() & Qt.WindowFullScreen
            )

    def closeEvent(self, evt):
        for graph in self.main_stack.graphs.values():
            graph.gen.close()
        super().closeEvent(evt)
```
