## Supplementary Git Repository for "Inexpensive multi-patient respiratory monitoring system for helmet ventilation during COVID-19 pandemic": qt.py

```
# Names are selected to be like PySide
from __future__ import annotations

from typing import Optional, TYPE_CHECKING

if TYPE_CHECKING:
    from typing_extensions import Protocol
else:

    class Protocol:
        pass


import os

if "PYQTGRAPH_QT_LIB" in os.environ:
    os.environ["PYQTGRAPH_QT_LIB"] = "PyQt5"

from pyqtgraph.Qt import QtCore, QtWidgets, QtGui

Qt = QtCore.Qt

try:
    Slot = QtCore.Slot
    Signal = QtCore.Signal
except AttributeError:
    Slot = QtCore.pyqtSlot
    Signal = QtCore.pyqtSignal


class BoxName(QtWidgets.QLabel):
    pass


class PopdownTitle(QtWidgets.QLabel):
    pass


# Layout factory functions
def HBoxLayout(*args, **kwargs) -> QtWidgets.QHBoxLayout:
    layout = QtWidgets.QHBoxLayout(*args, **kwargs)
    layout.setSpacing(0)
    layout.setContentsMargins(0, 0, 0, 0)
    return layout


def VBoxLayout(*args, **kwargs) -> QtWidgets.QVBoxLayout:
    layout = QtWidgets.QVBoxLayout(*args, **kwargs)
    layout.setSpacing(0)
    layout.setContentsMargins(0, 0, 0, 0)
    return layout


def FormLayout(*args, **kwargs) -> QtWidgets.QFormLayout:
    layout = QtWidgets.QFormLayout(*args, **kwargs)
    layout.setContentsMargins(0, 0, 0, 0)
    layout.setSpacing(0)
    return layout


def GridLayout(*args, **kwargs) -> QtWidgets.QGridLayout:
    layout = QtWidgets.QGridLayout(*args, **kwargs)
    layout.setContentsMargins(0, 0, 0, 0)
    layout.setSpacing(0)
    return layout


def update_textbox(textbox, text):
    if textbox.toPlainText() != text:
        val = textbox.verticalScrollBar().value()
        textbox.setText(text)
        textbox.verticalScrollBar().setValue(val)


def swap_grid(
    grid_layout: QtWidgets.QGridLayout,
    source: QtWidgets.QWidget,
    target: QtWidgets.QWidget,
) -> None:
    s_idx = grid_layout.indexOf(source)
    t_idx = grid_layout.indexOf(target)
    s_x, s_y, *_ = grid_layout.getItemPosition(s_idx)
    t_x, t_y, *_ = grid_layout.getItemPosition(t_idx)
    source_item = grid_layout.itemAtPosition(s_x, s_y)
    target_item = grid_layout.itemAtPosition(t_x, t_y)
    grid_layout.removeItem(source_item)
    grid_layout.removeItem(target_item)
    grid_layout.addItem(source_item, t_x, t_y)
    grid_layout.addItem(target_item, s_x, s_y)


class DraggableProtocol(Protocol):
    def childAt(self, _: QtCore.QPoint) -> Optional[QtWidgets.QWidget]:
        ...

    def close(self) -> None:
        ...

    def move(self, x: float, y: float) -> None:
        ...

    @property
    def offset(self) -> QtCore.QPoint:
        ...

    @offset.setter
    def offset(self, value: QtCore.QPoint) -> None:
        ...


class DraggableMixin:
    def __init__(self, *args, **kwargs):
        self.offset = None

    def mousePressEvent(self: DraggableProtocol, event):
        self.offset = event.pos()

        # Only required on Linux
        if self.childAt(event.pos()) is None:
            self.close()

    def mouseMoveEvent(self: DraggableProtocol, event):
        if self.offset is not None:
            x = event.globalX()
            y = event.globalY()
            x_w = self.offset.x()
            y_w = self.offset.y()
            self.move(x - x_w, y - y_w)

    def mouseReleaseEvent(self, a0: QtGui.QMouseEvent) -> None:
        self.offset = None
```
