## Supplementary Git Repository for "Inexpensive multi-patient respiratory monitoring system for helmet ventilation during COVID-19 pandemic": recover.py

```
from __future__ import annotations

import yaml
from processor.config import get_data_dir
from typing import Dict, Tuple
from dataclasses import dataclass


@dataclass
class Restore:
    ip_address: str
    position: Tuple[int, int]


def collect_restore() -> Dict[str, Restore]:
    results: Dict[str, Restore] = {}

    for filepath in (get_data_dir() / "nurse_layout").glob("*.yml"):
        with filepath.open() as f:
            d = yaml.safe_load(f)
            if "ip_address" in d and d.get("active"):
                i, j = d.get("position", (0, 0))
                results[filepath.stem] = Restore(
                    ip_address=d["ip_address"],
                    position=(i, j),
                )

    return results


def restore_limits(results: Dict[str, Restore]) -> Tuple[int, int]:
    return (
        max(r.position[0] for r in results.values()),
        max(r.position[1] for r in results.values()),
    )
```
