## Supplementary Git Repository for "Inexpensive multi-patient respiratory monitoring system for helmet ventilation during COVID-19 pandemic": test_common.py

```
import numpy as np

from nurse.common import rolling_mean


def test_rolling_mean():
    arr = np.array([1, 2, 3, 4, 5, 6, 7])
    res = np.array([1.5, 2, 3, 4, 5, 6, 6.5])
    rmean = rolling_mean(arr, 3)
    assert len(rmean) == 7
    np.testing.assert_allclose(res, rolling_mean(arr, 3))

    arr = np.array([0, 3, 0, 0, 0, 3, 0])
    res = np.array([1.5, 1, 1, 0, 1, 1, 1.5])
    np.testing.assert_allclose(res, rolling_mean(arr, 3))
```
