## Supplementary Git Repository for "Inexpensive multi-patient respiratory monitoring system for helmet ventilation during COVID-19 pandemic": nurse_read.py

```
#!/usr/bin/env python3

from processor.argparse import ArgumentParser

parser = ArgumentParser(log_dir=None, log_stem=None)
parser.add_argument("-n", default=2.0, type=float, help="Timeout between reporting")
parser.add_argument("--ip", default="127.0.0.1", help="Select an ip address")
parser.add_argument(
    "--port", type=int, help="Select a starting port (8100 recommended)"
)

arg = parser.parse_args()

import time
import numpy as np
import logging

from processor.local_generator import LocalGenerator
from processor.remote_generator import RemoteGenerator
from processor.generator import Generator

logger = logging.getLogger("povm")


gen: Generator

if arg.port is not None:
    address = f"tcp://{arg.ip}:{arg.port}"
    print(f"Remote: {address}")
    gen = RemoteGenerator(address=address, logger=logger)
else:
    print("Local Generator")
    gen = LocalGenerator(i=0, logger=logger)

print(f"Reporting every {arg.n} seconds, use Ctrl-C to exit.")

with gen:
    while True:
        time.sleep(arg.n)

        with np.printoptions(threshold=6, precision=4, floatmode="fixed", linewidth=95):
            print("--- Current status ---")
            print(f"gen.timestamps:    {gen.timestamps} ({len(gen.timestamps)} length)")
            print(f"gen.flow:          {gen.flow} ({len(gen.flow)} length)")
            print(f"gen.pressure:      {gen.pressure} ({len(gen.pressure)} length)")
            print(f"gen.volume (calc): {gen.volume} ({len(gen.volume)} length)")
            print()
```
