## Supplementary Git Repository for "Inexpensive multi-patient respiratory monitoring system for helmet ventilation during COVID-19 pandemic": nursegui.py

```
#!/usr/bin/env python3

from __future__ import annotations

import sys
import signal
import logging

from nurse.qt import QtWidgets, QtGui
from processor.argparse import ArgumentParser
from processor.listener import FindBroadcasts
from nurse.main_window import MainWindow

logger = logging.getLogger("povm")


def main(argv, *, window: bool, **kwargs):

    if "Fusion" in QtWidgets.QStyleFactory.keys():
        QtWidgets.QApplication.setStyle(QtWidgets.QStyleFactory.create("Fusion"))
    else:
        print("Fusion style is not available, display may be platform dependent")

    logger.info("Starting nursegui")

    app = QtWidgets.QApplication(argv)

    with FindBroadcasts() as listener:
        main_window = MainWindow(listener=listener, **kwargs)
        size = app.screens()[0].availableSize()
        if size.width() < 2000 or size.height() < 1200:
            main_window.resize(int(size.width() * 0.95), int(size.height() * 0.85))
            main_window.was_maximized = True
        else:
            main_window.resize(1920, 1080)
            main_window.was_maximized = False

        if not window:
            main_window.showFullScreen()
        elif main_window.was_maximized:
            main_window.showMaximized()
        else:
            main_window.showNormal()

        fs_shortcut = QtWidgets.QShortcut(QtGui.QKeySequence.FullScreen, main_window)
        fs_shortcut.activated.connect(main_window.toggle_fs)

        def ctrl_c(_sig_num, _stack_frame):
            signal.signal(signal.SIGINT, signal.SIG_DFL)
            main_window.close()

        signal.signal(signal.SIGINT, ctrl_c)
        sys.exit(app.exec())


if __name__ == "__main__":
    parser = ArgumentParser(
        description="Princeton Open Vent Monitor, nurse station graphical interface.",
        allow_abbrev=False,
        log_dir="nurse_log",
        log_stem="nursegui",
    )
    parser.add_argument("addresses", nargs="*", help="IP addresses to include")
    parser.add_argument(
        "--sim",
        action="store_true",
        help="Read from fake sim instead of remote generators (cannot be passed with addresses)",
    )
    parser.add_argument(
        "--fresh",
        action="store_true",
        help="Don't try to restore previous boxes, but instead only show detected boxes",
    )
    parser.add_argument(
        "--window", action="store_true", help="Open in window instead of fullscreen"
    )
    parser.add_argument(
        "--displays",
        "-n",
        type=int,
        help="# of displays (Dynamic if not given)",
    )

    args, unparsed_args = parser.parse_known_args()

    if args.displays is not None and len(args.addresses) > args.displays:
        print(
            "Can't start with more addresses than displays. "
            "Increase one or decrease the other."
        )
        sys.exit(1)

    addresses = [
        f"tcp://{addr}" + ("" if ":" in addr else ":8100") for addr in args.addresses
    ]

    if args.addresses and args.sim:
        print("Cannot give addresses and sim together")
        sys.exit(1)

    main(
        argv=sys.argv[:1] + unparsed_args,
        addresses=addresses,
        sim=args.sim,
        displays=args.displays,
        window=args.window,
        fresh=args.fresh,
    )
```
