## Supplementary Git Repository for "Inexpensive multi-patient respiratory monitoring system for helmet ventilation during COVID-19 pandemic": adctest.py

```
#!/usr/bin/env python3

# MCP3008 ADC readings - SPI handled by spidev.SpiDev()
import time
import signal
import spidev

# ------------------
# output file setup
# ------------------
# outputFileName = "patient.dat"
# f = open(outputFileName, "w")
# sys.stdout = f
# ------------------
# output file end of setup
# ------------------
# ------------------
# MCP3008 ADC setup
# ------------------
# Establish SPI device on Bus 0, Device 0
spiMCP3008 = spidev.SpiDev()
spiMCP3008.open(0, 0)
spiMCP3008.max_speed_hz = 500000
chanMP3V5004 = 0


def getADC(channel):
    # Check channel valid
    # if ((channel > 7) or (channel < 0)):
    #  return -1
    # Perform SPI (spiMCP3008.xfer2 keeps CS asserted)
    r = spiMCP3008.xfer2([1, (8 + channel) << 4, 0])
    # Reformat
    adcOut = ((r[1] & 3) << 8) + r[2]
    return adcOut


# ------------------
# MCP3008 ADC end of setup
# ------------------
# sdp3 interrupt handler
def sdp3_handler(signum, frame):
    ts = time.time()
    tmpADC = getADC(chanMP3V5004)
    print(ts, tmpADC)


signal.signal(signal.SIGALRM, sdp3_handler)
signal.setitimer(signal.ITIMER_REAL, 1, 0.01)  # 10Hz of readout

# event loop:
#    wait for readout of diff pressure sensor and pressure sensor (signal interrupt handler)
while True:
    signal.pause()
    # time.sleep(1)  # 1 second
    # update display with alarmThreshold1 and setThreshold1 status
```
