## Supplementary Git Repository for "Inexpensive multi-patient respiratory monitoring system for helmet ventilation during COVID-19 pandemic": backlight.py

```
#!/usr/bin/env python3

import pigpio
from typing import Optional


class Backlight:
    PIN_RED = 16
    PIN_GREEN = 20
    PIN_BLUE = 21

    def __init__(self, *, shade: int = 255, pi: Optional[pigpio.pi] = None):
        self.pi = pi
        self.shade = shade

    def __enter__(self) -> "Backlight":
        if self.pi is None:
            self.pi = pigpio.pi()
        return self

    def color(self, red: int, green: int, blue: int):
        assert self.pi is not None, 'Must use "with" to use'
        self.pi.set_PWM_dutycycle(self.PIN_RED, red)
        self.pi.set_PWM_dutycycle(self.PIN_GREEN, green)
        self.pi.set_PWM_dutycycle(self.PIN_BLUE, blue)

    def white(self, shade: int = None) -> None:
        shade = self.shade if shade is None else shade
        self.color(shade, shade, shade)

    def red(self, shade: int = None, *, light=False) -> None:
        zero = self.shade // 3 if light else 0
        shade = self.shade if shade is None else shade
        self.color(shade, zero, zero)

    def green(self, shade: int = None, *, light=False) -> None:
        zero = self.shade // 3 if light else 0
        shade = self.shade if shade is None else shade
        self.color(zero, shade, zero)

    def blue(self, shade: int = None, *, light=False) -> None:
        zero = self.shade // 3 if light else 0
        shade = self.shade if shade is None else shade
        self.color(zero, zero, shade)

    def cyan(self, shade: int = None, *, light=False) -> None:
        zero = self.shade // 3 if light else 0
        shade = self.shade if shade is None else shade
        self.color(zero, shade, shade)

    def magenta(self, shade: int = None, *, light=False) -> None:
        zero = self.shade // 3 if light else 0
        shade = self.shade if shade is None else shade
        self.color(shade, zero, shade)

    def yellow(self, shade: int = None, *, light=False) -> None:
        zero = self.shade // 3 if light else 0
        shade = self.shade if shade is None else shade
        self.color(shade, shade, zero)

    def orange(self, shade: int = None) -> None:
        shade = self.shade if shade is None else shade
        self.color(shade, shade // 4, 0)

    def black(self):
        self.color(0, 0, 0)

    def __exit__(self, *exc):
        pass


if __name__ == "__main__":
    with Backlight() as b:
        b.black()
```
