## Supplementary Git Repository for "Inexpensive multi-patient respiratory monitoring system for helmet ventilation during COVID-19 pandemic": buzzer.py

```
#!/usr/bin/env python3

import pigpio
import threading

from typing import Optional, TypeVar

T = TypeVar("T", bound="Buzzer")


class Buzzer:
    PIN = 19

    def __init__(self, *, pi: Optional[pigpio.pi] = None):
        self.pi = pi
        self._owns_pi = False
        self._done = threading.Event()
        self._lock = threading.Lock()
        self._thread: Optional[threading.Thread] = None

    def __enter__(self: T) -> T:
        if self.pi is None:
            self.pi = pigpio.pi()
            self._owns_pi = True
        return self

    def buzz(self, volume: int) -> None:
        assert self.pi is not None, 'Must use "with" to use'

        with self._lock:
            self.pi.set_PWM_dutycycle(self.PIN, volume)

    def _thread_pattern(
        self, volume: int, time_on: float, time_off: float, timer: Optional[float]
    ) -> None:
        start_time = time.monotonic()

        while not self._done.is_set():
            self.buzz(volume)
            self._done.wait(time_on)
            self.buzz(0)
            self._done.wait(time_off)
            if timer is not None and timer + start_time > time.monotonic():
                break
        self._done.clear()

    def clear(self) -> None:
        if self._thread is not None:
            self._done.set()
            self._thread.join()
            self._thread = None
        self.buzz(0)

    def buzz_pattern(
        self,
        volume: int,
        time_on: float,
        time_off: float = None,
        *,
        timer: Optional[float] = None,
    ) -> None:
        if time_off is None:
            time_off = time_on

        self._thread = threading.Thread(
            target=self._thread_pattern, args=(volume, time_on, time_off, timer)
        )

        self._thread.start()

    def __exit__(self, *exc) -> None:
        self.clear()
        if self._owns_pi and self.pi is not None:
            self.pi.stop()
            self.pi = None


if __name__ == "__main__":
    import time

    with Buzzer() as buzzer:
        print("Preparing to buzz")
        time.sleep(1)

        for level in [50, 100, 150, 200, 225, 250, 255]:
            print(f"Buzz level: {level}")
            buzzer.buzz(level)
            time.sleep(1)
        print("Buzz off")
        time.sleep(1)
        print("Buzz pattern test at .2s, 200 volume")
        buzzer.buzz_pattern(200, 0.05)
        time.sleep(2)
        buzzer.clear()
        print("Buzz off")
        time.sleep(1)

        print("Buzz pattern test at .1s, 255 volume")
        buzzer.buzz_pattern(250, 0.3)
        time.sleep(2)
        buzzer.clear()
```
