## Supplementary Git Repository for "Inexpensive multi-patient respiratory monitoring system for helmet ventilation during COVID-19 pandemic": lcd.py

```
#!/usr/bin/env python3
# mypy: disallow_untyped_defs
# mypy: disallow_incomplete_defs

from __future__ import annotations

import pigpio
import time
from typing import Union, Any
import enum


class Align(enum.Enum):
    RIGHT = enum.auto()
    CENTER = enum.auto()
    LEFT = enum.auto()


DEVICE_LCD = 0x3C  # Slave 0x78 << 1
pinRST = 26  # Soft Reset pin GPIO26


class LCD:
    def __init__(self, *, pi: pigpio.pi = None):
        self.pi = pi

    def __enter__(self) -> LCD:
        # Get pigio connection
        if self.pi is None:
            self.pi = pigpio.pi()

        # Get I2C bus handle
        self.hLCD = self.pi.i2c_open(6, DEVICE_LCD)

        # initialize
        self.reset()  # issue soft reset to LCD
        time.sleep(0.04)  # wait 40ms

        self.clear()  # Needed to fix flakiness when restarting
        time.sleep(0.002)
        self.ctrl(0x38)  # Function set - 8 bit, 2 line, norm height, inst table 0
        time.sleep(0.01)

        self.ctrl(0x39)  # Function set - 8 bit, 2 line, norm height, inst table 1
        time.sleep(0.01)

        self.ctrl(0x14)  # Set bias 1/5
        self.ctrl(0x78)  # Set contrast low
        self.ctrl(0x5E)  # ICON display, Booster on, Contrast high
        time.sleep(1.0)

        self.ctrl(0x6D)  # Font on, Amp ratio 6
        time.sleep(0.3)

        self.ctrl(0x0C)  # Display on, Cursor off, Cursor Pos off
        self.clear()
        time.sleep(0.002)

        self.ctrl(0x06)  # Entry mode increment

        return self

    def reset(self) -> None:
        assert self.pi is not None, 'Must use "with" to use'
        self.pi.write(pinRST, 0)
        time.sleep(0.002)
        self.pi.write(pinRST, 1)

    def ctrl(self, value: int) -> None:
        assert self.pi is not None, 'Must use "with" to use'
        self.pi.i2c_write_device(self.hLCD, [0x00, value])

    def text(self, text: str) -> None:
        assert self.pi is not None, 'Must use "with" to use'
        b = text.encode("ascii")
        self.pi.i2c_write_device(self.hLCD, [0x40] + [*b])

    def upper(
        self, text: str, pos: Union[int, Align] = Align.LEFT, fill: str = ""
    ) -> None:
        self._either(0x80, text, pos, fill)

    def lower(
        self, text: str, pos: Union[int, Align] = Align.LEFT, fill: str = ""
    ) -> None:
        self._either(0xC0, text, pos, fill)

    def _either(self, start: int, text: str, pos: Union[int, Align], fill: str) -> None:
        assert self.pi is not None, 'Must use "with" to use'
        if pos == Align.LEFT:
            pos = 0
        elif pos == Align.CENTER:
            pos = (20 - len(text)) // 2
        else:
            pos = 20 - len(text)

        if fill:
            text = fill * pos + text + fill * (20 - len(text))
            self.ctrl(start)
        else:
            self.ctrl(start + pos)

        self.text(text)

    def clear(self) -> None:
        self.ctrl(0x01)
        time.sleep(0.002)

    def __exit__(self, *exc: Any) -> None:
        assert self.pi is not None, 'Must use "with" to use'
        self.pi.i2c_close(self.hLCD)


if __name__ == "__main__":
    with LCD() as lcd:
        lcd.upper("HELLO", Align.CENTER)
        lcd.lower("WORLD", Align.CENTER)
        time.sleep(1)
        lcd.clear()
```
