## Supplementary Git Repository for "Inexpensive multi-patient respiratory monitoring system for helmet ventilation during COVID-19 pandemic": mac_address.py

```
from functools import lru_cache
import time
from pathlib import Path
from processor.device_names import address_to_name
import os

import logging

logger = logging.getLogger("povm")


@lru_cache(1)
def get_mac_addr() -> str:
    """
    Blocks until eth0 is up (linux) or returns all 0's on macOS (en0).
    """

    if not Path("/sys").exists():
        return "00:00:00:00:00:00"
    while True:
        try:
            mac = open("/sys/class/net/eth0/address").read().strip()
            logger.info(f"MAC Addr: {mac}")
            return mac
        except IOError:
            if "POVM_MACADDR" in os.environ:
                return os.environ["POVM_MACADDR"]
            time.sleep(1)


@lru_cache(1)
def get_box_name() -> str:
    try:
        return address_to_name(get_mac_addr()).title()
    except ValueError:
        return "<unknown>"
```
