## Supplementary Git Repository for "Inexpensive multi-patient respiratory monitoring system for helmet ventilation during COVID-19 pandemic": pwmtest.py

```
#!/usr/bin/env python3

# hardware interfaces:
# heater - PWM handled by pigpio.pi()
import pigpio

# ------------------
# PWM setup
# ------------------
# setup pin used for PWM
pinPWM = 13
pi = pigpio.pi()
# pi.set_PWM_range(pinPWM, 100) # renormalizes range to 100 instead of 255 if desired
# duty cycle out of 255
dc = 200
pi.set_PWM_dutycycle(pinPWM, dc)

pi.stop()
```
