## Supplementary Git Repository for "Inexpensive multi-patient respiratory monitoring system for helmet ventilation during COVID-19 pandemic": README.md

## Scripts for running simulation in patient mode
python3 patient_sim.py | python3 read_patient_sim.py

patient_sim.py reads simulation data at pseudo real-time speed, sends it via json to stdout. This runs for 40 seconds and then stops.
read_patient_sim.py reads stdin picks up the jsons that are sensor data and adds them to rolling buffers

## Data format (sensor -> patient event loop):

```json
{"v": 1, "t": 1587514788871, "P": 955.625, "F": -1.23, "temp":23.3}
```

One [JSON object](https://www.json.org/json-en.html) per line of text (delimited by `"\n"`, no `"\n"` allowed within a JSON object) at a rate of roughly 50 Hz. Field names are case-sensitive.

   * The `"v"` field (mandatory) is the version of the protocol; always `1`. Future versions are also integers (and version updates will be rare).
   * The `"t"` field (mandatory) is time as a floating-point number of milliseconds relative to an unspecified constant, which does not change through the interval in which the data collection box is connected to the nurse's station (with a given port number). Nominally, it is the number of milliseconds since Jan 1, 1970, but the offset isn't guaranteed.
   * The `"P"` field (mandatory) is [intrapleural pressure](https://en.wikipedia.org/wiki/Intrapleural_pressure) (difference between pressure in lungs and atmospheric pressure) in cm H2O (floating point). Positive pressure is greater than atmospheric; negative is less than atmospheric.
   * The `"F"` field (mandatory) is the flow rate in mL/sec (floating point). The volume can be computed by strictly integrating this field with respect to time. Positive flow is into the lungs; negative flow is out.
   * The `"temp"` field is optional, and in fact is only populated in about 1 out of 50 lines (1 Hz). It is the temperature of the flow sensor in degrees C (floating point).

## Data format (patient -> nurse)

Note: comments inserted for clarity only - JSON does not allow comments.

```yaml
{
  "version": 1,
  "source": "Sim.send",
  "parameters": {
    # Simulation parameters here
  },
  "calibration": {
    # Calibration parameters here
  },
  "time":1234.343,   # Current packet timestamp
  "alarms": {
    "MinFlow": false,
    # Status of patient alarms here
  },
  "data": {
    "timestamps": [1000,1002,1004,1006],
    "flows": [23.2, 12.2, 123.2, -2.2],
    "pressures": [1.2, 3.4, 5.6, 7.8],
    "volumes": [1.2, 3.4, 5.6, 7.8]
  }
}
```
