## Supplementary Git Repository for "Inexpensive multi-patient respiratory monitoring system for helmet ventilation during COVID-19 pandemic": rotary.py

```
#!/usr/bin/env python3
# mypy: disallow_untyped_defs
# mypy: disallow_incomplete_defs
from __future__ import annotations

import pigpio
from patient.rotary_live import LiveRotary
from processor.setting import Setting
import enum
import threading
from typing import Dict, Any, Optional, TypeVar, TYPE_CHECKING
import time
import logging

logger = logging.getLogger("povm")

if TYPE_CHECKING:
    from typing_extensions import Final

pinA: Final[int] = 17  # terminal A
pinB: Final[int] = 27  # terminal B
pinSW: Final[int] = 18  # switch
pinExt: Final[int] = 12  # Red pushbutton (added in v0.6)


class Mode(enum.Enum):
    EDIT = enum.auto()
    ALARM = enum.auto()


class Dir(enum.Enum):
    CLOCKWISE = 1
    COUNTERCLOCKWISE = -1


MT = TypeVar("MT", bound="MechanicalRotary")


class MechanicalRotary:
    def __init__(self, *, pi: Optional[pigpio.pi] = None):
        self.pi = pi
        self.pushed_in = False
        self.extra_in = False
        self._last_touched = time.monotonic()
        self.turns = 0
        self.levA = 0
        self.levB = 0
        self._last: Optional[int] = None
        self._ts = time.monotonic()
        self._filter = 0.02

    def __enter__(self: MT) -> MT:
        # Filter button pushes the normal way
        glitchFilterSW = 1_000  # microseconds
        glitchFilterExt = 10_000  # microseconds

        # Get pigio connection
        if self.pi is None:
            self.pi = pigpio.pi()

        self.pi.set_mode(pinA, pigpio.INPUT)
        self.pi.set_pull_up_down(pinA, pigpio.PUD_UP)

        self.pi.set_mode(pinB, pigpio.INPUT)
        self.pi.set_pull_up_down(pinB, pigpio.PUD_UP)

        self.pi.set_mode(pinSW, pigpio.INPUT)
        self.pi.set_pull_up_down(pinSW, pigpio.PUD_UP)
        self.pi.set_glitch_filter(pinSW, glitchFilterSW)

        self.pi.set_mode(pinExt, pigpio.INPUT)
        self.pi.set_pull_up_down(pinExt, pigpio.PUD_UP)
        self.pi.set_glitch_filter(pinExt, glitchFilterExt)

        # The callbacks get fired sometimes during startup
        time.sleep(0.2)

        self._rotary_turnedA = self.pi.callback(
            pinA, pigpio.EITHER_EDGE, self.rotary_turned
        )
        self._rotary_turnedB = self.pi.callback(
            pinB, pigpio.EITHER_EDGE, self.rotary_turned
        )
        self._rotary_switch = self.pi.callback(
            pinSW, pigpio.EITHER_EDGE, self.rotary_switch
        )
        self._rotary_extra = self.pi.callback(
            pinExt, pigpio.EITHER_EDGE, self.rotary_switch
        )

        return self

    def __exit__(self, *exc: Any) -> None:
        assert self.pi is not None, 'Must use "with" to use'
        self._rotary_turnedA.cancel()
        self._rotary_turnedB.cancel()
        self._rotary_switch.cancel()
        self._rotary_extra.cancel()

    def rotary_turned(self, ch: int, level: int, _tick: int) -> None:
        # Store last reading
        if ch == pinA:
            self.levA = level
        elif ch == pinB:
            self.levB = level

        # Debounce
        if self._last == ch:
            return
        self._last = ch

        # Pick the correct function to call
        if self.extra_in:
            function = self.extra_turn
        elif self.pushed_in:
            function = self.pushed_turn
        else:
            function = self.turn

        ts = time.monotonic()
        if level == 1 and ts > (self._ts + self._filter):
            if ch == pinA and self.levB == 1:
                self._ts = ts
                function(Dir.COUNTERCLOCKWISE)
            elif ch == pinB and self.levA == 1:
                self._ts = ts
                function(Dir.CLOCKWISE)

    def rotary_switch(self, ch: int, level: int, _tick: int) -> None:

        if ch == pinSW:
            # Allow rotations to tell if this is pushed in or out
            self.pushed_in = level == 0

            if level == 0:  # falling edge
                self.push()
            elif level == 1:  # rising edge
                self.release()

        elif ch == pinExt:
            # Allow rotations to tell if this is pushed in or out
            self.extra_in = level == 0

            if level == 0:  # falling edge
                self.extra_push()
            elif level == 1:  # rising edge
                self.extra_release()

    def pushed_turn(self, dir: Dir) -> None:
        self._last_touched = time.monotonic()
        logger.debug(f"Push + turn {dir}")

    def extra_turn(self, dir: Dir) -> None:
        self._last_touched = time.monotonic()
        logger.debug(f"Extra + turn {dir}")

    def turn(self, dir: Dir) -> None:
        self._last_touched = time.monotonic()
        logger.debug(f"Turned {dir}")

    def push(self) -> None:
        self._last_touched = time.monotonic()
        logger.debug("Pressed")

    def release(self) -> None:
        self._last_touched = time.monotonic()
        logger.debug("Released")

    def extra_push(self) -> None:
        self._last_touched = time.monotonic()
        logger.debug("Extra pressed")

    def extra_release(self) -> None:
        self._last_touched = time.monotonic()
        logger.debug("Extra released")


T = TypeVar("T", bound="Rotary")


class Rotary(LiveRotary, MechanicalRotary):
    def __init__(self, config: Dict[str, Setting], *, pi: pigpio.pi = None):
        LiveRotary.__init__(self, config)
        MechanicalRotary.__init__(self, pi=pi)

        self._current: int = 0
        self._slow_turn: int = 0

        # Rate of changing screens on the display
        self._change_rate: Final[int] = 4

        # Timestamp at which to start the alarm again - will be silenced until this time
        self._alarm_silence: float = time.monotonic()

        # Singleton Timer that will run alarm() when done.
        self._time_out_alarm: Optional[threading.Timer] = None

        # Singleton Timer that will start an alarm if not cancelled
        self._delay_timout_setter: Optional[threading.Timer] = None

        # Lock this to modify the delay timer
        self.delay_lock = threading.RLock()

    def last_interaction(self) -> float:
        return self._last_touched

    def time_left(self) -> float:
        """
        Amount of time left on the silencer. Negative if silence is off.
        """
        return self._alarm_silence - time.monotonic()

    def set_alarm_silence(self, value: float, *, reset: bool = True) -> None:
        with self.delay_lock:
            if self._time_out_alarm is not None:
                # If we are not resetting, do not touch anything
                if not reset:
                    return

                self._time_out_alarm.cancel()

            def timeout() -> None:
                self._time_out_alarm = None
                logger.debug("Silence timed out")
                self.alert()

            self._alarm_silence = time.monotonic() + value
            self._time_out_alarm = threading.Timer(value, timeout)
            self._time_out_alarm.start()
            logger.info(
                f"Setting silence timeout to {value} s, ends at {self._alarm_silence}"
            )
            if self._delay_timout_setter is not None:
                self._delay_timout_setter = None

    def delayed_set_alarm_silence(self, value: float, *, delay: float) -> None:
        "Activate alarm silence, but only after a time - can be canceled"
        with self.delay_lock:
            if self._delay_timout_setter is not None:
                self._delay_timout_setter.cancel()
                self._delay_timout_setter = None

            self._delay_timout_setter = threading.Timer(
                delay, self.set_alarm_silence, args=(value,)
            )
            self._delay_timout_setter.start()

    def turn(self, dir: Dir) -> None:
        self._slow_turn = (self._slow_turn + dir.value) % (
            len(self.config) * self._change_rate
        )

        old_current = self._current
        self._current = self._slow_turn // self._change_rate

        # This resets "auto-reset" settings (reset, advanced) when they are selected
        if self._current != old_current:
            self.value().active()

        super().turn(dir)

    def pushed_turn(self, dir: Dir) -> None:
        if dir == Dir.CLOCKWISE:
            self.value().up()
        else:
            self.value().down()

        self.changed()
        super().pushed_turn(dir)

    def key(self) -> str:
        return self._items[self._current]

    def value(self) -> Setting:
        return self.config[self._items[self._current]]

    @property
    def alarms(self) -> Dict[str, Any]:
        return self._alarms

    @alarms.setter
    def alarms(self, item: Dict[str, Any]) -> None:
        if item != self._alarms:
            self._alarms = item
            self.alert()

    def alert(self) -> None:
        pass

    def __enter__(self: T) -> T:
        # Get pigio connection
        if self.pi is None:
            self.pi = pigpio.pi()

        LiveRotary.__enter__(self)
        MechanicalRotary.__enter__(self)
        return self

    def __exit__(self, *exc: Any) -> None:
        if self._time_out_alarm is not None:
            self._time_out_alarm.cancel()
            self._time_out_alarm = None
        LiveRotary.__exit__(self, *exc)
        MechanicalRotary.__exit__(self, *exc)


if __name__ == "__main__":

    class TestRotary(MechanicalRotary):
        def __init__(self, *args: Any, **kwargs: Any):
            super().__init__(*args, **kwargs)
            self.turns = 0

        def pushed_turn(self, dir: Dir) -> None:
            print("PUSHED:", dir)

        def turn(self, dir: Dir) -> None:
            self.turns += dir.value
            print(self.turns)

        def push(self) -> None:
            print("Pushed")

    with TestRotary():
        forever = threading.Event()
        forever.wait()
```
