## Supplementary Git Repository for "Inexpensive multi-patient respiratory monitoring system for helmet ventilation during COVID-19 pandemic": rotary_gui.py

```
from patient.rotary_live import LiveRotary
from processor.setting import SelectionSetting, IncrSetting
from processor.display_settings import AdvancedSetting, CurrentSetting, CO2Setting
from processor.generator import Generator
import time

from nurse.qt import QtCore, QtWidgets, Slot, Signal, update_textbox

from typing import Dict, Any, Union


class RedrawSettings(QtCore.QObject):
    changed = Signal()


class RotaryGUI(LiveRotary):
    signal = RedrawSettings()

    def __init__(self, *args, **kwargs):
        self._last_changed = time.monotonic()
        super().__init__(*args, **kwargs)

    def external_update(self) -> None:
        self.signal.changed.emit()

    def touched(self) -> None:
        self._last_changed = time.monotonic()

    def changed(self) -> None:
        self.touched()
        super().changed()

    def last_interaction(self) -> float:
        return self._last_changed

    def time_left(self) -> float:
        return 0.0


class SelectionSettingGUI(QtWidgets.QComboBox):
    def __init__(self, rotary: LiveRotary, setting: SelectionSetting):
        super().__init__()
        self.setting = setting
        self.rotary = rotary

        for choice in setting._listing:
            self.addItem(f"{choice}{setting.unit if setting.unit else ''}")
        self.setCurrentIndex(setting._value)
        self.setEditable(False)
        self.currentIndexChanged.connect(self.change_rotary)

    @Slot(int)
    def change_rotary(self, index: int):
        self.setting._value = index
        self.rotary.changed()
        print(self.setting)


class AdvancedSettingGUI(QtWidgets.QComboBox):
    def __init__(self, rotary: RotaryGUI, setting: AdvancedSetting):
        super().__init__()
        self.setting = setting
        self.rotary = rotary

        for i in range(len(setting)):
            self.addItem(f"{setting.print_setting(i)}")
        self.setCurrentIndex(setting._value)
        self.setEditable(False)

        rotary.signal.changed.connect(self.redraw)
        self.currentIndexChanged.connect(self.touched)

    @Slot(int)
    def touched(self, _: int) -> None:
        self.rotary.touched()

    @Slot()
    def redraw(self) -> None:
        setting = self.setting
        for i in range(len(setting)):
            self.setItemText(i, f"{setting.print_setting(i)}")


class IncrSettingGUI(QtWidgets.QDoubleSpinBox):
    def __init__(self, rotary: LiveRotary, setting: IncrSetting):
        super().__init__()
        self.setting = setting
        self.rotary = rotary

        self.setRange(setting._min, setting._max)
        self.setSingleStep(setting._incr)
        self.setValue(setting._value)
        if setting.unit is not None:
            self.setSuffix(" " + setting.unit)

        self.valueChanged.connect(self.change_rotary)

    @Slot(float)
    def change_rotary(self, value: float):
        self.setting._value = value
        self.rotary.changed()


class CurrentSettingGUI(QtWidgets.QLabel):
    def __init__(self, rotary: RotaryGUI, setting: Union[CurrentSetting, CO2Setting]):
        super().__init__(f"{setting.print_setting()}")
        self.setting = setting
        self.rotary = rotary

        rotary.signal.changed.connect(self.redraw)

    @Slot()
    def redraw(self):
        self.setText(f"{self.setting.print_setting()}")


class UpdatingDisplay(QtWidgets.QTextEdit):
    def __init__(self, gen: Generator):
        super().__init__()

        self.gen = gen
        self.setEditable = False
        self.setMinimumWidth(100)

        self.qTimer = QtCore.QTimer()
        self.qTimer.setInterval(1000)  # 1 second
        self.qTimer.timeout.connect(self.update)
        self.qTimer.start(1000)


class AlarmWidget(UpdatingDisplay):
    @Slot()
    def update(self):
        cumulative = "\n".join(f"{k}: {v}" for k, v in self.gen.cumulative.items())
        update_textbox(self, cumulative)


class CumulativeWidget(UpdatingDisplay):
    @Slot()
    def update(self):
        def expand(s: Dict[str, Any]) -> str:
            return "".join(f"\n  {k}: {v}" for k, v in s.items())

        active_alarms = "\n".join(
            f"{k}: {expand(v)}" for k, v in self.gen.alarms.items()
        )
        update_textbox(self, active_alarms)


class MainWindow(QtWidgets.QMainWindow):
    def __init__(self, rotary: RotaryGUI, gen: Generator):
        super().__init__()

        rotary.signal = RedrawSettings()

        main = QtWidgets.QWidget()
        self.setCentralWidget(main)
        layout = QtWidgets.QHBoxLayout(main)

        form_layout = QtWidgets.QFormLayout()
        layout.addLayout(form_layout)

        for setting in rotary.values():
            if isinstance(setting, AdvancedSetting):
                widget = AdvancedSettingGUI(rotary, setting)
            elif isinstance(setting, IncrSetting):
                widget = IncrSettingGUI(rotary, setting)
            elif isinstance(setting, SelectionSetting):
                widget = SelectionSettingGUI(rotary, setting)
            elif isinstance(setting, (CurrentSetting, CO2Setting)):
                widget = CurrentSettingGUI(rotary, setting)
            else:
                raise RuntimeError("Invalid type of rotary item")

            widget.setMinimumWidth(250)
            form_layout.addRow(setting.name, widget)

        alarms_layout = QtWidgets.QVBoxLayout()
        layout.addLayout(alarms_layout)

        alarms_layout.addWidget(CumulativeWidget(gen), 1)
        alarms_layout.addWidget(AlarmWidget(gen), 1)
```
