## Supplementary Git Repository for "Inexpensive multi-patient respiratory monitoring system for helmet ventilation during COVID-19 pandemic": rotary_lcd.py

```
#!/usr/bin/env python3
# mypy: disallow_untyped_defs
# mypy: disallow_incomplete_defs
from __future__ import annotations

from processor.setting import Setting
from processor.display_settings import (
    ResetSetting,
    AdvancedSetting,
    CurrentSetting,
    CO2Setting,
)
from patient.rotary import Rotary, Dir
from patient.lcd import LCD, Align
from patient.backlight import Backlight
from patient.buzzer import Buzzer
from processor.config import config as _config
from patient.mac_address import get_box_name

import pigpio
from typing import Dict, Any, Optional
import time
import threading


class RotaryLCD(Rotary):
    def __init__(
        self,
        config: Dict[str, Setting],
        pi: pigpio.pi = None,
        event: Optional[threading.Event] = None,
    ):
        super().__init__(config, pi=pi)

        shade = _config["patient"]["brightness"].get(int)

        self.lcd = LCD(pi=pi)
        self.backlight = Backlight(shade=shade, pi=pi)
        self.buzzer = Buzzer(pi=pi)
        self.buzzer_volume: int = _config["patient"]["buzzer-volume"].get(int)
        self.lock = threading.Lock()
        self.orig_timer_setting = _config["patient"]["silence-timeout"].get(int)
        self.timer_setting: Optional[int] = None
        self.waiter = threading.Event() if event is None else event
        self.silence_holddown = _config["patient"]["silence-holddown"].get(float)

    def external_update(self) -> None:
        if (
            isinstance(self.value(), (CurrentSetting, CO2Setting))
            or self.time_left() > 0
            and not self.pushed_in
        ):
            with self.lock:
                self.upper_display()
                self.lower_display()

    def __enter__(self) -> RotaryLCD:
        self.lcd.__enter__()
        self.backlight.__enter__()
        self.buzzer.__enter__()

        self.backlight.magenta()
        self.lcd.upper("POVM Box name:")
        self.lcd.lower(f"{get_box_name():<20}")
        self.waiter.wait(3)

        self.backlight.green(light=True)
        self.lcd.upper("Turn to select alarm ")
        self.lcd.lower("Push and turn to set ")
        self.waiter.wait(2)
        self.backlight.white()

        super().__enter__()

        # Start with a 10 second silence countdown - reset is unlikely to be needed
        self.set_alarm_silence(10, reset=False)

        return self

    def __exit__(self, *exc: Any) -> None:
        self.backlight.cyan()
        self.lcd.clear()
        self.lcd.upper("Princeton Open Vent")
        self.lcd.lower("Patient loop closed")

        super().__exit__(*exc)
        self.buzzer.__exit__(*exc)
        self.backlight.__exit__(*exc)
        self.lcd.__exit__(*exc)

        if self.pi is not None:
            self.pi.stop()
            self.pi = None

    def reset(self) -> None:
        for value in self.config.values():
            value.reset()

    def release(self) -> None:
        value = self.value()
        if isinstance(value, ResetSetting) and value.at_maximum():
            self.reset()
            self.lcd.lower("Reset complete")
        super().release()

    def pushed_turn(self, dir: Dir) -> None:
        with self.lock:
            # Top display keeps ID number!
            super().pushed_turn(dir)
            value = self.value()
            if not value.STATIC_UPPER:
                self.upper_display()
            self.lower_display()

    def turn(self, dir: Dir) -> None:
        super().turn(dir)
        with self.lock:
            self.lower_display()
            self.upper_display()

    def extra_push(self) -> None:
        self.timer_setting = self.orig_timer_setting
        self.delayed_set_alarm_silence(999, delay=self.silence_holddown)
        super().extra_push()
        self.alert(False)
        self.lcd.upper("Silence duration", pos=Align.CENTER, fill=" ")
        self.lcd.lower(f"to {self.timer_setting} s", pos=Align.CENTER, fill=" ")

    def extra_release(self) -> None:
        if self.timer_setting is None:
            return

        with self.delay_lock:
            if self._delay_timout_setter is None:
                self.set_alarm_silence(self.timer_setting)
            else:
                self._delay_timout_setter.cancel()
                self._delay_timout_setter = None
        super().extra_release()
        self.alert(False)
        self.display()

        if self.timer_setting is not None:
            self.timer_setting = None

    def extra_turn(self, dir: Dir) -> None:
        if self.timer_setting is None:
            return
        if dir == Dir.CLOCKWISE and self.timer_setting < 995:
            self.timer_setting += 5
        elif dir == Dir.COUNTERCLOCKWISE and self.timer_setting > 0:
            self.timer_setting -= 5
        self.lcd.lower(f"to {self.timer_setting} s", pos=Align.CENTER, fill=" ")

    def alert(self, full: bool = True) -> None:
        with self.lock:
            time_left = self.time_left()
            if self.alarms and time_left < 0 and not self.extra_in:
                self.backlight.red()
                self.buzzer.buzz(self.buzzer_volume)
            elif not self.alarms:
                self.backlight.white()
                self.buzzer.clear()
            elif time_left > 0:
                self.backlight.yellow()
                self.buzzer.clear()

            if full:
                if time_left > 0:
                    self.set_alarm_silence(time_left, reset=False)

                if isinstance(self.value(), AdvancedSetting):
                    self.upper_display()
                else:
                    self.lower_display()

        super().alert()

    def _add_alarm_text(self, string: str) -> str:
        time_left = self.time_left()
        if time_left > 0:
            char = "Q" if self.alarms else "S"
            string = f"{string[:13]} {char}:{time_left:.0f}s"
            string = f"{string:<20}"
        elif self.alarms:
            n = len(self.alarms)
            if n == 1:
                string = string[:14] + " ALARM"
            else:
                string = string[:13] + f" {n}ALRMS"

        return string

    def upper_display(self) -> None:
        if self.extra_in:
            return
        current_name = self.value().lcd_name
        current_number = f"{self._current + 1}"
        if isinstance(self.value(), AdvancedSetting):
            current_number += chr(ord("a") + self.value()._value % 26)
        length_available = 20 - len(current_number) - 2
        if len(current_name) > length_available:
            print(f"Warning: Truncating {current_name!r}")
            current_name = current_name[:length_available]

        string = f"{current_number}: {current_name:<{length_available}}"

        if isinstance(self.value(), AdvancedSetting):
            string = self._add_alarm_text(string)

        self.lcd.upper(string)

    def lower_display(self) -> None:
        if self.extra_in:
            return
        current_item = self.value()
        string = f"{current_item:<20}"
        if len(string) > 20:
            print(f"Warning: Truncating {string!r}")
            string = string[:20]

        if not isinstance(self.value(), AdvancedSetting):
            string = self._add_alarm_text(string)

        self.lcd.lower(string)

    def display(self) -> None:
        with self.lock:
            self.lcd.clear()
            self.lower_display()
            self.upper_display()


if __name__ == "__main__":
    from processor.settings import get_live_settings

    with RotaryLCD(get_live_settings()) as rotary:
        rotary.display()

        while True:
            time.sleep(1)
```
