## Supplementary Git Repository for "Inexpensive multi-patient respiratory monitoring system for helmet ventilation during COVID-19 pandemic": rotary_live.py

```
# mypy: disallow_untyped_defs
# mypy: disallow_incomplete_defs

from __future__ import annotations

from processor.config import config
from processor.setting import Setting
from processor.rotary import LocalRotary
from typing import Optional, Dict, TypeVar, Any
import threading
import yaml
import logging
from pathlib import Path

logger = logging.getLogger("povm")

T = TypeVar("T", bound="LiveRotary")


class LiveRotary(LocalRotary):
    def __init__(self, config: Dict[str, Setting]):
        super().__init__(config)

        self._live_save_stop = threading.Event()
        self._live_save_thread: Optional[threading.Thread] = None
        self._live_save_change = threading.Event()

    def changed(self) -> None:
        self._live_save_change.set()
        super().changed()

    def _live_save(self, filename: Path, every: float) -> None:
        while not self._live_save_stop.is_set():
            if self._live_save_change.is_set():
                d = {
                    "rotary-live": {
                        key: {"default": self.config[key].default}
                        for key in config["rotary-live"]
                    },
                    "rotary": {
                        key: {"default": self.config[key].default}
                        for key in config["rotary"]
                    },
                }

                with open(filename, "w") as f:
                    yaml.dump(d, f)

                self._live_save_change.clear()

            self._live_save_stop.wait(every)

    def live_save(self, filename: Path, every: float) -> None:
        assert self._live_save_thread is None, "Must be called in a with statement"

        self._live_save_thread = threading.Thread(
            target=self._live_save, args=(filename, every)
        )
        self._live_save_thread.start()

    def live_load(self, filename: Path) -> None:
        try:
            with open(filename) as f:
                conf = yaml.load(f, Loader=yaml.SafeLoader)
        except FileNotFoundError:
            logger.info(f"File {filename} does not exist, not restoring defaults")
            return
        except IOError:
            logger.warning(f"Corrupted {filename}")
            return
        except yaml.YAMLError:
            logger.warning(f"Was unable to read YAML {filename}")
            return

        try:
            keys = set(conf["rotary-live"]) | set(conf["rotary"])
        except KeyError:
            logger.warning(f"Malformed keys in {filename}")
            return
        except TypeError:
            logger.warning(f"Empty {filename}")
            return

        if not keys - set(self.config):
            for group in ("rotary-live", "rotary"):
                for key in conf[group]:
                    self[key].default = conf[group][key]["default"]
        else:
            logger.warning(
                f"Was unable to load {filename} due to extra keys: {keys - set(self.config)}"
            )

    def __enter__(self: T) -> T:
        self._live_save_stop.clear()
        return super().__enter__()

    def __exit__(self, *exc: Any) -> None:
        self._live_save_stop.set()
        if self._live_save_thread is not None:
            self._live_save_thread.join()
            self._live_save_thread = None
        super().__exit__(*exc)
```
