## Supplementary Git Repository for "Inexpensive multi-patient respiratory monitoring system for helmet ventilation during COVID-19 pandemic": sdp3test.py

```
#!/usr/bin/env python3

# SDP3 diff pressure sensor - I2C handled by pigpio.pi()
import time
import signal
import pigpio
import binascii

# ------------------
# output file setup
# ------------------
# outputFileName = 'patient.dat'
# f = open(outputFileName,'w')
# sys.stdout = f
# ------------------
# output file end of setup
# ------------------
# ------------------
# SDP3 diff pressure sensor setup
# ------------------
DEVICE_SDP3 = 0x21  # grounded ADDR pin
# Get pigio connection
pi = pigpio.pi()
# Get I2C bus handle
hSDP3 = pi.i2c_open(1, DEVICE_SDP3)
# first issue stop command
pi.i2c_write_device(hSDP3, [0x3F, 0xF9])
# read product number and serial number
print("handle {}".format(hSDP3))
pi.i2c_write_device(hSDP3, [0x36, 0x7C])
pi.i2c_write_device(hSDP3, [0xE1, 0x02])
nbytes = 18
dataSDP3 = pi.i2c_read_device(hSDP3, nbytes)
print(dataSDP3)
bdataSDP3 = dataSDP3[1]
print(binascii.hexlify(bdataSDP3[0:2]))
print(binascii.hexlify(bdataSDP3[3:5]))
pnmsw = int.from_bytes(bdataSDP3[0:2], byteorder="big", signed=False)
pnlsw = int.from_bytes(bdataSDP3[3:5], byteorder="big", signed=False)
pn = (pnmsw << 16) | pnlsw
print(binascii.hexlify(bdataSDP3[6:8]))
print(binascii.hexlify(bdataSDP3[9:11]))
print(binascii.hexlify(bdataSDP3[12:14]))
print(binascii.hexlify(bdataSDP3[15:17]))
snmmsw = int.from_bytes(bdataSDP3[6:8], byteorder="big", signed=False)
snmsw = int.from_bytes(bdataSDP3[9:11], byteorder="big", signed=False)
snlsw = int.from_bytes(bdataSDP3[12:14], byteorder="big", signed=False)
snllsw = int.from_bytes(bdataSDP3[15:17], byteorder="big", signed=False)
sn = (snmmsw << 48) | (snmsw << 32) | (snlsw << 16) | snllsw
print(hex(pn), hex(sn))
# start continuous readout with averaging with differential pressure temperature compensation
pi.i2c_write_device(hSDP3, [0x36, 0x15])
# get initial values of differential pressure, temperature and the differential pressure scale factor
time.sleep(0.020)
nbytes = 9
dataSDP3 = pi.i2c_read_device(hSDP3, nbytes)
print(dataSDP3)
bdataSDP3 = dataSDP3[1]
dp = int.from_bytes(bdataSDP3[0:2], byteorder="big", signed=True)
# print(int.from_bytes(bdataSDP3[3:5], byteorder='big', signed=False))
# print(int.from_bytes(bdataSDP3[6:8], byteorder='big', signed=False))
# dp = (bdataSDP3[0] << 8) | bdataSDP3[1]
temp = (bdataSDP3[3] << 8) | bdataSDP3[4]
dpsf = (float)((bdataSDP3[6] << 8) | bdataSDP3[7])
print(time.time(), dp, temp, dpsf)
print("{} {:.4f} {:.4f}".format(time.time(), (float)(dp / dpsf), (float)(temp / 200.0)))


# sdp3 interrupt handler
def sdp3_handler(signum, frame):
    #  global dpsf
    ts = time.time()
    nbytes = 3
    tmpdataSDP3 = pi.i2c_read_device(hSDP3, nbytes)
    btmpdataSDP3 = tmpdataSDP3[1]
    tmpdp = int.from_bytes(btmpdataSDP3[0:2], byteorder="big", signed=True)
    tmpdict = dict({"t": ts, "F": tmpdp, "P": 0})
    print(tmpdict)


#  tmpdp = ((tmpdataSDP3[0] << 8) + tmpdataSDP3[1])*dpsf
#  print(ts, tmpdp)
signal.signal(signal.SIGALRM, sdp3_handler)
signal.setitimer(signal.ITIMER_REAL, 1, 0.01)  # 10Hz of readout
# ------------------
# SDP3 diff pressure sensor end of setup
# ------------------
while True:
    signal.pause()

# stop continuous measurement
pi.i2c_write_device(hSDP3, [0x3F, 0xF9])
pi.i2c_close(hSDP3)
pi.stop()
```
