## Supplementary Git Repository for "Inexpensive multi-patient respiratory monitoring system for helmet ventilation during COVID-19 pandemic": patient_loop.py

```
#!/usr/bin/env python3

from processor.argparse import ArgumentParser

parser = ArgumentParser(log_dir="patient_log", log_stem="patient_loop")
parser.add_argument("--port", "-p", type=int, default=8100, help="Select a port")
args = parser.parse_args()


import signal
import threading
from contextlib import ExitStack


from processor.settings import get_live_settings
from patient.rotary_lcd import RotaryLCD
from processor.collector import Collector
from processor.broadcast import Broadcast
from processor.config import get_data_dir


# Initialize LCD
with ExitStack() as stack:
    forever = threading.Event()

    def close(_number, _frame):
        print("Closing down server...")
        signal.signal(signal.SIGINT, signal.SIG_DFL)
        signal.signal(signal.SIGTERM, signal.SIG_DFL)
        forever.set()

    rotary = stack.enter_context(RotaryLCD(get_live_settings(), event=forever))
    collector = stack.enter_context(Collector(rotary=rotary, port=args.port))
    stack.enter_context(Broadcast("patient_loop", port=args.port, live=5))

    rotary.live_load(get_data_dir() / "povm-live.yml")
    rotary.live_save(get_data_dir() / "povm-live.yml", every=10)

    signal.signal(signal.SIGINT, close)
    signal.signal(signal.SIGTERM, close)

    forever.wait()
```
