## Supplementary Git Repository for "Inexpensive multi-patient respiratory monitoring system for helmet ventilation during COVID-19 pandemic": patient_sim.py

```
#!/usr/bin/env python3
from __future__ import annotations

from concurrent.futures import ThreadPoolExecutor
import time
import threading
import zmq
from zmq.decorators import context, socket
from typing import Iterator
import signal

from sim.start_sims import start_sims
from processor.argparse import ArgumentParser
from processor.broadcast import Broadcast


def frequency(length: float, event: threading.Event) -> Iterator[None]:
    """
    This pauses as long as needed after running
    """
    while not event.is_set():
        start_time = time.monotonic()
        yield
        diff = time.monotonic() - start_time
        left = length - diff
        if left > 0:
            event.wait(left)


class SimGenerator:
    def __init__(self):
        self.done = threading.Event()

    @context()
    @socket(zmq.PUB)
    def run(self, port: int, _ctx: zmq.Context, pub_socket: zmq.Socket):
        pub_socket.bind(f"tcp://*:{port}")

        with Broadcast("patient_sim", port=port):

            start_time = int(1_000 * time.monotonic())  # milliseconds
            (sim,) = start_sims(1, start_time, 12_000_000)  # milliseconds

            for _ in frequency(1 / 50, self.done):
                d = sim.get_next()
                mod = {
                    "t": int(time.monotonic() * 1000),
                    "f": d["F"],
                    "p": d["P"],
                }

                pub_socket.send_json(mod)


if __name__ == "__main__":
    parser = ArgumentParser(
        description="Serve values on network as JSON",
        log_dir="patient_log",
        log_stem="patient_sim",
    )
    parser.add_argument("--port", type=int, default=8100, help="First port to serve on")
    parser.add_argument("-n", type=int, default=1, help="How many ports to serve on")

    args = parser.parse_args()
    print(args)

    sim_gen = SimGenerator()

    def ctrl_c(_signal, _frame):
        print("You pressed Ctrl+C!")
        signal.signal(signal.SIGINT, signal.SIG_DFL)
        sim_gen.done.set()

    signal.signal(signal.SIGINT, ctrl_c)

    if args.n > 1:
        print("Serving; press Control-C quit")
        addresses = ((args.port + i) for i in range(args.n))
        with ThreadPoolExecutor(max_workers=args.n + 1) as e:
            e.map(sim_gen.run, addresses)
    else:
        sim_gen.run(args.port)
```
