## Supplementary Git Repository for "Inexpensive multi-patient respiratory monitoring system for helmet ventilation during COVID-19 pandemic": patientgui.py

```
#!/usr/bin/env python3

from processor.argparse import ArgumentParser

parser = ArgumentParser(log_dir="patient_log", log_stem="patientgui")
parser.add_argument("--port", "-p", type=int, default=8100, help="Select a port")
args = parser.parse_args()

import os
import signal
import sys

from nurse.qt import QtWidgets
from processor.settings import get_live_settings
from patient.rotary_gui import MainWindow, RotaryGUI
from processor.collector import Collector
from processor.broadcast import Broadcast
from processor.config import get_data_dir

# Hack to allow this to run on non-devices.
if "POVM_MACADDR" not in os.environ:
    os.environ["POVM_MACADDR"] = "00:00:00:00:00:00"


with RotaryGUI(get_live_settings()) as rotary, Collector(
    rotary=rotary, port=args.port
) as collector, Broadcast("patientgui", port=args.port, live=5):

    rotary.live_load(get_data_dir() / "povm-live.yml")
    rotary.live_save(get_data_dir() / "povm-live.yml", every=10)

    app = QtWidgets.QApplication([])
    main = MainWindow(rotary, collector)

    def ctrl_c(_number, _frame):
        main.close()

    signal.signal(signal.SIGINT, ctrl_c)

    main.showNormal()
    sys.exit(app.exec())
```
