## Supplementary Git Repository for "Inexpensive multi-patient respiratory monitoring system for helmet ventilation during COVID-19 pandemic": povm.yml

```
device:
  pressure:
    offset: 12.73
    scale: 0.0126
  flow:
    offset: 0.0
    scale: 0.7198

rotary:
  Avg Flow Min:
    type: Incr
    name: Avg Flow Min
    default: 50
    min: -150
    max: 150
    incr: 1
    unit: L/min

  Avg Flow Max:
    type: Incr
    name: Avg Flow Max
    default: 100
    min: -150
    max: 150
    incr: 1
    unit: L/min

  Avg Pressure Min:
    type: Incr
    name: Avg Pressure Min
    default: 5
    min: -20
    max: 100
    incr: 1
    unit: cm H20

  Avg Pressure Max:
    type: Incr
    name: Avg Pressure Max
    default: 12
    min: -20
    max: 100
    incr: 1
    unit: cm H2O

  RR Max:
    type: Incr
    name: RespRate Max
    default: 40
    min: 0
    max: 180
    incr: 1
    unit: 1/min
    zero: "OFF"

  Avg CO2 Max:
    type: Incr
    name: Avg CO2 Max
    default: 5000
    min: 0
    max: 10000
    incr: 100
    unit: ppm

  Stale Data Timeout:
    type: Incr
    name: Stale Data Timeout
    lcd_name: No breath since
    default: 10
    min: 0
    max: 60
    incr: 1
    unit: sec
    zero: "OFF"
```
