## Supplementary Git Repository for "Inexpensive multi-patient respiratory monitoring system for helmet ventilation during COVID-19 pandemic": analysis.py

```
import numpy as np
import scipy.integrate
import scipy.signal
from typing import Iterable, Dict
import logging
import enum

from processor.rotary import LocalRotary
from processor.config import config


class MaxMin(enum.Enum):
    max = enum.auto()
    min = enum.auto()


def pressure_deglitch_smooth(
    original_pressure,
    deglitch_cut=0.1,
):
    # 1/4 P[i-2] + 1/4 P[i-1] + 0 P[i] + 1/4 P[i+1] + 1/4 P[i+2] kernel
    pressure_average = 0.25 * (
        original_pressure[4:]
        + original_pressure[3:-1]
        + original_pressure[1:-3]
        + original_pressure[:-4]
    )

    # deglitching: large exclusions from the average of neighbors is
    #              replaced with an average of neighbors
    toreplace22 = abs(pressure_average - original_pressure[2:-2]) > deglitch_cut
    toreplace = np.zeros(len(original_pressure), np.bool_)
    toreplace[2:-2] = toreplace22

    pressure_out = original_pressure.copy()
    pressure_out[toreplace] = pressure_average[toreplace22]

    # smoothing: 2/5 P[i-2] + 1/5 P[i] + 2/5 P[i+2] kernel
    pressure_out[1:-1] = (
        0.4 * pressure_out[2:] + 0.2 * pressure_out[1:-1] + 0.4 * pressure_out[:-2]
    )
    return pressure_out


def _compute_cumulative_length(
    length: int, time: np.ndarray, window: np.ndarray
) -> float:
    if len(time) == 0:
        return 0.0
    try:
        ind = np.searchsorted(time, time[-1] - length * 1000)
    except ValueError:
        print("Time:", time)
        print("Value:", time[-1] - length * 1000)
        raise
    return np.mean(window[ind:])


def compute_cumulative(
    lengths: Iterable[int], time: np.ndarray, window: np.ndarray
) -> Dict[int, float]:

    return {
        length: _compute_cumulative_length(length, time, window) for length in lengths
    }


def flow_to_volume(realtime, old_realtime, flow, old_volume, critical_frequency):
    if old_realtime is None:
        shift = 0
    else:
        shift = old_volume[np.argmin(abs(old_realtime - realtime[0]))]

    out = scipy.integrate.cumtrapz(flow * 1000, realtime / 60.0, initial=0) + shift

    return scipy.signal.sosfilt(
        scipy.signal.butter(1, critical_frequency, "highpass", output="sos"), out
    )


class CantComputeDerivative(Exception):
    pass


def smooth_derivative(times, values, sig=0.2):
    values = values - values.mean()

    if len(times) < 1:
        raise CantComputeDerivative

    themin = np.min(times[1:] - times[:-1])
    if themin <= 0:
        raise CantComputeDerivative

    window_width = int(np.ceil(4 * sig / themin))
    if len(times) - window_width + 1 < 10 or window_width < 10:
        raise CantComputeDerivative

    windowed_times = np.lib.stride_tricks.as_strided(
        times,
        (len(times) - window_width + 1, window_width),
        (times.itemsize, times.itemsize),
    )
    windowed_values = np.lib.stride_tricks.as_strided(
        values,
        (len(values) - window_width + 1, window_width),
        (values.itemsize, values.itemsize),
    )

    centers = np.mean(windowed_times, axis=1)
    windowed_times_centered = windowed_times - centers[:, np.newaxis]
    windowed_weights = np.exp(-0.5 * windowed_times_centered ** 2 / sig ** 2)
    sumw = np.sum(windowed_weights, axis=1)
    sumwx = np.sum(windowed_weights * windowed_times_centered, axis=1)
    sumwy = np.sum(windowed_weights * windowed_values, axis=1)
    sumwxx = np.sum(
        windowed_weights * windowed_times_centered * windowed_times_centered, axis=1
    )
    sumwxy = np.sum(
        windowed_weights * windowed_times_centered * windowed_values, axis=1
    )
    delta = (sumw * sumwxx) - (sumwx * sumwx)

    with np.errstate(all="ignore"):
        intercept = ((sumwxx * sumwy) - (sumwx * sumwxy)) / delta
        slope = ((sumw * sumwxy) - (sumwx * sumwy)) / delta

    good = (~np.isnan(intercept)) & (~np.isnan(slope))

    return centers[good], intercept[good], slope[good]


def find_roots(
    times, values, derivative, threshold: float = 0.02, *, breath_thresh: float
):
    values_threshold = threshold * max(np.max(abs(values)), breath_thresh)
    derivative_threshold = threshold * np.max(abs(derivative))

    (A,) = np.nonzero(
        (values[:-1] < 0)
        & (values[1:] >= 0)
        & (0.5 * (derivative[:-1] + derivative[1:]) >= derivative_threshold)
    )
    (B,) = np.nonzero(
        (derivative[:-1] >= 0)
        & (derivative[1:] < 0)
        & (0.5 * (values[:-1] + values[1:]) >= values_threshold)
    )
    (C,) = np.nonzero(
        (values[:-1] >= -0.5 * values_threshold)
        & (values[1:] < -0.5 * values_threshold)
        & (0.5 * (derivative[:-1] + derivative[1:]) < -derivative_threshold)
    )
    (D,) = np.nonzero(
        (derivative[:-1] < 0)
        & (derivative[1:] >= 0)
        & (0.5 * (values[:-1] + values[1:]) < -values_threshold)
    )

    return (
        0.5 * (times[A] + times[A + 1]),
        0.5 * (times[B] + times[B + 1]),
        0.5 * (times[C] + times[C + 1]),
        0.5 * (times[D] + times[D + 1]),
    )


def find_breaths(A, B, C, D):
    # ensure that each type is sorted (though it probably already is)
    A.sort()
    B.sort()
    C.sort()
    D.sort()

    if len(A) == 0 or len(B) == 0 or len(C) == 0 or len(D) == 0:
        return []

    # where does the cycle start?
    which = np.argmin([A[0], B[0], C[0], D[0]])

    ins = [A, B, C, D]
    outs = []
    while len(A) > 0 or len(B) > 0 or len(C) > 0 or len(D) > 0:
        if len(ins[which]) > 1 and all(len(ins[i]) > 0 for i in range(4)):
            nextwhich = np.argmin([ins[i][1 if i == which else 0] for i in range(4)])

            # normal case: A -> B -> C -> D -> ... cycle
            if nextwhich == (which + 1) % 4:
                outs.append((which, ins[which][0]))
                ins[which] = ins[which][1:]

            # too many of one type: A -> B -> B -> B -> C -> D -> ...
            elif nextwhich == which:
                combine = [ins[which][0]]
                ins[which] = ins[which][1:]
                while True:
                    if (
                        any(len(x) == 0 for x in ins)
                        or np.argmin([ins[i][0] for i in range(4)]) != which
                    ):
                        break
                    combine.append(ins[which][0])
                    ins[which] = ins[which][1:]

                outs.append((which, np.mean(combine)))

            # missing one type: A -> C -> D -> ...
            else:
                fill_value = 0.5 * (ins[which][0] + ins[nextwhich][0])
                ins[(which + 1) % 4] = np.concatenate(
                    (np.array([fill_value]), ins[(which + 1) % 4])
                )

                outs.append((which, ins[which][0]))
                ins[which] = ins[which][1:]

        # edge condition: less than one cycle left
        elif len(ins[which]) > 0:
            popped = ins[which][0]
            ins[which] = ins[which][1:]

            if len(outs) > 0 and (outs[-1][0] + 1) % 4 == which:
                outs.append((which, popped))

        else:
            break

        which = (which + 1) % 4
        A, B, C, D = ins

    return outs


def measure_breaths(time, flow, volume, pressure, *, breath_thresh: float):
    try:
        smooth_time_f, smooth_flow, smooth_dflow = smooth_derivative(time, flow)
        smooth_time_p, smooth_pressure, smooth_dpressure = smooth_derivative(
            time, pressure
        )
        if len(smooth_time_f) < 4:
            return []

        turning_points = find_roots(
            smooth_time_f, smooth_flow, smooth_dflow, breath_thresh=breath_thresh
        )

        breath_times = find_breaths(*turning_points)

    except Exception as err:
        if not isinstance(err, CantComputeDerivative) and config["global"]["debug"].get(
            bool
        ):
            raise
        return []

    if len(time) == 0 or len(breath_times) == 0:
        return []

    breaths = []
    breath = {}
    for i, (which, t) in enumerate(breath_times):
        index = np.argmin(abs(time - t))

        if which == 0:
            breath["empty timestamp"] = t
            if 0 <= index < len(pressure):
                breath["empty pressure"] = pressure[index]
            if 0 <= index < len(volume):
                breath["empty volume"] = volume[index]
            if i >= 2:
                full_index = np.argmin(abs(time - breath_times[i - 2][1]))
                if 0 <= full_index < len(volume):
                    diff = volume[full_index] - breath["empty volume"]
                    if diff > 0:
                        breath["inspiratory tidal volume"] = diff
            if i >= 4:
                last_index = np.argmin(abs(time - breath_times[i - 4][1]))
                if 0 <= last_index < len(flow) and last_index < index:
                    breath["average flow"] = flow[last_index:index].sum() / (
                        index - last_index
                    )
                if 0 <= last_index < len(pressure) and last_index < index:
                    breath["average pressure"] = pressure[last_index:index].sum() / (
                        index - last_index
                    )

            breaths.append(breath)
            breath = {}

        elif which == 1:
            breath["inhale timestamp"] = t
            if 0 <= index < len(flow):
                breath["inhale flow"] = flow[index]
            if len(smooth_time_f) > 0:
                idx = np.argmin(abs(smooth_time_f - t))
                if 0 <= idx < len(smooth_flow):
                    breath["inhale dV/dt"] = smooth_flow[idx] * 1000 / 60.0
            if len(smooth_time_p) > 0:
                idx = np.argmin(abs(smooth_time_p - t))
                if 0 <= idx < len(smooth_dpressure):
                    breath["inhale dP/dt"] = smooth_dpressure[idx]
            if "inhale dV/dt" in breath and "inhale dP/dt" in breath:
                breath["inhale compliance"] = (
                    breath["inhale dV/dt"] / breath["inhale dP/dt"]
                )
            if i >= 2:
                start_index = np.argmin(abs(time - breath_times[i - 2][1]))
                if start_index < index:
                    breath["min pressure"] = np.min(pressure[start_index:index])

        elif which == 2:
            breath["full timestamp"] = t
            if 0 <= index < len(pressure):
                breath["full pressure"] = pressure[index]
            if 0 <= index < len(volume):
                breath["full volume"] = volume[index]
            if i >= 2:
                empty_index = np.argmin(abs(time - breath_times[i - 2][1]))
                if 0 <= empty_index < len(volume):
                    diff = breath["full volume"] - volume[empty_index]
                    if diff > 0:
                        breath["expiratory tidal volume"] = diff

        elif which == 3:
            breath["exhale timestamp"] = t
            if 0 <= index < len(flow):
                breath["exhale flow"] = flow[index]
            if len(smooth_time_f) > 0:
                idx = np.argmin(abs(smooth_time_f - t))
                if 0 <= idx < len(smooth_flow):
                    breath["exhale dV/dt"] = smooth_flow[idx] * 1000 / 60.0
            if len(smooth_time_p) > 0:
                idx = np.argmin(abs(smooth_time_p - t))
                if 0 <= idx < len(smooth_dpressure):
                    breath["exhale dP/dt"] = smooth_dpressure[idx]
            if "exhale dV/dt" in breath and "exhale dP/dt" in breath:
                breath["exhale compliance"] = (
                    breath["exhale dV/dt"] / breath["exhale dP/dt"]
                )
            if i >= 2:
                start_index = np.argmin(abs(time - breath_times[i - 2][1]))
                if start_index < index:
                    breath["max pressure"] = np.max(pressure[start_index:index])

    if (
        "empty timestamp" in breath
        or "inhale timestamp" in breath
        or "full timestamp" in breath
        or "exhale timestamp" in breath
    ):
        breaths.append(breath)

    return breaths


def combine_breaths(old_breaths, new_breaths):
    breaths = list(old_breaths)
    new_breaths = list(new_breaths)
    updated = []

    first_to_check = max(0, len(breaths) - len(new_breaths) - 1)
    for i in range(first_to_check, len(breaths)):
        drop = []
        for j in range(len(new_breaths)):
            # the smoothing sigma is 0.2 sec (see smooth_derivative), so cut at 3*0.2
            same = False
            if "empty timestamp" in breaths[i] and "empty timestamp" in new_breaths[j]:
                same = (
                    same
                    or abs(
                        breaths[i]["empty timestamp"]
                        - new_breaths[j]["empty timestamp"]
                    )
                    < 3 * 0.2
                )
            if (
                "inhale timestamp" in breaths[i]
                and "inhale timestamp" in new_breaths[j]
            ):
                same = (
                    same
                    or abs(
                        breaths[i]["inhale timestamp"]
                        - new_breaths[j]["inhale timestamp"]
                    )
                    < 3 * 0.2
                )
            if "full timestamp" in breaths[i] and "full timestamp" in new_breaths[j]:
                same = (
                    same
                    or abs(
                        breaths[i]["full timestamp"] - new_breaths[j]["full timestamp"]
                    )
                    < 3 * 0.2
                )
            if (
                "exhale timestamp" in breaths[i]
                and "exhale timestamp" in new_breaths[j]
            ):
                same = (
                    same
                    or abs(
                        breaths[i]["exhale timestamp"]
                        - new_breaths[j]["exhale timestamp"]
                    )
                    < 3 * 0.2
                )

            if same:
                # take all fields that are defined in either old or new, but preferring new if it's in both
                breaths[i] = {**breaths[i], **new_breaths[j]}
                updated.append(breaths[i])
                drop.append(j)
                break

        for j in drop[::-1]:
            del new_breaths[j]

    breaths.extend(new_breaths)
    breaths.sort(key=average_any_times)

    # The dicts in breaths, updated, and new_breaths are the same objects,
    # so updating the superset (breaths) affects them all.

    # Adding time-difference quantities.
    for i, breath in enumerate(breaths):
        if (
            "empty timestamp" in breath
            and "full timestamp" in breath
            and breath["empty timestamp"] > breath["full timestamp"]
        ):
            breath["exhale time"] = breath["empty timestamp"] - breath["full timestamp"]
        elif (
            i > 0
            and "empty timestamp" in breath
            and "full timestamp" in breaths[i - 1]
            and breath["empty timestamp"] > breaths[i - 1]["full timestamp"]
        ):
            breath["exhale time"] = (
                breath["empty timestamp"] - breaths[i - 1]["full timestamp"]
            )

        if (
            "full timestamp" in breath
            and "empty timestamp" in breath
            and breath["full timestamp"] > breath["empty timestamp"]
        ):
            breath["inhale time"] = breath["full timestamp"] - breath["empty timestamp"]
        elif (
            i > 0
            and "full timestamp" in breath
            and "empty timestamp" in breaths[i - 1]
            and breath["full timestamp"] > breaths[i - 1]["empty timestamp"]
        ):
            breath["inhale time"] = (
                breath["full timestamp"] - breaths[i - 1]["empty timestamp"]
            )

        if (
            i > 0
            and "empty timestamp" in breath
            and "empty timestamp" in breaths[i - 1]
            and breath["empty timestamp"] > breaths[i - 1]["empty timestamp"]
        ):
            breath["time since last"] = (
                breath["empty timestamp"] - breaths[i - 1]["empty timestamp"]
            )
        elif (
            i > 0
            and "full timestamp" in breath
            and "full timestamp" in breaths[i - 1]
            and breath["full timestamp"] > breaths[i - 1]["full timestamp"]
        ):
            breath["time since last"] = (
                breath["full timestamp"] - breaths[i - 1]["full timestamp"]
            )
        elif (
            i > 0
            and "inhale timestamp" in breath
            and "inhale timestamp" in breaths[i - 1]
            and breath["inhale timestamp"] > breaths[i - 1]["inhale timestamp"]
        ):
            breath["time since last"] = (
                breath["inhale timestamp"] - breaths[i - 1]["inhale timestamp"]
            )
        elif (
            i > 0
            and "exhale timestamp" in breath
            and "exhale timestamp" in breaths[i - 1]
            and breath["exhale timestamp"] > breaths[i - 1]["exhale timestamp"]
        ):
            breath["time since last"] = (
                breath["exhale timestamp"] - breaths[i - 1]["exhale timestamp"]
            )

    return breaths, updated, new_breaths


def average_any_times(breath):
    times = []
    if "empty timestamp" in breath:
        times.append(breath["empty timestamp"])
    if "inhale timestamp" in breath:
        times.append(breath["inhale timestamp"])
    if "full timestamp" in breath:
        times.append(breath["full timestamp"])
    if "exhale timestamp" in breath:
        times.append(breath["exhale timestamp"])
    return sum(times) / len(times)  # every breath has at least one


# default alpha is 0.3: value changes after about 3 breaths
def moving_average(cumulative, updated_fields, key, value, alpha=0.3):
    updated_fields.add(key)
    if key not in cumulative:
        return value
    else:
        return alpha * value + (1.0 - alpha) * cumulative[key]


def cumulative(cumulative, updated, new_breaths):
    cumulative = dict(cumulative)
    updated_fields = set()

    for breath in updated + new_breaths:
        timestamp = None
        if "empty timestamp" in breath and (
            timestamp is None or timestamp < breath["empty timestamp"]
        ):
            timestamp = breath["empty timestamp"]
        if "inhale timestamp" in breath and (
            timestamp is None or timestamp < breath["inhale timestamp"]
        ):
            timestamp = breath["inhale timestamp"]
        if "full timestamp" in breath and (
            timestamp is None or timestamp < breath["full timestamp"]
        ):
            timestamp = breath["full timestamp"]
        if "exhale timestamp" in breath and (
            timestamp is None or timestamp < breath["exhale timestamp"]
        ):
            timestamp = breath["exhale timestamp"]

        this_is_new = False
        if timestamp is not None:
            # the smoothing sigma is 0.2 sec (see smooth_derivative), so cut at 3*0.2
            if (
                "last breath timestamp" not in cumulative
                or cumulative["last breath timestamp"] + 3 * 0.2 < timestamp
            ):
                cumulative["last breath timestamp"] = timestamp
                updated_fields.add("last breath timestamp")
                this_is_new = True

        if this_is_new:
            if "time since last" in breath:
                cumulative["breath interval"] = moving_average(
                    cumulative,
                    updated_fields,
                    "breath interval",
                    breath["time since last"],
                )
                cumulative["RR"] = 60.0 / cumulative["breath interval"]
                updated_fields.add("RR")
            if "max pressure" in breath:
                cumulative["PIP"] = moving_average(
                    cumulative, updated_fields, "PIP", breath["max pressure"]
                )
            if "empty pressure" in breath:
                cumulative["PEEP"] = moving_average(
                    cumulative, updated_fields, "PEEP", breath["empty pressure"]
                )
            if "expiratory tidal volume" in breath:
                cumulative["TVe"] = moving_average(
                    cumulative, updated_fields, "TVe", breath["expiratory tidal volume"]
                )
            if "inspiratory tidal volume" in breath:
                cumulative["TVi"] = moving_average(
                    cumulative,
                    updated_fields,
                    "TVi",
                    breath["inspiratory tidal volume"],
                )
            if "TVe" in cumulative and "TVi" in cumulative:
                cumulative["TV"] = 0.5 * (cumulative["TVe"] + cumulative["TVi"])
                updated_fields.add("TV")
            if "inhale compliance" in breath:
                cumulative["inhale compliance"] = moving_average(
                    cumulative,
                    updated_fields,
                    "inhale compliance",
                    breath["inhale compliance"],
                )
            if "exhale compliance" in breath:
                cumulative["exhale compliance"] = moving_average(
                    cumulative,
                    updated_fields,
                    "exhale compliance",
                    breath["exhale compliance"],
                )
            if "RR" in cumulative and "TV" in cumulative:
                cumulative["breath volume rate"] = (
                    cumulative["TV"] * cumulative["RR"] / 1000.0
                )
                updated_fields.add("breath volume rate")
            if "inhale time" in breath:
                cumulative["inhale time"] = moving_average(
                    cumulative,
                    updated_fields,
                    "inhale time",
                    breath["inhale time"],
                )
            if "exhale time" in breath:
                cumulative["exhale time"] = moving_average(
                    cumulative,
                    updated_fields,
                    "exhale time",
                    breath["exhale time"],
                )
            if "inhale time" in cumulative and "exhale time" in cumulative:
                cumulative["I:E time ratio"] = (
                    cumulative["inhale time"] / cumulative["exhale time"]
                )
                updated_fields.add("I:E time ratio")
            if "average flow" in breath:
                cumulative["breath average flow"] = moving_average(
                    cumulative,
                    updated_fields,
                    "breath average flow",
                    breath["average flow"],
                )
            if "average pressure" in breath:
                cumulative["breath average pressure"] = moving_average(
                    cumulative,
                    updated_fields,
                    "breath average pressure",
                    breath["average pressure"],
                )

    return cumulative, updated_fields


def alarm_record(old_record, timestamp, value, ismax):
    if old_record is None:
        return {
            "first timestamp": timestamp,
            "last timestamp": timestamp,
            "extreme": value,
        }

    else:
        record = dict(old_record)
        record["last timestamp"] = timestamp
        if ismax and value > record["extreme"]:
            record["extreme"] = value
        elif not ismax and value < record["extreme"]:
            record["extreme"] = value
        return record


def sync_alarm(
    old_alarms: Dict[str, Dict[str, float]],
    logger: logging.Logger,
    key: str,
    timestamp: float,
    value: float,
    limit: float,
    direction: MaxMin,
) -> None:

    condition = (value < limit) if direction == MaxMin.min else (value > limit)

    if condition:
        old_alarms[key] = alarm_record(old_alarms.get(key), timestamp, value, True)
        if old_alarms[key]["first timestamp"] == timestamp:
            logger.info(
                f"Alarm {key!r} activated with value {old_alarms[key]['extreme']}"
            )
    elif key in old_alarms:
        time_active = timestamp - old_alarms[key]["first timestamp"]
        logger.info(
            f"Alarm {key!r} deactivated after being on for {time_active:g} seconds"
        )
        del old_alarms[key]


def avg_alarms(
    old_alarms: Dict[str, Dict[str, float]],
    rotary: LocalRotary,
    key: str,
    values: Dict[int, float],
    timestamp: float,
    logger: logging.Logger,
):
    """
    Return a dict with an alarm if alarm present and out of bounds.
    """

    avg_window = config["global"]["avg-window"].get(int)
    max_key = f"Avg {key.capitalize()} Max"
    min_key = f"Avg {key.capitalize()} Min"

    if max_key in rotary:
        sync_alarm(
            old_alarms,
            logger,
            max_key,
            timestamp,
            values[avg_window],
            rotary[max_key].value,
            MaxMin.max,
        )

    if min_key in rotary:
        sync_alarm(
            old_alarms,
            logger,
            min_key,
            timestamp,
            values[avg_window],
            rotary[min_key].value,
            MaxMin.min,
        )


def co2_alarm(
    old_alarms: Dict[str, Dict[str, float]],
    rotary: LocalRotary,
    values: np.ndarray,
    timestamp: float,
    logger: logging.Logger,
):
    """
    Return a dict with an alarm if alarm present and out of bounds.
    """

    max_key = "Avg CO2 Max"
    min_key = "Avg CO2 Min"

    if max_key in rotary:
        avg = float(np.mean(values))

        if max_key in rotary:
            sync_alarm(
                old_alarms,
                logger,
                max_key,
                timestamp,
                avg,
                rotary[max_key].value,
                MaxMin.max,
            )

        if min_key in rotary:
            sync_alarm(
                old_alarms,
                logger,
                min_key,
                timestamp,
                avg,
                rotary[min_key].value,
                MaxMin.min,
            )

    return old_alarms


def add_alarms(rotary, _updated, _new_breaths, cumulative, old_alarms, logger):
    alarms = {}

    if "PIP" in cumulative:
        if (
            "RR Max" in rotary
            and rotary["RR Max"].value > 0
            and "RR" in cumulative
            and cumulative["RR"] > rotary["RR Max"].value
        ):
            alarms["RR Max"] = alarm_record(
                old_alarms.get("RR Max"),
                cumulative["last breath timestamp"],
                cumulative["RR"],
                True,
            )

        if (
            "PIP Max" in rotary
            and "PIP" in cumulative
            and cumulative["PIP"] > rotary["PIP Max"].value
        ):
            alarms["PIP Max"] = alarm_record(
                old_alarms.get("PIP Max"),
                cumulative["last breath timestamp"],
                cumulative["PIP"],
                True,
            )

        if (
            "PIP Min" in rotary
            and "PIP" in cumulative
            and cumulative["PIP"] < rotary["PIP Min"].value
        ):
            alarms["PIP Min"] = alarm_record(
                old_alarms.get("PIP Min"),
                cumulative["last breath timestamp"],
                cumulative["PIP"],
                False,
            )

        if (
            "PEEP Max" in rotary
            and "PEEP" in cumulative
            and cumulative["PEEP"] > rotary["PEEP Max"].value
        ):
            alarms["PEEP Max"] = alarm_record(
                old_alarms.get("PEEP Max"),
                cumulative["last breath timestamp"],
                cumulative["PEEP"],
                True,
            )

        if (
            "PEEP Min" in rotary
            and "PEEP" in cumulative
            and cumulative["PEEP"] < rotary["PEEP Min"].value
        ):
            alarms["PEEP Min"] = alarm_record(
                old_alarms.get("PEEP Min"),
                cumulative["last breath timestamp"],
                cumulative["PEEP"],
                False,
            )

        if (
            "TVe Max" in rotary
            and "TVe" in cumulative
            and cumulative["TVe"] > rotary["TVe Max"].value
        ):
            alarms["TVe Max"] = alarm_record(
                old_alarms.get("TVe Max"),
                cumulative["last breath timestamp"],
                cumulative["TVe"],
                True,
            )

        if (
            "TVe Min" in rotary
            and "TVe" in cumulative
            and cumulative["TVe"] < rotary["TVe Min"].value
        ):
            alarms["TVe Min"] = alarm_record(
                old_alarms.get("TVe Min"),
                cumulative["last breath timestamp"],
                cumulative["TVe"],
                False,
            )

        if (
            "TVi Max" in rotary
            and "TVi" in cumulative
            and cumulative["TVi"] > rotary["TVi Max"].value
        ):
            alarms["TVi Max"] = alarm_record(
                old_alarms.get("TVi Max"),
                cumulative["last breath timestamp"],
                cumulative["TVi"],
                True,
            )

        if (
            "TVi Min" in rotary
            and "TVi" in cumulative
            and cumulative["TVi"] < rotary["TVi Min"].value
        ):
            alarms["TVi Min"] = alarm_record(
                old_alarms.get("TVi Min"),
                cumulative["last breath timestamp"],
                cumulative["TVi"],
                False,
            )

        for name in alarms:
            if name != "Stale Data" and name not in old_alarms:
                logger.info(
                    f"Alarm {name!r} activated with value {alarms[name]['extreme']}"
                )

        for name in old_alarms:
            if name != "Stale Data" and name not in alarms:
                try:
                    time_active = (
                        cumulative["last breath timestamp"]
                        - old_alarms[name]["first timestamp"]
                    )
                    logger.info(
                        f"Alarm {name!r} deactivated after being on for {time_active:g} seconds"
                    )
                except KeyError:
                    logger.error(f"Unable to find key in {name!r}")

    return alarms
```
