## Supplementary Git Repository for "Inexpensive multi-patient respiratory monitoring system for helmet ventilation during COVID-19 pandemic": argparse.py

```
from __future__ import annotations

import time
import argparse
import logging
import sys
from typing import Tuple, List, Optional

from processor.config import config, get_internal_file
from processor.logging import init_logger
from processor.broadcast import get_ip

logger = logging.getLogger("povm")


def excepthook(exctype, value, traceback):
    logger.error("uncaught exception", exc_info=(exctype, value, traceback))
    sys.__excepthook__(exctype, value, traceback)


class ArgumentParser(argparse.ArgumentParser):
    def __init__(self, *args, log_dir=Optional[str], log_stem=Optional[str], **kwargs):
        super().__init__(  # type: ignore
            *args, formatter_class=argparse.ArgumentDefaultsHelpFormatter, **kwargs
        )

        self.add_argument(
            "--config",
            default=str(get_internal_file("povm.yml")),
            help="YAML configuration file",
        )

        self.add_argument(
            "--debug",
            action="store_true",
            help="Start up in debug mode (log to screen)",
        )

        self.add_argument(
            "--iface",
            help="Wait for 1 IP address to be assigned on this interface before starting",
        )

        self.add_argument("--dir", help="Set a directory to log data to")

        self.log_dir = log_dir
        self.log_stem = log_stem

    def parse_known_args(
        self, *pargs, **kwargs
    ) -> Tuple[argparse.Namespace, List[str]]:
        # Note: This gets called by parse_args, so we don't need
        # to override both

        args, unparsed_args = super().parse_known_args(*pargs, **kwargs)

        if args.config:
            config.set_file(args.config)

        if "dir" in args:
            config.set_args({"global": {"datadir": args.dir}})

        if "debug" in args:
            config.set_args({"global": {"debug": args.debug}})

        if not config["global"]["debug"].get(bool):
            sys.excepthook = excepthook

        if self.log_dir is not None and self.log_dir is not None:
            init_logger(f"{self.log_dir}/{self.log_stem}.log")
        else:
            init_logger(None)

        logger = logging.getLogger("povm")

        if args.iface is not None:
            iface = args.iface
            logger.info(f"Checking (and waiting, if needed) for {iface}")
            faces = get_ip(iface)
            while not list(faces):
                faces = get_ip(iface)
                logger.info("Didn't find iface, waiting 1s")
                time.sleep(1)

        return args, unparsed_args
```
