## Supplementary Git Repository for "Inexpensive multi-patient respiratory monitoring system for helmet ventilation during COVID-19 pandemic": broadcast.py

```
#!/usr/bin/env python3
from __future__ import annotations

from zeroconf import ServiceInfo, Zeroconf
from ifaddr import get_adapters
from patient.mac_address import get_mac_addr, get_box_name
from typing import Iterator, Set, Optional
import threading
import ipaddress
import logging

logger = logging.getLogger("povm")


def get_ip(items: Optional[Iterator[str]] = None) -> Iterator[str]:
    if items is None:
        ifaces = (
            iface
            for iface in get_adapters()
            if iface.ips and iface.name not in ["lo", "lo0"]
        )
    else:
        ifaces = (
            iface for iface in get_adapters() if iface.ips and iface.name in items
        )

    for iface in ifaces:
        yield from (addr.ip for addr in iface.ips if addr.is_IPv4)


class Broadcast:
    def __init__(self, service: str, port: int = 8100, *, live: int = 0) -> None:
        """
        Service should be the name of the service you want to promote (nurse, sim, etc)
        Set a timeout for live to have this poll for new IP address assignments at this rate.
        """

        self.zeroconf = Zeroconf()
        self.port = port
        self.addrs: Set[str] = set()
        self.service = service
        self.live = live
        self.stop = threading.Event()
        self.thread: Optional[threading.Thread] = None
        self.info: Optional[ServiceInfo] = None

        # Workaround for this not always coming online when you start from a service
        self.times = 0

    def register(self):
        self.times += 1
        addrs = set(get_ip())
        if addrs != self.addrs or self.times in [1, 5, 25]:
            for addr in self.addrs - addrs:
                logger.info(f"Ending broadcast on {addr}")
            logger.info(f"Starting broadcast on {', '.join(addrs)}")

            if self.info is not None:
                self.zeroconf.unregister_service(self.info)

            self.zeroconf.close()
            self.zeroconf = Zeroconf()

            self.info = ServiceInfo(
                "_http._tcp.local.",
                f"Princeton Open Vent Monitor - {get_box_name()} - {self.port}._http._tcp.local.",
                addresses=[ipaddress.ip_address(ip).packed for ip in addrs],
                port=self.port,
                properties={
                    "type": "povm",
                    "mac_addr": get_mac_addr(),
                    "service": self.service,
                    "name": get_box_name(),
                    "v": "1",
                },
            )
            self.zeroconf.register_service(self.info)
            self.addrs = addrs

    def _run(self):
        try:
            while not self.stop.is_set():
                self.register()
                self.stop.wait(self.live)
            if self.info is not None:
                self.zeroconf.unregister_service(self.info)
        except Exception:
            logging.exception("Broadcast loop error!")
            raise

    def __enter__(self) -> Broadcast:
        if self.live > 0:
            self.thread = threading.Thread(target=self._run)
            self.thread.start()
        else:
            self.register()
        return self

    def __exit__(self, *exc) -> None:
        self.stop.set()
        if self.thread is not None:
            self.thread.join()
        elif self.info is not None:
            self.zeroconf.unregister_service(self.info)

        self.zeroconf.close()


if __name__ == "__main__":
    with Broadcast("broadcast"):
        forever = threading.Event()
        forever.wait()
```
