## Supplementary Git Repository for "Inexpensive multi-patient respiratory monitoring system for helmet ventilation during COVID-19 pandemic": collector.py

```
#!/usr/bin/env python3
from __future__ import annotations

from datetime import datetime
from typing import Optional
import time

from zmq.decorators import context, socket
import numpy as np
import zmq

from processor.config import config
from processor.display_settings import CurrentSetting, CO2Setting
from processor.generator import Generator
from processor.rotary import LocalRotary
from processor.thread_base import ThreadBase

from patient.mac_address import get_mac_addr, get_box_name


class CollectorThread(ThreadBase):
    def __init__(self, parent: Collector):
        self.parent = parent

        self._sn: Optional[int] = None
        self._file: Optional[str] = None

        self._flow_scale = config["device"]["flow"]["scale"].as_number()
        self._flow_offset = config["device"]["flow"]["offset"].as_number()
        self._pressure_scale = config["device"]["pressure"]["scale"].as_number()
        self._pressure_offset = config["device"]["pressure"]["offset"].as_number()

        super().__init__(parent)

    @context()
    @socket(zmq.SUB)
    @socket(zmq.PUB)
    def run(
        self, _ctx: zmq.Context, sub_socket: zmq.Socket, pub_socket: zmq.Socket
    ) -> None:
        sub_socket.connect("tcp://localhost:5556")
        sub_socket.subscribe(b"")

        # flow calibration
        from processor.flow_calibrator import flow_calibrator

        caliber = flow_calibrator()

        # Up to 60 seconds of data (roughly, not promised)
        pub_socket.hwm = 3000

        pub_socket.bind(f"tcp://*:{self.parent.port}")

        last = time.monotonic()
        while not self.parent.stop.is_set():
            ready_events = sub_socket.poll(0.1)
            for _ in range(ready_events):
                j = sub_socket.recv_json()
                t = j["t"]

                # Disconnected sensor block will send 0's
                if "F" not in j or "P" not in j:
                    f: float = 0
                    p: float = 0
                else:
                    f = caliber.Q(j["F"])
                    p = j["P"] * self._pressure_scale - self._pressure_offset

                pub_socket.send_json({"t": t, "f": f, "p": p})

                if "sn" in j:
                    self._sn = j["sn"]

                if "file" in j:
                    self._file = j["file"]

                with self.lock:
                    self._time.inject_value(t)
                    self._flow.inject_value(f)
                    self._pressure.inject_value(p)

                    extras = {}
                    if "C" in j:
                        extras["t"] = t
                        extras["C"] = j["C"]
                        extras["D"] = j["D"]
                        self._heat_time.inject_value(t)
                        self._heat_temp.inject_value(j["C"])
                        self._heat_duty.inject_value(j["D"])
                    if "CO2" in j:
                        extras["t"] = t
                        extras["CO2"] = j["CO2"]
                        extras["Tp"] = j["Tp"]
                        extras["H"] = j["H"]
                        self._co2_time.inject_value(t)
                        self._co2.inject_value(j["CO2"])
                        self._co2_temp.inject_value(j["Tp"])
                        self._humidity.inject_value(j["H"])

                # This runs every ~1 second, since it is only about that frequent from the device
                if extras:
                    pub_socket.send_json(extras)

            # Send rotary every ~1 second, regardless of status of input
            if time.monotonic() > (last + 1) or self.parent.rotary._changed.is_set():
                with self.parent.lock:
                    extra_dict = {
                        "rotary": self.parent.rotary.to_dict(),
                        "date": datetime.now().timestamp(),
                        "mac": get_mac_addr(),
                        "name": get_box_name(),
                        "last interact": self.parent.rotary.last_interaction(),
                        "time left": self.parent.rotary.time_left(),
                        "monotime": time.monotonic(),
                    }

                    if self._sn is not None:
                        extra_dict["sid"] = self._sn
                        if "Advanced" in self.parent.rotary:
                            setting = self.parent.rotary["Advanced"]
                            setting.sid = self._sn

                    if self._file is not None:
                        extra_dict["file"] = self._file
                        if "Advanced" in self.parent.rotary:
                            setting = self.parent.rotary["Advanced"]
                            setting.file = self._file

                    pub_socket.send_json(extra_dict)

                last = time.monotonic()
                self.parent.rotary._changed.clear()

    def access_collected_data(self) -> None:
        with self.parent.lock, self.lock:
            newel = self.parent._time.new_elements(self._time)
            self.parent._time.inject_batch(self._time, newel)
            self.parent._flow.inject_batch(self._flow, newel)
            self.parent._pressure.inject_batch(self._pressure, newel)

            newel = self.parent._heat_time.new_elements(self._heat_time)
            self.parent._heat_time.inject_batch(self._heat_time, newel)
            self.parent._heat_temp.inject_batch(self._heat_temp, newel)
            self.parent._heat_duty.inject_batch(self._heat_duty, newel)

            newel = self.parent._co2_time.new_elements(self._co2_time)
            self.parent._co2_time.inject_batch(self._co2_time, newel)
            self.parent._co2.inject_batch(self._co2, newel)
            self.parent._co2_temp.inject_batch(self._co2_temp, newel)
            self.parent._humidity.inject_batch(self._humidity, newel)


class Collector(Generator):
    def __init__(self, *, rotary: Optional[LocalRotary] = None, port: int = 8100):
        # Collectors never log data
        super().__init__(rotary=rotary, no_save=True)

        self._collect_thread: Optional[CollectorThread] = None
        self.port = port

    def _get_data(self) -> None:
        if self._collect_thread is not None:
            self._collect_thread.access_collected_data()

    def _analyze_full(self) -> None:
        super()._analyze_full()

        self.rotary.alarms = self.alarms

    def _analyze_timeseries(self) -> None:
        super()._analyze_timeseries()

        if "Current Setting" in self.rotary:
            cur_setting: CurrentSetting = self.rotary["Current Setting"]

            cur_setting.from_processor(
                F=self.average_flow.get(2),
                P=self.average_pressure.get(2),
                RR=self.cumulative.get("RR"),
            )

        if "CO2 Setting" in self.rotary:
            if len(self._co2):
                co2_setting: CO2Setting = self.rotary["CO2 Setting"]
                co2_setting.from_processor(
                    co2=float(np.mean(self._co2[-5:])),
                    temp=float(np.mean(self._co2_temp[-5:])),
                    humidity=float(np.mean(self._humidity[-5:])),
                )

        if "Current Setting" in self.rotary or "CO2 Setting" in self.rotary:
            self.rotary.external_update()

        self.rotary.alarms = self.alarms

    @property
    def pressure(self) -> np.ndarray:
        return np.asarray(self._pressure)

    def run(self) -> None:
        super().run()
        self._collect_thread = CollectorThread(self)
        self._collect_thread.start()

    def close(self) -> None:
        super().close()
        if self._collect_thread is not None:
            self._collect_thread.join()
            self._collect_thread = None


if __name__ == "__main__":
    with Collector() as coll:

        time.sleep(5)
        with coll.lock:
            print(f"Received {len(coll.time)} values in 5 seconds")
            print(f"Analyzer length is {len(coll.volume)}")
        time.sleep(5)

        with coll.lock:
            print(f"Received {len(coll.time)} values in 10 seconds")
            print(f"Analyzer length is {len(coll.volume)}")
            print(f"Alarms: {coll.alarms}")


# Sadly, since we are not making this a full proper python package (at the moment),
# we have to do the following to run this file for testing:
# python3 -m patient.collector
```
