## Supplementary Git Repository for "Inexpensive multi-patient respiratory monitoring system for helmet ventilation during COVID-19 pandemic": config.py

```
from __future__ import annotations

import confuse
from pathlib import Path
from functools import lru_cache
from typing import Union
import logging

logger = logging.getLogger("povm")

DIR = Path(__file__).parent.resolve()

config = confuse.Configuration("povm", "povm")

# Currently we are not a library; change if we change
config.clear()

# Defaults stored here
config_default = DIR / "config_default.yml"
config.set_file(config_default)


def get_internal_file(filename: Union[Path, str]) -> Path:
    # A bit of a hacky workaround until we have a proper package
    return DIR.parent / filename


@lru_cache(1)
def get_data_dir() -> Path:
    """
    * relative path: relative to package
    * Path with ~ is expanded
    * Absolute paths supported
    """
    path = Path(config["global"]["datadir"].get()).expanduser()
    path = path if path.is_absolute() else DIR.parent / path
    if not path.exists():
        path.mkdir(parents=True)
        logger.info(f"Created directory: {path}")
    else:
        logger.info(f"Logging to directory: {path}")
    return path
```
