## Supplementary Git Repository for "Inexpensive multi-patient respiratory monitoring system for helmet ventilation during COVID-19 pandemic": config_default.yml

```
device:
  pressure:
    offset: 12.73
    scale: 0.0126
  flow:
    offset: 0.0
    scale: 0.7198

global:
  debug: false
  analyze-every: 3 # seconds
  save-every: 20 # seconds
  cumulative-every: 10 # seconds
  run-every: 0.5 # seconds
  window-size: 1600 # 30 seconds @ 50 hertz + 100 extra (2 seconds)
  extras-window-size: 160 # reads out ~every second - CO2 (if present), temp, etc.
  datadir: . # relative, with home, or absolute
  avg-window: 10 # seconds (pick from limited list)
  breath-thresh: 50 # ml

patient:
  buzzer-volume: 0 # max 255 (200 ideal) TODO: set to 200
  brightness: 200 # max 255
  silence-timeout: 120 # seconds
  silence-holddown: 0.2 # seconds

rotary-live:
  Reset Setting:
    type: Reset
    name: PushTurn to Reset
    order: -2

  Advanced:
    type: Advanced
    order: -1

  Current Setting:
    type: Current
    order: 0

  CO2 Setting:
    type: CO2
    order: -3
    name: CO2 sensor
```
