## Supplementary Git Repository for "Inexpensive multi-patient respiratory monitoring system for helmet ventilation during COVID-19 pandemic": device_names.py

```
#!/usr/bin/env python3

from __future__ import annotations

from processor.config import get_internal_file
from typing import TYPE_CHECKING

if TYPE_CHECKING:
    from typing_extensions import Final


vendors = ["dc:a6:32:00:00:00"]  # only append to this list

adjectives = [x.strip() for x in open(get_internal_file("data/adjectives.txt"))]
nouns = [x.strip() for x in open(get_internal_file("data/nouns.txt"))]

assert len(adjectives) == 4096
assert len(nouns) == 4096


def lcg_generator(
    state: int,
    multiplier: int = 1140671485,
    addend: int = 12820163,
    pmod: int = 2 ** 24,
) -> int:
    """
    Performs one step in a pseudorandom sequence from state (the input argument)
    to state the return value.

    The return value is an integer, uniformly distributed in [0, pmod).

    See https://en.wikipedia.org/wiki/Linear_congruential_generator
    """
    state = (multiplier * state + addend) % pmod
    return state


def address_to_addr(address: str) -> int:
    """
    Converts a MAC address string (hexidecimal with colons) into an integer.
    """
    return int(address.replace(":", ""), 16)


def addr_to_address(addr: int) -> str:
    """
    Converts a MAC address integer (48 bit) into a string (lowercase hexidecimal
    with colons).
    """
    out = "{0:012x}".format(addr)
    return ":".join(out[2 * i : 2 * i + 2] for i in range(6))


vendor_mask: Final[int] = 0xFFFFFF000000
adjective_mask: Final[int] = 0x000000FFF000
noun_mask: Final[int] = 0x000000000FFF


def address_to_name(address: str) -> str:
    """
    Converts a MAC address string (hexidecimal with colons) into a two-word name.

    The string is not guaranteed unique, but it is one of 16 million (4096**2)
    possibilities.

    This mapping is guaranteed to be stable (same MAC address in always yields
    the same name out) even if

       1. New vendors are added at the END of the processor.device_names.vendors
          list.
       2. Lines in data/adjectives.txt or data/nouns.txt that have never been
          used before are blanked out.

    (No other changes to the algorithm are known to provide such guarantees.)

    The output name will also be 20 characters or less. There is a 1% chance of
    collision and retry.

    The possibility of an infinite loop has not been theoretically ruled out
    (e.g. infinite collision and retry, somehow always hitting that 1% case),
    though if such a case is encountered in the future, we can fix it by blanking
    out a line on a word that leads to the loop.
    """
    addr = address_to_addr(address)
    state = addr & ~vendor_mask

    vendor_id = 0
    for vid, vendor_address in enumerate(vendors):
        vendor_addr = address_to_addr(vendor_address)
        if (addr & vendor_mask) == (vendor_addr & vendor_mask):
            vendor_id = vid
            break
    else:
        raise ValueError(f"unrecognized vendor: {addr_to_address(addr & vendor_mask)}")

    for _ in range(vendor_id + 1):
        state = lcg_generator(state)

    while True:
        adjective = adjectives[(state & adjective_mask) >> 12]
        noun = nouns[state & noun_mask]
        if adjective != "" and noun != "" and len(adjective) + len(noun) < 20:
            return adjective + " " + noun
        state = lcg_generator(state)


if __name__ == "__main__":
    import argparse

    parser = argparse.ArgumentParser(
        description="Convert a MAC address to a device name"
    )
    parser.add_argument("mac_address", nargs="+", help="MAC address(es) to convert")
    args = parser.parse_args()
    for addr in args.mac_address:
        print(f"{addr}: {address_to_name(addr)}")
```
