## Supplementary Git Repository for "Inexpensive multi-patient respiratory monitoring system for helmet ventilation during COVID-19 pandemic": display_settings.py

```
from processor.setting import SelectionSetting, DisplaySetting
from typing import Optional
from patient.mac_address import get_mac_addr
from processor.device_names import address_to_name
from pathlib import Path
from processor.version import get_version


class AdvancedSetting(SelectionSetting):
    STATIC_UPPER = False

    def __init__(self, *, rate: int = 2):

        string_listing = ["Box name", "MAC addr", "SensorID", "Log file", "Version"]

        # Sensor ID
        self.sid: int = 0
        self.file: str = ""

        super().__init__(0, string_listing, name="Advanced", rate=rate)

    @property
    def value(self) -> str:
        if self._value == 0:
            try:
                return address_to_name(get_mac_addr()).title()
            except ValueError:
                return "<Unknown>"
        elif self._value == 1:
            try:
                return get_mac_addr()
            except ValueError:
                return "<Unknown>"
        elif self._value == 2:
            if self.sid:
                return f"{self.sid:016X}"
            else:
                return "No sensor detected"
        elif self._value == 3:
            if self.file:
                return Path(self.file).name[-24:-4]
            else:
                return "Not recording"
        elif self._value == 4:
            return get_version() or "Unable to retrieve"
        else:
            raise NotImplementedError("Setting must be in range 0-3")

    @value.setter
    def value(self, value: int):
        pass

    def active(self) -> None:
        self._value = 0

    @property
    def lcd_name(self) -> str:
        return self._listing[self._value]

    # For the GUI
    def print_setting(self, value: int) -> str:
        c = self._value
        self._value = value
        res = f"{self.lcd_name}: {self.value}"
        self._value = c
        return res


class CO2Setting(DisplaySetting):
    STATIC_UPPER = False

    def __init__(self, *, name: str):

        self._co2: Optional[float] = None
        self._temp: Optional[float] = None
        self._humidity: Optional[float] = None

        super().__init__(name=name)

    def from_processor(
        self, co2: Optional[float], temp: Optional[float], humidity: Optional[float]
    ):

        self._co2 = co2
        self._temp = temp
        self._humidity = humidity

    @property
    def lcd_name(self):
        if self._co2 is None:
            return self._name
        return f"CO2: {self._co2:5.1f} H:{self._humidity:4.1f}"

    def __str__(self) -> str:
        if self._co2 is None:
            return "No sensor"
        return f"T: {self._temp: 5.2f}"

    def print_setting(self) -> str:
        return f"{self.lcd_name} {self}"


class CurrentSetting(DisplaySetting):
    STATIC_UPPER = False

    def __init__(self, *, name: str):

        self._F: Optional[float] = None
        self._P: Optional[float] = None
        self._RR: Optional[float] = None

        super().__init__(name=name)

    def from_processor(
        self, F: Optional[float], P: Optional[float], RR: Optional[float]
    ):
        self._F = F
        self._P = P
        self._RR = RR

    @property
    def lcd_name(self):
        if self._F is None or self._P is None:
            return self._name

        return f"F:{self._F:<6.1f} P:{self._P:.2f}"

    def __str__(self) -> str:
        if self._RR is None:
            return "RR: ---"
        return f"RR: {self._RR:.0f}"

    def print_setting(self) -> str:
        return f"{self.lcd_name} {self}"


class ResetSetting(SelectionSetting):
    def __init__(self, name: str, rate: int = 1):
        super().__init__(
            0,
            [f"[{x*'x':9}]" for x in range(9)] + ["[  RESET  ]"],
            name=name,
            rate=rate,
        )

    def active(self) -> None:
        self._value = 0

    @property
    def default(self) -> float:
        return 0.0

    @default.setter
    def default(self, val: float):
        pass

    def at_maximum(self):
        return self._value == (len(self._listing) - 1)
```
