## Supplementary Git Repository for "Inexpensive multi-patient respiratory monitoring system for helmet ventilation during COVID-19 pandemic": flow_calibrator.py

```
#!/usr/bin/env python3

import numpy as np
import scipy.interpolate
import yaml
import os
from typing import Union
from pathlib import Path

DIR = Path(__file__).parent.resolve()
DEFAULT_BLOCK = str(DIR / "flowcalib_data" / "flowcalib_ave200619.yml")


def get_yaml(yml_file):
    stream = open(yml_file, "r")
    params = yaml.safe_load(stream)
    qs = np.zeros(len(params))
    deltaPs = np.zeros(len(params))
    for i, d in enumerate(params):
        qs[i] = d["Q"]
        deltaPs[i] = d["deltaP"]

    return qs, deltaPs


class flow_calibrator:
    def __init__(self, block: str = DEFAULT_BLOCK) -> None:
        func = None
        if block == "simple":
            func = self.simple
        if block.endswith("yaml") or block.endswith("yml"):
            qs, deltaPs = get_yaml(block)
            self.interp = scipy.interpolate.interp1d(
                deltaPs, qs, kind="cubic", fill_value="extrapolate"
            )
            func = self.extrap1d

        assert func is not None
        self.func = func

    def simple(self, deltaP):
        #        return np.copysign(np.abs(deltaP) ** (4 / 7),deltaP)*0.7198/0.09636372314370535
        return np.copysign(np.abs(deltaP / 0.02962975) ** (4 / 7), deltaP)

    def extrap1d(self, in_deltaP):
        if not isinstance(in_deltaP, np.ndarray):
            deltaP = np.array(in_deltaP)
        else:
            deltaP = in_deltaP

        retval = self.interp(np.abs(deltaP))
        xs = self.interp.x
        beyond = deltaP > xs[-1]
        retval[beyond] = self.interp(xs[-1]) * np.power(
            deltaP[beyond] / xs[-1], 0.5
        )  # 4 / 7 is an alternative

        return np.copysign(retval, deltaP)

    def Q(self, f) -> Union[np.ndarray, float]:
        return self.func(f / 60.0)  # put f into Pa to get deltaP


if __name__ == "__main__":
    print("Checking old software calibration")
    caliber = flow_calibrator("simple")
    fs = np.arange(0.0, 33000.0, 1000.0)
    for f in fs:
        print(f, caliber.Q(f))

    print("Checking array syntax too")
    print(fs)
    print(caliber.Q(fs))

    print("Checking the default")
    caliber_def = flow_calibrator()
    print(caliber.Q(fs))

    fs = np.arange(0.0, 33000.0, 1000.0)
    res1 = caliber.Q(fs)

    files = ["flowcalib_all.yml", "flowcalib_xometry.yml", "flowcalib_ave200619.yml"]
    labels = ["All flowblocks", "Xometry flowblocks", "Selected June 18 flowblocks"]

    import matplotlib.pyplot as plt

    res_list = []
    for i, f in enumerate(files):
        caliber_f = flow_calibrator(
            os.path.join(os.path.dirname(__file__), "flowcalib_data", f)
        )
        res_list.append(caliber_f.Q(fs))
        plt.plot(fs, res_list[-1], label=labels[i])

    plt.plot(fs, res1, label="Previous software calibration")
    plt.xlabel("measured f")
    plt.ylabel("Q (L/min)")
    plt.legend(loc="best")
    plt.show()
```
