## Supplementary Git Repository for "Inexpensive multi-patient respiratory monitoring system for helmet ventilation during COVID-19 pandemic": flowcalib_all.yml

```
-
  Q: 0.
  deltaP: 0.
-
  Q: 9.74
  deltaP: 2.0838
-
  Q: 20.067
  deltaP: 6.5345
-
  Q: 30.185
  deltaP: 13.615
-
  Q: 39.998
  deltaP: 22.392
-
  Q: 49.976
  deltaP: 33.309
-
  Q: 60.008
  deltaP: 46.424
-
  Q: 70.148
  deltaP: 62.638
-
  Q: 79.823
  deltaP: 81.139
-
  Q: 89.668
  deltaP: 102.9
-
  Q: 100.02
  deltaP: 128.72
```
