## Supplementary Git Repository for "Inexpensive multi-patient respiratory monitoring system for helmet ventilation during COVID-19 pandemic": flowcalib_ave200619.yml

```
-
  Q: 0.0
  deltaP: 0.0
-
  Q: 1.0
  deltaP: 0.13433118438194872
-
  Q: 2.0
  deltaP: 0.29461267092953114
-
  Q: 3.0
  deltaP: 0.471244447812402
-
  Q: 4.0
  deltaP: 0.6647799280473008
-
  Q: 5.0
  deltaP: 0.8757725246509668
-
  Q: 6.0
  deltaP: 1.104775650640139
-
  Q: 7.0
  deltaP: 1.3523427190315576
-
  Q: 8.0
  deltaP: 1.619027142841961
-
  Q: 9.0
  deltaP: 1.905382335088089
-
  Q: 10.0
  deltaP: 2.211961708786681
-
  Q: 11.0
  deltaP: 2.5393186769544753
-
  Q: 12.0
  deltaP: 2.8880143725084486
-
  Q: 13.0
  deltaP: 3.258790747538581
-
  Q: 14.0
  deltaP: 3.6525801625111147
-
  Q: 15.0
  deltaP: 4.070324740951361
-
  Q: 16.0
  deltaP: 4.512966606384623
-
  Q: 17.0
  deltaP: 4.98144788233621
-
  Q: 18.0
  deltaP: 5.476710692331422
-
  Q: 19.0
  deltaP: 5.999697159895568
-
  Q: 20.0
  deltaP: 6.551369461883587
-
  Q: 21.0
  deltaP: 7.132541267490982
-
  Q: 22.0
  deltaP: 7.7427702539287075
-
  Q: 23.0
  deltaP: 8.380828245887109
-
  Q: 24.0
  deltaP: 9.04523935625748
-
  Q: 25.0
  deltaP: 9.734522365831944
-
  Q: 26.0
  deltaP: 10.447196055402618
-
  Q: 27.0
  deltaP: 11.181779205761615
-
  Q: 28.0
  deltaP: 11.936790597701062
-
  Q: 29.0
  deltaP: 12.710749134535119
-
  Q: 30.0
  deltaP: 13.502206409019259
-
  Q: 31.0
  deltaP: 14.310238723298383
-
  Q: 32.0
  deltaP: 15.135131780934156
-
  Q: 33.0
  deltaP: 15.977470249833583
-
  Q: 34.0
  deltaP: 16.837841922969044
-
  Q: 35.0
  deltaP: 17.7168345933129
-
  Q: 36.0
  deltaP: 18.615036053837528
-
  Q: 37.0
  deltaP: 19.53303409751529
-
  Q: 38.0
  deltaP: 20.47141651731857
-
  Q: 39.0
  deltaP: 21.430771119283843
-
  Q: 40.0
  deltaP: 22.41171415585843
-
  Q: 41.0
  deltaP: 23.414812329827082
-
  Q: 42.0
  deltaP: 24.440073585626184
-
  Q: 43.0
  deltaP: 25.487314950301847
-
  Q: 44.0
  deltaP: 26.556353330632014
-
  Q: 45.0
  deltaP: 27.647005633394556
-
  Q: 46.0
  deltaP: 28.75908876536745
-
  Q: 47.0
  deltaP: 29.89241963332859
-
  Q: 48.0
  deltaP: 31.04681514405594
-
  Q: 49.0
  deltaP: 32.222092204327375
-
  Q: 50.0
  deltaP: 33.41809206249345
-
  Q: 51.0
  deltaP: 34.63509061251415
-
  Q: 52.0
  deltaP: 35.87389970149652
-
  Q: 53.0
  deltaP: 37.1353703080552
-
  Q: 54.0
  deltaP: 38.420351624849594
-
  Q: 55.0
  deltaP: 39.729692844539045
-
  Q: 56.0
  deltaP: 41.064243159783
-
  Q: 57.0
  deltaP: 42.42485176324081
-
  Q: 58.0
  deltaP: 43.81236784757186
-
  Q: 59.0
  deltaP: 45.22764060543556
-
  Q: 60.0
  deltaP: 46.67153663090511
-
  Q: 61.0
  deltaP: 48.14498673562198
-
  Q: 62.0
  deltaP: 49.648342638326454
-
  Q: 63.0
  deltaP: 51.1816114018193
-
  Q: 64.0
  deltaP: 52.74479479709318
-
  Q: 65.0
  deltaP: 54.33789459514075
-
  Q: 66.0
  deltaP: 55.960912566954725
-
  Q: 67.0
  deltaP: 57.61385048352775
-
  Q: 68.0
  deltaP: 59.29671011585255
-
  Q: 69.0
  deltaP: 61.009496295317575
-
  Q: 70.0
  deltaP: 62.752318119488585
-
  Q: 71.0
  deltaP: 64.52560549999231
-
  Q: 72.0
  deltaP: 66.32980689317074
-
  Q: 73.0
  deltaP: 68.16532416951136
-
  Q: 74.0
  deltaP: 70.03255911019357
-
  Q: 75.0
  deltaP: 71.93191349639694
-
  Q: 76.0
  deltaP: 73.86378910930105
-
  Q: 77.0
  deltaP: 75.82858773008519
-
  Q: 78.0
  deltaP: 77.82671113992899
-
  Q: 79.0
  deltaP: 79.85856111999212
-
  Q: 80.0
  deltaP: 81.9245108754713
-
  Q: 81.0
  deltaP: 84.02459260084716
-
  Q: 82.0
  deltaP: 86.15812152386691
-
  Q: 83.0
  deltaP: 88.32419875002277
-
  Q: 84.0
  deltaP: 90.52192370830832
-
  Q: 85.0
  deltaP: 92.75039582771709
-
  Q: 86.0
  deltaP: 95.0087145372426
-
  Q: 87.0
  deltaP: 97.2959792658784
-
  Q: 88.0
  deltaP: 99.61128944261793
-
  Q: 89.0
  deltaP: 101.95374449645482
-
  Q: 90.0
  deltaP: 104.32244385638256
-
  Q: 91.0
  deltaP: 106.71648695139478
-
  Q: 92.0
  deltaP: 109.13497321048496
-
  Q: 93.0
  deltaP: 111.5770020626465
-
  Q: 94.0
  deltaP: 114.0416729368731
-
  Q: 95.0
  deltaP: 116.52808526215831
-
  Q: 96.0
  deltaP: 119.03533846749542
-
  Q: 97.0
  deltaP: 121.56253198187831
-
  Q: 98.0
  deltaP: 124.10876523430028
-
  Q: 99.0
  deltaP: 126.67313765375498
```
