## Supplementary Git Repository for "Inexpensive multi-patient respiratory monitoring system for helmet ventilation during COVID-19 pandemic": flowcalib_xometry.yml

```
-
  Q: 0
  deltaP: 0
-
  Q: 9.7175
  deltaP: 2.0167
-
  Q: 19.956
  deltaP: 6.3291
-
  Q: 30.01
  deltaP: 13.079
-
  Q: 39.975
  deltaP: 21.805
-
  Q: 49.995
  deltaP: 32.571
-
  Q: 59.91
  deltaP: 45.333
-
  Q: 70.011
  deltaP: 61.297
-
  Q: 79.849
  deltaP: 80.186
-
  Q: 89.733
  deltaP: 102.25
-
  Q: 100.04
  deltaP: 127.93
```
