## Supplementary Git Repository for "Inexpensive multi-patient respiratory monitoring system for helmet ventilation during COVID-19 pandemic": gen_record.py

```
from __future__ import annotations

from typing import Optional
from dataclasses import dataclass
from logging import Logger
from processor.device_names import address_to_name


@dataclass
class GenRecord:
    logger: Logger

    _mac: Optional[str] = None
    _sid: int = 0
    _name: Optional[str] = None

    # Nurse only, but in master class to make simpler
    _nurse_name: str = ""

    @property
    def safemac(self) -> str:
        """Returns a safe filename (Windows does not allow colons)"""
        keepcharacters = (".", "_")
        return "".join(
            c for c in self.mac if c.isalnum() or c in keepcharacters
        ).rstrip()

    @property
    def mac(self) -> str:
        """
        The mac address. Returns <unknown> if the address is not known.
        """
        return "<unknown>" if self._mac is None else self._mac

    @mac.setter
    def mac(self, value: str):
        if self._mac is None or self._mac != value:
            self._mac = value
            self.logger.info(f"MAC addr: {self._mac}")
            self.mac_changed()

    @property
    def sid(self) -> int:
        """
        Sensor ID, as an integer. Printout with "X" format.
        """
        return self._sid

    @sid.setter
    def sid(self, value: int):
        if self._sid != value:
            self._sid = value
            self.logger.info(f"Sensor ID: {self._sid:X}")
            self.sid_changed()

    @property
    def box_name(self) -> str:
        """
        The name of the box, or <unknown>.
        """
        if self._name is not None:
            return self._name

        if self._mac is None:
            return "<unknown>"

        try:
            return address_to_name(self._mac).title()
        except ValueError:
            return self.mac

    @box_name.setter
    def box_name(self, value: Optional[str]):
        if self._name != value:
            self._name = value
            self.logger.info(f"Box name: {self._name}")
            self.mac_changed()

    @property
    def stacked_name(self) -> str:
        """
        Return the box name stacked using a newline
        If unknown, return Box name: <unknown>.
        """

        if self._mac is None or self._mac == "00:00:00:00:00:00":
            return "Box name\n<unknown>"
        try:
            return "\n".join(address_to_name(self._mac).title().split())
        except ValueError:
            return self.mac

    @property
    def title(self) -> str:
        """
        The title to show in the dialog box. Will show box_name if unset.
        """
        return self._nurse_name

    @title.setter
    def title(self, value: str):
        if self._nurse_name is None or self._nurse_name != value:
            self._nurse_name = value
            self.logger.info(f"Changed title to {self._nurse_name!r}")
            self.title_changed()

    def title_changed(self) -> None:
        """
        Modify in subclasses to add special callbacks here.
        """

    def mac_changed(self) -> None:
        """
        Modify in subclasses to add special callbacks here.
        """

    def sid_changed(self) -> None:
        """
        Modify in subclasses to add special callbacks here.
        """
```
