## Supplementary Git Repository for "Inexpensive multi-patient respiratory monitoring system for helmet ventilation during COVID-19 pandemic": generator.py

```
from __future__ import annotations

import abc
import enum
import threading
import time
import warnings
import logging

import numpy as np
from typing import Dict, Any, List, Optional, TypeVar, TYPE_CHECKING
from pathlib import Path
from datetime import datetime

import processor.analysis
from processor.rotary import LocalRotary
from processor.settings import get_remote_settings
from processor.config import config
from processor.rolling import Rolling
from processor.saver import CSVSaverTS, CSVSaverCML, JSONSSaverBreaths, FieldInfo
from processor.gen_record import GenRecord

if TYPE_CHECKING:
    from typing_extensions import Final


class Status(enum.Enum):
    OK = "OK"
    ALERT = "!"
    SILENT = "S"
    ALERT_SILENT = "S!"
    DISCON = "D"
    NONE = "?"


T = TypeVar("T", bound="Generator")


class Generator(abc.ABC):
    # Keeps track of how many generators have been created; each gets a unique ID
    # that lasts for the session.
    _total_generators: int = 0

    def __init__(
        self,
        *,
        rotary: processor.rotary.LocalRotary = None,
        logger: logging.Logger = None,
        no_save: bool = False,
        gen_record: GenRecord = None,
    ) -> None:

        # The size of the rolling window
        self.window_size = config["global"]["window-size"].get(int)  # seconds

        # The size of the extras window (CO2 if present, etc.)
        self.extras_window_size = config["global"]["extras-window-size"].get(
            int
        )  # seconds

        # The raw timestamps
        self._time = Rolling(window_size=self.window_size, dtype=np.int64)

        # The flow
        self._flow = Rolling(window_size=self.window_size)

        # The pressure
        self._pressure = Rolling(window_size=self.window_size)

        # Heating timestamps
        self._heat_time = Rolling(window_size=self.extras_window_size, dtype=np.int64)

        # The heating element temp
        self._heat_temp = Rolling(window_size=self.extras_window_size)

        # The heating element duty cycle
        self._heat_duty = Rolling(window_size=self.extras_window_size)

        # CO2 timestamps
        self._co2_time = Rolling(window_size=self.extras_window_size, dtype=np.int64)

        # The CO2 levels (if hardware present)
        self._co2 = Rolling(window_size=self.extras_window_size)

        # The humidity levels (if hardware present)
        self._humidity = Rolling(window_size=self.extras_window_size)

        # The temperature levels (if hardware present)
        self._co2_temp = Rolling(window_size=self.extras_window_size)

        # The volume, expected to be generated by ._analyze_timeseries()
        self._volume = np.array([], dtype=np.double)

        # Minimum bias volume, not shown on screen, used to compute volume differences,
        # generated by ._analyze_timeseries()
        self._minbias_volume = np.array([], dtype=np.double)

        # The previous collection of realtime values, as floats
        self._old_realtime: Optional[np.ndarray] = None

        # Defines the "zero" of volume
        self._volume_unshifted_min: Optional[float] = None

        # The difference between the volume we show and volume with a mean of 0
        self._volume_shift = 0.0

        # The cumulative running windows for flow
        self._flow_cumulative = {
            2: 0.0,
            10: 0.0,
        }

        # The cumulative running windows for pressure
        self._pressure_cumulative = {
            2: 0.0,
            10: 0.0,
        }

        # The list of breaths
        self._breaths: List[Dict[str, float]] = []

        # The list of cumulative values from the analysis
        self._cumulative: Dict[str, float] = {}

        # Timestamps on all cumulative keys for calculating staleness
        self._cumulative_timestamps: Dict[str, Any] = {}

        # The active alarms
        self._alarms: Dict[str, Dict[str, float]] = {}

        # The active average alarms
        self._avg_alarms: Dict[str, Dict[str, float]] = {}

        # The rotary with alarm settings
        self.rotary = LocalRotary(get_remote_settings()) if rotary is None else rotary

        # How often to rerun analyze (full analyze)
        self.analyze_every = config["global"]["analyze-every"].as_number()  # seconds

        # How often to process data in automatic mode (partial analyze)
        self.run_every = config["global"]["run-every"].as_number()  # seconds

        # Last updated datetime.now()
        self.last_update: Optional[datetime] = None

        # Last analyze run in local time - used by analyze_as_needed
        self._last_ana = time.monotonic()

        # Last partial analyze for plotting
        self._last_get: Optional[float] = None

        # Status of the generator alarms, set in get_data
        self.status: Status = Status.NONE

        # Path to write data to (needs name change)
        self._logging: Optional[Path] = None

        # This lock ensures validity of the access when threading.
        # This may be "reentered" by analyze calling the properties; that is fine.
        self.lock = threading.RLock()

        # A thread to run the analyze loop in the background
        self._run_thread: Optional[threading.Thread] = None

        # A stop signal to turn off the thread
        self.stop = threading.Event()

        # A quick way to get the debug status
        self._debug = config["global"]["debug"].get(bool)

        # The breath threshold
        self.breath_thresh: float = config["global"]["breath-thresh"].as_number()

        # An incrementing unique ID (for logging, perhaps)
        self.gen_id: Final[int] = self._total_generators
        self._total_generators += 1

        # The logger instance
        self.logger: logging.Logger = logger or logging.getLogger("povm")

        # Used by GUI to bundle information
        self.record = GenRecord(self.logger) if gen_record is None else gen_record

        # Saver instances
        self.saver_ts: Optional[CSVSaverTS] = None
        self.saver_heat: Optional[CSVSaverTS] = None
        self.saver_co2: Optional[CSVSaverTS] = None
        self.saver_cml: Optional[CSVSaverCML] = None
        self.saver_breaths: Optional[JSONSSaverBreaths] = None

        if no_save:
            return

        handlers = self.logger.handlers
        file_handlers = [
            handler for handler in handlers if isinstance(handler, logging.FileHandler)
        ]
        if file_handlers:
            handler: logging.FileHandler = file_handlers[0]
            log_path = Path(handler.baseFilename).parent

            self.saver_ts = CSVSaverTS(
                (
                    FieldInfo("t", "_time"),
                    FieldInfo("f", "_flow", ".2"),
                    FieldInfo("p", "_pressure", ".3"),
                ),
                self,
                log_path / "ts.csv",
                config["global"]["save-every"].as_number(),
            )

            self.saver_heat = CSVSaverTS(
                (
                    FieldInfo("t", "_heat_time"),
                    FieldInfo("C", "_heat_temp", ".4"),
                    FieldInfo("D", "_heat_duty", ".4"),
                ),
                self,
                log_path / "heat.csv",
                config["global"]["save-every"].as_number(),
            )

            self.saver_co2 = CSVSaverTS(
                (
                    FieldInfo("t", "_co2_time"),
                    FieldInfo("CO2", "_co2", ".5"),
                    FieldInfo("Tp", "_co2_temp", ".3"),
                    FieldInfo("H", "_humidity", ".3"),
                ),
                self,
                log_path / "co2.csv",
                config["global"]["save-every"].as_number(),
            )

            self.saver_cml = CSVSaverCML(
                self,
                log_path / "cml.csv",
                config["global"]["cumulative-every"].as_number(),
            )
            self.saver_breaths = JSONSSaverBreaths(self, log_path / "breaths.jsons")
        else:
            self.logger.info(
                "No file-based logging attached, not saving time series or cumulatives"
            )

    def run(self) -> None:
        """
        Start running an analysis loop. Calling get_data and analyze_as_needed manually are not recommended
        while this is running in the background. Be sure to close/exit context to close and clean up the thread.

        Started by the context manager
        """

        self.stop.clear()
        self.logger.info("Starting run")

        if self.saver_ts is not None:
            self.saver_ts.enter()
        if self.saver_heat is not None:
            self.saver_heat.enter()
        if self.saver_co2 is not None:
            self.saver_co2.enter()
        if self.saver_cml is not None:
            self.saver_cml.enter()
        if self.saver_breaths is not None:
            self.saver_breaths.enter()

        for k, v in self.rotary.to_dict().items():
            self.logger.info(f"rotary: {k} set to {v['value']} (initial value)")

        self._run_thread = threading.Thread(target=self._logging_run)
        self._run_thread.start()

    def _logging_run(self) -> None:
        try:
            self._run()
        except Exception:
            self.logger.exception("Unexpected error in analysis!")
            raise

    def _run(self) -> None:
        """
        Must be run in the background, by run()
        """

        while not self.stop.is_set():
            with self.lock:
                self._get_data()
                self.analyze_as_needed()
            self.stop.wait(self.run_every)

    def analyze_as_needed(self) -> None:
        """
        Run basic analysis, and more complex analysis only if needed.
        """
        self._analyze_timeseries()

        if time.monotonic() - self._last_ana > self.analyze_every:
            self._analyze_full()

            self._last_ana = time.monotonic()

            self._set_alarms()

            if self.saver_cml:
                self.saver_cml.save()

        if self.saver_ts:
            self.saver_ts.save()

        if self.saver_heat:
            self.saver_heat.save()

        if self.saver_co2:
            self.saver_co2.save()

    def _set_alarms(self):
        "Overridden in remote generator to include silenced alarms. Collector doens't care."
        self.status = Status.ALERT if self.alarms else Status.OK

    @abc.abstractmethod
    def _get_data(self):
        """
        Copy in the remote/local datastream to internal cache.
        """

    def _analyze_timeseries(self) -> None:
        """
        Quick analysis that's easier to run often, makes volume (run by `analyze` too)
        """
        avg_window = config["global"]["avg-window"].get(int)

        realtime = self.realtime
        if len(realtime) > 0:
            if self._logging is not None:
                if self._logging.exists() and not self._logging.is_dir():
                    warnings.warn(f"{self._logging} is not a directory; not logging")
                else:
                    if not self._logging.exists():
                        self._logging.mkdir()

                    if self._old_realtime is None or len(self._old_realtime) == 0:
                        start_index = 0
                    else:
                        start_index = (
                            np.argmin(abs(realtime - self._old_realtime[-1])) + 1
                        )

                    with open(self._logging / f"time_{id(self)}.dat", "ba") as file:
                        file.write(
                            (realtime[start_index:] * 1000).astype("<u8").tostring()
                        )

                    with open(self._logging / f"flow_{id(self)}.dat", "ba") as file:
                        file.write(self.flow[start_index:].astype("<f4").tostring())

                    with open(self._logging / f"time_{id(self)}.dat", "ba") as file:
                        file.write(self.pressure[start_index:].astype("<f4").tostring())

            self._flow_cumulative = processor.analysis.compute_cumulative(
                self._flow_cumulative.keys(), self.timestamps, self.flow
            )

            self._pressure_cumulative = processor.analysis.compute_cumulative(
                self._pressure_cumulative.keys(), self.timestamps, self.pressure
            )

            if len(self.realtime) > 0:
                processor.analysis.avg_alarms(
                    self._avg_alarms,
                    self.rotary,
                    "flow",
                    self._flow_cumulative,
                    self.realtime[-1],
                    self.logger,
                )
                processor.analysis.avg_alarms(
                    self._avg_alarms,
                    self.rotary,
                    "pressure",
                    self._pressure_cumulative,
                    self.realtime[-1],
                    self.logger,
                )

                if len(self.co2_realtime) > 0:
                    processor.analysis.co2_alarm(
                        self._avg_alarms,
                        self.rotary,
                        self.co2[self.co2_time < avg_window],
                        self.realtime[-1],
                        self.logger,
                    )

            self._volume = processor.analysis.flow_to_volume(
                realtime,
                self._old_realtime,
                self.flow,
                self._volume - self._volume_shift,
                critical_frequency=0.004,
            )
            self._old_realtime = realtime

            # Removed "volume minimum is minimum ever"
            # Added "volume minimum is minimum on screen"
            #
            # if self._volume_unshifted_min is None:
            #     self._volume_unshifted_min = np.min(self._volume)
            # else:
            #     self._volume_unshifted_min = min(
            #         self._volume_unshifted_min, np.min(self._volume)
            #     )
            self._volume_unshifted_min = np.min(self._volume[-15 * 50 :])
            #
            # END

            self._volume_shift = (
                -self._volume_unshifted_min
                if self._volume_unshifted_min is not None
                else 0
            )
            self._volume = self._volume + self._volume_shift

            self._minbias_volume = processor.analysis.flow_to_volume(
                realtime,
                None,
                self.flow - np.mean(self.flow),
                None,
                critical_frequency=0.0004,
            )

    def _analyze_full(self) -> None:
        """
        Full analysis of breaths.
        """

        realtime = self.realtime

        updated = []
        new_breaths = []
        updated_fields = set()

        if len(realtime) > 0:
            breaths = processor.analysis.measure_breaths(
                realtime,
                self.flow,
                self._minbias_volume,
                self.pressure,
                breath_thresh=self.breath_thresh,
            )

            if len(breaths) > 0:
                (
                    all_breaths,
                    updated,
                    new_breaths,
                ) = processor.analysis.combine_breaths(self._breaths, breaths)

                if self.saver_breaths is not None:
                    self.saver_breaths.save_breaths(all_breaths[:-30])
                self._breaths = all_breaths[-30:]

                self._cumulative, updated_fields = processor.analysis.cumulative(
                    self._cumulative, updated, new_breaths
                )

        self._alarms = processor.analysis.add_alarms(
            self.rotary,
            updated,
            new_breaths,
            self._cumulative,
            self._alarms,
            self.logger,
        )

        timestamp = time.time()
        cumulative_timestamps = dict(self._cumulative_timestamps)
        cumulative_timestamps[""] = timestamp
        for field in updated_fields:
            cumulative_timestamps[field] = timestamp
        self._cumulative_timestamps = cumulative_timestamps

        stale_threshold = (
            self.rotary["Stale Data Timeout"].value
            if "Stale Data Timeout" in self.rotary
            else (10 if self._debug else None)
        )

        # This can be None or 0 to deactivate
        if stale_threshold:
            default = timestamp - stale_threshold
            stale = {}
            for field in self._cumulative:
                last_update_timediff = timestamp - self._cumulative_timestamps.get(
                    field, default
                )
                if last_update_timediff >= stale_threshold:
                    stale[field] = last_update_timediff

            old_stale = self._alarms.get("Stale Data", {})
            for name in stale:
                if name not in old_stale:
                    self.logger.info(f"Stale data alarm for {repr(name)} activated")

            for name in old_stale:
                if name not in stale:
                    self.logger.info(f"Stale data alarm for {repr(name)} deactivated")

            if len(stale) > 0:
                self._alarms["Stale Data"] = stale

    @property
    def tardy(self) -> float:
        """
        The amount of time since last update.
        """
        return (
            (time.monotonic() - self._last_get) if self._last_get is not None else 0.0
        )

    @property
    def time(self) -> np.ndarray:
        """
        The time array, with the most recent time as 0, with an adjustment based on `last_update`. Mostly for plotting.
        """
        timestamps = self.timestamps

        if len(timestamps) > 0:
            return -(timestamps - timestamps[-1]) / 1000 + self.tardy
        else:
            return timestamps

    @property
    def co2_time(self) -> np.ndarray:
        """
        The co2 time array, with the most recent time as 0, with an adjustment based on `last_update`. Mostly for plotting.
        """
        co2_timestamps = np.asarray(self._co2_time)
        timestamps = self.timestamps

        if len(co2_timestamps) > 0 and len(timestamps) > 0:
            return -(co2_timestamps - timestamps[-1]) / 1000 + self.tardy
        else:
            return co2_timestamps

    @property
    def realtime(self) -> np.ndarray:
        """
        The actual time in seconds (arbitrary monotonic device clock)
        """
        return self.timestamps / 1000

    @property
    def co2_realtime(self) -> np.ndarray:
        """
        The actual time for the CO2 time
        """

        return np.asarray(self._co2_time) / 1000

    @property
    def timestamps(self) -> np.ndarray:
        """
        Raw timestamps.
        """
        return np.asarray(self._time)

    @property
    def flow(self) -> np.ndarray:
        """
        Raw flow data.
        """
        return np.asarray(self._flow)

    @property
    def co2(self) -> np.ndarray:
        """
        The CO2 values, if present.
        """

        return np.asarray(self._co2)

    @property
    @abc.abstractmethod
    def pressure(self) -> np.ndarray:
        """
        Raw pressure data. (some generators implement a filter here)
        """
        pass

    @property
    def volume(self) -> np.ndarray:
        """
        Computed volume.
        """
        return self._volume

    @property
    def breaths(self) -> List[Any]:
        """
        The last 30 computed breaths.
        """
        return self._breaths

    @property
    def alarms(self) -> Dict[str, Dict[str, float]]:
        """
        A dictionary of alarms, each with first and last time, and extreme value.
        """
        return {**self._alarms, **self._avg_alarms}

    @property
    def cumulative(self) -> Dict[str, float]:
        """
        A dictionary of resulting values, like RR, from the analysis.
        """
        return self._cumulative

    @property
    def cumulative_timestamps(self):
        """
        Timestamps on all cumulative keys for calculating staleness
        """

        return self._cumulative_timestamps

    @property
    def average_flow(self) -> Dict[int, float]:
        """
        The cumulative running averages
        """
        return self._flow_cumulative

    @property
    def average_pressure(self) -> Dict[int, float]:
        """
        The cumulative running averages
        """
        return self._pressure_cumulative

    def close(self) -> None:
        """
        Always close or use a context manager if running threads!
        """
        self.logger.info("Stopping run")
        self.stop.set()
        if self._run_thread is not None:
            self._run_thread.join()

        if self.saver_ts is not None:
            self.saver_ts.close()
        if self.saver_heat is not None:
            self.saver_heat.close()
        if self.saver_co2 is not None:
            self.saver_co2.close()
        if self.saver_cml is not None:
            self.saver_cml.close()
        if self.saver_breaths is not None:
            self.saver_breaths.close()

    def __enter__(self: T) -> T:
        self.run()
        return self

    def __exit__(self, *exc) -> None:
        self.close()
```
