## Supplementary Git Repository for "Inexpensive multi-patient respiratory monitoring system for helmet ventilation during COVID-19 pandemic": listener.py

```
#!/usr/bin/env python3
from __future__ import annotations

from zeroconf import Zeroconf, ServiceBrowser, ServiceListener
import threading
import queue
import logging
import ipaddress
from typing import Set, Callable
from dataclasses import dataclass
from processor.device_names import address_to_name

logger = logging.getLogger("povm")


@dataclass
class Detector:
    address: ipaddress.IPv4Address
    port: int
    mac: str
    service: str
    name: str

    @property
    def url(self):
        return f"tcp://{ipaddress.ip_address(self.address)}:{self.port}"

    def __str__(self):
        return f"{self.name} @ {ipaddress.ip_address(self.address)}:{self.port}"

    def __hash__(self):
        return hash((self.address, self.port))


class Listener(ServiceListener):
    def __init__(self):
        self.detected: Set[Detector] = set()
        self.inject: Callable[[], None] = lambda: None
        self.queue: queue.Queue[Detector] = queue.Queue()

    def _injects(self, addrs: Set[Detector]):
        new = addrs - self.detected
        self.detected |= addrs
        for item in new:
            self.queue.put(item)
        self.inject()

    def add_service(self, zeroconf: Zeroconf, service_type: str, name: str) -> None:
        addrs = self._add_if_unseen("added", zeroconf, service_type, name)
        self._injects(addrs)

    def update_service(self, zeroconf: Zeroconf, service_type: str, name: str) -> None:
        addrs = self._add_if_unseen("updated", zeroconf, service_type, name)
        self._injects(addrs)

    def remove_service(self, zeroconf: Zeroconf, service_type: str, name: str) -> None:
        self._add_if_unseen("updated", zeroconf, service_type, name)

    def _add_if_unseen(
        self, status: str, zeroconf: Zeroconf, service_type: str, name: str
    ) -> Set[Detector]:
        if "Princeton Open Vent" in name:
            info = zeroconf.get_service_info(service_type, name)
            if not info:
                logger.info(f"Service {name} not {status}, missing info!")
                return set()

            macaddr = info.properties.get(b"mac_addr", b"<unknown>").decode()
            service = info.properties.get(b"service", b"<unknown>").decode()
            box_name = info.properties.get(b"name", b"").decode()
            if not box_name:
                try:
                    box_name = address_to_name(macaddr)
                except ValueError:
                    box_name = macaddr

            addresses = {
                Detector(
                    ipaddress.ip_address(ip), info.port or 0, macaddr, service, box_name
                )
                for ip in info.addresses
            }

            logger.info(f"Service {name} {status}: {addresses}")
            return addresses

        else:
            logger.debug(f"Service {name} not {status}")
            return set()


class FindBroadcasts:
    def __init__(self):
        self.zeroconf = Zeroconf()
        self.listener = Listener()

    def __enter__(self) -> FindBroadcasts:
        self.browser = ServiceBrowser(self.zeroconf, "_http._tcp.local.", self.listener)
        return self

    def __exit__(self, *exc) -> None:
        self.zeroconf.close()

    @property
    def detected(self) -> Set[Detector]:
        return self.listener.detected

    @property
    def inject(self) -> Callable[[], None]:
        return self.listener.inject

    @inject.setter
    def inject(self, func: Callable[[], None]):
        self.listener.inject = func

    @property
    def queue(self) -> queue.Queue:
        return self.listener.queue


if __name__ == "__main__":
    fb = FindBroadcasts()

    def proc():
        while not fb.queue.empty():
            print("Detected:", fb.queue.get())

    fb.inject = proc

    try:
        with fb:
            forever = threading.Event()
            forever.wait()
    except KeyboardInterrupt:
        print(f"\nDuring run, detected {len(fb.detected)}:", *fb.detected, sep="\n  ")
        print("\n\nIPs:", *(x.address for x in fb.detected))
```
