## Supplementary Git Repository for "Inexpensive multi-patient respiratory monitoring system for helmet ventilation during COVID-19 pandemic": local_generator.py

```
import time
from datetime import datetime
import numpy as np
import logging

from sim.start_sims import start_sims
from processor.generator import Generator, Status
from processor.gen_record import GenRecord


class LocalGenerator(Generator):
    def __init__(self, *, i: int, logger: logging.Logger, gen_record: GenRecord = None):
        super().__init__(logger=logger, gen_record=gen_record)
        self.status = Status.OK

        self.record.mac = f"dc:a6:32:00:00:{i:02x}"

        self._start_time = int(1000 * time.monotonic())
        (self._sim,) = start_sims(1, self._start_time, 12000000)

    def _get_data(self):
        t = int(time.monotonic() * 1000)
        events = t - self._time[-1] if len(self._time) else self._start_time
        times, flow, pressure = self._sim.get_from_timestamp(t, events)

        to_add = self._time.new_elements(times)
        if to_add:
            self._time.inject(times[-to_add:])
            self._flow.inject(flow[-to_add:])
            self._pressure.inject(pressure[-to_add:])

        self._last_get = self._time[-1] / 1000
        self.last_update = datetime.now()

    @property
    def flow(self):
        return np.asarray(self._flow)

    @property
    def pressure(self):
        return np.asarray(self._pressure)

    @property
    def timestamps(self):
        return np.asarray(self._time)
```
