## Supplementary Git Repository for "Inexpensive multi-patient respiratory monitoring system for helmet ventilation during COVID-19 pandemic": logging.py

```
from __future__ import annotations

from pathlib import Path
from typing import Optional
import logging
from datetime import datetime
import time

from processor.config import config, get_data_dir


def name_of_next(mypath: Path) -> Path:
    """
    Get the next available file
    """
    while True:
        dt = datetime.now().strftime("%Y%m%d_%H%M%S")
        name = "{n}_{dt}{s}".format(n=mypath.stem, dt=dt, s=mypath.suffix)
        new_file_path = mypath.with_name(name)
        if not new_file_path.exists():
            return new_file_path
        else:
            time.sleep(1)


def init_logger(logstr: Optional[str] = None) -> None:
    """
    logstr should be nurse_log/nursegui.log or similar (or None for screen only, even non-debug)
    """

    logger = logging.getLogger("povm")
    formatter = logging.Formatter(
        "%(asctime)s - %(levelname)s - %(message)s",
        "%Y-%m-%d %H:%M:%S",
    )

    if logstr is None or config["global"]["debug"].get(bool):
        ch = logging.StreamHandler()
        logger.setLevel(logging.DEBUG)
        ch.setFormatter(formatter)
        logger.addHandler(ch)
    else:
        file_path = get_data_dir() / logstr
        file_path.parent.mkdir(exist_ok=True)
        logfile_incr = name_of_next(file_path)
        fh = logging.FileHandler(str(logfile_incr))
        logger.setLevel(logging.INFO)
        fh.setFormatter(formatter)
        logger.addHandler(fh)


def make_nested_logger(nested: int) -> logging.Logger:
    logger = logging.getLogger("povm")
    print("Making nested logger", nested)

    if len(logger.handlers) != 1:
        raise RuntimeError("Too many handlers")

    handler: logging.Handler = logger.handlers[0]

    nested_logger: logging.Logger = logging.getLogger(f"povm-{nested:02}")
    formatter = logging.Formatter(
        f"%(asctime)s - %(levelname)s - {nested:02} - %(message)s",
        "%Y-%m-%d %H:%M:%S",
    )
    nested_logger.setLevel(logger.level)

    if isinstance(handler, logging.FileHandler):
        main_path = Path(handler.baseFilename).with_suffix("")
        main_path.mkdir(exist_ok=True)
        new_path = main_path / f"{nested:02}"
        new_path.mkdir(exist_ok=True)

        fh = logging.FileHandler(new_path / f"{main_path.stem}.log")
        fh.setLevel(logging.INFO)
        fh.setFormatter(formatter)
        nested_logger.addHandler(fh)

    elif isinstance(handler, logging.StreamHandler):
        ch = logging.StreamHandler()
        ch.setFormatter(formatter)
        nested_logger.addHandler(ch)

    else:
        raise RuntimeError(f"Logger handler {handler} is not of a known type!")

    return nested_logger
```
