## Supplementary Git Repository for "Inexpensive multi-patient respiratory monitoring system for helmet ventilation during COVID-19 pandemic": README.md

## Developer's guide


The key class is `Generator`, which implements a universal base for collecting and monitoring the data. It holds rolling windows of data, and can `analyze_as_needed`.

The `Collector` takes data from the single readings (run by `patient_*.py` code).

The `LocalGenerator` makes simulated data. The `RemoteGenerator` collects data via HTTP from a server serving data (probably from a `Collector`). `GeneratorThread` is the tool that `RemoteGenorator` runs to collect data.

The main collector has a built in thread that can be started with `.run(delay=0.2)`. Access to all properties should be protected with a `with self.lock`.

Key methods and properties:

* `analyze_as_needed()`: Run basic analysis, and more complex analysis only if needed.
    - `analyze()`: Full analysis of breaths.
    - `analyze_timeseries()`: Quick analysis that's easier to run often, makes volume (run by `analyze` too)
* `get_data()`: Copy in the remote/local datastream to internal cache
* `prepare(*, from_timestamp=None)`: Prepare a dict for transmission via json. Does *not* call `get_data()`.
* `close()`: Always close or use a context manager if running threads!

* `time`: The time array, with the most rececnt time as 0, with an adjustment based on `last_update`. Mostly for plotting.
* `realtime`: The actual time in seconds (arbitrary monotonic device clock)
* `timestamps`: Raw timestamps.
* `flow`: The raw flow data.
* `pressure`: The raw pressure data.
* `volume`: Computed volume.
* `breaths`: The last 30 computed breaths.
* `alarms`: A dictionary of alarms, each with first and last time, and extreme value.
* `cumulative`: A dictionary of resulting values, like RR, from the analysis.
* `cumulative_timestamps`: Timestamps on all cumulative keys for calculating staleness
* `cumulative_bywindow`: Dict of running averages for flow and pressure

You should close/leave context manger on any threaded collectors.
