## Supplementary Git Repository for "Inexpensive multi-patient respiratory monitoring system for helmet ventilation during COVID-19 pandemic": remote_generator.py

```
from __future__ import annotations

from processor.analysis import pressure_deglitch_smooth


import numpy as np
import zmq
from zmq.decorators import context, socket
import time
from datetime import datetime
from typing import Optional, Dict
import logging

from processor.generator import Status, Generator
from processor.gen_record import GenRecord
from processor.thread_base import ThreadBase


class RemoteThread(ThreadBase):
    def __init__(self, parent: RemoteGenerator):
        self.parent = parent
        self._address = self.parent.address
        self.status = Status.DISCON

        self._last_update: Optional[datetime] = None
        self._last_get: Optional[float] = None
        self.rotary_dict: Dict[str, Dict[str, float]] = {}
        self.mac: Optional[str] = None
        self.box_name: Optional[str] = None
        self.sid = 0
        self.last_interact: Optional[float] = None
        self.time_left: Optional[float] = None
        self.monotime: Optional[float] = None

        super().__init__(parent)

    def run(self) -> None:
        try:
            self._logging_run()
        except Exception:
            self.parent.logger.exception("Unexpected error in remote collection!")
            raise

    @context()
    @socket(zmq.SUB)
    def _logging_run(self, _ctx: zmq.Context, sub_socket: zmq.Socket) -> None:

        sub_socket.connect(self._address)
        sub_socket.subscribe(b"")

        while not self.parent.stop.is_set():
            number_events = sub_socket.poll(1 * 1000)
            for _ in range(number_events):
                self._last_update = datetime.now()
                root = sub_socket.recv_json()
                if "mac" in root:
                    with self.lock:
                        self.mac = root["mac"]
                if "name" in root:
                    with self.lock:
                        self.box_name = root["name"]
                if "sid" in root:
                    with self.lock:
                        self.sid = root["sid"]
                if "rotary" in root:
                    with self.lock:
                        self.rotary_dict = root["rotary"]
                if "last interact" in root:
                    with self.lock:
                        self.last_interact = root["last interact"]
                if "monotime" in root:
                    with self.lock:
                        self.monotime = root["monotime"]
                if "time left" in root:
                    with self.lock:
                        self.time_left = root["time left"]
                if "f" in root:
                    with self.lock:
                        self._time.inject_value(root["t"])
                        self._flow.inject_value(root["f"])
                        self._pressure.inject_value(root["p"])
                        self._last_get = time.monotonic()

                        if self.status == Status.DISCON:
                            self.parent.logger.info(
                                f"(Re)Connecting to {self._address} successful"
                            )
                            self.status = Status.OK

                if "C" in root:
                    with self.lock:
                        self._heat_time.inject_value(root["t"])
                        self._heat_temp.inject_value(root["C"])
                        self._heat_duty.inject_value(root["D"])

                if "CO2" in root:
                    with self.lock:
                        self._co2_time.inject_value(root["t"])
                        self._co2.inject_value(root["CO2"])
                        self._co2_temp.inject_value(root["Tp"])
                        self._humidity.inject_value(root["H"])

            if number_events == 0 and self.status != Status.DISCON:
                with self.lock:
                    self.status = Status.DISCON
                    self.parent.logger.info(f"Dropped connection to {self._address}")

    def access_collected_data(self) -> None:
        with self.parent.lock, self.lock:
            self.parent.last_update = self._last_update
            self.parent._last_get = self._last_get
            self.parent.last_interact = self.last_interact
            self.parent.current_monotonic = self.monotime
            self.parent.time_left = self.time_left

            newel = self.parent._time.new_elements(self._time)
            self.parent._time.inject_batch(self._time, newel)
            self.parent._flow.inject_batch(self._flow, newel)
            self.parent._pressure.inject_batch(self._pressure, newel)

            newel = self.parent._heat_time.new_elements(self._heat_time)
            self.parent._heat_time.inject_batch(self._heat_time, newel)
            self.parent._heat_temp.inject_batch(self._heat_temp, newel)
            self.parent._heat_duty.inject_batch(self._heat_duty, newel)

            newel = self.parent._co2_time.new_elements(self._co2_time)
            self.parent._co2_time.inject_batch(self._co2_time, newel)
            self.parent._co2.inject_batch(self._co2, newel)
            self.parent._co2_temp.inject_batch(self._co2_temp, newel)
            self.parent._humidity.inject_batch(self._humidity, newel)

            if self.status == Status.DISCON:
                self.parent.status = Status.DISCON

            # These log and perform (simple, please!) callbacks
            if self.mac is not None:
                self.parent.record.mac = self.mac
            if self.box_name is not None:
                self.parent.record.box_name = self.box_name
            if self.sid != 0:
                self.parent.record.sid = self.sid

            if len(self._time) > 0:
                self.parent._last_ts = self._time[-1]

            for k, v in self.rotary_dict.items():
                if k in self.parent.rotary:
                    if self.parent.rotary[k].value != v["value"]:
                        self.parent.rotary[k].value = v["value"]
                        self.parent.logger.info(f"rotary: {k} set to {v['value']}")


class RemoteGenerator(Generator):
    def __init__(
        self,
        *,
        address: str = "tcp://127.0.0.1:8100",
        logger: logging.Logger,
        gen_record: GenRecord = None,
    ):
        super().__init__(logger=logger, gen_record=gen_record)
        self._address = address

        self.status = Status.DISCON
        self._last_ts: int = 0

        # Last interaction timestamp (only set on remote generators)
        self.last_interact: Optional[float] = None

        # Time left on alarm silence (only on remote generators)
        self.time_left: Optional[float] = None

        # Current monotonic time from last access
        self.current_monotonic: Optional[float] = None

        self._remote_thread: Optional[RemoteThread] = None

    def run(self) -> None:
        super().run()
        self._remote_thread = RemoteThread(self)
        self._remote_thread.start()

    def _get_data(self) -> None:
        if self._remote_thread is not None:
            self._remote_thread.access_collected_data()

        if self._debug:
            with self.lock:
                if np.any(self._time[:-1] > self._time[1:]):
                    self.logger.error("Time array is not sorted!")

    def _set_alarms(self) -> None:
        if self.time_left is not None and self.time_left > 0:
            self.status = Status.ALERT_SILENT if self.alarms else Status.SILENT
        else:
            super()._set_alarms()

    @property
    def address(self) -> str:
        return self._address

    @property
    def pressure(self) -> np.ndarray:
        return pressure_deglitch_smooth(np.asarray(self._pressure))

    def close(self) -> None:
        super().close()
        if self._remote_thread is not None:
            self._remote_thread.join()
```
