## Supplementary Git Repository for "Inexpensive multi-patient respiratory monitoring system for helmet ventilation during COVID-19 pandemic": rolling.py

```
from __future__ import annotations

import numpy as np

from typing import Union, List


class Rolling:
    def __init__(self, init=None, *, window_size: int, dtype=None):
        if dtype is None:
            if init:
                dtype = np.asarray(init).dtype
            else:
                dtype = np.double

        self._values = np.empty((window_size * 2,), dtype=dtype)
        self._start = 0
        self._window_size = window_size
        self._current_size = 0

        if init is not None:
            self.inject(init)

    def clear(self) -> None:
        self._start = 0
        self._current_size = 0

    @property
    def window_size(self) -> int:
        return self._window_size

    def inject_value(self, value: float) -> None:
        """
        High performance version of inject.
        """

        fill_start = (
            self._start
            if self._current_size == self._window_size
            else self._current_size
        )

        self._values[fill_start] = value
        self._values[fill_start + self._window_size] = value

        if self._current_size < self._window_size:
            self._current_size += 1
        else:
            self._start += 1
            self._start %= self._window_size

    def inject(self, values: Union[List[float], np.ndarray]) -> None:
        """
        Add a value or an array of values to the end of the rolling buffer.  It
        is currently invalid to input a 2D array.

        Single values can be injected much more quickly with inject_value.
        """

        # Make sure input is an array, truncate if larger than rolling buffer
        values = np.asarray(values)

        if values.size > self._window_size:
            values = values[-self._window_size :]
        if values.ndim > 1:
            raise RuntimeError("Only 0D and 1D supported")

        fill_start = (
            self._start
            if self._current_size == self._window_size
            else self._current_size
        )

        # Filling the first copy is always valid
        self._values[fill_start : fill_start + values.size] = values

        # If we go off the end, wrap around
        overlap = fill_start + values.size - self._window_size
        if overlap > 0:
            self._values[self._window_size + fill_start :] = values[
                : self._window_size - fill_start
            ]
            self._values[:overlap] = values[self._window_size - fill_start :]
        else:
            self._values[
                fill_start
                + self._window_size : fill_start
                + self._window_size
                + values.size
            ] = values

        # Update current size and starting position
        self._current_size = min(self._current_size + values.size, self._window_size)
        self._start = (
            fill_start + values.size - (self._current_size - self._window_size)
        ) % self._window_size

    def explain(self) -> str:
        """Gives a nice text display of the internal structure"""
        mask = np.ones_like(self._values, dtype=np.bool)
        mask[: self._current_size] = False
        mask[self._window_size : self._window_size + self._current_size] = False
        data = np.ma.masked_array(data=self._values, mask=mask)
        return f"{data} -> {self}"

    def __repr__(self) -> str:
        r = repr(np.asarray(self))
        start = "Rolling(" + ("\n" if "\n" in r else "")
        return start + r[6:-1] + f", window_size={self._window_size})"

    def __str__(self) -> str:
        return str(np.array(self))

    def __eq__(self, other: object) -> bool:
        if isinstance(other, Rolling):
            return len(self) == len(other) and np.allclose(self, other)
        else:
            return NotImplemented

    def __array__(self) -> np.ndarray:
        data = self._values[self._start : self._start + self._current_size]
        data.flags.writeable = False
        return data

    def __getitem__(self, arg):
        return np.asarray(self).__getitem__(arg)

    def __len__(self) -> int:
        return self._current_size

    def inject_batch(self, other: Rolling, newel: int) -> None:
        """
        Inject "newel" items from other to self.
        """
        if newel:
            self.inject(other[-newel:])

    def new_elements(self, addition: Union[Rolling, np.ndarry]) -> int:
        """
        Given two sorted arrays, find the number of elements in the second array
        that are past the end of the first array.

        WARNING: Be sure to protect when this returns 0 or use inject_batch!
        """

        if len(self) < 1:
            return len(addition)

        final_value = self[-1]
        ind = np.searchsorted(addition, final_value)
        if ind == len(addition):
            return 0
        else:
            return len(addition) - ind - (final_value == addition[ind])
```
