## Supplementary Git Repository for "Inexpensive multi-patient respiratory monitoring system for helmet ventilation during COVID-19 pandemic": rotary.py

```
#!/usr/bin/env python3
from __future__ import annotations

from typing import List, Dict, Any, ValuesView, TypeVar, Iterable, ItemsView

from processor.setting import Setting
import threading


T = TypeVar("T", bound="LocalRotary")


class LocalRotary:
    def __init__(self, config: Dict[str, Setting]):
        self.config = config
        self._alarms: Dict[str, Any] = {}

        # Will be set by changing a value, unset by access (to_dict)
        self._changed = threading.Event()

        # Cached for simplicity (dicts are ordered)
        self._items: List[str] = list(self.config.keys())

    def changed(self):
        """
        This should always be called when an item in the rotary is changed
        """

        self._changed.set()

    def to_dict(self) -> Dict[str, Any]:
        "Convert config to dict"
        return {
            k: v.to_dict()
            for k, v in self.config.items()
            if k not in ["C02 Setting", "Current Setting", "Reset Setting"]
        }

    @property
    def alarms(self) -> Dict[str, Dict[str, float]]:
        return self._alarms

    @alarms.setter
    def alarms(self, item: Dict[str, Dict[str, float]]):
        self._alarms = item

    def __getitem__(self, item: str):
        return self.config[item]

    def values(self) -> ValuesView[Setting]:
        return self.config.values()

    def items(self) -> ItemsView[str, Setting]:
        return self.config.items()

    def __repr__(self) -> str:
        out = f"{self.__class__.__name__}(\n"
        for key, value in self.config.items():
            out += f"  {key} : {value}\n"
        return out + "\n)"

    def __enter__(self: T) -> T:
        return self

    def __exit__(self, *args) -> None:
        return None

    def __contains__(self, key: str):
        return key in self.config

    def __iter__(self) -> Iterable[str]:
        return iter(self.config)

    def external_update(self) -> None:
        "Update the display after a live setting (CurrentSetting) is changed externally"
        pass

    def time_left(self) -> float:
        "Amount of time left on timer"
        raise NotImplementedError()

    def last_interaction(self) -> float:
        "Timestamp of last interaction"
        raise NotImplementedError()
```
