## Supplementary Git Repository for "Inexpensive multi-patient respiratory monitoring system for helmet ventilation during COVID-19 pandemic": saver.py

```
from __future__ import annotations

import time
from pathlib import Path
from typing import TYPE_CHECKING, Dict, List, NamedTuple, Sequence, Iterator
from datetime import datetime
import numpy as np
import json

if TYPE_CHECKING:
    from processor.generator import Generator


class Saver:
    def __init__(self, gen: Generator, filepath: Path, save_every: float):
        self.parent = gen
        self.filepath = filepath
        self.file = open(filepath, "a")
        self.last_save = time.monotonic()
        self.save_every = save_every

    def __bool__(self) -> bool:
        return time.monotonic() - self.last_save > self.save_every

    def enter(self) -> None:
        self.file.close()
        self.file = open(self.filepath, "a")
        self.header()

    def save(self) -> None:
        self.file.flush()
        self.last_save = time.monotonic()

    def close(self) -> None:
        self.file.close()

    def header(self) -> None:
        self.file.write(f"# Nursetime: {datetime.now().isoformat()}\n")


class FieldInfo(NamedTuple):
    id: str
    name: str
    fmt: str = ""


class CSVSaverTS(Saver):
    def __init__(
        self,
        fields: Sequence[FieldInfo],
        gen: Generator,
        filepath: Path,
        save_every: float,
    ):
        super().__init__(gen, filepath, save_every)
        self._last_timestamp = 0
        self.fields = fields

    def header(self) -> None:
        super().header()
        values = (f.id for f in self.fields)
        print(*values, sep=", ", file=self.file)

    def save(self) -> None:

        time_name = self.fields[0].name
        parent_time = getattr(self.parent, time_name)

        if len(parent_time) < 1:
            return

        ind = np.searchsorted(parent_time, self._last_timestamp, side="right")
        slices: Iterator[np.ndarray] = (
            getattr(self.parent, f.name)[ind:] for f in self.fields
        )

        for items in zip(*slices):
            item_format = zip(items, (f.fmt for f in self.fields))
            values = (format(item, fmt) for item, fmt in item_format)
            print(*values, sep=", ", file=self.file)

        self._last_timestamp = parent_time[-1]
        super().save()


class CSVSaverCML(Saver):
    def save(self) -> None:
        self.file.write(json.dumps(self.parent.cumulative))
        self.file.write("\n")
        super().save()


class JSONSSaverBreaths(Saver):
    def __init__(self, gen: Generator, filepath: Path):
        super().__init__(gen, filepath, 0.0)

    def __bool__(self) -> bool:
        return False

    def save_breaths(self, breaths: List[Dict[str, float]]) -> None:
        for breath in breaths:
            self.file.write(json.dumps(breath))
            self.file.write("\n")
        self.save()

    def header(self) -> None:
        pass
```
