## Supplementary Git Repository for "Inexpensive multi-patient respiratory monitoring system for helmet ventilation during COVID-19 pandemic": setting.py

```
#!/usr/bin/env python3

# mypy: disallow_untyped_defs
# mypy: disallow_incomplete_defs

import threading
from typing import List, Any, Dict, Union, Optional
import abc

Number = Union[float, int]


class Setting(abc.ABC):
    STATIC_UPPER = True

    def __init__(
        self,
        *,
        name: str,
        unit: str = None,
        lcd_name: str = None,
        zero: str = None,
        rate: int = 1,
    ):
        self._name = name
        self._lcd_name: Optional[str] = lcd_name or name
        self.unit = unit
        self._lock = threading.Lock()
        self._value: Any = None
        self._original_value: Any = None
        self._zero = zero

        # Rate at which settings change
        self._rate = rate

        # Buffer for computing above rate
        self._buffer = 0

    @property
    def name(self) -> str:
        return self._name

    @property
    def lcd_name(self) -> str:
        return self._name if self._lcd_name is None else self._lcd_name

    @property
    def value(self) -> Any:
        with self._lock:
            return self._value

    @value.setter
    def value(self, val: Any) -> None:
        # Only used to set values remotely
        with self._lock:
            self._value = val

    def reset(self) -> None:
        with self._lock:
            self._value = self._original_value

    @property
    def default(self) -> Any:
        """
        Override in subclass if there's a special way to save this setting
        """
        with self._lock:
            return self._value

    @default.setter
    def default(self, value: Any) -> None:
        with self._lock:
            self._value = value

    def __str__(self) -> str:
        if self._zero is not None and self._value == 0:
            return str(self._zero)
        else:
            unit = "" if self.unit is None else f" {self.unit}"
            return f"{self.value}{unit}"

    def __repr__(self) -> str:
        return f"{self.__class__.__name__}({self}, name={self.name})"

    def to_dict(self) -> Dict[str, Union[str, float]]:
        return {"name": self.name, "value": self.value}

    def __format__(self, format_spec: str) -> str:
        return str(self).__format__(format_spec)

    def up(self) -> None:
        self._buffer += 1
        self._buffer %= self._rate
        if self._buffer == 0:
            self._up_()

    def down(self) -> None:
        self._buffer -= 1
        self._buffer %= self._rate
        if self._buffer == 0:
            self._down_()

    @abc.abstractmethod
    def _up_(self) -> None:
        pass

    @abc.abstractmethod
    def _down_(self) -> None:
        pass

    def active(self) -> None:
        """
        Called when this becomes the active item.
        """


class DisplaySetting(Setting):
    def __init__(self, *, name: str, lcd_name: str = None):
        super().__init__(name=name, lcd_name=lcd_name)

    def _up_(self) -> None:
        pass

    def _down_(self) -> None:
        pass


class IncrSetting(Setting):
    def __init__(
        self,
        default: Number,
        *,
        min: Number,
        max: Number,
        incr: Number,
        name: str,
        unit: str = None,
        lcd_name: str = None,
        zero: str = None,
        rate: int = 1,
    ):
        "Note: incr should be a nice floating point number"

        super().__init__(unit=unit, name=name, lcd_name=lcd_name, zero=zero, rate=rate)

        self._min = min
        self._max = max
        self._value = default
        self._original_value = default
        self._incr = incr

    def _up_(self) -> None:
        "Return true if not at limit"
        with self._lock:
            if self._value < self._max:
                self._value += self._incr

    def _down_(self) -> None:
        "Return true if not at limit"
        with self._lock:
            if self._value > self._min:
                self._value -= self._incr


class SelectionSetting(Setting):
    def __init__(
        self,
        default: int,
        listing: List[Any],
        *,
        name: str,
        unit: str = None,
        lcd_name: str = None,
        zero: str = None,
        rate: int = 2,
    ):

        assert (
            0 <= default < len(listing)
        ), "Default must be an index into the list given"

        super().__init__(unit=unit, name=name, lcd_name=lcd_name, zero=zero, rate=rate)

        self._value: int = default
        self._original_value: int = default
        self._listing: List[Any] = listing
        self._zero: Optional[str] = zero

    def _up_(self) -> None:
        "Return true if not at limit"
        with self._lock:
            if self._value < len(self._listing) - 1:
                self._value += 1

    def _down_(self) -> None:
        "Return true if not at limit"
        with self._lock:
            if self._value > 0:
                self._value -= 1

    def __len__(self) -> int:
        return len(self._listing)

    @property
    def value(self) -> Any:
        with self._lock:
            return self._listing[self._value]

    @value.setter
    def value(self, val: float) -> None:
        # Only used to set values remotely
        with self._lock:
            self._value = self._listing.index(val)
```
