## Supplementary Git Repository for "Inexpensive multi-patient respiratory monitoring system for helmet ventilation during COVID-19 pandemic": settings.py

```
#!/usr/bin/env python3
from __future__ import annotations

from processor.setting import IncrSetting, SelectionSetting, Setting
from processor.display_settings import (
    AdvancedSetting,
    CurrentSetting,
    ResetSetting,
    CO2Setting,
)
from processor.config import config

from typing import Dict, Tuple, Optional
from confuse import ConfigView
from pprint import pprint


def get_setting(c: ConfigView) -> Tuple[Optional[int], Setting]:
    type_name = c["type"].get()
    order = c["order"].get(int) if "order" in c else None
    if type_name == "Incr":
        return (
            order,
            IncrSetting(
                default=c["default"].as_number(),
                min=c["min"].as_number(),
                max=c["max"].as_number(),
                incr=c["incr"].as_number(),
                name=c["name"].get(),
                unit=c["unit"].get() if "unit" in c else None,
                rate=c["rate"].get(int) if "rate" in c else 1,
                zero=c["zero"].get() if "zero" in c else None,
                lcd_name=c["lcd_name"].get() if "lcd_name" in c else None,
            ),
        )
    elif type_name == "Selection":
        return (
            order,
            SelectionSetting(
                default=c["default"].get(int),
                listing=[v.as_number() for v in c["items"]],
                name=c["name"].get(),
                unit=c["unit"].get() if "unit" in c else None,
                zero=c["zero"].get() if "zero" in c else None,
                rate=c["rate"].get(int) if "rate" in c else 2,
                lcd_name=c["lcd_name"].get() if "lcd_name" in c else None,
            ),
        )
    elif type_name == "Current":
        return (
            order,
            CurrentSetting(name="Current:"),
        )
    elif type_name == "CO2":
        return (
            order,
            CO2Setting(name="Current:"),
        )
    elif type_name == "Reset":
        return (
            order,
            ResetSetting(
                name=c["name"].get(),
                rate=c["rate"].get(int) if "rate" in c else 1,
            ),
        )
    elif type_name == "Advanced":
        return order, AdvancedSetting(rate=c["rate"].get(int) if "rate" in c else 2)
    else:
        raise RuntimeError(
            f"Invalid type {type_name!r}, needs to be defined in setting.py/settings.py"
        )


def reorder(indict: Dict[str, Setting], ordering: Dict[str, int]) -> Dict[str, Setting]:
    master: Dict[str, Setting] = {}

    above_values = sorted(v for v in set(ordering.values()) if v >= 0)
    below_values = sorted(v for v in set(ordering.values()) if v < 0)

    # Values that have a positive order go first (0, then 1, etc.)
    for value in above_values:
        keys = {k for k, v in ordering.items() if v == value}
        master.update({k: v for k, v in indict.items() if k in keys})

    # Values that have no order go in the middle
    master.update({k: v for k, v in indict.items() if k not in ordering})

    # Values with a negative order go at the end (-3, -2, -1)
    for value in below_values:
        keys = {k for k, v in ordering.items() if v == value}
        master.update({k: v for k, v in indict.items() if k in keys})

    return master


def get_live_settings() -> Dict[str, Setting]:
    "Get the live version of the configuration dictionary"

    d: Dict[str, Setting] = {}
    ordering: Dict[str, int] = {}
    all_settings = [config["rotary-live"], config["rotary"]]
    for settings in all_settings:
        for setting in settings:
            order, d[setting] = get_setting(settings[setting])
            if order is not None:
                ordering[setting] = order

    return reorder(d, ordering)


def get_remote_settings() -> Dict[str, Setting]:
    "Get the non-live version of the configuration dictionary"
    d: Dict[str, Setting] = {}
    ordering: Dict[str, int] = {}
    all_settings = [config["rotary"]]
    for settings in all_settings:
        for setting in settings:
            order, d[setting] = get_setting(settings[setting])
            if order is not None:
                ordering[setting] = order

    return reorder(d, ordering)


if __name__ == "__main__":
    print("Live ----/n")
    pprint(get_live_settings())
    print("/nRemote ----/n")
    pprint(get_remote_settings())
```
