## Supplementary Git Repository for "Inexpensive multi-patient respiratory monitoring system for helmet ventilation during COVID-19 pandemic": test_rolling.py

```
from numpy.testing import assert_allclose
import numpy as np

from processor.rolling import Rolling


def test_rolling_single():
    r = Rolling(window_size=3)

    assert len(r) == 0
    assert r._start == 0
    assert_allclose(np.asarray(r), [])

    r.inject(1)
    assert len(r) == 1
    assert r._start == 0
    assert_allclose(np.asarray(r), [1.0])

    r.inject(2)
    assert len(r) == 2
    assert r._start == 0
    assert_allclose(np.asarray(r), [1.0, 2.0])

    r.inject(3)
    assert len(r) == 3
    assert r._start == 0
    assert_allclose(np.asarray(r), [1.0, 2.0, 3.0])

    r.inject(4)
    assert len(r) == 3
    assert r._start == 1
    assert_allclose(np.asarray(r), [2.0, 3.0, 4.0])

    r.inject(5)
    assert len(r) == 3
    assert r._start == 2
    assert_allclose(np.asarray(r), [3.0, 4.0, 5.0])

    r.inject(6)
    assert len(r) == 3
    assert r._start == 0
    assert_allclose(np.asarray(r), [4.0, 5.0, 6.0])

    r.inject(7)
    assert len(r) == 3
    assert r._start == 1
    assert_allclose(np.asarray(r), [5.0, 6.0, 7.0])


def test_rolling_single_value():
    r = Rolling(window_size=3)

    assert len(r) == 0
    assert r._start == 0
    assert_allclose(np.asarray(r), [])

    r.inject_value(1)
    assert len(r) == 1
    assert r._start == 0
    assert_allclose(np.asarray(r), [1.0])

    r.inject_value(2)
    assert len(r) == 2
    assert r._start == 0
    assert_allclose(np.asarray(r), [1.0, 2.0])

    r.inject_value(3)
    assert len(r) == 3
    assert r._start == 0
    assert_allclose(np.asarray(r), [1.0, 2.0, 3.0])

    r.inject_value(4)
    assert len(r) == 3
    assert r._start == 1
    assert_allclose(np.asarray(r), [2.0, 3.0, 4.0])

    r.inject_value(5)
    assert len(r) == 3
    assert r._start == 2
    assert_allclose(np.asarray(r), [3.0, 4.0, 5.0])

    r.inject_value(6)
    assert len(r) == 3
    assert r._start == 0
    assert_allclose(np.asarray(r), [4.0, 5.0, 6.0])

    r.inject_value(7)
    assert len(r) == 3
    assert r._start == 1
    assert_allclose(np.asarray(r), [5.0, 6.0, 7.0])


def test_rolling_double():
    r = Rolling(window_size=3)

    r.inject([1, 2])
    assert len(r) == 2
    assert r._start == 0
    assert_allclose(np.asarray(r), [1.0, 2.0])

    r.inject([3, 4])
    assert len(r) == 3
    assert r._start == 1
    assert_allclose(np.asarray(r), [2.0, 3.0, 4.0])

    r.inject([5, 6])
    assert len(r) == 3
    assert r._start == 0
    assert_allclose(np.asarray(r), [4.0, 5.0, 6.0])


def test_rolling_full():
    r = Rolling(window_size=3)

    r.inject([1, 2, 3])
    assert len(r) == 3
    assert r._start == 0
    assert_allclose(np.asarray(r), [1.0, 2.0, 3.0])

    r.inject([4, 5, 6])
    assert len(r) == 3
    assert r._start == 0
    assert_allclose(np.asarray(r), [4.0, 5.0, 6.0])


def test_rolling_overlap():
    r = Rolling(window_size=3)

    r.inject([1, 2, 3, 4])
    assert len(r) == 3
    assert r._start == 0
    assert_allclose(np.asarray(r), [2.0, 3.0, 4.0])

    r.inject([5, 6, 7, 8])
    assert len(r) == 3
    assert r._start == 0
    assert_allclose(np.asarray(r), [6.0, 7.0, 8.0])


def test_getitem():
    r = Rolling(window_size=3)
    r.inject([1, 2, 3])
    assert r[0] == 1.0
    assert_allclose(r[1:], [2.0, 3.0])


def test_new_elements():
    r = Rolling([1, 2, 3, 4, 5], window_size=5)
    assert r.new_elements([4, 5, 6, 7]) == 2
    assert r.new_elements([5, 6, 7]) == 2
    assert r.new_elements([4, 5]) == 0
    assert r.new_elements([2, 3]) <= 0
    assert r.new_elements([8, 9]) == 2
    assert r.new_elements([3, 8, 9]) == 2

    r = Rolling(window_size=5)
    assert r.new_elements([3, 8, 9]) == 3


def test_batch():
    r = Rolling([1, 2, 3, 4, 5], window_size=6)
    arr = np.array([5, 6, 7, 8])
    newel = r.new_elements(arr)
    assert newel == 3

    r.inject_batch(arr, newel)

    assert_allclose(r[:], [3, 4, 5, 6, 7, 8])
```
