## Supplementary Git Repository for "Inexpensive multi-patient respiratory monitoring system for helmet ventilation during COVID-19 pandemic": thread_base.py

```
#!/usr/bin/env python3
from __future__ import annotations

import threading

import numpy as np

from processor.generator import Generator
from processor.rolling import Rolling


class ThreadBase(threading.Thread):
    def __init__(self, parent: Generator):
        self._time = Rolling(window_size=parent.window_size, dtype=np.int64)
        self._flow = Rolling(window_size=parent.window_size)
        self._pressure = Rolling(window_size=parent.window_size)

        self._heat_time = Rolling(window_size=parent.extras_window_size, dtype=np.int64)
        self._heat_temp = Rolling(window_size=parent.extras_window_size)
        self._heat_duty = Rolling(window_size=parent.extras_window_size)

        self._co2_time = Rolling(window_size=parent.extras_window_size, dtype=np.int64)
        self._co2 = Rolling(window_size=parent.extras_window_size)
        self._co2_temp = Rolling(window_size=parent.extras_window_size)
        self._humidity = Rolling(window_size=parent.extras_window_size)

        self.lock = threading.Lock()

        super().__init__()
```
