## Supplementary Git Repository for "Inexpensive multi-patient respiratory monitoring system for helmet ventilation during COVID-19 pandemic": version.py

```
from __future__ import annotations

from functools import lru_cache
from pathlib import Path
import subprocess

DIR = Path(__file__).parent.resolve()


@lru_cache(1)
def get_version() -> str:
    try:
        return (
            subprocess.check_output(["git", "describe", "--tags"], cwd=DIR, text=True)
            .strip()
            .lstrip("v")
        )
    except subprocess.CalledProcessError:
        return ""
```
