## Supplementary Git Repository for "Inexpensive multi-patient respiratory monitoring system for helmet ventilation during COVID-19 pandemic": README.md

# princeton-penn-flowmeter
Software for patient and nurse stations for multi-patient ventilator
monitoring.

The hardware at the patient includes two sensors to measure flow and pressure
mounted on a flow block and equipped with a heating circuit to reduce condensation,
a single board computer (SBC, in this case a Raspberry Pi model 4b/4GB), a 2x20
character LCD display with RBG backlight, a rotary encoder and a piezo buzzer.
The sensor time series data from up to 20 patient boxes are transmitted to a nurse
monitoring station, where a graphical GUI presents an aggregate view of the
time series. The nurse monitoring station can either be an SBC or a more
standard laptop/desktop machine. Analysis code runs on both the patient box
and on the nurse monitoring station to derive quantities for both alarms
and display.

The software is written in Python and PyQt for portability.

# Installation

#### On a local machine:

```bash
git clone https://github.com/Princeton-Penn-Vents/princeton-penn-flowmeter
cd princeton-penn-flowmeter
conda env create
conda activate flowmeter
```

(and git pull, then conda env update to update)

### On a Raspberry Pi running Raspbian:

Boot up a fresh copy of Raspbian. At the setup screen, click next to proceed.
Select United States, American English, and New York for the country and
timezone settings. Set a password.  Check the box saying there's a black
border (there is one). You can select a network if you want to connect it
to the internet. Press "next" when asked about updating if you connected
to the internet. Press OK then restart.

Open the settings (Raspberry menu -> `Preferences` -> `Raspberry Pi Configuration`).
Rename hostname, as needed.
In the display tab, select "Disable" for `Screen Blanking`.  For a patient box,
select "Disable" for `Overscan`.

Select the Interfaces tab to enable hardware interfaces.

For a patient box, you need: I2C enabled, SPI enabled.  These are also set in config.txt.
Optional:  SSH enabled.
Reboot required.

Click on terminal icon and execute:

```bash
git clone https://github.com/Princeton-Penn-Vents/princeton-penn-flowmeter
# if a username is requested, then a typo has been made in the name, try again carefully

cd princeton-penn-flowmeter
sudo apt update
sudo apt upgrade

sudo apt install python3-pyqt5 python3-zmq # Required on the base system, included in NOOBs
sudo apt install python3-scipy
sudo python3 -m pip install pyqtgraph pyzmq confuse zeroconf
sudo python3 -m pip install "setuptools>=44" getmac setuptools_scm[toml]
```

For development, these quality-of-life additions help:

```bash
sudo apt install vim htop
python3 -m pip install black pytest mypy
```

<details><summary>Networking for the nurse box: (click to expand)</summary>

Automatic discovery make make this no longer required; even the default auto-IP selection
should work.

This should only be done for *one* nurse box, even if you connect two nurse stations to a network,
only one of them should have the following setup:

Nurse box networking (assuming debian family, like Ubuntu, when naming specifics):

```bash
sudo apt install isc-dhcp-server
sudo cp config/isc-dhcp-server /etc/default/     # OVERWRITES
sudo cp config/dhcpd.conf /etc/dhcp/             # OVERWRITES
sudo cp config/10-eth0-povm.config /etc/network/interfaces.d/
sudo systemctl enable isc-dhcp-server
```

The new nurse station IP (192.168.3.3) will come up automatically and the DHCP
server will start on next computer restart.

</details>

<details><summary>Additional information: (click to expand)</summary>

Edit the file `/etc/default/isc-dhcp-server` and set `eth0` as the interface to serve:

```
INTERFACESv4="eth0"
```

Edit the file `/etc/dhcp/dhcpd.conf` to include these lines:

```
option domain-name "local";
option domain-name-servers ns1.local, ns2.local;
default-lease-time 6000;
max-lease-time 72000;
ddns-update-style none;
authoritative;
subnet 192.168.3.0 netmask 255.255.255.0 {
  range 192.168.3.20 192.168.3.250;
  option subnet-mask 255.255.255.0;
  option routers 192.168.3.1;
  option broadcast-address 192.168.3.255;
  option domain-name-servers 192.168.3.2;
  option domain-search "local";
}
```

Finally, edit the `/etc/network/interface/` file or add a file in `/etc/network/interface.d/` with these lines:

```
iface eth0 inet static
    address 192.168.3.2/24
    gateway 192.168.3.1
```

Finally, bring up your interface and start/enable the service:

```bash
sudo ifup eth0
sudo systemctl enable isc-dhcp-server
sudo systemctl start isc-dhcp-server
```

Connections to the outside world / internet can be done through wireless, which
is unaffected by the above settings.

</details>


For patient box setup:

```bash
cd princeton-penn-flowmeter
sudo cp config/config.txt /boot/config.txt
sudo cp config/patientdevice.service /lib/systemd/system
sudo cp config/patientloop.service /lib/systemd/system
sudo systemctl enable pigpiod
sudo systemctl enable patientdevice
sudo systemctl enable patientloop
```
Now, you have all the code and automatic startup setup.
On reboot, all patient processes will be launched automatically.

<details><summary>Additional information: (click to expand)</summary>

To operate patient box manually:

To execute the readout, click on a terminal icon, then in the shell:

```
cd princeton-penn-flowmeter
./stopall
python3 ./device_loop.py —-file test.out
```

This will start printing temperature settings every second to the screen - will
thermalize at 40C with the temperature servo, and it will be recording pressure
(in ADC counts from 0 to 4095) and flow rate (signed integer 0 to 32,767) and
the time in milliseconds (with a precision of microseconds).  The RPi4 does not
operate a clock when powered down, but rather begins from where it left off
when shutdown.

You can `^C` at any time and look at the data. This will print it to the screen:

```bash
cat test0000.out
```

where the 0000 will increment every time the process is restarted through a
local directory scan.  Logging of data is by default in `./device_log/`.

Operation of the LCD display and rotary and to serve data to the nurseguii from
the device_loop, one can run locally:

```
cd princeton-penn-flowmeter
python3 ./patient_loop.py
```

The local IP address of eth0 can be found using ifconfig.  A remote nurse
station can receive the data from the `patient_loop` by starting:

```
cd princeton-penn-flowmeter
python3 ./nursegui.py -n 1 --port 8100 --ip <patientboxIP>
```

For reference, instead of a full copy of config.txt, the changes that need to be made to `/boot/config.txt` are specifically:

```
dtparam=i2c_arm=on,baudrate=200000
dtparam=spi=on
```

These lines are added:

```
dtoverlay=i2c1,pins_2_3
dtoverlay=i2c6,pins_22_23
```

#### pigpio cleanup, if needed:

```bash
sudo killall pigpiod
```

</details>

<details><summary>Setup a Windows monitoring station</summary>

From a fresh install, setup the machine with Windows. Keep the machine *off*
the internet during the setup procedure! Microsoft will *require* an account
otherwise.

Install all updates; you should be on at least Windows 10 version 1909. I
recommend upgraded the built-in Edge browswer to the Chromium based Edge
browser, as well.

Download the Windows package manager, available in releases here:
<https://github.com/microsoft/winget-cli> Install it.

Install the following from cmd/PowerShell (your choice, but run as administrator):

```
winget install Git.Git
winget install Microsoft.WindowsTerminal
winget install Anaconda.Miniconda3
```


You have a conda terminal in your start menu now. From that terminal, download the
repository and setup:

```cmd
git clone git://github.com/Princeton-Penn-Vents/princeton-penn-flowmeter.git
cd princeton-penn-flowmeter
conda env create
explorer .
```

Finally, and assuming all paths are "normal", copy/drag the patientmonitor.bat to
the desktop (I also drag out the terminals that are useful there, too).

</details>

---

## Common test procedures:

#### Start up a batch of local simulations

```bash
./nursegui.py -n 20 --sim --debug --window
```

(You can always click the `+` to add a device - and you can even add a new sim if you started with `--sim`)

#### Start simulations and stream through sockets

Terminal 1:

```bash
./patientsim.py --port 8100 -n 20
```

Terminal 2 (same computer or on a local network):

```bash
./nursegui.py --debug --window
```

#### Replay a log file

Add `--ff <timestamp>` to fast-forward to a timestamp before starting playback to the socket.

```bash
# Terminal 1
./device_json_to_socket.py data/20200422_helmet.out

# Terminal 2
./patientgui.py

# Terminal 3
./nursegui.py --debug --window
```

---

# Acknowledgements

This project is supported by Princeton University and by NSF Cooperative Agreement OAC-1836650.
