## Supplementary Git Repository for "Inexpensive multi-patient respiratory monitoring system for helmet ventilation during COVID-19 pandemic": setup.cfg

```
[mypy]
mypy_path = .stubs
warn_unused_configs = True
pretty = True
python_version = 3.7
warn_unused_ignores = True
check_untyped_defs = True
files=**/*.py

[mypy-numpy.*]
ignore_missing_imports = True

[mypy-scipy.*]
ignore_missing_imports = True

[mypy-matplotlib.*]
ignore_missing_imports = True

[mypy-PyQt5.*]
ignore_missing_imports = True

[mypy-pyqtgraph.*]
ignore_missing_imports = True

[mypy-ifaddr.*]
ignore_missing_imports = True

# [mypy-processor.*]
# disallow_untyped_defs = True
# disallow_incomplete_defs = True

# [mypy-patient.*]
# disallow_untyped_defs = True
# disallow_incomplete_defs = True


[flake8]
ignore = E203, E231, E501, E722, W503, E402, B950
select = C,E,F,W,B,B9
```
