## Supplementary Git Repository for "Inexpensive multi-patient respiratory monitoring system for helmet ventilation during COVID-19 pandemic": aggregate.py

```
#!/usr/bin/env python3
import json


def aggregate_for_nursing(time_stamp, jsons):

    for i, j_str in enumerate(jsons):
        j_info = json.loads(j_str)
        if i == 0:
            nurse_info = {
                "version": j_info["v"],
                "time": time_stamp,
                "alarms": {},
                "data": {"timestamps": [], "flows": [], "pressures": []},
            }
        nurse_info["data"]["timestamps"].append(j_info["t"])
        nurse_info["data"]["pressures"].append(j_info["P"])
        nurse_info["data"]["flows"].append(j_info["F"])

    return json.dumps(nurse_info).encode("ascii")


if __name__ == "__main__":
    from sim.ventsim import VentSim

    from datetime import datetime

    now_time = int(1000 * datetime.now().timestamp())

    sim = VentSim(now_time, 1000000)
    jsons = []
    for _ in range(3000):
        d = sim.get_next()
        jsons.append(json.dumps(d).encode("ascii"))
    output = aggregate_for_nursing(now_time, jsons)
    print(len(output))
    print(json.loads(output))
```
