## Supplementary Git Repository for "Inexpensive multi-patient respiratory monitoring system for helmet ventilation during COVID-19 pandemic": sim_configs.yml

```
nominal_breather:
  - sample_length: 20.0
  - breath_interval: 5000.0
  - max_flow: 15.0
  - tidal_volume:
    - mean: 0.6
    - sigma: 0.1
  - peep: 4
  - compliance_func: "constant_compliance"
  - starting_volume: 0.0
  - breath_variation: 300.0
  - max_breath_interval: 9000.0
  - measurement_error_flow: 1.0
  - measurement_error_pressure: 0.1
  - average_flow: 60.0

nominal_nonbreather:
  - sample_length: 20.0
  - breath_interval: 5000.0
  - max_flow: 15.0
  - tidal_volume:
    - mean: 0.5
    - sigma: 0.1
  - peep: 4
  - compliance_func: "constant_compliance"
  - starting_volume: 0.0
  - breath_variation: 50.0
  - max_breath_interval: 9000.0
  - measurement_error_flow: 1.0
  - measurement_error_pressure: 0.1
  - average_flow: 60.0

alarm_nonbreather:
  - sample_length: 20.0
  - breath_interval: 5000.0
  - max_flow: 15.0
  - tidal_volume:
    - min: 0.0
    - max: 0.3
  - peep: 4
  - compliance_func: "constant_compliance"
  - starting_volume: 0.0
  - breath_variation: 50.0
  - max_breath_interval: 9000.0
  - measurement_error_flow: 1.0
  - measurement_error_pressure: 0.1
  - average_flow: 60.0
```
