## Supplementary Git Repository for "Inexpensive multi-patient respiratory monitoring system for helmet ventilation during COVID-19 pandemic": start_sims.py

```
#!/usr/bin/env python3
from __future__ import annotations

from .ventsim import VentSim
import random
import os
from typing import Any, Dict


def start_sims(nSim, start_time, sim_time):

    sims = []
    for i in range(nSim):

        params: Dict[str, Any] = {}

        simulator = VentSim(start_time, sim_time)
        simulator.load_configs(
            os.path.join(os.path.dirname(os.path.realpath(__file__)), "sim_configs.yml")
        )
        if i == 3:
            simulator.use_config("alarm_nonbreather")
        else:
            if random.random() < 0.5:
                simulator.use_config("nominal_breather", params)
            else:
                simulator.use_config("nominal_nonbreather", params)

        simulator.initialize_sim()

        # advance it somewhat
        # t = sim.get_batch(100 * random.random())
        sims.append(simulator)

    return sims
```
