## Supplementary Git Repository for "Inexpensive multi-patient respiratory monitoring system for helmet ventilation during COVID-19 pandemic": ventsim.py

```
#!/usr/bin/env python3
from __future__ import annotations

import numpy as np
import yaml
import logging
from typing import Tuple, Any, Dict, Optional


logger = logging.getLogger("povm")


def constant_compliance(**kwargs):
    return 0.1  # L / cm H2O


known_compliance_functions = {"constant_compliance": constant_compliance}


class VentSim:
    def __init__(
        self,
        curr_time: float,
        sim_time_max: float,
        params: Optional[Dict[str, Any]] = None,
    ):
        if params is None:
            params = {}

        self.current_bin = 0
        self.curr_time = curr_time
        self.sim_time = sim_time_max

        self.sample_length = params.get("sample_length", 20.0)
        self.breath_interval = params.get("breath_interval", 7000.0)
        self.max_flow = params.get("max_flow", 15.0)
        self.tidal_volume = params.get("tidal_volume", 0.6)
        self.peep = params.get("peep", 4)
        self.compliance_func = params.get("compliance_func", constant_compliance)
        self.starting_volume = params.get("starting_volume", 0.0)
        self.breath_variation = params.get("breath_variation", 300.0)
        self.max_breath_interval = params.get("max_breath_interval", 9000.0)
        self.measurement_error_pressure = params.get("measurement_error_pressure", 0.0)
        self.measurement_error_flow = params.get("measurement_error_flow", 0.0)
        self.average_flow = params.get("average_flow", 0.0)

    def initialize_sim(self) -> None:
        logger.info("Running sim with these parameters")
        self.print_config()
        self.precompute()

    def load_configs(self, yml_file: str) -> None:

        stream = open(yml_file, "r")
        params = yaml.safe_load(stream)
        self.configs = params

    def interpret_yaml_key(self, val):
        if type(val) == list:
            possible_keys = {"mean": -1, "sigma": -1, "min": -1, "max": -1}
            for t_dict in val:
                for key in t_dict:
                    if key in possible_keys:
                        possible_keys[key] = t_dict[key]
                    else:
                        raise RuntimeError(f"Unexpected value {key}")

            if possible_keys["mean"] > -1 and possible_keys["sigma"] > -1:
                return np.random.normal(possible_keys["mean"], possible_keys["sigma"])
            if possible_keys["min"] > -1 and possible_keys["max"] > -1:
                return np.random.uniform(possible_keys["min"], possible_keys["max"])
            raise RuntimeError("Missing mean/sigma or min/max values")
        else:
            return val  # its a value

    def use_config(self, config, params: Optional[Dict[str, float]] = None) -> None:
        if params is None:
            params = {}
        assert config in self.configs, "missing configuration " + config
        new_config = self.configs[config]
        for t_dict in new_config:
            for key in t_dict:
                if key not in params:
                    setattr(self, key, self.interpret_yaml_key(t_dict[key]))
                    logger.info(f"{key}, {getattr(self, key)}")
                else:
                    setattr(self, key, params[key])
        if isinstance(self.compliance_func, str):
            assert self.compliance_func in known_compliance_functions, (
                "missing compliance function " + self.compliance_func
            )
            self.compliance_func = known_compliance_functions[self.compliance_func]
        logger.info("Changed ventsim configuration to {config}")

    def print_config(self) -> None:
        logger.info(f"Sample length (ms) {self.sample_length}")
        logger.info(f"Breathing interval (ms) {self.breath_interval}")
        logger.info(f"Maximum flow (mL/m) {self.max_flow}")
        logger.info(f"Tidal volume (L) {self.tidal_volume}")
        logger.info(f"PEEP (cm H2O) {self.peep}")
        logger.info(f"Staring volume (L) {self.starting_volume}")
        logger.info(f"Breath variation (sigma in ms) {self.breath_variation}")
        logger.info(f"Maximum interval between breaths (ms) {self.max_breath_interval}")

    def precompute(self) -> None:
        self.breath_starts = self.get_breath_starts()
        self.flow = self.nominal_flow()
        self.times = np.arange(
            0, 1.2 * self.sim_time, self.sample_length, dtype=np.int64
        )[: len(self.flow)]
        self.volume = self.nominal_volume()
        self.pressure = self.nominal_pressure()
        self.flow = self.average_flow + self.flow

    def extend(self) -> None:
        self.curr_time += self.sim_time
        self.precompute()
        self.current_bin = 0

    def get_breath_starts(self) -> np.array:
        """
        returns:
        array of breath start times
        """

        max_breaths = (
            1.2 * self.sim_time / self.breath_interval
        )  # safety margin for fluctuating this later
        deltas = np.random.normal(
            self.breath_interval, self.breath_variation, int(max_breaths)
        )
        deltas = np.minimum(deltas, self.max_breath_interval)
        breath_starts = np.append(
            0, np.cumsum(deltas)
        )  # put a breath at the beginning..
        return breath_starts

    def nominal_flow(self) -> np.array:

        """
        expected parameters:
        inhale_exhale_ratio (optional, default is 0.5) (unused)
        """

        # bins = int(self.sim_time * self.sampling_rate)
        bins = int(
            self.breath_starts[np.where(self.breath_starts > self.sim_time)][0]
            / self.sample_length
        )
        flow = np.zeros(bins)

        flow_start_bins = (self.breath_starts) // self.sample_length

        # max_flow is in L/minute
        flow_end_bins = flow_start_bins + self.tidal_volume / self.sample_length / (
            self.max_flow / 60000.0
        )

        flow_start_bins = flow_start_bins.astype(int)
        flow_end_bins = flow_end_bins.astype(int)
        breath_integrals = np.zeros(len(flow_start_bins))

        for i in range(len(flow_start_bins)):
            if flow_start_bins[i] < bins:
                flow[flow_start_bins[i] : min(bins, flow_end_bins[i])] = self.max_flow
                breath_integrals[i] = np.sum(
                    flow[flow_start_bins[i] : min(bins, flow_end_bins[i])]
                )

        exp_zero_bins = flow_start_bins[1:]
        exp_min_bins = flow_end_bins[:-1]
        times = np.arange(0, 1.2 * self.sim_time, self.sample_length)
        # later times = times.astype(int)

        for i in range(len(exp_min_bins)):
            if exp_min_bins[i] < bins:
                b_min = exp_min_bins[i] + 2
                b_max = min(bins, exp_zero_bins[i]) - 1
                flow[b_min:b_max] = np.log(
                    (times[b_min:b_max] - times[flow_start_bins[i]])
                    / (times[b_max] - times[flow_start_bins[i]])
                )
                flow[b_min:b_max] *= (
                    -1.0 * breath_integrals[i] / np.sum(flow[b_min:b_max])
                )

        return flow

    def nominal_volume(self):
        # convert to L from L/m
        return (
            np.cumsum(self.flow) * (self.sample_length / (60.0 * 1000.0))
            + self.starting_volume
        )

    def nominal_pressure(self):
        # We can add **kwargs and join it with locals if we need to go further up the chain
        compliance_val = self.compliance_func
        pressure = np.zeros(len(self.volume))
        pressure[0] = self.peep
        delta_p = (
            self.volume[1:] - self.volume[:-1]
        ) / compliance_val()  # compliance is DeltaV/DeltaP
        pressure[1:] = pressure[0] + np.cumsum(delta_p)
        return pressure

    def get_next(self) -> Dict[str, Any]:
        if self.current_bin >= len(self.times):
            self.extend()
        d = {
            "v": 1,
            "t": int(self.curr_time + self.times[self.current_bin]),
            "F": float(
                self.flow[self.current_bin]
                + np.random.normal(0, self.measurement_error_flow, 1)
            ),
            "P": float(
                self.pressure[self.current_bin]
                + np.random.normal(0, self.measurement_error_pressure, 1)
            ),
            "temp": 23.3,
        }

        self.current_bin += 1
        return d

    def get_batch(self, nMilliSeconds: float):
        nbins = int(nMilliSeconds / self.sample_length)
        if self.current_bin + nbins > len(self.times):
            self.extend()
        fbin = self.current_bin
        lbin = self.current_bin + nbins

        d = {
            "version": 1,
            "source": "simulation",
            "parameters": {},
            "data": {
                "timestamps": (self.curr_time + self.times[fbin:lbin])
                .astype(int)
                .tolist(),
                "flows": (
                    self.flow[fbin:lbin]
                    + np.random.normal(0, self.measurement_error_flow, lbin - fbin)
                ).tolist(),
                "pressures": (
                    self.pressure[fbin:lbin]
                    + np.random.normal(0, self.measurement_error_pressure, lbin - fbin)
                ).tolist(),
            },
        }

        self.current_bin += nbins
        return d

    def get_all(self):
        return (
            self.times.astype(int),
            self.flow
            + np.random.normal(0, self.measurement_error_flow, len(self.times)),
            self.volume,
            self.pressure
            + np.random.normal(0, self.measurement_error_pressure, len(self.times)),
        )

    def get_from_timestamp(
        self, t: int, nMilliSeconds: int
    ) -> Tuple[np.ndarray, np.ndarray, np.ndarray]:
        lbin = np.searchsorted(self.times, t - self.curr_time, side="left")
        fbinT = 0
        nbins = int(nMilliSeconds / self.sample_length)
        if lbin == len(self.times):
            fbinT = np.searchsorted(
                self.times, t - nMilliSeconds - self.curr_time, side="left"
            )
            if fbinT != len(self.times):
                lbinT = np.searchsorted(self.times, self.sim_time, side="left")
                saveTimes = self.times[fbinT:lbinT]
                saveFlow = self.flow[fbinT:lbinT]
                savePressure = self.pressure[fbinT:lbinT]

            self.extend()
            lbin = np.searchsorted(self.times, t - self.curr_time, side="left")
            assert lbin != len(
                self.times
            ), "something wrong in timestamps - or use more simulation chunks"

            if fbinT == len(self.times):
                if lbin < nbins:
                    fbin = 0
                else:
                    fbin = lbin - nbins

                return (
                    (self.curr_time + self.times[fbin:lbin]).astype(int),
                    self.flow[fbin:lbin]
                    + np.random.normal(0, self.measurement_error_flow, lbin - fbin),
                    self.pressure[fbin:lbin]
                    + np.random.normal(0, self.measurement_error_pressure, lbin - fbin),
                )
            else:
                return (
                    (self.curr_time + np.append(saveTimes, self.times[:lbin])).astype(
                        int
                    ),
                    np.append(saveFlow, self.flow[:lbin])
                    + np.random.normal(
                        0, self.measurement_error_flow, lbin + lbinT - fbinT
                    ),
                    np.append(savePressure, self.pressure[:lbin])
                    + np.random.normal(
                        0, self.measurement_error_pressure, lbin + lbinT - fbinT
                    ),
                )

        else:
            if lbin < nbins:
                fbin = 0
            else:
                fbin = lbin - nbins

            return (
                (self.curr_time + self.times[fbin:lbin]).astype(int),
                self.flow[fbin:lbin]
                + np.random.normal(0, self.measurement_error_flow, lbin - fbin),
                self.pressure[fbin:lbin]
                + np.random.normal(0, self.measurement_error_pressure, lbin - fbin),
            )


if __name__ == "__main__":

    import matplotlib.pyplot as plt
    import time

    now_time = 1000 * time.monotonic()
    print(now_time)

    simulator = VentSim(now_time, 1200000)
    simulator.load_configs("sim_configs.yml")
    simulator.use_config("nominal_breather")
    simulator.initialize_sim()

    for _ in range(0, 10):
        print(simulator.get_next())

    print("testing get from timestamp features")

    time.sleep(5)
    d = simulator.get_from_timestamp(int(1000 * time.monotonic()), 10000)
    print(len(d[0]))
    time, flow, volume, pressure = simulator.get_all()

    fig = plt.figure()
    plt.subplot(3, 1, 1)
    (line_flow,) = plt.plot(time, flow)
    plt.xlabel("Time (seconds)")
    plt.ylabel("Flow (L/m)")

    plt.subplot(3, 1, 2)
    (line_volume,) = plt.plot(time, volume)
    plt.xlabel("Time (seconds)")
    plt.ylabel("Volume (L)")

    plt.subplot(3, 1, 3)
    (line_pressure,) = plt.plot(time, pressure)
    plt.xlabel("Time (seconds)")
    plt.ylabel("Pressure (cm H2O)")

    plt.show()
```
