## Supplementary Git Repository for "Inexpensive multi-patient respiratory monitoring system for helmet ventilation during COVID-19 pandemic": timeseries_to_breaths.py

```
#!/usr/bin/env python3

from processor.argparse import ArgumentParser

parser = ArgumentParser()
parser.add_argument("input", help="Calibrated timeseries CSV file")
parser.add_argument(
    "--drop-header",
    action="store_true",
    help="If enabled, the header line will be dropped",
)
args = parser.parse_args()

from processor.config import config

import csv

import numpy

from processor import analysis

# Documentation for all analysis products, including breath records
# https://github.com/Princeton-Penn-Vents/princeton-penn-flowmeter/blob/master/docs/analysis-products.md

order_and_mapping = [
    # header name (human readable with units), breath key name (can't change)
    ("inhale timestamp (sec)", "inhale timestamp"),
    ("inhale flow (L/min)", "inhale flow"),
    ("inhale dV/dt (mL/sec)", "inhale dV/dt"),
    ("inhale dP/dt (cm H2O/sec)", "inhale dP/dt"),
    ("inhale compliance (ml/cm H2O)", "inhale compliance"),
    ("min pressure (cm H2O)", "min pressure"),
    ("full timestamp (sec)", "full timestamp"),
    ("full pressure (cm H2O)", "full pressure"),
    ("full volume (mL)", "full volume"),
    ("expiratory tidal volume (mL)", "expiratory tidal volume"),
    ("inspiratory tidal volume (mL)", "inspiratory tidal volume"),
    ("inhale time (sec)", "inhale time"),
    ("exhale timestamp (sec)", "exhale timestamp"),
    ("exhale flow (L/min)", "exhale flow"),
    ("exhale dV/dt (mL/sec)", "exhale dV/dt"),
    ("exhale dP/dT (cm H2O/sec)", "exhale dP/dt"),
    ("exhale compliance (ml/cm H2O)", "exhale compliance"),
    ("max pressure (cm H2O)", "max pressure"),
    ("empty timestamp (sec)", "empty timestamp"),
    ("empty pressure (cm H2O)", "empty pressure"),
    ("empty volume (mL)", "empty volume"),
    ("exhale time (sec)", "exhale time"),
    ("average flow (L/min)", "average flow"),
    ("average pressure (cm H2O)", "average pressure"),
    ("time since last (sec)", "time since last"),
]

time = []
pressure = []
flow = []
volume = []
minbias_volume = []

with open(args.input) as fin:
    # skip the header
    next(fin)

    reader = csv.reader(fin)
    for t_raw, p_raw, f_raw, v_raw, mv_raw in reader:
        t, p, f, v, mv = (
            float(t_raw),
            float(p_raw),
            float(f_raw),
            float(v_raw),
            float(mv_raw),
        )
        time.append(t)
        pressure.append(p)
        flow.append(f)
        volume.append(v)
        minbias_volume.append(mv)

time = numpy.array(time)
pressure = numpy.array(pressure)
flow = numpy.array(flow)
volume = numpy.array(volume)
minbias_volume = numpy.array(minbias_volume)

breath_thresh = config["global"]["breath-thresh"].as_number()
breaths = analysis.measure_breaths(
    time, flow, minbias_volume, pressure, breath_thresh=breath_thresh
)

breaths, updated, new_breaths = analysis.combine_breaths([], breaths)

if not args.drop_header:
    print(", ".join(head for head, key in order_and_mapping))

for breath in breaths:
    line = []
    for _head, key in order_and_mapping:
        line.append(str(breath.get(key, "nan")))
    print(", ".join(line))
```
