## Supplementary Data Files 1 for "Inexpensive multi-patient respiratory monitoring system for helmet ventilation during COVID-19 pandemic": POVMRespiratoryAnalysisExample.ipynb

```
# %% [code]
# Princeton Open Ventillation Monitor Analysis - Breaths
import json
import numpy as np
import matplotlib.pyplot as plt
import scipy.integrate
import scipy.signal
import scipy.interpolate
import yaml

# %% [code]
# these lines can be commented out if data is available locally
!wget --no-check-certificate 'https://docs.google.com/uc?export=download&id=1UmOcL4tCoN8YbwTYiS4UIViAWlFx-O2I' -O device_raw_waveforms_expiratory.json
#!wget --no-check-certificate 'https://docs.google.com/uc?export=download&id=1g1rYMiZ1s7NKfBJjzH_Fwf3qxoWpqzRO' -O device_raw_waveforms_inspiratory.json
!wget --no-check-certificate 'https://docs.google.com/uc?export=download&id=11GFvDS-f3tY1G113ut-T64_dF-Lj4awN' -O device_raw_waveforms_auxport.json
!wget --no-check-certificate 'https://docs.google.com/uc?export=download&id=1HgL8yJN3Jp8iVTp2kvM9mcmroqdWj0th' -O flowcalib_ave200619.yml

# %% [code]
# yaml (yet-another-markup-language) file reading function
# function for reading flow calibration file
def get_yaml(yml_file):
    stream = open(yml_file, "r")
    params = yaml.safe_load(stream)
    qs = np.zeros(len(params))
    deltaPs = np.zeros(len(params))
    for i, d in enumerate(params):
        qs[i] = d["Q"]
        deltaPs[i] = d["deltaP"]

    return qs, deltaPs

# %% [code]
# this reads a flow calibration file and sets up a calibration function
qs, deltaPs = get_yaml("flowcalib_ave200619.yml")
interp = scipy.interpolate.interp1d(
                deltaPs, qs, kind="cubic", fill_value="extrapolate"
            )
extrap1d = lambda deltaP : interp(np.abs(deltaP)/60.0)

# %% [code]
# respiratory data reading function (pressure and flow calibrations applied)
def read_our(filename):
    with open(filename) as file:
        time, pressure, flow = [], [], []
        for line in file:
            try:
                row = json.loads(line)
            except:
                pass
            else:
                time.append(row["t"] / 1000.0)
                pressure.append(0.0126 * row["P"] - 12.73)
#                flow.append(0.7198 * np.sign(row["F"])*abs(row["F"])**(4/7))
                flow.append(np.sign(row["F"])*extrap1d(row["F"]))
        return np.array(time), np.array(pressure), np.array(flow)

# %% [code]
# reading of auxiliary and expiratory port data
time, pressure, flow = read_our("device_raw_waveforms_auxport.json")
time2, pressure2, flow2 = read_our("device_raw_waveforms_expiratory.json")

# %% [code]
# timestamp sychronization for auxiliary and expriatory ports 
timesync = 448.587
timesync2 = 344.856
sync = np.where(time == timesync)[0][0]
sync2 = np.where(time2 == timesync2)[0][0]
print(sync,time[sync])
print(sync2,time2[sync2])

# %% [code]
# data periods
from datetime import datetime
starttime = datetime(2020, 9, 14, 10, 52, 10)

# Uncomment the successive datastable1 and datastable1end lines to look at each separate period of the data
# ----------------------------------
# 83 L/min helmet breathing stable period 1
#datastable1 = datetime(2020, 9, 14, 11, 12, 21)
#datastable1end = datetime(2020, 9, 14, 11, 14, 58)
# 83 L/min helmet breathing stable period 2
datastable1 = datetime(2020, 9, 14, 11, 15, 20)
datastable1end = datetime(2020, 9, 14, 11, 19, 29)
# 83 L/min external breathing
#datastable1 = datetime(2020, 9, 14, 10, 58, 14)
#datastable1end = datetime(2020, 9, 14, 10, 58, 54)
# 105 L/min external breathing
#datastable1 = datetime(2020, 9, 14, 11, 4, 15)
#datastable1end = datetime(2020, 9, 14, 11, 4, 59)
# 52 L/min external breathing
#datastable1 = datetime(2020, 9, 14, 11, 1, 18)
#datastable1end = datetime(2020, 9, 14, 11, 2, 28)
#null data 83 L/min
#datastable1 = datetime(2020, 9, 14, 10, 54, 5)
#datastable1end = datetime(2020, 9, 14, 10, 55, 50)
#all data
#datastable1 = datetime(2020, 9, 14, 10, 52, 10)
#datastable1end = datetime(2020, 9, 14, 11, 22, 34)
duration = datastable1 - starttime
duration_in_sec = duration.total_seconds()
duration1 = datastable1end - datastable1
duration1_in_sec = duration1.total_seconds()
print(duration_in_sec,duration1_in_sec)

# %% [code]
# translation of times to indicies
timestart = timesync+duration_in_sec
sync = np.where(time >= timestart)[0][0]
timesyncend = timestart+duration1_in_sec
window = np.where(time >= timesyncend)[0][0] - sync
off = 6
inx0 = sync + off
inx1 = sync + off + window
print(inx0,inx1)
timestart2 = timesync2+duration_in_sec
sync2 = np.where(time2 >= timestart2)[0][0]
off2 = 0
inx2 = sync2 + off2
inx3 = sync2 + off2 + window
print(inx2,inx3)

# %% [code]
# high-pass filter parameter to maintain an average 
critical_freq = 0.004 # monitoring
#critical_freq = 0.0004 # minbias

# %% [code]
# filtering of volume baseline
rawvolume = scipy.integrate.cumtrapz(flow, time / 60.0, initial=0)
ourvolume = scipy.signal.sosfilt(scipy.signal.butter(1, critical_freq, "highpass", output="sos"), rawvolume)
ourvolume -= ourvolume[inx0:inx1].min()
rawvolume2 = scipy.integrate.cumtrapz(flow2, time2 / 60.0, initial=0)
ourvolume2 = scipy.signal.sosfilt(scipy.signal.butter(1, critical_freq, "highpass", output="sos"), rawvolume2)
ourvolume2 -= ourvolume2[inx2:inx3].min()
volume_by_average = scipy.integrate.cumtrapz(flow - flow[inx0:inx1].mean(), time / 60.0, initial=0)
volume_by_average -= volume_by_average[inx0:inx1].min()
volume_by_average2 = scipy.integrate.cumtrapz(flow2 - flow2[inx2:inx3].mean(), time2 / 60.0, initial=0)
volume_by_average2 -= volume_by_average2[inx2:inx3].min()

# %% [code]
# processing of pressure data to remove spurious spikes
pressure_average = 0.25*(
    pressure[4:] +
    pressure[3:-1] +
    pressure[1:-3] +
    pressure[:-4])

toreplace22 = abs(
    pressure_average -
    pressure[2:-2]) > 0.1
toreplace = np.zeros(
    len(pressure), np.bool_)
toreplace[2:-2] = toreplace22

pressure_deglitched = pressure.copy()
pressure_deglitched[toreplace] = \
    pressure_average[toreplace22]

pressure_average2 = 0.25*(
    pressure2[4:] +
    pressure2[3:-1] +
    pressure2[1:-3] +
    pressure2[:-4])

toreplace222 = abs(
    pressure_average2 -
    pressure2[2:-2]) > 0.1
toreplace2 = np.zeros(
    len(pressure2), np.bool_)
toreplace2[2:-2] = toreplace222

pressure_deglitched2 = pressure2.copy()
pressure_deglitched2[toreplace2] = \
    pressure_average2[toreplace222]

# %% [code]
# plot formatting commands
plt.rcParams.update({'xtick.labelsize': 20})
plt.rcParams.update({'ytick.labelsize': 20})
plt.rcParams.update({'lines.linewidth': 2.0})
plt.rcParams.update({'axes.linewidth': 2.0})
plt.rcParams.update({'axes.labelpad': 20.0})
plt.rcParams.update({'axes.xmargin': 0.0})
#plt.rcParams.update({'axes.ymargin': 0.0})
plt.rcParams.update({'xtick.major.size': 10.0})
plt.rcParams.update({'ytick.major.size': 10.0})
plt.rcParams.update({'xtick.major.width': 2.0})
plt.rcParams.update({'ytick.major.width': 2.0})
plt.rcParams.update({'xtick.major.pad': 10.0})
plt.rcParams.update({'ytick.major.pad': 10.0})
plt.rcParams.update({'xtick.direction': 'in'})
plt.rcParams.update({'ytick.direction': 'in'})
plt.rcParams.update({'font.family': 'serif'})
plt.rcParams.update({'font.size': 20})
plt.rcParams.update({'figure.dpi': 300})
plt.rcParams.update({'savefig.dpi': 300})
plt.rcParams.update({'figure.facecolor': 'white'})
plt.rcParams.update({'figure.figsize': (25,15)})

# %% [code]
# plot of auxiliary port and expiratory path monitoring readings
fig, (ax1, ax2, ax3) = plt.subplots(3, 1, figsize=(25, 15))
deglitched, = ax1.plot(time[inx0:inx1] - time[inx0], pressure_deglitched[inx0:inx1], color='orange')
deglitched2, = ax1.plot(time2[inx2:inx3] - time2[inx2], pressure_deglitched2[inx2:inx3], color='blue')
ax1.legend([deglitched,deglitched2], ["auxport", "expiratory"], loc="upper right")
ax1.set_ylabel("pressure (cm H₂O)")
ax1.set_xlim(time2[inx2] - time2[inx2], time2[inx3] - time2[inx2])
qlflow, = ax2.plot(time[inx0:inx1] - time[inx0], flow[inx0:inx1], color='orange')
expflow, = ax2.plot(time2[inx2:inx3] - time2[inx2], flow2[inx2:inx3], color='blue')
ax2.legend([qlflow,expflow], ["auxport", "expiratory"], loc="upper right")
ax2.set_ylabel("flow (L/min)")
ax2.set_xlim(time2[inx2] - time2[inx2], time2[inx3] - time2[inx2])
#byaverage, = ax3.plot(time[inx0:inx1] - time[inx0], volume_by_average[inx0:inx1], color='orange')
byhighpass, = ax3.plot(time[inx0:inx1] - time[inx0], ourvolume[inx0:inx1], color='orange')
#byaverage2, = ax3.plot(time2[inx2:inx3] - time2[inx2], volume_by_average2[inx2:inx3], color='blue')
byhighpass2, = ax3.plot(time2[inx2:inx3] - time2[inx2], ourvolume2[inx2:inx3], color='blue')
#ax3.legend([byaverage, byhighpass], ["corrected by flow average", "corrected by high-pass filter"])
ax3.legend([byhighpass,byhighpass2], ["auxport", "expiratory"], loc="upper right")
ax3.set_ylabel("volume (L)")
ax3.set_xlim(time2[inx2] - time2[inx2], time2[inx3] - time2[inx2])
ax3.set_xlabel("time since start (sec)")

# %% [code]
# alternative method for analyzing respirator rate
from scipy.signal import find_peaks

# parameters that identify patient's breaths
RRmin = 3.  # 3 Hz min respiratory rate (bpm)
RRmax = 60. # 60 Hz max respiratory rate (bpm)
peak_dist_min = int(60.*50/RRmax) # index distance computed using 50 Hz (=30*50/min) of flow readings
#Vmin = 0.2 # high side 50mL min volume (L)
#Vmin = 0.03 # low side 50mL min volume (L)
Vmin = 0.05 # 50mL min volume (L)
Vwidth_min = int(60.*50/RRmax/2.) # 25*0.02s = 0.5s
Vwidth_max = int(60.*50/RRmin) # 1000*0.02s = 20.0s equivalent to 3 Hz

fig, ax1 = plt.subplots(1, 1, figsize=(25, 15))
#fig, (ax1, ax2) = plt.subplots(2, 1, figsize=(25, 15))
#byaverage, = ax3.plot(time[inx0:inx1] - time[inx0], volume_by_average[inx0:inx1], color='orange')
byhighpass, = ax1.plot(time[inx0:inx1] - time[inx0], ourvolume[inx0:inx1], color='orange')
#byaverage2, = ax3.plot(time2[inx2:inx3] - time2[inx2], volume_by_average2[inx2:inx3], color='blue')
byhighpass2, = ax1.plot(time2[inx2:inx3] - time2[inx2], ourvolume2[inx2:inx3], color='blue')
#ax3.legend([byaverage, byhighpass], ["corrected by flow average", "corrected by high-pass filter"])
ax1.legend([byhighpass,byhighpass2], ["auxport", "expiratory"], loc="upper right")
ax1.set_ylabel("volume (L)")
ax1.set_xlim(time2[inx2] - time2[inx2], time2[inx3] - time2[inx2])
ax1.set_xlabel("time since start (sec)")

p = pressure_deglitched[inx0:inx1]
p2 = pressure_deglitched2[inx2:inx3]
ppeaks, pprops = find_peaks(p, distance=peak_dist_min)
ppeaks2, pprops2 = find_peaks(p2, distance=peak_dist_min)
pp = p[ppeaks]
pp2 = p2[ppeaks2]
print("pressure aux",pp.mean(), pp.std(), "pressure exp", pp2.mean(), pp2.std())
f = flow[inx0:inx1]
f2 = flow2[inx2:inx3]
fpeaks, fprops = find_peaks(f, distance=peak_dist_min)
fpeaks2, fprops2 = find_peaks(f2, distance=peak_dist_min)
fp = f[fpeaks]
fp2 = f2[fpeaks2]
print("flow aux",fp.mean(), fp.std(), "flow exp", fp2.mean(), fp2.std())
v = ourvolume[inx0:inx1]
v2 = ourvolume2[inx2:inx3]
vpeaks, vprops = find_peaks(v, prominence=Vmin, distance=peak_dist_min, width=(Vwidth_min,Vwidth_max))
vpeaks2, vprops2 = find_peaks(v2, prominence=Vmin, distance=peak_dist_min, width=(Vwidth_min,Vwidth_max))
vp = v[vpeaks]
vp2 = v2[vpeaks2]
t = time[inx0:inx1] - time[inx0]
tf = t[vpeaks]
ax1.plot(tf,vp, 'x', color='orange')
ax1.vlines(x=tf,ymin=vp-vprops["prominences"],ymax=vp, color='orange')
ax1.hlines(y=vprops["width_heights"], xmin=t[vprops["left_ips"].astype(int)], xmax=t[vprops["right_ips"].astype(int)], color='#ff7f0e')
t2 = time2[inx2:inx3] - time2[inx2]
tf2 = t2[vpeaks2]
ax1.plot(tf2,vp2, 'x', color='blue')
ax1.vlines(x=tf2,ymin=vp2-vprops2["prominences"],ymax=vp2, color='blue')
ax1.hlines(y=vprops2["width_heights"], xmin=t2[vprops2["left_ips"].astype(int)], xmax=t2[vprops2["right_ips"].astype(int)], color='#1f77b4')
if len(vpeaks2)>0:
    print("volume exp", vprops2["prominences"].mean(), vprops2["prominences"].std())
if len(vpeaks)>0:
    print("volume aux", vprops["prominences"].mean(), vprops["prominences"].std())
# comparing volumes requires that the same breaths are identified in the auxiliary port and expiratory paths
if len(vpeaks2) == len(vpeaks) and len(vpeaks)>0:
    vf = vprops2["prominences"]/vprops["prominences"]
    print("aux/exp frac", vf[1:].mean()-1.0,vf[1:].std()) # drop first breath
    volfrac, = ax2.plot(tf, vf, label="auxport/expiratory", color='black')
    ax2.legend(loc="upper right")
    ax2.set_ylabel("volume ratio (L)")
    ax2.set_xlim(time2[inx2] - time2[inx2], time2[inx3] - time2[inx2])
    ax2.set_ylim(0.7,1.25)
    ax2.set_xlabel("time since start (sec)")

# %% [code]
# Respiratory rate from this alternative calculation
RRbpm = 0.
if len(vpeaks2)>0:
    RRbpm = 60.*(len(vpeaks2)-1)/(tf2[-1]-tf2[0])
print("number of breaths = {:6d}".format(len(vpeaks2)))
print("Respiratory Rate = {:4.1f} bpm".format(RRbpm))

# %% [code]
# processing of flow data
def smooth_derivative(times, values, sig=0.2):
    values = values - values.mean()

    if len(times) < 1:
        raise CantComputeDerivative

    themin = np.min(times[1:] - times[:-1])
    if themin <= 0:
        raise CantComputeDerivative

    window_width = int(np.ceil(4 * sig / themin))
    if len(times) - window_width + 1 < 10 or window_width < 10:
        raise CantComputeDerivative

    windowed_times = np.lib.stride_tricks.as_strided(
        times,
        (len(times) - window_width + 1, window_width),
        (times.itemsize, times.itemsize),
    )
    windowed_values = np.lib.stride_tricks.as_strided(
        values,
        (len(values) - window_width + 1, window_width),
        (values.itemsize, values.itemsize),
    )

    centers = np.mean(windowed_times, axis=1)
    windowed_times_centered = windowed_times - centers[:, np.newaxis]
    windowed_weights = np.exp(-0.5 * windowed_times_centered ** 2 / sig ** 2)
    sumw = np.sum(windowed_weights, axis=1)
    sumwx = np.sum(windowed_weights * windowed_times_centered, axis=1)
    sumwy = np.sum(windowed_weights * windowed_values, axis=1)
    sumwxx = np.sum(
        windowed_weights * windowed_times_centered * windowed_times_centered, axis=1
    )
    sumwxy = np.sum(
        windowed_weights * windowed_times_centered * windowed_values, axis=1
    )
    delta = (sumw * sumwxx) - (sumwx * sumwx)

    with np.errstate(all="ignore"):
        intercept = ((sumwxx * sumwy) - (sumwx * sumwxy)) / delta
        slope = ((sumw * sumwxy) - (sumwx * sumwy)) / delta

    good = (~np.isnan(intercept)) & (~np.isnan(slope))

    return centers[good], intercept[good], slope[good]

# %% [code]
# identifying features of the breath
def find_roots(
    times, values, derivative, threshold: float = 0.02, *, breath_thresh: float
):
    values_threshold = threshold * max(np.max(abs(values)), breath_thresh)
    derivative_threshold = threshold * np.max(abs(derivative))

    (A,) = np.nonzero(
        (values[:-1] < 0)
        & (values[1:] >= 0)
        & (0.5 * (derivative[:-1] + derivative[1:]) >= derivative_threshold)
    )
    (B,) = np.nonzero(
        (derivative[:-1] >= 0)
        & (derivative[1:] < 0)
        & (0.5 * (values[:-1] + values[1:]) >= values_threshold)
    )
    (C,) = np.nonzero(
        (values[:-1] >= -0.5 * values_threshold)
        & (values[1:] < -0.5 * values_threshold)
        & (0.5 * (derivative[:-1] + derivative[1:]) < -derivative_threshold)
    )
    (D,) = np.nonzero(
        (derivative[:-1] < 0)
        & (derivative[1:] >= 0)
        & (0.5 * (values[:-1] + values[1:]) < -values_threshold)
    )

    #print(times[A],times[B],times[C],times[D])
    plt.plot(times, values)
    plt.plot(times[A],values[A],'x',color='red')
    plt.plot(times[B],values[B],'x',color='green')
    plt.plot(times[C],values[C],'x',color='blue')
    plt.plot(times[D],values[D],'x',color='magenta')
    #plt.plot(times[B],values[B]-values[D])
    return (
        0.5 * (times[A] + times[A + 1]),
        0.5 * (times[B] + times[B + 1]),
        0.5 * (times[C] + times[C + 1]),
        0.5 * (times[D] + times[D + 1]),
    )

# %% [code]
# finding breaths
def find_breaths(A, B, C, D):
    # ensure that each type is sorted (though it probably already is)
    A.sort()
    B.sort()
    C.sort()
    D.sort()

    if len(A) == 0 or len(B) == 0 or len(C) == 0 or len(D) == 0:
        return []

    print("identifying breaths",len(A),len(B),len(C),len(D))
    # where does the cycle start?
    which = np.argmin([A[0], B[0], C[0], D[0]])

    ins = [A, B, C, D]
    outs = []
    while len(A) > 0 or len(B) > 0 or len(C) > 0 or len(D) > 0:
        if len(ins[which]) > 1 and all(len(ins[i]) > 0 for i in range(4)):
            nextwhich = np.argmin([ins[i][1 if i == which else 0] for i in range(4)])

            # normal case: A -> B -> C -> D -> ... cycle
            if nextwhich == (which + 1) % 4:
                outs.append((which, ins[which][0]))
                ins[which] = ins[which][1:]

            # too many of one type: A -> B -> B -> B -> C -> D -> ...
            elif nextwhich == which:
                combine = [ins[which][0]]
                ins[which] = ins[which][1:]
                while True:
                    if (
                        any(len(x) == 0 for x in ins)
                        or np.argmin([ins[i][0] for i in range(4)]) != which
                    ):
                        break
                    combine.append(ins[which][0])
                    ins[which] = ins[which][1:]

                outs.append((which, np.mean(combine)))

            # missing one type: A -> C -> D -> ...
            else:
                fill_value = 0.5 * (ins[which][0] + ins[nextwhich][0])
                ins[(which + 1) % 4] = np.concatenate(
                    (np.array([fill_value]), ins[(which + 1) % 4])
                )

                outs.append((which, ins[which][0]))
                ins[which] = ins[which][1:]

        # edge condition: less than one cycle left
        elif len(ins[which]) > 0:
            popped = ins[which][0]
            ins[which] = ins[which][1:]

            if len(outs) > 0 and (outs[-1][0] + 1) % 4 == which:
                outs.append((which, popped))

        else:
            break

        which = (which + 1) % 4
        A, B, C, D = ins

    return outs

# %% [code]
# measuring breaths
def measure_breaths(time, flow, volume, pressure, *, breath_thresh: float):
    try:
        smooth_time_f, smooth_flow, smooth_dflow = smooth_derivative(time, flow)
        smooth_time_p, smooth_pressure, smooth_dpressure = smooth_derivative(
            time, pressure
        )
        if len(smooth_time_f) < 4:
            return []

        #plt.plot(smooth_time_f, smooth_flow)
        turning_points = find_roots(
            smooth_time_f, smooth_flow, smooth_dflow, breath_thresh=breath_thresh
        )

        #print(turning_points)
        breath_times = find_breaths(*turning_points)

    except Exception as err:
        if not isinstance(err, CantComputeDerivative):
            raise
        return []
    
    if len(time) == 0 or len(breath_times) == 0:
        return []

    breaths = []
    breath = {}
    for i, (which, t) in enumerate(breath_times):
        index = np.argmin(abs(time - t))

        if which == 0:
            breath["empty timestamp"] = t
            if 0 <= index < len(pressure):
                breath["empty pressure"] = pressure[index]
            if 0 <= index < len(volume):
                breath["empty volume"] = volume[index]
            if i >= 2:
                full_index = np.argmin(abs(time - breath_times[i - 2][1]))
                if 0 <= full_index < len(volume):
                    diff = volume[full_index] - breath["empty volume"]
                    if diff > 0:
                        breath["inspiratory tidal volume"] = diff
            if i >= 4:
                last_index = np.argmin(abs(time - breath_times[i - 4][1]))
                if 0 <= last_index < len(flow) and last_index < index:
                    breath["average flow"] = flow[last_index:index].sum() / (
                        index - last_index
                    )
                if 0 <= last_index < len(pressure) and last_index < index:
                    breath["average pressure"] = pressure[last_index:index].sum() / (
                        index - last_index
                    )

            breaths.append(breath)
            breath = {}

        elif which == 1:
            breath["inhale timestamp"] = t
            if 0 <= index < len(flow):
                breath["inhale flow"] = flow[index]
            if len(smooth_time_f) > 0:
                idx = np.argmin(abs(smooth_time_f - t))
                if 0 <= idx < len(smooth_flow):
                    breath["inhale dV/dt"] = smooth_flow[idx] * 1000 / 60.0
            if len(smooth_time_p) > 0:
                idx = np.argmin(abs(smooth_time_p - t))
                if 0 <= idx < len(smooth_dpressure):
                    breath["inhale dP/dt"] = smooth_dpressure[idx]
            if "inhale dV/dt" in breath and "inhale dP/dt" in breath:
                breath["inhale compliance"] = (
                    breath["inhale dV/dt"] / breath["inhale dP/dt"]
                )
            if i >= 2:
                start_index = np.argmin(abs(time - breath_times[i - 2][1]))
                if start_index < index:
                    breath["min pressure"] = np.min(pressure[start_index:index])

        elif which == 2:
            breath["full timestamp"] = t
            if 0 <= index < len(pressure):
                breath["full pressure"] = pressure[index]
            if 0 <= index < len(volume):
                breath["full volume"] = volume[index]
            if i >= 2:
                empty_index = np.argmin(abs(time - breath_times[i - 2][1]))
                if 0 <= empty_index < len(volume):
                    diff = breath["full volume"] - volume[empty_index]
                    if diff > 0:
                        breath["expiratory tidal volume"] = diff

        elif which == 3:
            breath["exhale timestamp"] = t
            if 0 <= index < len(flow):
                breath["exhale flow"] = flow[index]
            if len(smooth_time_f) > 0:
                idx = np.argmin(abs(smooth_time_f - t))
                if 0 <= idx < len(smooth_flow):
                    breath["exhale dV/dt"] = smooth_flow[idx] * 1000 / 60.0
            if len(smooth_time_p) > 0:
                idx = np.argmin(abs(smooth_time_p - t))
                if 0 <= idx < len(smooth_dpressure):
                    breath["exhale dP/dt"] = smooth_dpressure[idx]
            if "exhale dV/dt" in breath and "exhale dP/dt" in breath:
                breath["exhale compliance"] = (
                    breath["exhale dV/dt"] / breath["exhale dP/dt"]
                )
            if i >= 2:
                start_index = np.argmin(abs(time - breath_times[i - 2][1]))
                if start_index < index:
                    breath["max pressure"] = np.max(pressure[start_index:index])

    if (
        "empty timestamp" in breath
        or "inhale timestamp" in breath
        or "full timestamp" in breath
        or "exhale timestamp" in breath
    ):
        breaths.append(breath)

    return breaths


# %% [code]
# analysis of threshold for identifying breaths
#bThresh = 450.0 # max value 450.0 for stable breathing and 200.0 for A/C points, works down to 0 threshold
bThresh = 50.0 # min value 35.0 for null data
breaths = measure_breaths(t2, f2, v2, p2, breath_thresh=bThresh)
print(len(breaths),breaths)
```
