## Supplementary Data Files 1 for "Inexpensive multi-patient respiratory monitoring system for helmet ventilation during COVID-19 pandemic": POVMWaveformAnalysisExample.ipynb

```
# %% [code]
# Princeton Open Ventillation Monitor Analysis
import json
import numpy as np
import matplotlib.pyplot as plt
import scipy.integrate
import scipy.signal
import scipy.interpolate
import yaml

# %% [code]
# these lines can be commented out if data is available locally
!wget --no-check-certificate 'https://docs.google.com/uc?export=download&id=1UmOcL4tCoN8YbwTYiS4UIViAWlFx-O2I' -O device_raw_waveforms_expiratory.json
#!wget --no-check-certificate 'https://docs.google.com/uc?export=download&id=1g1rYMiZ1s7NKfBJjzH_Fwf3qxoWpqzRO' -O device_raw_waveforms_inspiratory.json
!wget --no-check-certificate 'https://docs.google.com/uc?export=download&id=11GFvDS-f3tY1G113ut-T64_dF-Lj4awN' -O device_raw_waveforms_auxport.json
!wget --no-check-certificate 'https://docs.google.com/uc?export=download&id=1HgL8yJN3Jp8iVTp2kvM9mcmroqdWj0th' -O flowcalib_ave200619.yml

# %% [code]
# function for reading flow calibration file
def get_yaml(yml_file):
    stream = open(yml_file, "r")
    params = yaml.safe_load(stream)
    qs = np.zeros(len(params))
    deltaPs = np.zeros(len(params))
    for i, d in enumerate(params):
        qs[i] = d["Q"]
        deltaPs[i] = d["deltaP"]

    return qs, deltaPs

# %% [code]
# read flow calibration and setup correction function
qs, deltaPs = get_yaml("flowcalib_ave200619.yml")
interp = scipy.interpolate.interp1d(
                deltaPs, qs, kind="cubic", fill_value="extrapolate"
            )
extrap1d = lambda deltaP : interp(np.abs(deltaP)/60.0)

# %% [code]
# function to read datafiles
def read_our(filename):
    with open(filename) as file:
        time, pressure, flow = [], [], []
        for line in file:
            try:
                row = json.loads(line)
            except:
                pass
            else:
                time.append(row["t"] / 1000.0)
                pressure.append(0.0126 * row["P"] - 12.73)
#                flow.append(0.7198 * np.sign(row["F"])*abs(row["F"])**(4/7))
                flow.append(np.sign(row["F"])*extrap1d(row["F"]))
        return np.array(time), np.array(pressure), np.array(flow)

# %% [code]
# read datafiles
time, pressure, flow = read_our("device_raw_waveforms_auxport.json")
time2, pressure2, flow2 = read_our("device_raw_waveforms_expiratory.json")

# %% [code]
# timestamp synchronization of data array indices
timesync = 448.587
timesync2 = 344.856
sync = np.where(time == timesync)[0][0]
sync2 = np.where(time2 == timesync2)[0][0]
print(sync,time[sync])
print(sync2,time2[sync2])

# %% [code]
# data periods
from datetime import datetime
starttime = datetime(2020, 9, 14, 10, 52, 10)
# 83 L/min helmet breathing stable period 1
#datastable1 = datetime(2020, 9, 14, 11, 12, 21)
#datastable1end = datetime(2020, 9, 14, 11, 14, 58)
# 83 L/min helmet breathing stable period 2
#datastable1 = datetime(2020, 9, 14, 11, 15, 20)
#datastable1end = datetime(2020, 9, 14, 11, 19, 29)
# 83 L/min external breathing
datastable1 = datetime(2020, 9, 14, 10, 58, 14)
datastable1end = datetime(2020, 9, 14, 10, 58, 54)
##datastable1 = datetime(2020, 9, 14, 10, 57, 44)
##datastable1end = datetime(2020, 9, 14, 10, 59, 17)
# 105 L/min external breathing
#datastable1 = datetime(2020, 9, 14, 11, 4, 15)
#datastable1end = datetime(2020, 9, 14, 11, 4, 59)
##datastable1 = datetime(2020, 9, 14, 11, 4, 5)
##datastable1end = datetime(2020, 9, 14, 11, 5, 4)
# 52 L/min external breathing
#datastable1 = datetime(2020, 9, 14, 11, 1, 18)
#datastable1end = datetime(2020, 9, 14, 11, 2, 28)
##datastable1 = datetime(2020, 9, 14, 11, 0, 58)
##datastable1end = datetime(2020, 9, 14, 11, 2, 35)
#null data 83 L/min
#datastable1 = datetime(2020, 9, 14, 10, 53, 55)
#datastable1end = datetime(2020, 9, 14, 10, 55, 57)
#all data
#datastable1 = datetime(2020, 9, 14, 10, 52, 10)
#datastable1end = datetime(2020, 9, 14, 11, 22, 34)
duration = datastable1 - starttime
duration_in_sec = duration.total_seconds()
duration1 = datastable1end - datastable1
duration1_in_sec = duration1.total_seconds()
print(duration_in_sec,duration1_in_sec)

# %% [code]
# further time sycnhronization
timestart = timesync+duration_in_sec
sync = np.where(time >= timestart)[0][0]
timesyncend = timestart+duration1_in_sec
window = np.where(time >= timesyncend)[0][0] - sync
off = 6
inx0 = sync + off
inx1 = sync + off + window
print(inx0,inx1)
timestart2 = timesync2+duration_in_sec
sync2 = np.where(time2 >= timestart2)[0][0]
off2 = 0
inx2 = sync2 + off2
inx3 = sync2 + off2 + window
print(inx2,inx3)

# %% [code]
# baseline filtering for volume calcultions
critical_freq = 0.004 # monitoring
#critical_freq = 0.0004 # minbias

# %% [code]
# volume data processing
rawvolume = scipy.integrate.cumtrapz(flow, time / 60.0, initial=0)
ourvolume = scipy.signal.sosfilt(scipy.signal.butter(1, critical_freq, "highpass", output="sos"), rawvolume)
ourvolume -= ourvolume[inx0:inx1].min()
rawvolume2 = scipy.integrate.cumtrapz(flow2, time2 / 60.0, initial=0)
ourvolume2 = scipy.signal.sosfilt(scipy.signal.butter(1, critical_freq, "highpass", output="sos"), rawvolume2)
ourvolume2 -= ourvolume2[inx2:inx3].min()

# %% [code]
# alternative volume data processing
volume_by_average = scipy.integrate.cumtrapz(flow - flow[inx0:inx1].mean(), time / 60.0, initial=0)
volume_by_average -= volume_by_average[inx0:inx1].min()
volume_by_average2 = scipy.integrate.cumtrapz(flow2 - flow2[inx2:inx3].mean(), time2 / 60.0, initial=0)
volume_by_average2 -= volume_by_average2[inx2:inx3].min()

# %% [code]
# pressure data processing (deglitching)
pressure_average = 0.25*(
    pressure[4:] +
    pressure[3:-1] +
    pressure[1:-3] +
    pressure[:-4])

toreplace22 = abs(
    pressure_average -
    pressure[2:-2]) > 0.1
toreplace = np.zeros(
    len(pressure), np.bool_)
toreplace[2:-2] = toreplace22

pressure_deglitched = pressure.copy()
pressure_deglitched[toreplace] = \
    pressure_average[toreplace22]

pressure_average2 = 0.25*(
    pressure2[4:] +
    pressure2[3:-1] +
    pressure2[1:-3] +
    pressure2[:-4])

toreplace222 = abs(
    pressure_average2 -
    pressure2[2:-2]) > 0.1
toreplace2 = np.zeros(
    len(pressure2), np.bool_)
toreplace2[2:-2] = toreplace222

pressure_deglitched2 = pressure2.copy()
pressure_deglitched2[toreplace2] = \
    pressure_average2[toreplace222]

# %% [code]
# plot style
plt.rcParams.update({'xtick.labelsize': 20})
plt.rcParams.update({'ytick.labelsize': 20})
plt.rcParams.update({'lines.linewidth': 2.0})
plt.rcParams.update({'axes.linewidth': 2.0})
plt.rcParams.update({'axes.labelpad': 20.0})
plt.rcParams.update({'axes.xmargin': 0.0})
#plt.rcParams.update({'axes.ymargin': 0.0})
plt.rcParams.update({'xtick.major.size': 10.0})
plt.rcParams.update({'ytick.major.size': 10.0})
plt.rcParams.update({'xtick.major.width': 2.0})
plt.rcParams.update({'ytick.major.width': 2.0})
plt.rcParams.update({'xtick.major.pad': 10.0})
plt.rcParams.update({'ytick.major.pad': 10.0})
plt.rcParams.update({'xtick.direction': 'in'})
plt.rcParams.update({'ytick.direction': 'in'})
plt.rcParams.update({'font.family': 'serif'})
plt.rcParams.update({'font.size': 20})
plt.rcParams.update({'figure.dpi': 300})
plt.rcParams.update({'savefig.dpi': 300})
plt.rcParams.update({'figure.facecolor': 'white'})

# %% [code]
# plots of pressure, flow and volume for auxport and expiratory paths
#xlim0 = 0.
#xlim1 = 40.
xlim0 = time2[inx2] - time2[inx2]
xlim1 = time2[inx3] - time2[inx2]
fig, (ax1, ax2, ax3) = plt.subplots(3, 1, figsize=(25, 15))
#rawpressure, = ax1.plot(time[inx0:inx1] - time[0], pressure[inx0:inx1], color='#ff7f0e')
deglitched, = ax1.plot(time[inx0:inx1] - time[inx0], pressure_deglitched[inx0:inx1], color='#ff7f0e')
#rawpressure2, = ax1.plot(time2[inx0:inx1] - time2[0], pressure2[inx0:inx1], color='#ff7f0e')
deglitched2, = ax1.plot(time2[inx2:inx3] - time2[inx2], pressure_deglitched2[inx2:inx3], color='#1f77b4')
#ax1.legend([rawpressure, deglitched], ["raw pressure", "deglitched"], loc="upper right")
ax1.legend([deglitched,deglitched2], ["auxport", "expiratory"], loc="upper right")
ax1.set_ylabel("pressure (cm H₂O)")
ax1.set_xlim(xlim0, xlim1)
qlflow, = ax2.plot(time[inx0:inx1] - time[inx0], flow[inx0:inx1], color='#ff7f0e')
expflow, = ax2.plot(time2[inx2:inx3] - time2[inx2], flow2[inx2:inx3], color='#1f77b4')
ax2.legend([qlflow,expflow], ["auxport", "expiratory"], loc="upper right")
ax2.set_ylabel("flow (L/min)")
ax2.set_xlim(xlim0, xlim1)
#byaverage, = ax3.plot(time[inx0:inx1] - time[0], volume_by_average[inx0:inx1], color='#ff7f0e')
byhighpass, = ax3.plot(time[inx0:inx1] - time[inx0], ourvolume[inx0:inx1], color='#ff7f0e')
#byaverage2, = ax3.plot(time2[inx0:inx1] - time2[0], volume_by_average2[inx0:inx1], color='#ff7f0e')
byhighpass2, = ax3.plot(time2[inx2:inx3] - time2[inx2], ourvolume2[inx2:inx3], color='#1f77b4')
#ax3.legend([byaverage, byhighpass], ["corrected by flow average", "corrected by high-pass filter"])
ax3.legend([byhighpass,byhighpass2], ["auxport", "expiratory"], loc="upper right")
ax3.set_ylabel("volume (L)")
ax3.set_xlim(xlim0, xlim1)
ax3.set_xlabel("time since start (sec)")
#fig.savefig("/tmp/helmet-data.png")

# %% [code]
# function to read CO2 data
def read_co2(filename):
    with open(filename) as file:
        time, co2 = [], []
        for line in file:
            try:
                row = json.loads(line)
            except:
                pass
            else:
                if len(row) > 10:
                  time.append(row["t"] / 1000.0)
                  co2.append(row["CO2"])
        return np.array(time), np.array(co2)

# %% [code]
# read CO2 data
timeco2, co2 = read_co2("device_raw_waveforms_expiratory.json")

# %% [code]
# time synchronization for CO2 data
timestart = timesync+duration_in_sec
syncco2 = np.where(timeco2 >= timestart)[0][0]
timesyncco2end = timestart+duration1_in_sec
windowco2 = np.where(timeco2 >= timesyncco2end)[0][0] - syncco2
offco2 = -52
inx0co2 = syncco2 + offco2
inx1co2 = syncco2 + offco2 + windowco2
print(inx0co2,inx1co2)

# %% [code]
# plots of pressure, flow, volume and CO2 for auxport and expiratory paths
#xlim0 = 0.
#xlim1 = 40.
xlim0 = time2[inx2] - time2[inx2]
xlim1 = time2[inx3] - time2[inx2]
fig, (ax1, ax2, ax3, ax4) = plt.subplots(4, 1, figsize=(25, 15))
#rawpressure, = ax1.plot(time[inx0:inx1] - time[0], pressure[inx0:inx1], color='#ff7f0e')
deglitched, = ax1.plot(time[inx0:inx1] - time[inx0], pressure_deglitched[inx0:inx1], color='#ff7f0e')
#rawpressure2, = ax1.plot(time2[inx0:inx1] - time2[0], pressure2[inx0:inx1], color='#ff7f0e')
deglitched2, = ax1.plot(time2[inx2:inx3] - time2[inx2], pressure_deglitched2[inx2:inx3], color='#1f77b4')
#ax1.legend([rawpressure, deglitched], ["raw pressure", "deglitched"], loc="upper right")
ax1.legend([deglitched,deglitched2], ["auxport", "expiratory"], loc="upper right")
ax1.set_ylabel("pressure (cm H₂O)")
ax1.set_xlim(xlim0, xlim1)
qlflow, = ax2.plot(time[inx0:inx1] - time[inx0], flow[inx0:inx1], color='#ff7f0e')
expflow, = ax2.plot(time2[inx2:inx3] - time2[inx2], flow2[inx2:inx3], color='#1f77b4')
#co2plot, = ax2.plot(timeco2[inx0co2:inx1co2] - timeco2[inx0co2], co2[inx0co2:inx1co2]/100.0, color='green')
ax2.legend([qlflow,expflow], ["auxport", "expiratory"], loc="upper right")
ax2.set_ylabel("flow (L/min)")
ax2.set_xlim(xlim0, xlim1)
#byaverage, = ax3.plot(time[inx0:inx1] - time[0], volume_by_average[inx0:inx1], color='#ff7f0e')
byhighpass, = ax3.plot(time[inx0:inx1] - time[inx0], ourvolume[inx0:inx1], color='#ff7f0e')
#byaverage2, = ax3.plot(time2[inx0:inx1] - time2[0], volume_by_average2[inx0:inx1], color='#ff7f0e')
byhighpass2, = ax3.plot(time2[inx2:inx3] - time2[inx2], ourvolume2[inx2:inx3], color='#1f77b4')
#ax3.legend([byaverage, byhighpass], ["corrected by flow average", "corrected by high-pass filter"])
ax3.legend([byhighpass,byhighpass2], ["auxport", "expiratory"], loc="upper right")
ax3.set_ylabel("volume (L)")
ax3.set_xlim(xlim0, xlim1)
ax3.set_xlabel("time since start (sec)")
co2plot, = ax4.plot(timeco2[inx0co2:inx1co2] - timeco2[inx0co2], co2[inx0co2:inx1co2], color='green')
ax4.legend([co2plot], ["CO$_2$"], loc="upper right")
ax4.set_ylabel("CO$_2$ (ppm)")
ax4.set_ylim(0.0,6000.0)
ax4.set_xlim(xlim0, xlim1)
ax4.set_xlabel("time since start (sec)")
#fig.savefig("/tmp/helmet-data-co2.png")

# %% [code]
# plots of pressure, flow, volume and CO2 for expiratory path
#xlim0 = 0.
#xlim1 = 40.
xlim0 = time2[inx2] - time2[inx2]
xlim1 = time2[inx3] - time2[inx2]
fig, (ax1, ax2, ax3, ax4) = plt.subplots(4, 1, figsize=(25, 15))
#rawpressure, = ax1.plot(time[inx0:inx1] - time[0], pressure[inx0:inx1], color='#ff7f0e')
#deglitched, = ax1.plot(time[inx0:inx1] - time[inx0], pressure_deglitched[inx0:inx1], color='#ff7f0e')
#rawpressure2, = ax1.plot(time2[inx0:inx1] - time2[0], pressure2[inx0:inx1], color='#ff7f0e')
deglitched2, = ax1.plot(time2[inx2:inx3] - time2[inx2], pressure_deglitched2[inx2:inx3], color='#1f77b4')
#ax1.legend([rawpressure, deglitched], ["raw pressure", "deglitched"], loc="upper right")
#ax1.legend([deglitched,deglitched2], ["auxport", "expiratory"], loc="upper right")
ax1.set_ylabel("pressure (cm H₂O)")
ax1.set_xlim(xlim0, xlim1)
#qlflow, = ax2.plot(time[inx0:inx1] - time[inx0], flow[inx0:inx1], color='#ff7f0e')
expflow, = ax2.plot(time2[inx2:inx3] - time2[inx2], flow2[inx2:inx3], color='#1f77b4')
#co2plot, = ax2.plot(timeco2[inx0co2:inx1co2] - timeco2[inx0co2], co2[inx0co2:inx1co2]/100.0, color='green')
#ax2.legend([qlflow,expflow], ["auxport", "expiratory"], loc="upper right")
ax2.set_ylabel("flow (L/min)")
ax2.set_xlim(xlim0, xlim1)
#byaverage, = ax3.plot(time[inx0:inx1] - time[0], volume_by_average[inx0:inx1], color='#ff7f0e')
#byhighpass, = ax3.plot(time[inx0:inx1] - time[inx0], ourvolume[inx0:inx1], color='#ff7f0e')
#byaverage2, = ax3.plot(time2[inx0:inx1] - time2[0], volume_by_average2[inx0:inx1], color='#ff7f0e')
byhighpass2, = ax3.plot(time2[inx2:inx3] - time2[inx2], ourvolume2[inx2:inx3], color='#1f77b4')
#ax3.legend([byaverage, byhighpass], ["corrected by flow average", "corrected by high-pass filter"])
#ax3.legend([byhighpass,byhighpass2], ["auxport", "expiratory"], loc="upper right")
ax3.set_ylabel("volume (L)")
ax3.set_xlim(xlim0, xlim1)
ax3.set_xlabel("time since start (sec)")
co2plot, = ax4.plot(timeco2[inx0co2:inx1co2] - timeco2[inx0co2], co2[inx0co2:inx1co2], color='green')
#ax4.legend([co2plot], ["CO$_2$"], loc="upper right")
ax4.set_ylabel("CO$_2$ (ppm)")
ax4.set_ylim(0.0,6000.0)
ax4.set_xlim(xlim0, xlim1)
ax4.set_xlabel("time since start (sec)")
fig.savefig("/tmp/helmet-data-co2.png")

# %% [code]
# comparison of volumes of auxport and expritory paths
#xlim0 = 0.
#xlim1 = 40.
xlim0 = time2[inx2] - time2[inx2]
xlim1 = time2[inx3] - time2[inx2]
fig, (ax1, ax2) = plt.subplots(2, 1, figsize=(25, 15))
#byaverage, = ax3.plot(time[inx0:inx1] - time[0], volume_by_average[inx0:inx1], color='#ff7f0e')
byhighpass, = ax1.plot(time[inx0:inx1] - time[inx0], ourvolume[inx0:inx1], color='#ff7f0e')
#byaverage2, = ax3.plot(time2[inx0:inx1] - time2[0], volume_by_average2[inx0:inx1], color='#ff7f0e')
byhighpass2, = ax1.plot(time2[inx2:inx3] - time2[inx2], ourvolume2[inx2:inx3], color='#1f77b4')
#ax3.legend([byaverage, byhighpass], ["corrected by flow average", "corrected by high-pass filter"])
ax1.legend([byhighpass,byhighpass2], ["auxport", "expiratory"], loc="upper right")
ax1.set_ylabel("volume (L)")
ax1.set_xlim(xlim0, xlim1)
ax1.set_xlabel("time since start (sec)")
from scipy.signal import find_peaks
RRmin = 3.  # 3 Hz min respiratory rate (bpm)
RRmax = 60. # 60 Hz max respiratory rate (bpm)
peak_dist_min = int(60.*50/RRmax) # index distance computed using 50 Hz (=30*50/min) of flow readings
Vmin = 0.05 # 50mL min volume (L)
Vwidth_min = int(60.*50/RRmax/2.) # 25*0.02s = 0.5s
Vwidth_max = int(60.*50/RRmin) # 1000*0.02s = 20.0s equivalent to 3 Hz
p = pressure_deglitched[inx0:inx1]
p2 = pressure_deglitched2[inx2:inx3]
ppeaks, pprops = find_peaks(p, distance=peak_dist_min)
ppeaks2, pprops2 = find_peaks(p2, distance=peak_dist_min)
pp = p[ppeaks]
pp2 = p2[ppeaks2]
print("pressure aux",pp.mean(), pp.std(), "pressure exp", pp2.mean(), pp2.std())
f = flow[inx0:inx1]
f2 = flow2[inx2:inx3]
fpeaks, fprops = find_peaks(f, distance=peak_dist_min)
fpeaks2, fprops2 = find_peaks(f2, distance=peak_dist_min)
fp = f[fpeaks]
fp2 = f2[fpeaks2]
print("flow aux",fp.mean(), fp.std(), "flow exp", fp2.mean(), fp2.std())
v = ourvolume[inx0:inx1]
v2 = ourvolume2[inx2:inx3]
vpeaks, vprops = find_peaks(v, prominence=Vmin, distance=peak_dist_min, width=(Vwidth_min,Vwidth_max))
vpeaks2, vprops2 = find_peaks(v2, prominence=Vmin, distance=peak_dist_min, width=(Vwidth_min,Vwidth_max))
vp = v[vpeaks]
vp2 = v2[vpeaks2]
t = time[inx0:inx1] - time[inx0]
tf = t[vpeaks]
ax1.plot(tf,vp, 'x', color='#ff7f0e')
ax1.vlines(x=tf,ymin=vp-vprops["prominences"],ymax=vp, color='#ff7f0e')
ax1.hlines(y=vprops["width_heights"], xmin=t[vprops["left_ips"].astype(int)], xmax=t[vprops["right_ips"].astype(int)], color='#ff7f0e')
t2 = time2[inx2:inx3] - time2[inx2]
tf2 = t2[vpeaks2]
ax1.plot(tf2,vp2, 'x', color='#1f77b4')
ax1.vlines(x=tf2,ymin=vp2-vprops2["prominences"],ymax=vp2, color='#1f77b4')
ax1.hlines(y=vprops2["width_heights"], xmin=t2[vprops2["left_ips"].astype(int)], xmax=t2[vprops2["right_ips"].astype(int)], color='#1f77b4')
vf = vprops2["prominences"]/vprops["prominences"]
print("volume aux", vprops["prominences"].mean(), vprops["prominences"].std())
print("volume exp", vprops2["prominences"].mean(), vprops2["prominences"].std())
print("aux/exp frac", vf[1:].mean()-1.0,vf[1:].std()) # drop first breath
volfrac, = ax2.plot(tf, vf, label="auxport/expiratory", color='black')
ax2.legend(loc="upper right")
ax2.set_ylabel("volume ratio (L)")
ax2.set_xlim(xlim0, xlim1)
ax2.set_ylim(0.7,1.25)
ax2.set_xlabel("time since start (sec)")
fig.savefig("/tmp/helmet-data.png")

# %% [code]
# histogram of CO2 data
fig, ax = plt.subplots(1, 1, figsize=(15, 15))
#ax.hist(co2[inx0co2:inx1co2],bins=np.arange(0,6300,300), density=True)
bin_edges=np.arange(0,6500,300)
Nullco2=co2[100:163]
HST2co2=co2[761:889]
AP52co2=co2[328:364]
AP83co2=co2[233:254]
AP105co2=co2[419:442]
print(inx2,time2[inx2],inx0co2,timeco2[inx0co2],timeco2[100])
finx = lambda i : np.where(time2 > timeco2[i] + time2[inx2] - timeco2[inx0co2])[0][0]
Nullflow2=flow2[finx(100):finx(163)]
HST2flow2=flow2[finx(761):finx(889)]
AP52flow2=flow2[finx(328):finx(364)]
AP83flow2=flow2[finx(233):finx(254)]
AP105flow2=flow2[finx(419):finx(442)]
ax.hist(Nullco2,bins=bin_edges, density=True, histtype='step')
ax.hist(HST2co2,bins=bin_edges, density=True, histtype='step')
ax.hist(AP52co2,bins=bin_edges, density=True, histtype='step')
ax.hist(AP83co2,bins=bin_edges, density=True, histtype='step')
ax.hist(AP105co2,bins=bin_edges, density=True, histtype='step')
ax.set_ylim(0.0,0.003)
ax.set_ylabel("Number of CO$_2$ Readings/2 sec")
ax.set_xlabel("CO$_2$ (ppm)")
fig.savefig("/tmp/helmet-data-co2-hist.png")
print(inx0co2,inx1co2,np.mean(co2[inx0co2:inx1co2]),np.std(co2[inx0co2:inx1co2]))
# HST Stab1e 1 669 749 3491.773126220703 622.083534096037
# HST Stable 2 761 889 3316.8309803009033 731.7660514714353
# AuxPort 52 L/min 328 364 5010.739732530382 464.92444413853315
# AuxPort 83 L/min 233 254 2710.000558035714 287.2526062913333
# AuxPort 105 L/min 419 442 2176.2248269786005 478.1798992473555
# null data 83 L/min 100 163 412.1304713657924 92.42008635199817

# %% [code]
# plots of CO2 data versus flow
plt.rcParams.update({'axes.xmargin': 0.2})
#plt.rcParams.update({'axes.ymargin': 0.0})
plt.rcParams.update({'lines.linewidth': 2.0})
plt.rcParams.update({'lines.markersize': 20.0})
fig, ax = plt.subplots(1, 1, figsize=(20, 10))
mean = np.zeros(5)
rms = np.zeros(5)
fmean = np.zeros(5)
frms = np.zeros(5)
mean[0] = np.mean(Nullco2)
rms[0] = np.std(Nullco2)
mean[1] = np.mean(HST2co2)
rms[1] = np.std(HST2co2)
mean[2] = np.mean(AP52co2)
rms[2] = np.std(AP52co2)
mean[3] = np.mean(AP83co2)
rms[3] = np.std(AP83co2)
mean[4] = np.mean(AP105co2)
rms[4] = np.std(AP105co2)
fmean[0] = np.mean(Nullflow2)
frms[0] = np.std(Nullflow2)
fmean[1] = np.mean(HST2flow2)
frms[1] = np.std(HST2flow2)
fmean[2] = np.mean(AP52flow2)
frms[2] = np.std(AP52flow2)
fmean[3] = np.mean(AP83flow2)
frms[3] = np.std(AP83flow2)
fmean[4] = np.mean(AP105flow2)
frms[4] = np.std(AP105flow2)
#flow = np.array([83.0,83.0,52.0,83.5,105.0])
print(fmean,frms,mean,rms)
Nullco2plot = ax.errorbar(x=fmean[0],y=mean[0],xerr=frms[0], yerr=rms[0], marker='.', linestyle='', color='black')
HST2co2plot = ax.errorbar(x=fmean[1],y=mean[1],xerr=frms[1], yerr=rms[1], marker='.', linestyle='', color='blue')
APco2plot = ax.errorbar(x=fmean[2:5],y=mean[2:5],xerr=frms[2:5], yerr=rms[2:5], marker='.', linestyle='', color='green')
ax.set_ylabel("CO$_2$ (ppm)")
ax.set_xlabel("Flow (L/min)")
ax.legend([Nullco2plot,HST2co2plot,APco2plot], ["Medical Air","Helmet Breathing","Auxiliary Port Breathing"], loc="upper right")
```
